# Proteomic Network Analysis of Alzheimer’s Disease Cerebrospinal Fluid Reveals Alterations Associated with *APOE* ε4 Genotype and Atomoxetine Treatment

**DOI:** 10.1101/2023.10.29.23297651

**Authors:** Eric B. Dammer, Anantharaman Shantaraman, Lingyan Ping, Duc M. Duong, Ekaterina S. Gerasimov, Suda Parimala Ravindran, Valborg Gudmundsdottir, Elisabet A. Frick, Gabriela T. Gomez, Keenan A. Walker, Valur Emilsson, Lori L. Jennings, Vilmundur Gudnason, Daniel Western, Carlos Cruchaga, James J. Lah, Thomas S. Wingo, Aliza P. Wingo, Nicholas T. Seyfried, Allan I. Levey, Erik C.B. Johnson

## Abstract

Alzheimer’s disease (AD) is currently defined at the research level by the aggregation of amyloid-β (Aβ) and tau proteins in brain. While biofluid biomarkers are available to measure Aβ and tau pathology, few biomarkers are available to measure the complex pathophysiology that is associated with these two cardinal neuropathologies. Here we describe the proteomic landscape of cerebrospinal fluid (CSF) changes associated with Aβ and tau pathology in 300 individuals as assessed by two different proteomic technologies—tandem mass tag (TMT) mass spectrometry and SomaScan. Harmonization and integration of both data types allowed for generation of a robust protein co-expression network consisting of 34 modules derived from 5242 protein measurements, including disease-relevant modules associated with autophagy, ubiquitination, endocytosis, and glycolysis. Three modules strongly associated with the apolipoprotein E ε4 (*APOE* ε4) AD risk genotype mapped to oxidant detoxification, mitogen associated protein kinase (MAPK) signaling, neddylation, and mitochondrial biology, and overlapped with a previously described lipoprotein module in serum. Neddylation and oxidant detoxification/MAPK signaling modules had a negative association with *APOE* ε4 whereas the mitochondrion module had a positive association with *APOE* ε4. The directions of association were consistent between CSF and blood in two independent longitudinal cohorts, and altered levels of all three modules in blood were associated with dementia over 20 years prior to diagnosis. Dual-proteomic platform analysis of CSF samples from an AD phase 2 clinical trial of atomoxetine (ATX) demonstrated that abnormal elevations in the glycolysis CSF module—the network module most strongly correlated to cognitive function—were reduced by ATX treatment. Individuals who had more severe glycolytic changes at baseline responded better to ATX. Clustering of individuals based on their CSF proteomic network profiles revealed ten groups that did not cleanly stratify by Aβ and tau status, underscoring the heterogeneity of pathological changes not fully reflected by Aβ and tau. AD biofluid proteomics holds promise for the development of biomarkers that reflect diverse pathologies for use in clinical trials and precision medicine.

## Introduction

Alzheimer’s disease (AD) is a growing public health problem with few effective treatments. AD therapeutic development has traditionally focused on two proteins that define the disease at the neuropathological level—amyloid-β (Aβ) and microtubule associated protein tau (MAPT, or tau)(*1*). However, it has become increasingly appreciated that AD represents a complex neurological disorder with many other pathological changes not readily observable by classical neuropathological examination, including alterations in proteostasis, synaptic and neuronal signaling, endocytic trafficking, mitochondrial function, and energy metabolism, among others(*2-4*). Examining how these pathological changes evolve over the course of the disease and are related to one another and to Aβ and tau pathology remains a challenge in the absence of tools to interrogate these brain processes in individuals during life.

Cerebrospinal fluid (CSF) is the most proximate biofluid to brain in which to assess normal and abnormal brain physiology. Proteomic analysis of AD CSF to assess disease-associated changes has most commonly been performed using unbiased mass spectrometry-based approaches and has led to a common set of robust changes observed across multiple studies(*3, 5-9*). More recently, antibody-based proximity extension assay (Olink) and modified nucleic acid aptamer (SomaScan) targeted assay platforms have been used to characterize AD CSF proteomic changes(*10-15*). However, relatively few studies have examined how proteomic measurements compare across these platforms in AD. A recent study by our group analyzed AD CSF by tandem mass tag mass spectrometry (TMT-MS), Olink, and SomaScan platforms in 36 individuals and found proteomic changes common and unique to each platform(*10*). Integration of these different proteomic data types allowed for deep profiling of CSF samples beyond that afforded by any one platform, and for technical assessment of protein measurements common to more than one platform. Such deep profiling is important for capturing biological changes related to AD, some of which may be observable by proteomic analysis of cadaveric AD brain tissue, and many of which are not observable by traditional neuropathological examination. The ability to assess the response of different pathological alterations in AD beyond Aβ and tau levels to pharmacologic and non-pharmacologic therapeutic interventions at various stages of disease is critical to broadening and advancing the treatment landscape for AD. Also important is the ability to measure which pathological alterations are present in any given patient in the preclinical or symptomatic phases of the disease, regardless of Aβ and tau pathological burden, for effective personalized therapeutic approaches to disease modification.

In an effort to advance our AD biomarker toolkit to assess the heterogenous pathology of AD, here we measure greater than 4,500 unique protein gene products after application of quality control criteria in control and AD CSF using TMT-MS and SomaScan in 300 individuals, and analyze the resulting harmonized data with protein co-expression to generate a highly robust CSF network that reveals significant changes in autophagy, ubiquitination, synaptic vesicle, glycolysis, redox, and endocytosis pathways, among others, in AD. We identify three co-expression modules whose levels are strongly influenced by the apolipoprotein E ε4 (*APOE* ε4) genotype—the strongest genetic risk factor for late-onset AD—and assess the relationship of blood levels of these modules with AD risk. Towards clinical application of our findings, we demonstrate that a recent AD phase II clinical trial of atomoxetine—a promising disease-modifying medication which blocks reuptake of norepinephrine in brain and is currently approved by the FDA for treatment of attention deficit hyperactivity disorder—showed mitigation of pathologic brain glycolytic metabolic alterations as identified in our AD CSF proteomic network. We observe that patients with more severe alterations in our proteomic glycolytic biomarker panel at baseline have a larger response in this panel with ATX treatment. Finally, by using key features of the CSF proteome network to cluster control and AD participants, we find that many participants who would meet criteria as controls based on CSF Aβ and tau levels in fact appear more similar to those that meet criteria for AD based on CSF proteome features. Conversely, many participants who meet criteria for AD by Aβ and tau levels appear more similar to controls at the proteome level, illustrating the value of comprehensive proteomic analysis of AD biofluids for the realization of precision medicine approaches in AD.

## Results

### Cross-Platform Comparison of TMT-MS and SomaScan Protein Measurements in CSF

The general scheme of our study is shown in **Figure 1**. We analyzed CSF from 140 control and 160 AD participants by TMT-MS and SomaScan proteomic approaches (**Supplementary Tables 1, 2**). TMT-MS is a direct measurement of protein abundance, whereas SomaScan is an indirect affinity-based measurement. The two proteomic technologies therefore provide a complementary measure of the CSF proteome with certain advantages and disadvantages, as previously described(*10*). AD was determined based on the NIA research definition using the CSF total tau to Aβ ratio(*16-18*), with the threshold ratio for AD derived empirically from a Gaussian mixture model analysis of all participants from our center. TMT-MS was performed in multiple batches with multiple fractions per batch to increase depth of proteome coverage, but without depletion of highly abundant proteins to avoid potential bias introduced by protein depletion(*10*). SomaScan modified aptamer-based measurements included 7,596 assays, of which 7,288 targeted human proteins that were used in the analysis. We applied stringent quality control (QC) and pre-processing measures to SomaScan and TMT-MS data prior to analysis. Background signal variance was removed in the SomaScan data, and batch effects were removed from the TMT-MS data (**Supplementary Figure 1A, B**). Because CSF is collected by lumbar puncture, the possibility exists for blood contamination introduced during sampling. While blood contamination was not observable in our samples by visual inspection, we observed a subtle signal of blood contamination by proteomic inspection in a minority of samples in both TMT-MS and SomaScan datasets (**Supplementary Figure 1C**). Although the effects on the data were small, we nevertheless removed this variance from both TMT-MS and SomaScan datasets prior to analysis (**Supplementary Figure 1D, E**). SomaScan is designed for analysis of blood, and we have previously shown that many aptamer signals are at or near noise-level when applied to analysis of CSF(*10*). We removed SomaScan assays from consideration that did not meet our limit of detection (LOD) criteria (i.e., at least three standard deviations above background noise-level), and filtered additional low-signal assays based on an empirical signal-to-noise (S:N) correlation threshold with TMT-MS measurements (**Supp Figure 2A, B**) as previously described(*10*). This approach assumes that low signal measurements in SomaScan that do not correlate well with TMT-MS measurements of the same protein are unlikely to be robust protein measurements, and therefore removing these measurements up to a certain S:N threshold to maximize correlation with TMT-MS measurements enriches for true signals. These QC steps resulted in a final count of 4,098 (56%) assays used in the analyses out of the original 7,288 human SomaScan assays available. We validated our QC approach by comparing the overall fraction of assays discarded based on our QC criteria *versus* the number of assays on the same SomaScan platform that have been shown to have signals in CSF affected by genetic variation (i.e., that have a protein quantitative trait locus, or pQTL)(*19*) (**Supplementary Figure 2C**). 88% of SomaScan aptamers with CSF pQTLs were retained after the LOD and S:N filtering QC steps, demonstrating that our SomaScan aptamer QC approach retained biologically meaningful assays while discarding most low S:N assays. We also directly tested the proportion of CSF pQTLs in our cohort that were in the retained versus discarded aptamer pools (**Supplementary Figure 2D, Supplementary Table 3**). We identified only 1 protein with pQTLs out of 2,854 proteins tested in the discarded pool (0.04%), and 152 proteins with pQTLs out of 3,863 proteins tested in the retained pool (3.9%), further validating our QC approach. In a final QC step, we removed proteins with missing measurements in greater than 75% of the 300 CSF samples, ensuring remaining measurements were sufficiently present in both AD and control cases for statistical analyses (**Supplementary Figure 2E, F**). Therefore, the final number of unique proteins (or gene symbols) retained for analysis was 3,649 for SomaScan and 2,195 for TMT-MS, with a total of 4,576 unique proteins measured between the two platforms (**Figure 2A**). TMT-MS preferentially measured complement proteins and immunoglobulins, whereas SomaScan preferentially measured nuclear proteins (**Supplementary Figure 3A**). Median correlation between measurements common to the two platforms was 0.59, with a broad distribution of correlation values (**Figure 2B, Supplementary Table 4, Extended Data**). The median correlation was not significantly different when analyzing only proteins that were differentially abundant in AD (**Supplementary Figure 3B**). Some AD-relevant proteins such as chitotriosidase-1 (CHIT1), neurogranin (NRGN), and SPARC-related modular calcium-binding protein 1 (SMOC1) correlated well across platforms (**Figure 2C**), while others such as amyloid-β precursor protein (APP), apolipoprotein E (APOE), and chitinase-3-like protein 1 (CHI3L1, also known as YKL-40) correlated poorly (**Figure 2D**). The use of an orthogonal direct measurement platform such as TMT-MS allowed for the interpretation of SomaScan protein measurements in which multiple aptamers target the same protein, such as Parkinson disease protein 7 (PARK7) and α-1-antichymotrypsin (SERPINA3) (**Figure 2E**). Differential abundance analysis on both proteomic datasets demonstrated a similar number of increased proteins in AD, with an overlap of approximately 21 percent between the platforms (**Figure 2F, G; Supplementary Tables 5and 6**) due to differences in coverage and statistical significance. More proteins were observed to be decreased in the SomaScan data (**Figure 2G**). Strength and statistical significance of differential abundance did not always correlate perfectly with significance of correlation between the platforms, as illustrated by protein complement C3 where the aptamer with the best correlation to the TMT-MS measurement was not the most significantly differentially abundant SOMAmer to this protein in the SomaScan data (**Figure 2H**). Both platforms covered a similar percentage of proteins from a previously reported AD brain co-expression network (**Supplementary Figure 4A**)(*2*). In summary, we were able to measure greater than 4,500 proteins in CSF by two orthogonal proteomic approaches across 300 AD and control individuals, and after thorough QC, compare the measurements within subject between platforms. These analyses demonstrated the unique and common attributes of each platform for assessing disease-relevant protein changes in AD CSF, and the utility of using multiple platforms for robust assessment of potential AD biomarkers.

**Figure 1.**
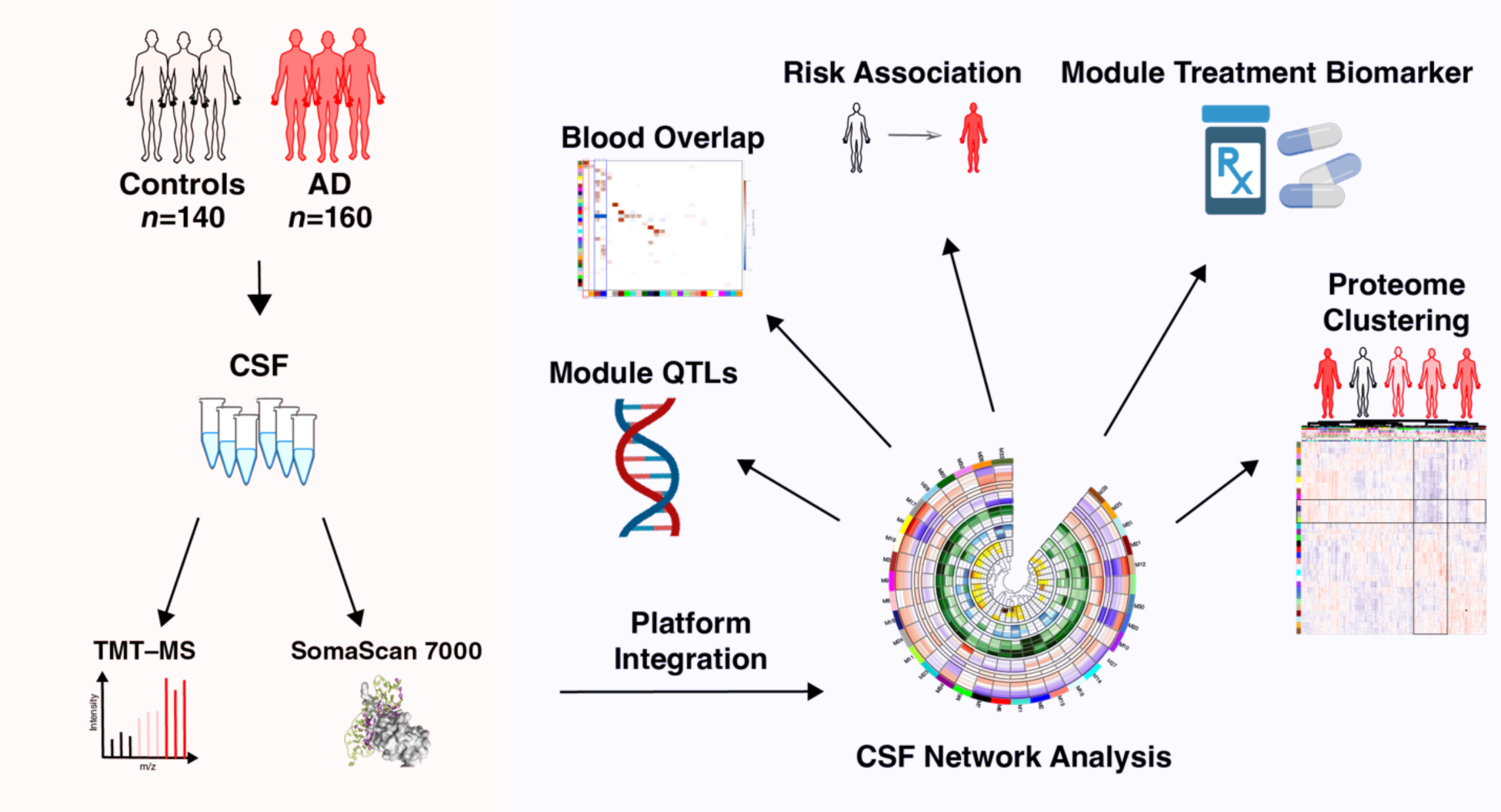
Study Overview. Cerebrospinal fluid (CSF) was sampled from 140 controls and 160 AD participants as defined by their CSF tTau/Aβ_1–42_ ratio. CSF proteomes for each person were obtained by tandem mass tag mass spectrometry (TMT-MS) and SomaScan 7000 assays. After data quality control and cross-platform comparison, data from both platforms were integrated to generate an AD CSF protein co-expression network. Genetic influence on the network was assessed by module quantitative trait locus analysis (modQTL). The network was compared to a blood network as described in Emilsson et al.(*24*), and association of CSF module proteins in blood with risk of AD was assessed in the AGES-Reykjavik and ARIC studies. The CSF AD network was used to assess the effect of pharmacologic intervention with atomoxetine on disease-relevant pathophysiology. Finally, key hub proteins from across the network were used to cluster participants into groups based on similarity in their CSF proteomic features regardless of their CSF Aβ and tTau status.

**Figure 2.**
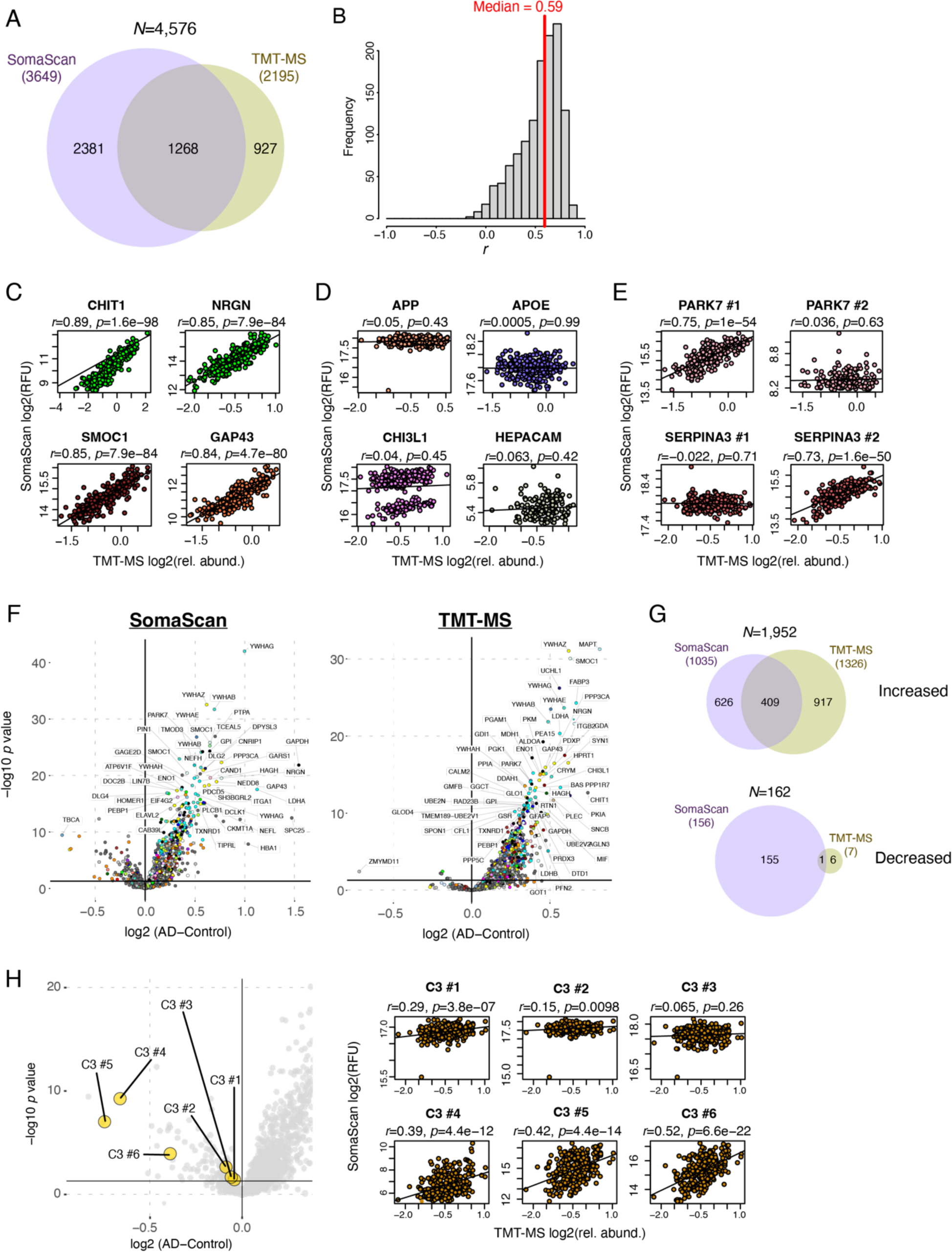
Cross-Platform Proteomic Comparisons. (A) Overlap of unique proteins measured by TMT-MS and SomaScan in CSF across 300 individuals after quality control filtering in both platforms. Numbers represent counts of gene symbols. Only unique gene symbol overlap was considered. (B) Distribution of within-subject correlation of protein measurements shared between platforms. Measurements were required to have a minimum of 74 total observations and 3 measurements per diagnostic group in each platform (*n*=1274). The vertical red line indicates the median correlation. (C-E) Illustration of AD-relevant proteins that are strongly correlated between platforms (C), poorly correlated between platforms (D), and variably correlated depending on the SOMAmer used for correlation (E). (F) Differential abundance of proteins in AD as measured by SomaScan (left) and MS (right). Proteins increased in AD are located in the upper right quadrant. (G) Overlap of significantly increased (top) or decreased (bottom) proteins in each platform. (H) The six different SOMAmers for C3 protein highlighted in the SomaScan differential abundance volcano plot shown in panel (F) (left), and correlation to the MS C3 measurement for each SOMAmer (right). Significance of differential abundance was determined at *p*<0.05. Correlations were performed using Pearson correlation with Student’s test for significance. APOE, apolipoprotein E; APP, amyloid-β precursor protein; C3, complement C3; CHI3L1, chitinase-3-like protein 1; CHIT1, chitotriosidase-1; GAP43, neuromodulin; HEPACAM, hepatic and glial cell adhesion molecule; NRGN, neurogranin; PARK7, Parkinson disease protein 7; rel. abund., relative abundance; RFU, relative fluorescence units; SERPINA3, α-1-antichymotrypsin; SMOC1, SPARC-related modular calcium-binding protein 1.

### Integration of Proteomic Measurements to Identify Pathological Alterations in AD CSF

We leveraged proteomic measurements from TMT-MS and SomaScan to construct a protein co-expression network of AD CSF from a total of 5,242 protein assays between the two platforms (**Figure 3A, Supplementary Tables 7and 8, Extended Data**). Co-expression analysis aims to cluster proteins into groups (or “modules”) that are related to one another based on their common variation across individuals. These protein modules can represent various biological processes, pathways, and cell types. As we have previously demonstrated, co-expression allows for the integration of protein measurements across different platforms, as different measurements of the same protein will cluster within the same modules if the measurements are correlated(*10*). Our AD co-expression network consisted of 34 modules, most of which contained measurements from both platforms (**Supplementary Figure 4B**). Most modules were significantly correlated to at least one endophenotype of AD such as CSF levels of total tau (tTau), tau phosphorylated at residue 181 (pTau181), and Aβ_1–42_ as measured by enzyme-linked immunosorbent assay (ELISA). The module most strongly correlated to CSF tTau levels was M4 Autophagy/Ubiquitination, which contained MAPT itself as measured by TMT-MS (**Figure 3B**). Other modules strongly correlated to tTau were M12 Endocytosis/Ubiquitin binding and M17 Synaptic vesicle/SNAP-SNARE complex. The module most strongly correlated to CSF Aβ_1–42_ level was M20 Glycolysis/Redox homeostasis. M20 was also the module most strongly correlated to cognitive function (as assessed by the Montreal Cognitive Assessment, or MoCA) in the network (*r*=–0.64, *p*=1.1e^–35^), and was defined primarily by TMT-MS measurements. The M2 Complement/Coagulation module was the most highly preserved module to a previously described brain network, reflecting shared co-expression of complement biology between compartments. Strikingly, the network revealed three modules that were highly correlated to the number of *APOE* ε4 alleles present in the participants and each module was defined only by SomaScan measurements—M26 Neddylation, M33 Oxidant detoxification/Mitogen activated protein kinase (MAPK) signaling, and M34 Mitochondrion (**Figure 4**). Neddylation is a protein post-translation modification in which the ubiquitin-like protein NEDD8 is added to protein substrates, and is overactivated in many human cancers(*20-22*). Levels of M26 Neddylation and M33 Oxidant detoxification/MAPK signaling were significantly decreased in AD CSF and negatively correlated with increasing number of *APOE* ε4 alleles (**Figure 4A-D**). Both were modestly correlated with cognitive function (M33 *r*=0.23, *p*=8.0e^–5^; M26 *r*=0.33, *p*=8.4e^–9^) and Aβ_1–42_ level (M33 *r*=0.44, *p*=1.3e^–15^; M26 *r*=0.38, *p*=1.4e^–11^). M33 Oxidant detoxification/MAPK signaling was not strongly correlated with CSF tTau or pTau181 levels, but levels of M26 Neddylation were significantly negatively correlated with CSF tTau and pTau181 levels (tTau *r*=–0.45, *p*=2e^–16^; pTau181 *r*=–0.43, *p*=1.7e^–14^). M34 Mitochondrion was strongly increased in AD and with *APOE* ε4, with modest correlations to cognitive function, tTau and pTau181, and Aβ_1–42_ (**Figure 4E, F**). None of these *APOE*-related modules were well defined in our previously described TMT-MS AD brain network as assessed by overrepresentation and module preservation analyses (**Figure 3A**)(*2*). Notably, a MAPK signaling module previously described in brain was strongly related to AD pathology and cognitive decline, yet was increased in AD in opposite direction to the M33 Oxidant detoxification/MAPK signaling module in CSF(*2*). In similar discordant fashion, mitochondrial modules in brain are consistently decreased in AD(*2, 3, 7*), opposite of that observed with M34 Mitochondrion in CSF. Interestingly, while M26 Neddylation was not preserved at the level of co-expression in brain, the first principal component of protein expression of this module (or module eigenprotein) was decreased in both CSF and brain (**Figure 3A**), showing concordance in direction of change between compartments for this set of proteins.

**Figure 3.**
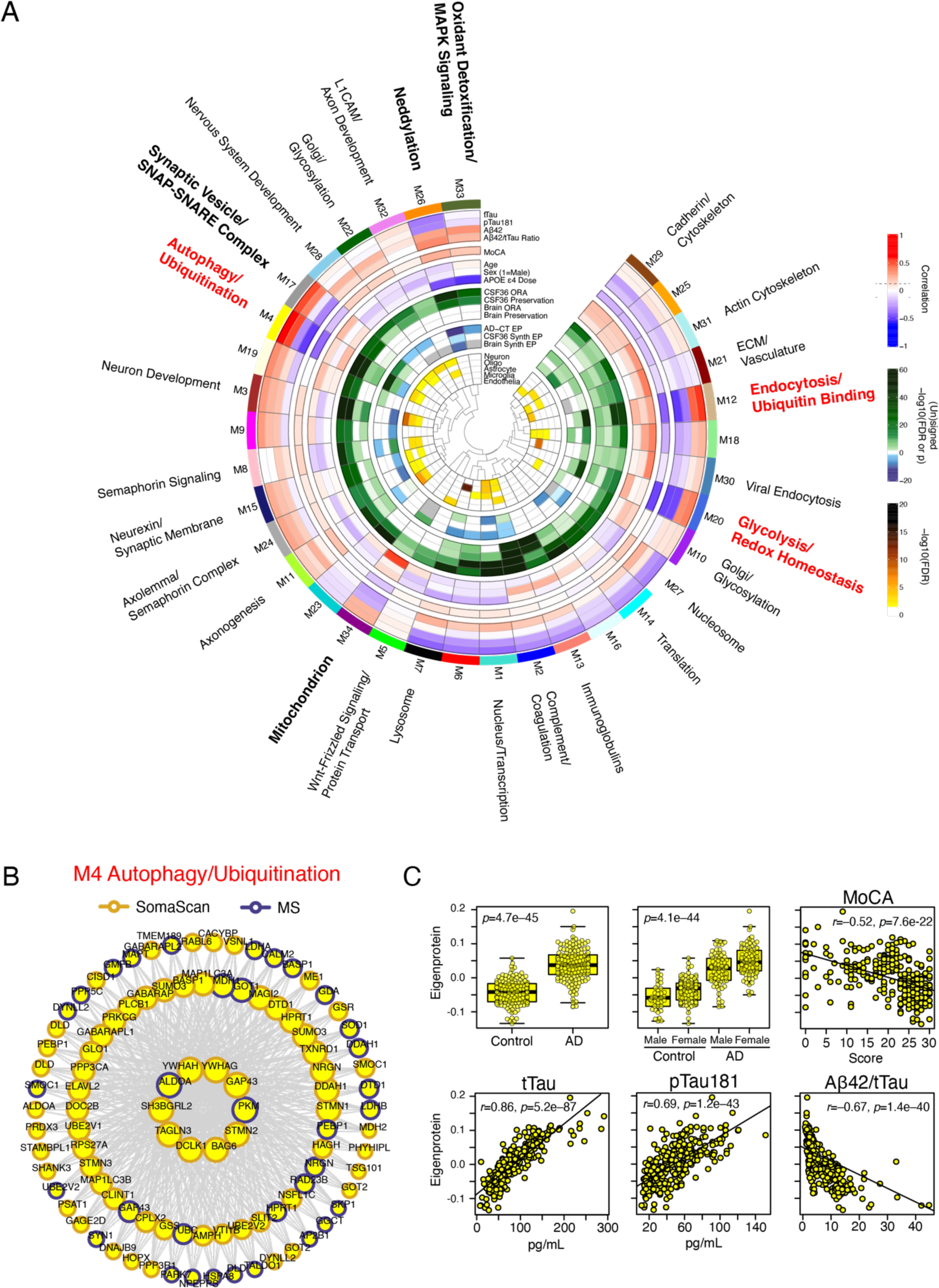
Multi-Platform AD CSF Protein Co-Expression Network. (A) A protein co-expression network of 5242 protein assays from SomaScan and TMT-MS platforms measured across 296 individuals after outlier removal identified 34 co-expression modules capturing different biological processes (outer ring). The module eigenprotein, or first principal component of module expression, was correlated with AD endophenotypes including CSF levels of total tau (tTau), tau phosphorylated at residue 181 (pTau181), amyloid-β 1-42 levels (Aβ_1–42_), tTau/Aβ_1–42_ ratio, and Montreal Cognitive Assessment (MoCA, higher scores reflect better cognitive function). Module eigenproteins were also correlated with age, sex, and number of *APOE* ε4 alleles (*APOE* ε4 dose). Correlations were considered significant at an absolute value of approximately *r*=0.1 (*p*=0.05, dashed lines in legend). Modules were tested for preservation in a previous CSF network generated on 36 individuals(*10*) (CSF36) by overrepresentation analysis (ORA) and network preservation statistics (preservation), as well as an AD brain network(*2*) (brain ORA, brain preservation). Green shading indicates preservation at varying degrees of significance. Module eigenproteins were tested for significant change in AD (AD-CT EP), significant change in AD in the CSF36 network (CSF36 synth EP), and significant change in AD in the brain network (brain synth EP). Color shading indicates direction and level of significance (blue=decreased in AD, green=increased in AD, gray=not calculated). Modules were also tested for overlap of protein membership with brain cell type specific protein markers for neurons, oligodendrocytes (oligo), astrocytes, microglia, and endothelia. Color shading indicates degree of significant overlap. L1CAM, neuronal cell adhesion molecule L1; ECM, extracellular matrix. (B) The top 100 proteins by intramodule correlation for the M4 Autophagy/Ubiquitination module. Larger circles represent stronger correlation, with the largest circles representing module “hub” proteins. Proteins are outlined by the proteomic platform by which they were measured. (C) M4 module eigenprotein levels in AD compared to controls, and correlation with AD endophenotypes. Correlations were performed with bicor. Differences between groups were assessed by *t* test or one-way ANOVA. For overlap and preservation tests, see Methods.

**Figure 4.**
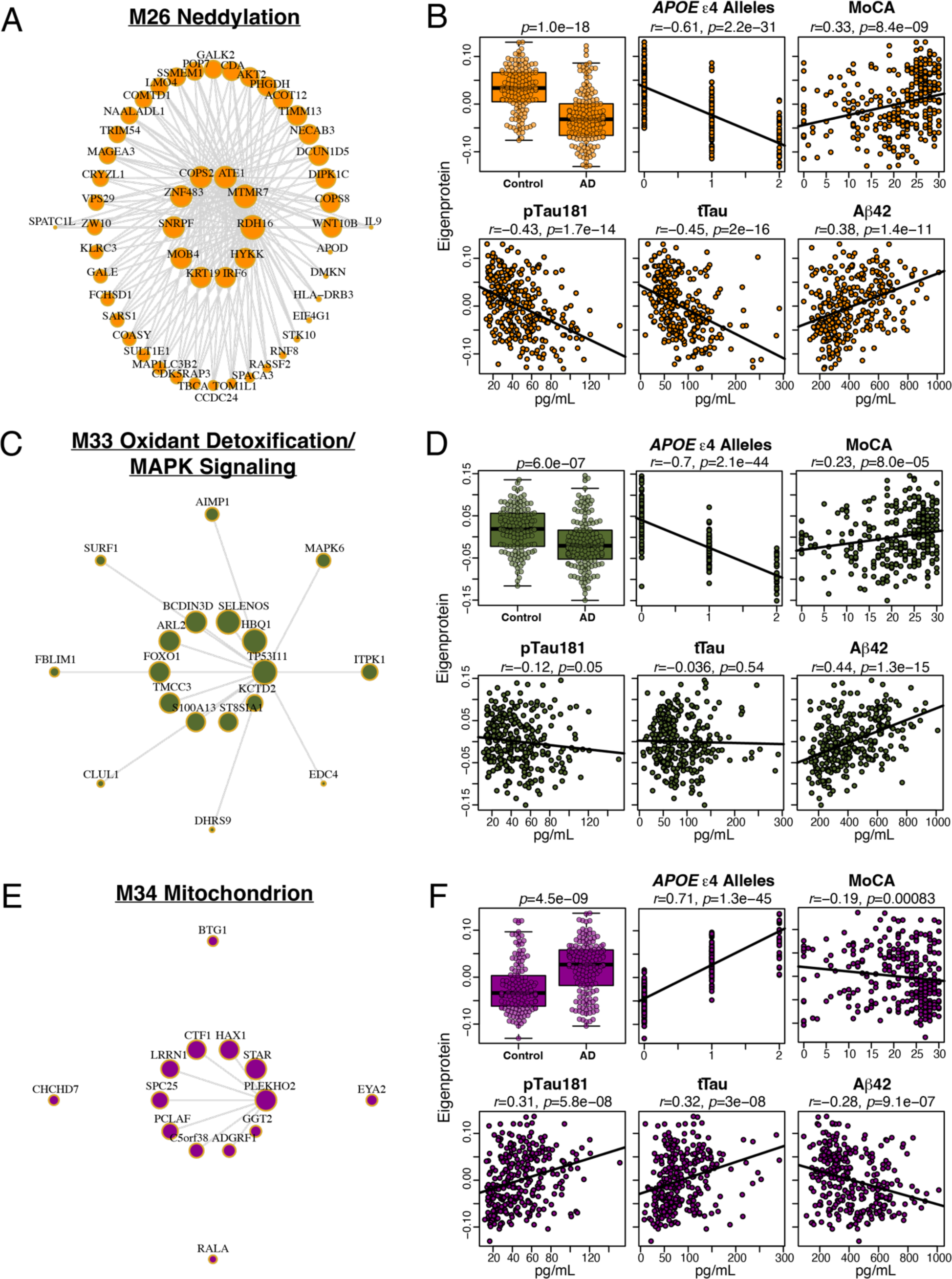
CSF AD Network Modules Correlated with *APOE* ε4 Dose. (A-F) Protein module membership, eigenprotein levels in control and AD, and correlation with number of *APOE* ε4 alleles and AD endophenotypes for M26 Neddylation (A, B), M33 Oxidant detoxification/MAPK signaling (C, D), and M34 Mitochondrion (E, F). Correlations were performed with bicor. Differences between groups were assessed by *t* test and adjusted for age and sex.

We validated the association of these three CSF modules with *APOE* ε4 by performing a module quantitative trait locus (mQTL) analysis(*2*). We observed a strong association of the *APOE* single nucleotide polymorphism (SNP) rs429358 that partially defines the *APOE* ε4 genotype with levels of M26, M33, and M34 (**Table 1**). These associations remained significant after adjustment for potential population stratification using a genomic control (GC) correction approach(*23*). In addition to the *APOE* ε4 associations, we observed significant associations after GC correction with SNPs near Pre-B-cell leukemia transcription factor 3 (PBX3) for the M32 L1CAM/Axon development module, CYFIP-related Rac1 interactor A (CYRIA) for the M9 Ambiguous module, and Ras guanyl-releasing protein 3 (RASGRP3) for the M20 Glycolysis/Redox homeostasis module, suggesting that these modules were also affected by genetic variation (Table 1). In summary, we were able to leverage both proteomic platforms to construct an AD CSF co-expression network that revealed multiple pathological changes beyond Aβ and tau dyshomeostasis, including changes under control of the *APOE* ε4 genotype.

**Table 1.** CSF Module Quantitative Trait Loci. SNP, single nucleotide polymorphism; CHR, chromosome; BP, base pair; A1, allele; mod-QTL, module quantitative trait locus; GC, genomic control. *”Yes” indicates genome-wide significant level at *p*<5 x 10^-8^

| SNP | CHR | BP | A1 | Nearest Coding Gene to SNP | BETA | P value | Module | mod-QTL | Significant after GC Correction* |
| --- | --- | --- | --- | --- | --- | --- | --- | --- | --- |
| **rs429358** | 19 | 45411941 | C | **APOE** | **-0.07** | **1.0E-52** | **M33 Oxidant Detoxification/MAPK Signaling** | *trans* | yes |
| **rs429358** | 19 | 45411941 | C | **APOE** | **-0.05** | **1.3E-27** | **M26 Neddylation** | *trans* | yes |
| rs10819083 | 9 | 128660207 | C | PBX3 | -0.03 | 1.1E-08 | M32 L1CAM/Axon Development | *trans* | yes |
| rs13409992 | 2 | 16575331 | T | CYRIA | -0.05 | 1.4E-08 | M9 Ambiguous | *trans* | yes |
| **rs429358** | 19 | 45411941 | C | **APOE** | **0.07** | **7.0E-64** | **M34 Mitochondrion** | *trans* | yes |
| rs6442288 | 3 | 11847081 | T | TAMM41 | 0.05 | 4.5E-08 | M13 Immunoglobulins | *trans* | no |
| rs539717210 | 7 | 23347402 | A | MALSU1 | 0.06 | 4.8E-08 | M16 Ambiguous | *trans* | no |
| rs11745580 | 5 | 149378425 | T | TIGD6 | 0.04 | 2.9E-08 | M27 Nucleosome | *trans* | no |
| rs2887087 | 2 | 33695073 | T | RASGRP3 | 0.02 | 3.5E-09 | M20 Glycolysis/Redox Homeostasis | *trans* | yes |
| rs736334 | 22 | 51139178 | T | SHANK3 | 0.03 | 2.8E-08 | M25 Ambiguous | *trans* | No |

### AD CSF Network Overlap with Blood Network Highlights Conservation of APOE-Related Modules Between Compartments

We assessed whether the three CSF modules influenced by the *APOE* ε4 genotype (M26, M33, and M34) were also represented in blood. To do so, we used a serum network generated from greater than 4,100 proteins measured by SomaScan in over 5,400 Icelandic individuals(*24*), and performed overrepresentation analysis across the networks (**Figure 5, Supplementary Table 9**). We found that most CSF modules had significant overlap with at least one serum module, but that only about half of serum modules had significant overlap with CSF modules. CSF M26, M33, and M34 overlapped with serum M11, with CSF M33 having particularly strong overlap with serum M11. Serum M11 has previously been shown to be a lipoprotein module that is genetically regulated by *APOE*, suggesting that the genetic regulation of this serum module overlapped with the genetic regulation of the *APOE*-regulated modules in CSF. CSF modules related to synaptic and neuronal biology (M32 L1CAM/Axon Development, M17 Synaptic Vesicle/SNAP-SNARE Complex, M3 Neuron Development, M15 Neurexin/Synaptic Membrane, M24 Axolemma/Semaphorin Complex, and M11 Axonogenesis) also overlapped with serum modules related to similar biology (M27 Axon Development/Semaphorin Complex and M26 Neuron Development/Ephrin Signaling), suggesting that neuronal biology as captured by CSF co-expression is also partly reflected in serum in an independent cohort.

**Figure 5.**
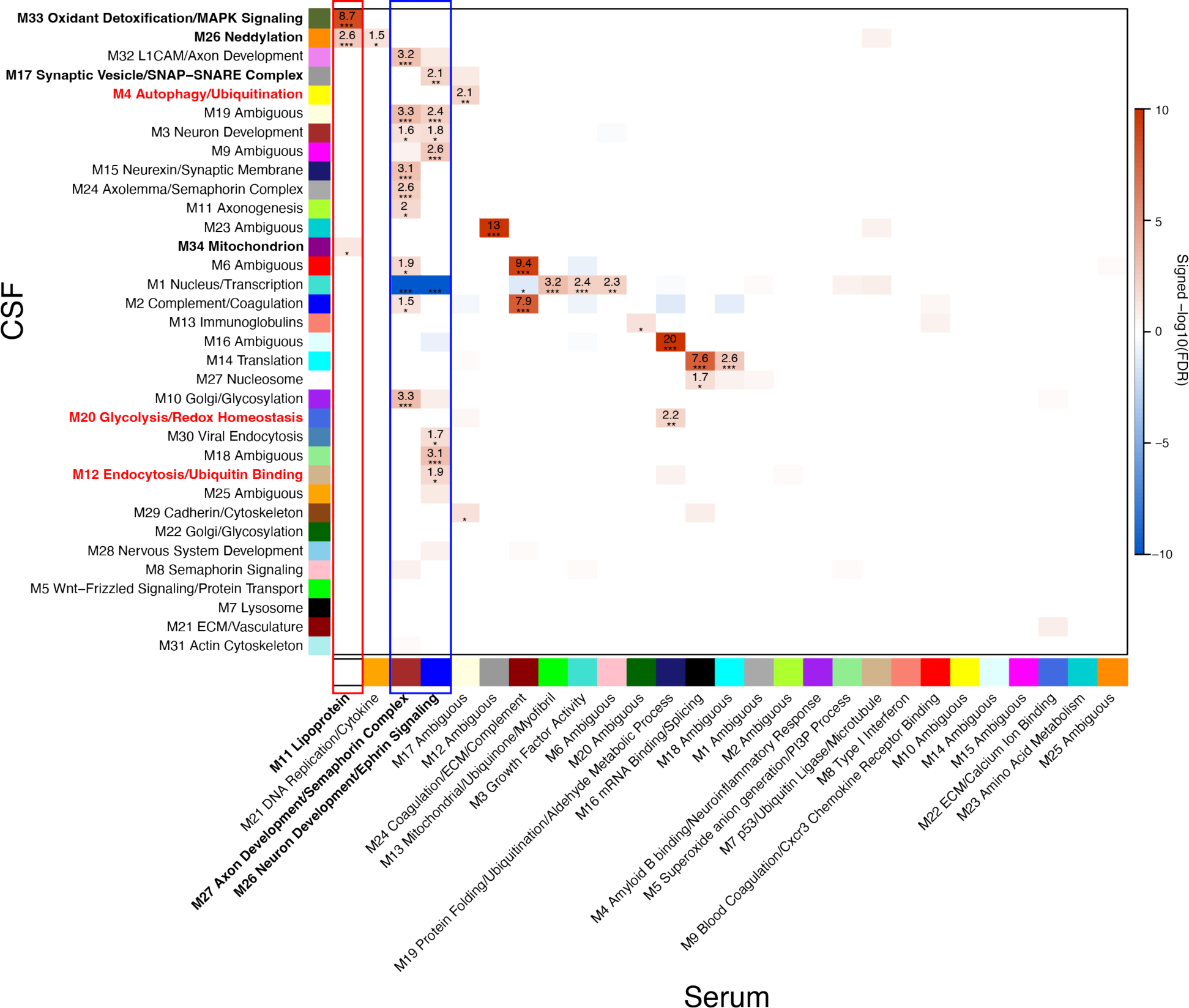
Overlap of CSF AD Network With Blood Network. CSF AD network modules were tested for overlap with serum network modules previously described in Emilsson *et al*.(*24*) CSF M33 Oxidant detoxification/MAPK signaling, M26 Neddylation, and M34 Mitochondrion modules significantly overlapped with the serum M11 Lipoprotein module (red box). Many other CSF modules overlapped with serum M27 Axon development/Semaphorin complex and M26 Neuron development/Ephrin signaling modules (blue box). Significance of overlap was determined using overrepresentation analysis and Fisher exact test with Benjamini-Hochberg correction. Red indicates overrepresentation, blue indicates underrepresentation.

### Levels of M33 Oxidant Detoxification/MAPK Signaling and M34 Mitochondrion Modules in Blood are Associated with Incident Dementia at Least 21 Years Prior to Diagnosis

Given the overlap of M26, M33, and M34 with blood, we next determined whether levels of these CSF *APOE*-related modules were also associated with *APOE* ε4 in blood. We calculated a synthetic eigenprotein (or first principal component) for 32 out of 34 CSF modules in serum using SomaScan protein measurements in 5,457 Icelanders from the AGES-Reykjavik cohort, a prospective population-based study (**Supplementary Tables 10and 11**)(*25*). We then tested for association of these modules with *APOE* ε4 in serum. We found all three CSF *APOE* ε4 modules were associated with *APOE* ε4 in serum in the same directions observed in CSF, with M33 and M34 particularly strongly associated (**Table 2**). Because AGES-Reykjavik is a longitudinal study with median follow-up of 12.8 years, we also tested whether levels of these three modules were associated with incident AD. M33 and M34 were associated with both prevalent and incident AD, with the association consistent with the directionality of relationship with *APOE* ε4 and dependent upon exclusion of *APOE* from the model (**Table 3**). These associations remained significant after a permutation test for randomization of protein module membership. We observed weaker association with incident AD for two other modules—M5 Wnt-Frizzled Signaling/Protein Transport and M31 Actin Cytoskeleton—although neither module was strongly associated with AD in CSF. We also tested associations of CSF module synthetic eigenproteins calculated in plasma with *APOE* ε4 and incident dementia in the Atherosclerosis Risk in Communities (ARIC) study (*N*=11,596), a multi-center longitudinal study of cardiovascular health that includes cognitive assessment(*26*) (**Supplementary Table 12**). M26 Neddylation, M33 Oxidant detoxification/MAPK signaling, and M34 Mitochondrion modules were found to have consistent associations with *APOE* ε4 in ARIC (**Table 2**). At a mean follow-up of 21 years we observed plasma levels of all three modules were associated with incident dementia in a model adjusted for cardiovascular risk factors given the nature of the ARIC cohort, with the association dependent upon *APOE* (**Table 3**). The associations were weaker but remained significant in a model that did not adjust for cardiovascular risk factors (**Supplementary Table 13**). In summary, the protein co-expression modules M26 Neddylation, M33 Oxidant detoxification/MAPK signaling, and M34 Mitochondrion observed in CSF that strongly correlated to *APOE* ε4 were also associated with *APOE* ε4 in blood in the same direction of change, and changes in the blood levels of these modules were associated with the development of AD or dementia up to 21 years later.

**Table 2.** Association of CSF *APOE* ε4 module synthetic eigenproteins in blood with *APOE* ε4 in the AGES-Reykjavik and ARIC cohorts. SE, standard error.

| Module | AGES-Reykjavik |  |  | ARIC |  |  |
| --- | --- | --- | --- | --- | --- | --- |
|  | Beta | SE | P | Beta | SE | P |
| M34 Mitochondrion | 0.86 | 0.03 | 3.7E-232 | 0.71 | 0.02 | 1.5E-283 |
| M33 Oxidant Detoxification/MAPK Signaling | -0.88 | 0.02 | 3.6E-272 | -0.84 | 0.02 | ~0 |
| M26 Neddylation | -0.10 | 0.03 | 2.7E-04 | -0.63 | 0.02 | 4.7E-217 |

**Table 3.** Association of CSF module synthetic eigenproteins in serum with incident and prevalent AD in the AGES-Reykjavik cohort and in plasma with incident dementia in the ARIC cohort. In AGES-Reykjavik incident AD (*n*=655) was determined a median of 13 years after baseline measurement. In ARIC, incident dementia (*n*=2270) was determined a mean of 21 years after baseline measurement. Further information on the cohorts is provided in **Supplementary** Tables 10 and 12. Associations were performed with and without inclusion of *APOE* in the model. In AGES-Reykjavik, the model was adjusted for age and sex. In ARIC, the model was adjusted for demographic variables, cardiovascular risk factors, and kidney function. Empirical *p* values were calculated after module membership permutation.

|  |  | Incident AD (AGES) or Dementia (ARIC) |  |  |  |  | Prevalent AD |  |  |  |  |
| --- | --- | --- | --- | --- | --- | --- | --- | --- | --- | --- | --- |
| Module | Model | HR | Lower CI | Upper CI | *P* | *emp P* | OR | Lower CI | Upper CI | *P* | *emp P* |
| AGES-Reykjavik (*N*=5,457) |  |  |  |  |  |  |  |  |  |  |  |
| M34 Mitochondrion | No *APOE* | 1.23 | 1.14 | 1.32 | 1.8E-07 | <1.0 E-03 | 1.25 | 1.07 | 1.46 | 4.0E-03 | 0.021 |
| M34 Mitochondrion | *APOE* | 1.05 | 0.96 | 1.14 | 0.31 | 0.58 | 1.00 | 0.84 | 1.19 | 0.99 | 0.99 |
| M33 Oxidant Detoxification/MAPK Signaling | No *APOE* | 0.81 | 0.75 | 0.88 | 2.6E-07 | 3.0E-03 | 0.70 | 0.60 | 0.82 | 6.2E-06 | 1.0E-03 |
| M33 Oxidant Detoxification/MAPK Signaling | *APOE* | 1.01 | 0.92 | 1.11 | 0.86 | 0.94 | 0.89 | 0.74 | 1.08 | 0.24 | 0.38 |
| M5 Wnt-Frizzled Signaling/Protein Transport | No *APOE* | 0.86 | 0.79 | 0.93 | 2.0E-04 | 0.03 | 1.04 | 0.88 | 1.22 | 0.64 | 0.67 |
| M5 Wnt-Frizzled Signaling/Protein Transport | *APOE* | 0.87 | 0.80 | 0.95 | 1.1E-03 | 0.03 | 1.09 | 0.92 | 1.29 | 0.33 | 0.61 |
| M31 Actin Cytoskeleton | No *APOE* | 0.85 | 0.78 | 0.92 | 5.9E-05 | 0.02 | 0.90 | 0.77 | 1.06 | 0.20 | 0.27 |
| M31 Actin Cytoskeleton | *APOE* | 0.89 | 0.82 | 0.97 | 6.8E-03 | 0.12 | 0.95 | 0.80 | 1.12 | 0.53 | 0.69 |
| ARIC (*N*=11,596) |  |  |  |  |  |  |  |  |  |  |  |
| M33 Oxidant Detoxification/MAPK Signaling | No *APOE* | 0.88 | 0.84 | 0.92 | 2.1E-09 | <1.0 E-03 |  |  |  |  |  |
| M33 Oxidant Detoxification/MAPK Signaling | *APOE* | 0.99 | 0.95 | 1.04 | 0.67 | 0.85 |  |  |  |  |  |
| M26 Neddylation | No *APOE* | 0.89 | 0.85 | 0.93 | 3.0E-07 | 4.0E-03 |  |  |  |  |  |
| M26 Neddylation | *APOE* | 0.98 | 0.94 | 1.03 | 0.43 | 0.71 |  |  |  |  |  |
| M34 Mitochondrion | No *APOE* | 1.10 | 1.06 | 1.14 | 2.1E-06 | 5.0E-03 |  |  |  |  |  |
| M34 Mitochondrion | *APOE* | 0.99 | 0.95 | 1.04 | 0.75 | 0.88 |  |  |  |  |  |
| M18 Ambiguous | No *APOE* | 1.10 | 1.05 | 1.16 | 9.0E-05 | 0.03 |  |  |  |  |  |
| M18 Ambiguous | *APOE* | 1.12 | 1.06 | 1.17 | 8.4E-06 | 0.02 |  |  |  |  |  |

### AD CSF Network Module Levels are Influenced by Treatment with Atomoxetine

Protein co-expression modules derived from proteomic analysis of CSF can provide insight into different aspects of AD pathophysiology, but whether they can be responsive to therapeutic intervention and potentially serve as biomarkers in clinical trials and clinical practice is unknown. To address this question, we analyzed CSF from a recently described phase 2 clinical trial of the norepinephrine reuptake inhibitor atomoxetine (ATX) in individuals with mild cognitive impairment (MCI) due to AD(*27*). Norepinephrine tone in brain is decreased in AD through dysfunction of the locus coeruleus, one of the first brain structures affected in AD(*28, 29*). Increasing brain norepinephrine levels has positive effects in preclinical AD models(*30*), and was shown to have a beneficial effect on CSF tTau and pTau181 levels and brain metabolic function as assessed by [¹⁸F]fluorodeoxyglucose positron emission tomography (FDG-PET) after 6 months of ATX treatment in the phase 2 clinical trial. The trial had a cross-over design with a run-in placebo arm and an ATX washout arm. We analyzed the CSF proteomes of each participant with TMT-MS and SomaScan before and after treatment with ATX and generated a ratio of the ATX-treated sample to the within-subject placebo or baseline sample. We compared this within-subject ATX ratio to a within-subject placebo to baseline ratio to gauge treatment effects due to ATX (**Figure 6A**). We observed four modules that were significantly influenced by ATX treatment in the trial cohort: M20 Glycolysis/Redox homeostasis, M21 extracelllular matrix (ECM)/Vasculature, M14 Translation, and M16 Ambiguous (**Supplementary Table 14, Extended Data**). Given the strong association of M20 Glycolysis/Redox homeostasis with AD endophenotypes and the plausible connection of M21 ECM/vasculature to potential effects on vasculature physiology with norepinephrine, we focused further analyses on these two modules. The M20 Glycolysis/Redox homeostasis module eigenprotein, representing *n*=65 protein measurements, was strongly elevated in AD (**Figure 6B**). Levels of M20 were significantly reduced in the clinical trial after 6 months of ATX treatment (**Figure 6C**). Individuals with higher CSF levels of M20 prior to treatment with ATX generally showed a larger decrease in M20 levels after ATX treatment (**Figure 6D**). One individual had very high levels of M20 at baseline and responded strongly to ATX; however, the correlation remained significant when using a correlation measure, bicor, that is insensitive to outliers. The M21 ECM/Vasculature module was also elevated in the AD network, but in contrast to M20, ATX treatment led to an increase in the levels of this module (**Figure 6E, F**). Pre-treatment levels of M21 were not correlated with an ATX treatment response (**Figure 6G**). In an exploratory analysis, we assessed whether we might be able to improve prediction of M20 treatment response beyond that afforded by baseline M20 levels with a ratio of proteins that were individually correlated with M20 response (**Supplementary Figure 5**). After a combinatorial search for correlation predictors (**Supplementary Figure 5A**), we found that a ratio of summed normalized abundance of four proteins positively correlated to M20 response (UBE2L3, ubiquitin-conjugating enzyme E2 L3; CLIC1, chloride intracellular channel protein 1; BST1, ADP-ribosyl cyclase/cyclic ADP-ribose hydrolase 2; and FBP1, fructose-1,6-bisphosphatase 1) divided by a summed normalized abundance of four negatively correlated to M20 response (IL9, interleukin-9; BPIFA2, BPI fold-containing family A member 2; TXNDC5, thioredoxin domain-containing protein 5; and B4GALT1, beta-1,4-galactosyltransferase 1) increased the prediction correlation to –0.86 (**Supplementary Figure 5B**). As expected, all eight proteins comprising the ratio varied significantly by M20 response (**Supplementary Figure 5C**), although none of these proteins was present within M20. In summary, we observed that panels of CSF proteins defined by their co-expression and representing different biological processes affected in AD changed with ATX treatment—a potential disease-modifying medication for AD—and in the case of M20, this response could partially be predicted by baseline M20 levels or an M20-responsive protein ratio.

**Figure 6.**
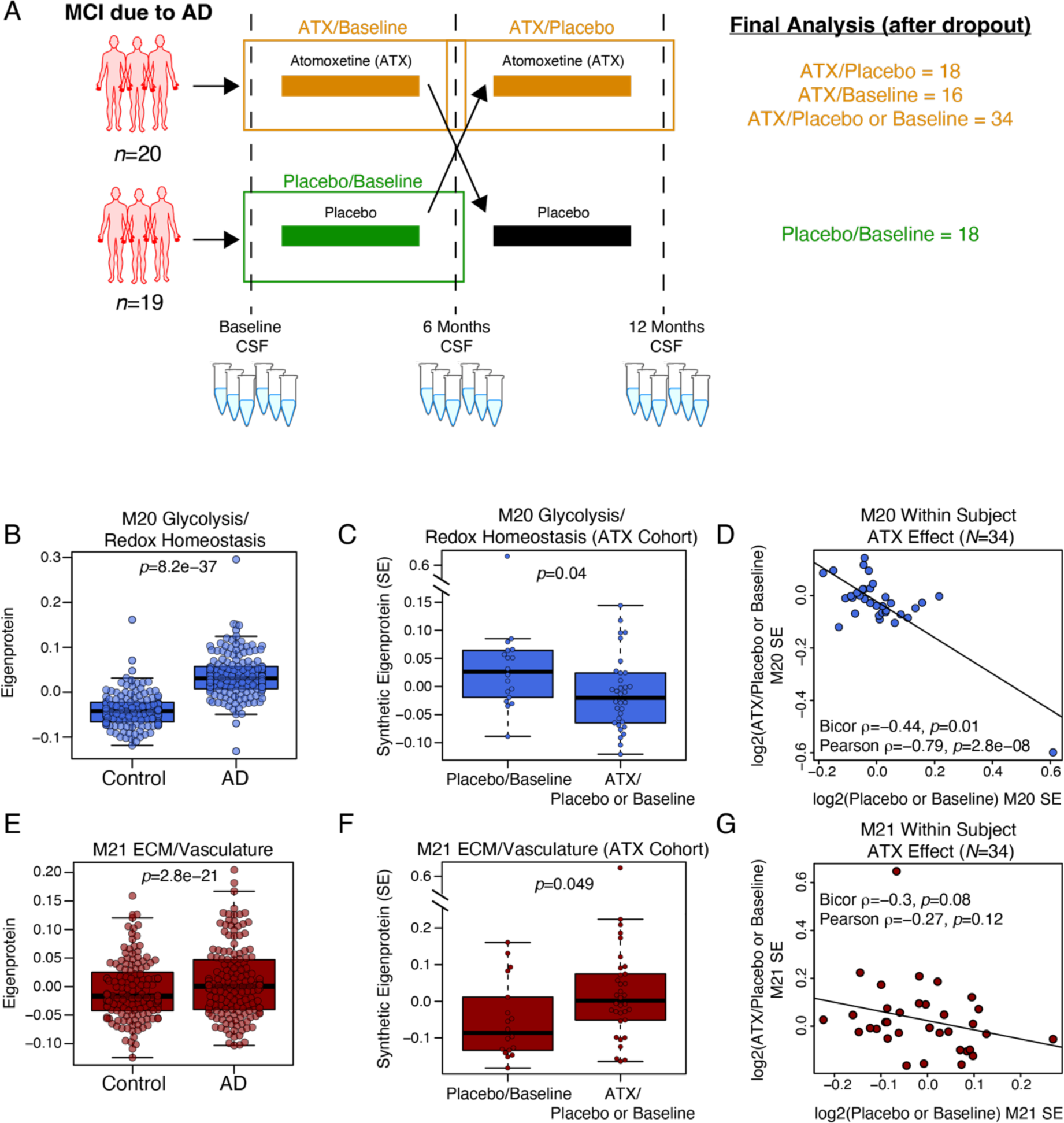
AD CSF Network Modules Influenced by Treatment with Atomoxetine. (A) Scheme for atomoxetine (ATX) trial design. All participants had MCI due to AD and baseline CSF sampling. One arm (*n*=20) was treated initially with ATX for 6 months, then moved to a washout phase, while the other arm (*n*=19) was treated initially with placebo for 6 months and then moved to ATX treatment. The total trial length was 12 months, with CSF sampling at baseline, 6 months, and 12 months. After dropout, a total of 34 ATX samples were compared to 18 ATX-naïve samples, using ratios to baseline or placebo to assess within-subject treatment effects. The ATX washout samples were not analyzed. (B) M20 Glycolysis/Redox Homeostasis eigenprotein levels in control and AD groups in the CSF AD network demonstrating increased M20 levels in AD. (C) M20 levels after ATX treatment compared to ATX-naïve samples in the ATX trial cohort. (D) Correlation of M20 response to ATX treatment with baseline or placebo levels of M20. (E) M21 ECM/vasculature eigenprotein levels in control and AD groups in the CSF AD network demonstrating increased M21 levels in AD. (F) M21 levels after ATX treatment compared to ATX-naïve samples in the ATX trial cohort. (G) Correlation of M21 response to ATX treatment with baseline or placebo levels of M21. ATX treatment effects for all CSF network modules are provided in **Supplementary Table 14 and Extended Data**. Differences between groups were assessed by *t* test. Correlations were performed using bicor and Pearson test.

### Clustering Participants on their CSF Proteomes Identifies Pathological Heterogeneity Not Reflected by Aβ and Tau

We identified many CSF network modules that were correlated to CSF Aβ and tau levels and were increased in AD as defined by CSF Aβ and tau levels. To what extent participants were similar or dissimilar in their CSF proteomic network profile and how such similarity was related to their disease classification based on Aβ and tau and response to ATX was unclear. To address this question, we identified the top ten proteins by correlation to the module eigenprotein (i.e., hub proteins) for each network module and used these proteins to cluster participants into groups based on their similarity across these proteins with the same clustering algorithm used to define the protein modules (**Figure 7A)**. This clustering approach yielded ten different groups of participants based on their CSF AD proteomic network features (**Figure 7B**). The ten groups could be further grouped into three general hierarchical clusters (or superclusters) based on the similarity of their CSF proteomes. The ten groups could also be identified using a separate data dimensionality reduction technique called uniform manifold approximation and projection (UMAP), although the separation of participant groups was not as distinct as the separation of protein groups with UMAP (**Supplementary Figure 6A, B**). Out of the ten groups, only one group (group 10) had uniform membership based on Aβ and tau diagnostic criteria. Groups 3 and 6 were most closely related to group 10 but differed in key proteome features such as higher levels of neuronal modules M8 Semaphorin signaling, M15 Neurexin/Synaptic membrane, M24 Axolemma/Semaphorin complex, and M11 Axonogenesis. Group 6 was different from groups 3 and 10 in the same supercluster based on higher levels of *APO*E-related modules M33 Oxidant detoxification/MAPK signaling and M26 Neddylation and lower levels of M34 Mitochondrion. Groups 1, 2, and 7 were related in the same supercluster and also contained a mixture of AD and control cases based on Aβ and tau criteria, but differed in many key CSF proteome features including lower levels of the neuronal modules mentioned above, lower levels of M4 Autophagy/Ubiquitination, and higher levels of M2 Complement/Coagulation. The third supercluster comprised of groups 4, 5, 8, and 9 contained two groups with nearly all control cases as defined by Aβ and tau (groups 4 and 5), but also a group that had a high proportion of AD cases (group 8). One feature that distinguished this “AD-like” group from the other “control-like” groups in the supercluster was elevated levels of the M20 Glycolysis/Redox homeostasis module. To test whether membership in any of the data-driven participant clusters might be associated with the M20 response to ATX treatment, we divided the 34 AD participants that were treated with ATX into tertiles based on the degree of post-ATX reduction in M20 Glycolysis/Redox homeostasis levels, and projected their baseline (i.e., prior to ATX treatment) proteomes onto the groups using UMAP (**Supplementary Figure 6C**). To simplify the analysis and visualization, we considered only groups that had minimal representation of AD-like cases, which excluded groups 4, 5, and 9. Multi-dimension proteome feature projection showed that none of the ATX tertiles cleanly projected to a group, suggesting that the clustering of participants by CSF proteomic features did not identify a category of individuals who could be predicted to respond to ATX based on their baseline CSF proteome. In summary, we identified 10 groups of participants related by similarity in their CSF proteomes. Although the best responders to ATX treatment did not cluster within one of the groups, the clustering suggested that CSF proteome features significantly varied within AD and control diagnostic classifications defined by Aβ and tau levels.

**Figure 7.**
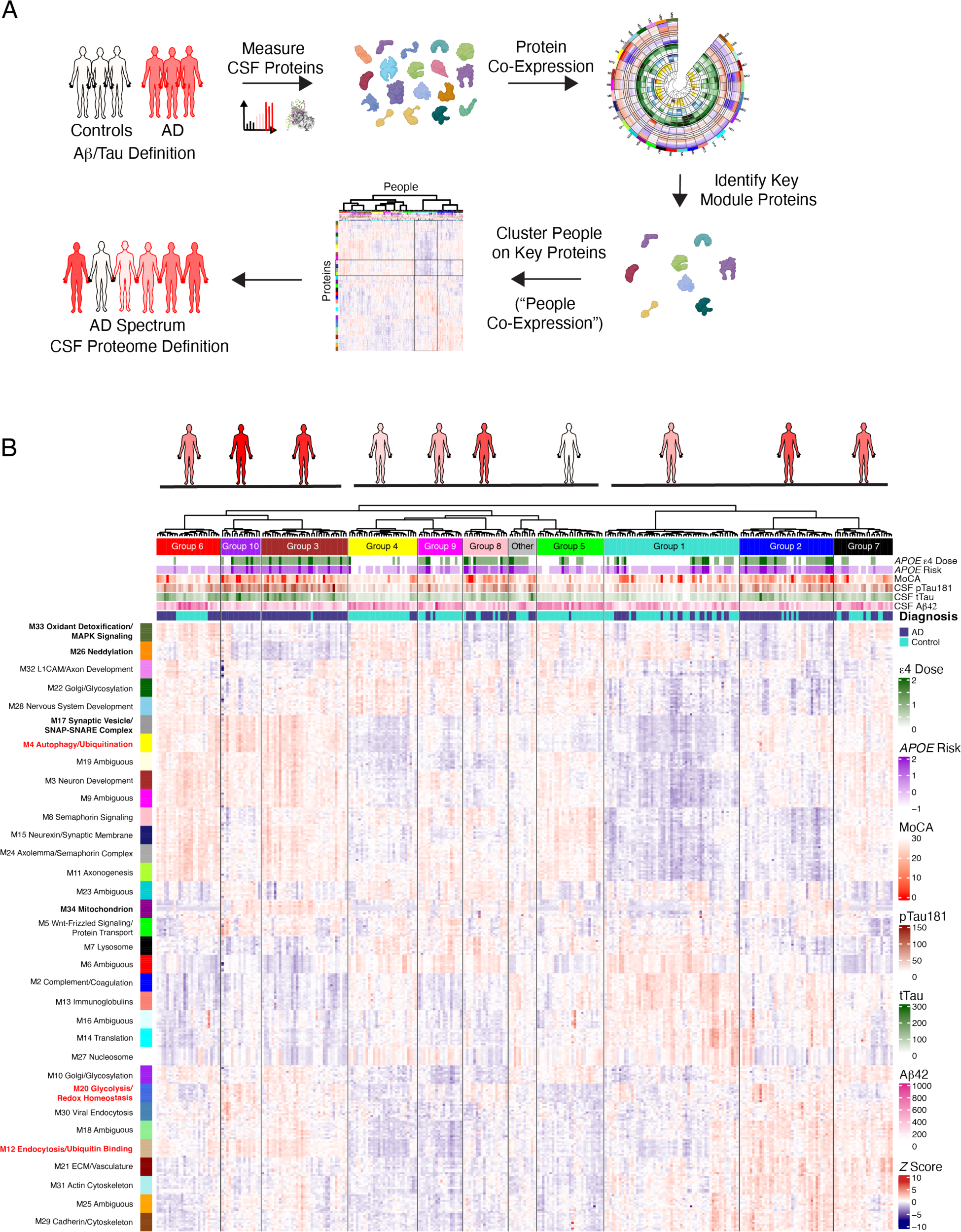
Grouping Individuals Based on CSF AD Proteome Network Features. (A) Scheme for defining groups of people based on their proteomes. CSF protein levels were measured by SomaScan and TMT-MS and used to construct a protein co-expression network. The top ten hub proteins from each module were then used to cluster individuals using the same hierarchical clustering algorithm used to construct the protein co-expression network, defining groups of people based on similarity across CSF network hub proteins. (B) *z* scored relative protein abundance heatmap of the top ten hub proteins in each CSF network module, with individuals (*n*=296) grouped into ten clusters based on the similarity across hub proteins. Figures at the top represent each group, with underlines highlighting the three superclusters. Individuals are shaded based on the number of AD cases within the group as defined by CSF Aβ and tau levels. In addition to diagnostic category, measures of *APOE* ε4 allele number (ε4 Dose; 0, 1, or 2), *APOE* risk (–1, ε2/3; 0, ε3/3, 1, ε3/4; 2, ε4/4), Montreal Cognitive Assessment (MoCA, higher scores indicate better cognitive function), CSF pTau181, CSF tTau, and CSF Aβ_1–42_ are provided for each participant.

## Discussion

In this study we used multi-platform proteomics in AD CSF to identify and measure pathological processes other than Aβ and tau dyshomeostasis associated with AD. Three co-expression clusters (or modules) of proteins associated with oxidant detoxification/MAPK signaling, neddylation, and mitochondrial biology were strongly associated with the *APOE* ε4 AD risk genotype, and altered blood levels of these modules were associated with the development of dementia up to 21 years later. We demonstrated that treatment with atomoxetine in the MCI stage of AD reduced pathological elevation in the M20 Glycolysis/Redox homeostasis module in CSF, suggesting that unbiased data-driven techniques can be used to develop biomarkers that reflect different treatment-responsive AD pathophysiologies. Using CSF proteomic network features, we defined 10 different groups of participants that did not cleanly separate according to a CSF total tau to Aβ ratio classification, highlighting the utility of multi-dimensional techniques for assessment of disease state or type. Refinement of such an approach will likely enable advancement of precision medicine therapeutic approaches for AD.

Our CSF AD network generated from 300 participants showed a high degree of overlap with a previous pilot AD CSF network generated from 36 participants in which we also included a smaller number of protein measurements from the Olink platform, illustrating the robustness of the network approach to define AD pathophysiology in AD CSF. Here the more highly powered network allowed us to observe modules related to *APOE* ε4 that were absent in the pilot network. These modules were composed exclusively of SomaScan measurements. It is possible that these modules could be captured by MS or Olink approaches with sufficient depth of coverage. Indeed, mitochondrial biology in the AD CSF proteome has previously been captured by extensive pre-fractionation and TMT-MS analysis(*31*), but such an approach is not feasible for analysis of large cohorts. The M20 Glycolysis/Redox homeostasis module was the module most strongly correlated to cognitive function in the network and was composed of mostly TMT-MS measurements. As proteomic technologies advance in coverage depth and throughput, additional AD pathophysiology will undoubtedly be revealed in CSF. A multi-platform analytical approach is therefore useful to leverage the benefits provided by any single platform, as well as to increase robustness of study findings.

The M4 Autophagy/Ubiquitination module was the module most strongly correlated to AD biomarkers in the network, with a correlation of *r*=0.86 to tTau, suggesting that the proteostasis signature in CSF is strongly associated with tau pathology. Although we did not have tau-PET measures for these participants, it is possible that this signature is correlated with tau neurofibrillary tangle (NFT) levels in brain. Future studies that include tau-PET as a trait will be informative for assessing which CSF modules are most strongly correlated to this hallmark AD pathophysiology. Interestingly, M4 contained proteins that were recently observed to be changed in autosomal dominant AD (ADAD) CSF 20 to 30 years prior to the onset of cognitive symptoms, including SPARC-related modular calcium-binding protein 1 (SMOC1), 14-3-3 protein zeta (YWHAZ),14-3-3 protein gamma (YWHAG), and pyruvate kinase PKM (PKM)(*32*). Elevation in SMOC1 levels occurred concomitantly with the formation of Aβ plaques and prior to elevations in pTau or Aβ PET markers. Markers of proteostatic stress response were also elevated early in the ADAD study. While longitudinal data in CSF were not available in the present study, the findings in ADAD suggest that an alteration in proteostasis—either through an increased compensatory response or failure of the response—is an early observable AD pathological change in CSF that may precede NFT formation.

Direction of change in CSF modules with development of AD is not always consistent with the direction of change for the analogous modules in brain. While disease-related modules such as M20 Glycolysis/Redox homeostasis and M11 Axonogenesis are increased and decreased, respectively, in CSF concordant with previously observed changes in glycolysis and neuronal biology in AD brain(*2, 3*), other modules such as M2 Complement/Coagulation, M17 Synaptic vesicle/SNAP-SNARE complex, M33 Oxidant detoxification/MAPK signaling and M34 Mitochondrion show opposite direction of change compared to brain. These findings suggest that protein biomarkers for AD pathological processes may depend on the compartment in which they are measured, as previously suggested(*10*). In the case of the complement module which is highly conserved across compartments, levels are elevated in both blood and brain but decreased in CSF, perhaps suggesting deposition of complement in brain tissue with subsequent reduced levels in CSF analogous to the behavior of Aβ_1–42_.

We observed three CSF modules that were highly correlated to *APOE* ε4—M26 Neddylation, M33 Oxidant detoxification/MAPK signaling, and M34 Mitochondrion. M34 Mitochondrion was strongly positively correlated to *APOE* ε4 and demonstrated increased levels in AD CSF, whereas the other two modules were negatively correlated to *APOE* ε4 and demonstrated decreased levels in AD CSF. The associations with *APOE* ε4 were also present in the same direction when these modules were analyzed in blood in two different cohorts. Furthermore, altered levels of these modules in blood were associated with risk of developing AD dementia over two decades prior to diagnosis. Whether these modules represent systemic changes observable in both blood and CSF that influence brain health, or whether they are markers of primary brain pathophysiology that develops decades prior to symptom onset is unclear. It has previously been shown that *APOE* ε4 is associated with reduced brain metabolic function in early and midlife(*33, 34*). *APOE* ε4 fragments have also been shown to be inhibitors of mitochondrial function in neurons(*35, 36*). These observations potentially suggest a brain-derived origin of the M34 Mitochondrion signature. Neddylation is a process closely related to ubiquitination through conjugation of the ubiquitin-like protein NEDD8 to proteins via E3 ligases, and has been shown to be involved in proteostasis and maintaining synaptic integrity(*20, 37*). Multiple intracellular protein aggregates and inclusion bodies observed in different neurodegenerative diseaseas are neddylated(*21*), which could support a primary brain origin for the M26 neddylation module. However, changes in neddylation could also reflect a general systemic failure in proteostasis. Recently, changes in blood levels of proteins such as growth/differentiation factor 15 (GDF15) that have no known expression in brain have been associated with all-cause dementia up to 25 years later(*38*), highlighting the possibility that the *APOE* ε4-associated modules observed in this study may simply reflect a systemic process observable in both blood and CSF that influences AD risk. Additional studies that clarify the tissue source and time-resolved effects of the observed *APOE* ε4-associated modules are needed.

We leveraged our AD CSF network to interrogate the effects of atomoxetine treatment on the AD CSF proteome. We found significant drug treatment effects at the module level, including reduction in the M20 Glycolysis/Redox homeostasis module. This module showed the strongest correlation to cognitive function in our study. While a significant cognitive benefit was not observed in the short phase 2 clinical trial of ATX, this observation is consistent with other beneficial outcomes observed in the trial such as reduced CSF levels of tTau and pTau(*27*). Notably, M20 was correlated to both tTau and pTau levels in our network. This type of data-driven approach to biomarker development for different AD pathophysiologies may be a promising way to understand drug treatment effects on other pathways beyond Aβ and tau.

In an effort towards developing precision-medicine approaches that leverage patient-specific proteomic information, we explored the ability to predict response to ATX treatment at three different levels: individual protein, module, and network cluster. At the individual protein level, the strongest baseline measurement correlation to M20 treatment response was with Ubiquitin-conjugating enzyme E2 L3 (UBE2L3) with a correlation of *r*=–0.6. This was slightly better than module level prediction, where baseline levels of M20 correlated with treatment response at *r*=– 0.44. We were able to identify a combination of 8 proteins that could increase this correlation to *r*=–0.86. While this discovery approach to protein biomarker combination may suffer from overdetermination due to the large number of proteins tested and the small number of patients in the clinical trial, further validation could be obtained in future phase 3 trials of ATX. Because none of the 8 proteins was a member of the M20 module, we decided to use the entire CSF network to potentially identify a category of patient who might best respond to ATX treatment considering reduction in M20 levels as the outcome measure. We identified 10 different potential groups of participants through clustering on AD CSF network features, and although none of these groups clearly captured those who best responded to ATX, this type of approach may be a way to identify responders to other treatments for AD and for other neurodegenerative diseases.

Recent studies have used TMT-MS proteomics in CSF to subtype patients into three or five groups(*39, 40*). In these studies, AD was defined on the basis of CSF Aβ levels only and proteins that were differentially expressed in the AD group were used to cluster subjects within the AD defined group using non-negative matrix factorization. In our study we used the CSF tTau/Aβ_1–42_ ratio to classify participants into control and AD groups regardless of cognitive status, and included control participants in the unbiased clustering approach using WGCNA on the top ten hub proteins from each CSF network module. We found that the ten groups of participants did not cleanly stratify on AD and control diagnoses using Aβ and tau, suggesting that disease classification or staging based on the CSF proteome captures more heterogeneity than these classical markers. The supercluster consisting of groups 3, 6, and 10 had a similar proteomic profile consistent with what might be considered “classic” AD with low Aβ and high tTau and pTau, and cognitive impairment. While all three groups had similar elevations in M4 Autophagy/Ubiquitination and M17 Synaptic vesicle/SNAP-SNARE complex modules, the groups were resolved largely based on *APOE* ε4-related module levels and a lack of increase in some neuronal modules in group 10 (M8 semaphorin signaling, M15 neurexin/synaptic membrane, M24 axolemma/semaphorin complex, and M11 axonogenesis). Fascinatingly, control participants in group 6 had a similar proteome profile to the AD participants but with generally higher levels of Aβ, lower levels of tTau and pTau, and lower levels of M20 Glycolysis/Redox homeostasis, suggesting a potential “resilient” phenotype or an early disease stage in which observable disease changes occur before significant changes in Aβ and tau. Another supercluster consisting of groups 1, 2, and 7 contained a mix of AD and control participants with group 1 demonstrating low M4 Autophagy/Ubiquitination levels and low neuronal markers, yet elevated M2 Complement/Coagulation and M13 Immunoglobulin modules perhaps suggesting increased blood-brain barrier permeability and/or inflammation. Groups 2 and 7 demonstrated similar elevations in M21 ECM/Vasculature, M31 Actin cytoskeleton, M25 Ambiguous, and M29 Cadherin/Cytoskeleton, reflecting a general elevation in structural proteins. The disease phenotype this profile represents is unclear but may be related to a proposed subtype of AD related to changes in choroid plexus biology(*39*). The type of analysis presented here clearly illustrates the heterogeneity of CSF proteomic changes that do not necessarily correlate with Aβ and tau levels. Future work on longitudinal datasets will be required to resolve to what extent these profiles reflect disease subtypes *versus* disease stage.

In summary, multidimensional profiling of the AD CSF proteome as illustrated in this study is a promising approach to further our understanding of AD pathophysiology and develop biomarkers for this varied pathophysiology. Such an approach holds promise to advance precision medicine approaches for AD.

## Methods

### Participants and Case Classification

All CSF samples used in this study were collected under the auspices of the Emory Goizueta Alzheimer’s Disease Research Center (ADRC) and Emory Healthy Brain Study (EHBS). The cohort consisted of 140 healthy controls and 160 patients with AD as defined by the NIA research framework(*16*). Basic demographic data were obtained from the Goizueta ADRC and EHBS. Control and AD participants received standardized cognitive assessments in the Emory Cognitive Neurology Clinic, Goizueta ADRC, or EHBS. CSF was collected and banked according to the 2014 National Institute on Aging best practice guidelines for Alzheimer’s Disease Centers (https://alz.washington.edu/BiospecimenTaskForce.html). CSF samples were subjected to ELISA Aβ_1–42_, total tau, and pTau181 analysis by the INNO-BIA AlzBio3 Luminex Assay(*41*). ELISA values were used to support diagnostic classification based on established AD biomarker cutoff criteria(*42, 43*). We derived a cutoff tTau/Aβ_1–42_ ratio of 0.182 as the classification threshold for AD based on a Gaussian mixture model analysis of all research participants analyzed on the AlzBio3 platform at our center. The ratio of 0.182 best separated the two distinct populations observed for this ratio as measured by the AlzBio3 assay. *APOE* genotype was determined by extracting DNA from the plasma buffy using the GenePure kit (Qiagen) following the manufacturer’s recommended protocol, then determining the rs7412 and rs429358 genotypes using either an Affymetrix Precision Medicine Array (Affymetrix) or TaqMan assays (ThermoFisher Scientific C_904973_10 and C_3084793_20). *APOE* genotype was confirmed by assessing the *APOE* proteotype in all samples. For any samples that were discrepant between genotype and proteotype, the proteotype was used. All samples were analyzed by both TMT-MS and SomaScan 7k platform assays. All Emory research participants provided informed consent under protocols approved by the Institutional Review Board at Emory University. Case metadata are provided in **Supplementary Table 1.**

### Quantification of Proteins by SomaLogic SomaScan Modified Aptamers

Proteins were quantified by SomaScan as previously described(*44-46*). Aliquots of CSF and plasma from each subject were sent to SomaLogic (SomaLogic, Boulder, CO) for analysis using the modified aptamer SomaScan assay (v4.1). All samples passed quality control measures and were randomized by SomaLogic prior to analysis on single plates. Results were reported as relative fluorescence units (RFUs) for relative quantification of protein abundance.

### CSF Protein Preparation and Digestion for Tandem Mass Tag Mass Spectrometry (TMT-MS) Analysis

Equal volumes (50 μL of each sample) of CSF were digested with lysyl endopeptidase (LysC, Wako 125-05061) and trypsin (ThermoFisher Scientific 90058). Briefly, each sample was reduced and alkylated with 1 μL of 0.5 M tris-2(-carboxyethyl)-phosphine (TCEP) and 5 μL of 0.4 M chloroacetamide (CAA) at 90°C for 10 min, followed by water bath sonication for 5 min. The same volume of 8 M urea buffer [56 μL, 8 M urea in 10 mM Tris, 100 mM NaH_2_PO_4_ (pH 8.5)] was added to each sample after cooling the samples to room temperature, along with LysC (2.5 μg). After overnight digestion, 336 μL of 50 mM ammonium bicarbonate (ABC) was added to each sample to dilute the urea concentration to 1 M, along with trypsin (5 μg). After 12 hours, the trypsin digestion was stopped by adding final concentration of 1% formic acid (FA) and 0.1% trifluoroacetic acid (TFA).

### Isobaric TMT Peptide Labeling

Before TMT labeling, the digested peptides were desalted using 30 mg HLB columns (Waters). Briefly, the columns were activated with 1 mL of methanol, then equilibrated with 2 × 1 mL 0.1% TFA. The acidified samples were loaded following by washing with 2 × 1 mL 0.1% TFA. Elution was performed with 2 × 0.5 mL 50% acetonitrile. To normalize protein quantification across batches, global internal standard (GIS) samples were generated for each sample set by combining 100 μL aliquots from each sample elution. All individual samples and GIS pooled standards were dried by speed vacuum (Labconco). The TMT 16-plex kit (ThermoFisher Scientific, A44520, lot number VH311511) was used for labeling, which divided CSF sample sets into 22 TMT batches with 13 or 14 samples plus 2 GIS in each batch. The sample and channel distribution are provided in **Supplementary Table 1**. 5 mg of each channel reagent was dissolved in 200 μL anhydrous acetonitrile. Each CSF peptide sample was resuspended in 50 μL of 100 mM TEAB buffer, and 10 μL of TMT reagent solution was subsequently added. The labeling was stopped after 1 h with 3 μL of 5% hydroxylamine for CSF and the peptide solutions were then combined according to the batch arrangement. The combined TMT samples were desalted with 100 mg of Sep-Pak C18 columns. The elutions were dried under speed vacuum.

### High-pH Off-line Fractionation

Dried samples were re-suspended in high pH loading buffer (0.07% vol/vol NH_4_OH, 0.045% vol/vol FA, 2% vol/vol ACN) and loaded onto a Water’s BEH column (2.1 mm x 150 mm with 1.7 μm particles). A Vanquish UPLC system (ThermoFisher Scientific) was used to carry out the fractionation. Solvent A consisted of 0.0175% (vol/vol) NH_4_OH, 0.01125% (vol/vol) FA, and 2% (vol/vol) ACN; solvent B consisted of 0.0175% (vol/vol) NH_4_OH, 0.01125% (vol/vol) FA, and 90% (vol/vol) ACN. The sample elution was performed over a 25 min gradient with a flow rate of 0.6 mL/min with a gradient from 0 to 50% solvent B. A total of 96 individual equal volume fractions were collected across the gradient. Fractions were concatenated to 48 and dried to completeness using vacuum centrifugation.

### TMT Mass Spectrometry

All fractions (∼1μg) were loaded and eluted using a EasyNLC 1200 (ThermoFisher Scientific) on an in-house packed 25 cm, 100 μm internal diameter (i.d.) capillary column with 1.9 μm Reprosil-Pur C18 beads (Dr. Maisch, Ammerbuch, Germany) over a 35 min gradient from 1% to 99% buffer B (80 ACN with 0.1% FA). Mass spectrometry was performed with a high-field asymmetric waveform ion mobility spectrometry (FAIMS) Pro-equipped Orbitrap Lumos (ThermoFisher Scientific) in positive ion mode using data-dependent acquisition with 1 s top speed cycles. Each cycle consisted of one full MS scan followed by as many MS/MS events that could fit within the given 2 s cycle time limit. MS scans were collected at a resolution of 120,000 (410-1600 m/z range, 4x10^5^ AGC, 50 ms maximum ion injection time, FAIMS compensation voltage of –45 and –65). All higher energy collision-induced dissociation (HCD) MS/MS spectra were acquired at a resolution of 50,000 (0.7 m/z isolation width, 35% collision energy, 200% normalized AGC target, 86 ms maximum ion time). Dynamic exclusion was set to exclude previously sequenced peaks for 30 s within a 10-ppm isolation window.

### Database Searches and Protein Quantification

All raw files were searched using Proteome Discoverer (version 2.4.1.15, ThermoFisher Scientific) with Sequest HT. The spectra were searched against a human UniProt database downloaded April 2015 (90,411 target sequences). Search parameters included 10 ppm precursor mass window, 0.05 Da product mass window, dynamic modifications methionine (+15.995 Da), deamidated asparagine and glutamine (+0.984 Da), phosphorylated serine, threonine and tyrosine (+79.966 Da), and static modifications for carbamidomethyl cysteines (+57.021 Da) and N-terminal and lysine-tagged TMT (+304.207 Da depending on the dataset). Percolator was used to filter peptide spectral matches (PSMs) to 1% FDR. Peptides were grouped using strict parsimony and only razor and unique peptides were used for protein level quantitation. Reporter ions were quantified from MS2 scans using an integration tolerance of 20 ppm with the most confident centroid setting. Only unique and razor (i.e., parsimonious) peptides were considered for quantification.

### Protein Abundance Data Processing SomaLogic SomaScan Assay

The SomaScan .adat file containing relative fluorescence unit (RFU) abundances for the 344 samples including 300 experimental CSF samples, 12 blank buffer (background or noise measurements), calibrator, and QC samples was loaded using the SomaDataIO package v3.1.0 in R v4.0.2. Multidimensional scaling (MDS) indicated that background measures were different in one of the four plates, contributing to noise and increasing the threshold for limit of detection (LODs). We stabilized background measurements which were in excess across the triplicate measures of higher noise in PLT11726 buffer signals by subtraction of the difference of medians between the median buffer signals of background measurements in the three plates with lower background signal and the median background measurement in the plate with higher background signal. LOD was then calculated as the median stabilized background measurement plus three standard deviations (SDs) of all background measurements. After this subtraction, 6,024 assays had at least 25% of values above LOD, whereas only 4,307 assays were above LOD before noise stabilization. Sample measurements in this assay subset that were below LOD were retained but considered as missing values for downstream analyses except where noted. Next, buffer measurements were used to calculate signal-to-noise (S:N) ratios for unnormalized RFU abundance data. S:N ratios were calculated by subtracting the within-assay median buffer signal from the unlogged assay signal (RFU), then dividing by the median buffer signal. We performed two-dimensional MDS on the SomaScan data matrix rows (assays) for all assays with a median maximum S:N (maximum of either control or AD median S:N) above 10. We noted two distinct clusters of cases. Differential protein abundance between the two clusters of cases as calculated by *t* test indicated that nine blood gene product proteins (HBA1, hemoglobin subunit alpha; HBB, hemoglobin subunit beta; HBG1, hemoglobin subunit gamma-1; FGA, fibrinogen alpha chain; FGB, fibrinogen beta chain; FGG, fibrinogen gamma chain; HP, haptoglobin; CAT, catalase; CA1, carbonic anhydrase 1) were the most differentially abundant between the two clusters, suggesting blood contamination. Prior to addressing blood contamination, SomaScan samples were checked for status as sample network connectivity outliers by calculating Z.ku (sample connectivity *z* score) from an adjacency matrix calculation using the available log2 protein abundance matrix. Briefly, a normalized adjacency calculation on this matrix was performed using the Weighted Correlation Network Analysis (WGCNA) (v1.72-1) bicor function with pairwise complete observations only, squared, multiplied by 0.5 and added to 0.5. Then, sample connectivity (ku) was calculated using the WGCNA fundamentalNetworkConcepts function, followed by *z*-transformation before checking Z.ku for outlier status below –3.0 *z* score units (>3 SD below the mean ku). Four samples with Z.ku more than 3 SD below the mean were thereby removed; these are indicated in **Supplementary Table 1**. We then proceeded to calculate the first principal component of the nine-protein blood signature using calculation of a synthetic WGCNA eigenprotein, and performed nonparametric bootstrap regression of this signature blood eigenprotein. Following regression, we confirmed that the differences between samples in the prior two clusters were ablated as viewed by PCA, MDS, and histograms of a recalculated blood signature.

Because SomaScan CSF data had low signal, a second-pass filter step was applied to remove assays that did not meet an empirically-derived S:N threshold. This threshold was determined by correlating SomaScan assays with TMT-MS assays at varying S:N cutoff values (0, 0.15, 0.25, 0.3, 0.35, 0.4, 0.45, 0.50, 0.625, 0.75, 1, 2, 4, and 8), and selecting the S:N value (0.3) that maximized median Pearson correlation with proteins measured in common between SomaScan and TMT-MS. After application of first- and second-pass filters and removal of control aptamers, 4098 CSF human SomaScan assays were retained for subsequent analyses.

### TMT-MS

Only proteins that were identified and summarized as high confidence (<1% FDR) by Proteome Discoverer (PD) were used for analysis. The 3,870 UniProt protein identifier accessions provided by PD were further annotated with Hugo Gene Nomenclature Committee (HGNC) official gene symbols. TMT reporter intensities (abundances) that had not undergone normalization by PD were used for analysis to preserve inherent protein abundance differences between control and AD subjects. Batch correction was performed by dividing abundances for each protein within each batch by the GIS. GIS measurements were then removed, and proteins with more than 75 percent (*n* > 225/300) missing values were excluded from consideration. The number of remaining protein isoforms after missing value control was 2334.

The nine blood-associated proteins which separated samples in SomaScan data prior to regression were checked in the resulting TMT-MS protein log_2_ relative abundance matrix by two-dimensional MDS. Separation (77 high, 223 low blood signature) was found, and further refined as a five-protein subset signature in the MS data (HP, HBB, HBG1, CAT, and CA1). To harmonize cases with the SomaScan data, TMT-MS samples matching the four SomaScan outliers detected by low connectivity Z scores as noted above were likewise removed from consideration prior to correction of blood contamination. The first principal component of the five-protein signature was then regressed using nonparametric bootstrap regression as performed in the SomaScan data. MDS following regression found no further clustering of sample subsets.

### Proteome Coverage Overlap, Ontology Enrichment, and Missing Data Analysis

Unique gene symbols measured in each platform were counted, and overlap was visualized using the venneuler R package (v1.1-0) venneuler function. Visualization of overlap of differentially expressed proteins between the two platforms used the same function. Enrichment of gene ontologies (GO) in different Venn categories was calculated as a Fisher exact test *p* value transformed to *z* score using GOparallel (https://www.github.com/edammer/GOparallel). GOparallel implements piano R package functions and downloads monthly updated gene sets curated by the Bader Lab (at https://baderlab.org/GeneSets as described in Reimand *et al*. (*47*)), and produces visualizations of the output. The same procedure was used to determine ontology enrichment for network modules (Extended Data). Regarding missing data (**Supplementary Figure 2**), SomaScan assays included assay measurements below LOD for the network analysis, but these values were censored as missing for differential abundance determination (volcano analysis). Missing data in 2334 TMT-MS isoforms was considered at the level of batch, as all measurements within a batch result from the same MS/MS fragmentation. All 2334 isoforms were subject to volcano analysis, but no isoforms with any missing data were considered in the 1144 isoforms of TMT-MS data merged with 4098 SOMAScan assays for network analysis.

### Protein Abundance Correlation Analysis

Proteins measured in common between the two platforms within the same biofluid were correlated across all samples using the corAndPvalue function in the WGCNA R package (v1.72-1) (**Supplementary Table 4, Extended Data**). In the case of multiple SomaScan assays for the same protein, the assay with the identical UniProt protein accession, or secondarily, a SOMAmer measuring an identical gene product, was selected. When multiple cross-platform UniProt accession or gene symbol matches occurred, the SOMAmer with the highest correlation was selected. We constructed a population histogram of all Pearson correlations for distinct gene products or UniProt accessions (representing distinct protein isoforms) and identified the median rho for each population of paired measurements between the two platforms.

### Differential Expression Analysis

Differences between AD and control were assessed on the log_2_(abundance) measurements over all proteins after data processing as described above, which included signal cleanup, filtering on missingness, and, in the case of SomaScan CSF data, control of excessively low S:N assays. SomaScan assay values below LOD were considered as missing. Volcano plots and the underlying stats were calculated using a custom in house script (https://www.github.com/edammer/parANOVA) and plotted as volcanoes via the plotly (v4.9.2.1) R package function ggplotly. Individual volcano points were colored by membership in the 44 brain network modules described in Johnson *et al*.(*2*). A stacked circular barplot depicting total protein coverage in brain by module and coverage of differentially expressed proteins in each platform was rendered using the R package ggplot2.

### Harmonization of Platform Protein Abundance Prior to Network Analysis

TMT-MS ion counts in neat CSF with no missing values across the 22 TMT batches (*n*=1,144), and SomaScan RFUs (*n*=4,098 assays), totaling 5,242 assays in CSF, were assembled for 296 case samples measured on both platforms. Only truly missing values were considered as unavailable; values below LOD were retained, although assays removed due to S:N filtering were excluded. Data were transposed prior to removal of platform-specific effects as a batch effect using the TAMPOR algorithm. Proteins were considered as samples (columns) and samples as rows for the two-way table median polish of ratio using TAMPOR. Common proteins measured across both platforms were used as the GIS (*n*=452) to calculate the central tendency of data within and across platforms used for the denominators in the TAMPOR algorithm, as previously described(*48*). Normalized data used in subsequent network analyses was of the form log_2_(abundance/central tendency) of the common proteins in all platforms. No protein assay had any missing values.

### Protein Co-Expression Network Analysis

A CSF proteome network was constructed using the harmonized protein abundances. The Weighted Correlation Network Analysis (WGCNA) algorithm (v1.72-1) was used for network generation. No outliers were detected using the WGCNA sample network connectivity outlier algorithm. The WGCNA blockwiseModules function was run on the CSF harmonized abundances with the following parameters: power=14, deepSplit=4, minModuleSize=10, mergeCutHeight=0.07, TOMdenom=”mean”, bicor correlation, signed network type, PAM staging and PAM respects dendro as TRUE, and a maxBlockSize larger than the total number of protein assays. Module memberships were then iteratively reassigned to enforce kME table consistency, as previously described(*2*). The resulting network assignments were visualized as modules using the iGraph(v1.4.1) package using a custom implementation available as the buildIgraphs function at https://www.github.com/edammer/netOps. Module eigenprotein correlations and significance were visualized in circular heatmaps using the circlize(v0.4.15), dendextend(v1.17.1), and dendsort(0.3.4) R packages. Synthetic eigenproteins for brain and CSF networks were calculated as previously described(*2*) leveraging the top 20 percent of hubs and a minimum overlap of four proteins. For synthetic eigenproteins translated either from brain or into brain, the existing data for 8,619 proteins underlying the brain network were mapped to labels in the CSF network using a mapping rubric to cross-reference protein labels. Specifically, (1) an exact UniProt ID match to that in labels of the form Symbol|UniprotID|platform took precedence for labels with MS as the platform, followed by (2) symbol matches with MS as the platform. This was followed by (3) an exact UniProt ID to a SomaScan row, followed by (4) a symbol match with SomaScan as the platform. In this way, unmatched proteins across any pair of networks were minimized.

### Network Preservation

Pairwise, directional preservation between CSF and plasma, plasma and CSF, and brain to each of the biofluid networks and vice versa was performed using the WGCNA (v1.72-1) modulePreservation function with 500 permutations after harmonizing protein assay labels as described above. The same 4-point rubric described above was used for matching (relabeling) brain network member labels before performing module preservation. Zsummary composite *z* score for 8 underlying network parameters was calculated and visualized by circular heatmap as significance (–log_10_(Benjamini-Hochberg adjusted *p* values), corresponding to the Zsummary scores obtained.

### Cell Type Marker Enrichment Analyses

Cell type-specific enriched marker gene symbol lists were used as previously published to perform a Fisher’s exact one-tailed test for enrichment(*2*). Benjamini-Hochberg correction was applied to all resulting *p* values.

### Network Module Overlap Comparison

Overrepresentation analysis (ORA) was performed to find gene product overlap significance of modules in the current two-platform network with those of the 38-module CSF network published previously using three platforms(*10*), and with a 44-module brain network(*2*). Two-tailed hypergeometric overlap was used to obtain significance (*p*) for each module pair across networks. *P* was further FDR-corrected using the Benjamini-Hochberg method and considered as –log(FDR). External SomaScan data (4,137 human protein targets on a custom platform) for a serum network were obtained from Emilsson *et al*.(*24*). Data on serum network module membership of gene products was used to perform ORA as above for determining overlap with CSF network modules. These measures of overlap significance were visualized using a custom in house script, or with our circular network visualization script. Serum network module ontologies were assigned in consistent fashion with the CSF network module ontologies based on GO analysis.

### Association Analyses in the AGES-Reykjavik Cohort

The Age, Gene/Environment Susceptibility (AGES)-Reykjavik Study cohort is a single-center prospective population-based study of deeply phenotyped subjects (*n*=5,764, mean age 76.6 ± 5.6 years) and survivors of the 40-year-long prospective Reykjavik study, an epidemiologic study to understand aging in the context of gene/environment interaction by focusing on four biologic systems: vascular, neurocognitive (including sensory), musculoskeletal, and body composition/metabolism(*25*). Alzheimer’s diagnosis at AGES-Reykjavik baseline and a 5-year follow-up visit was carried out using a three-step procedure as previously described(*49*). The follow-up time was up to 16.9 years, with the last individual being diagnosed with AD 16 years from baseline. The AGES study was approved by the NBC in Iceland (approval number VSN-00-063), the National Institute on Aging Intramural Institutional Review Board, and the Data Protection Authority in Iceland.

The proteomic measurements in AGES have been described in detail elsewhere(*50*) and were available for 5,457 participants. Briefly, a custom version of the SomaScan platform (Novartis V3-5K) was applied based on the slow-off rate modified aptamer (SOMAmer) protein profiling technology including 4,782 SOMAmers that bind to 4,137 human proteins. Serum was prepared using a standardized protocol(*51*) from blood samples collected after an overnight fast and stored in 0.5 mL aliquots at –80°C. Serum samples that had not been previously thawed were used for the protein measurements. All samples were run as a single set at SomaLogic Inc. (Boulder, CO, US). All SOMAmers that passed quality control had median intra-assay and inter-assay coefficient of variation (CV) < 5% or equivalent to reported variability.

Protein measurement data were centered, scaled and Box-Cox transformed, and extreme outliers excluded as previously described(*50*). Synthetic CSF protein modules were calculated in serum protein data using the moduleEigengenes function from the WGCNA R package(*52*). The synthetic CSF protein modules were limited to the top 20% proteins in each module ranked by kME value except for the two smallest modules (M33 and M34) for which a cutoff of kME >=0.8 was used. The associations of the synthetic CSF protein modules in serum with prevalent (*n*=167) and incident (*n*=655) AD were examined via logistic regression and Cox proportional-hazards model, respectively. All models included adjustment for age and sex, and a secondary model additionally included *APOE* ε4 allele count. Individuals with prevalent non-AD dementia (*n*=163) were excluded from all analyses, and those with prevalent AD were excluded when testing for associations with incident AD. Empirical *p* values were calculated based on results obtained by 1,000 permutations of protein module membership.

### Association Analyses in the ARIC Cohort

The Atherosclerosis Risk in Communities (ARIC) cohort is a community-based study that enrolled 15,792 participants between 1987 and 1989 from Jackson MS; the northwestern suburbs of Minneapolis, MN; Forsyth County, NC; and Washington County, MD. Participants were initially evaluated every three years in-person. Plasma proteomic data in this study were from ARIC visit 2 (*N*=11,596). Institutional review boards approved the study protocols at each participating center. Additional details on the ARIC cohort, dementia ascertainment, and the SomaScan proteomic measurements in ARIC are described elsewhere(*26, 38*). A total of 5,284 SOMAmers were used in the analysis. Association analyses in plasma were conducted in the same fashion as the analyses in serum in AGES. Association of incident dementia with module plasma eigenproteins was performed using Cox regression models. Three models were tested: a model unadjusted for covariates; a model adjusting for demographic variables (baseline age, sex, and race-center); and a model adjusting for demographic variables (baseline age, sex, race-center, education), cardiovascular risk factors (body mass index, diabetes, hypertension, smoking status), and kidney function (eGFR-creatinine). Adjusted models were also tested with and without *APOE* included in the models. *P* values were adjusted using a false discovery rate procedure. Empirical *p* values were also calculated based on results obtained by 1,000 permutations of protein module membership.

### Sample-based Network (sWGCNA) for Sample Clustering/Classification

A sample network classifying the 296 CSF samples into ten modules was constructed using WGCNA (v1.72-1). The WGCNA blockwiseModules function was run on the top ten protein assay hubs of 33 out of the 34 modules in the joint SomaScan-MS CSF protein network, as ranked by intramodule kME (bicor calculation). Due to its large and non-specific nature, module 1 was excluded from the calculation. Samples were assigned to ten modules using the following blockwiseModules function parameters: power=8, deepSplit=2, minModuleSize=10, mergeCutHeight=0.07, TOMdenom=”mean”, bicor correlation, signed network type, PAM staging and PAM respects dendro as TRUE, and a maxBlockSize larger than the total number of samples. Module memberships were then iteratively reassigned to enforce kME table consistency, as previously described(*2*). Sample modules and their organization of top ten protein assay hubs from the prior protein network were then visualized as a heatmap using the ComplexHeatmap package (v2.14.0) pheatmap function, clustering hierarchically using correlation distance only within sample modules.

### UMAP Visualization of Independently Clustered Samples

Python v3.10 was leveraged by the R reticulate package (v1.28) to run UMAP to visualize the similarity of samples in two-dimensional manifold space. Samples were represented by the *Z*-transformed top 10 hub proteins of the same 33 (of 34) protein network modules used for the sample-based network generated using WGCNA as described above. Thus, input was a 330 x 296 matrix. The ExtraTreesClassifier from sklearn.tree was trained on 70 percent of the data, with n_estimators=80, 8 min_samples_leaf, max_features=’auto’, bootstrap=True, and class_weight=’balanced’, and generated leaves to which samples were assigned. These were embedded in low-dimensional space by the UMAP function using a hamming metric. Later, the same process was repeated with only AD samples from AD sample-containing sWGCNA clusters, adding in the Z-transformed abundance profiles for the same hub proteins in the atomoxetine (ATX) drug trial dataset, and restricting from the 330 top 10 hubs of the modules considered the 315 proteins available in both the ATX and 296-sample cohort.

### Atomoxetine Dataset Integration

CSF samples collected longitudinally from patients enrolled in a drug repurposing trial of atomoxetine (ATX) for mild cognitive impairment due to Alzheimer’s disease were sequenced and quantified for proteins both by TMT-MS as previously described(*27*) and SomaScan 7k modalities. Data pre-processing for both TMT-MS and SomaScan ATX datasets was conducted in the same fashion as described above. Briefly, TMT data were batch corrected, and both MS and SomaScan data were regressed to remove blood contamination in a subset of samples. Then, the SomaScan data were double-filtered both for maximum values below LOD (<75%, or at least 22/87 values above LOD), and for S:N achieving the most improved cross-platform correlation with a minimum of protein assays removed, which for the ATX dataset was a ratio of 0.45 (noise-subtracted signal mean) to 1. The filtering resulted in a two-pass filtered SomaScan matrix of 4,721 x 87 (assays x samples), and the TMT-MS data with missingness controlled to <75% was a matrix of 2,286 x 87. These data were combined, with any TMT-MS isoforms missing measurements (N=999) removed, and the remaining 6,008 assays plus isoform measurements were harmonized using transposed TAMPOR as described above leveraging 531 common proteins measured on both platforms with no missing values as a placeholder for TAMPOR-required global internal standard measures.

Following harmonization of the two-platform ATX dataset, the following calculations were performed: (1) response within patient, calculated as log_2_(drug-treated/baseline or placebo) (N=34) and log_2_(placebo/baseline) (N=18) where the protein abundance matrix was determined for all 6008 proteins or assays in the no-missing harmonized data; and (2) synthetic eigenproteins based on the top 20 percent of kME (bicor)-defined hubs of the 34-module network build on the 296-sample cohort with a minimum of four modules members for (a) the full width data of 87 samples, and (b) for the log_2_(ratio) (N=34+18) drug and placebo response matrix (6,008 x 52) resulting from calculation (1) (Extended Data).

### M20 ATX Drug Response Predictor Using Multiprotein Ratios from Drug-naïve CSF

The synthetic eigenprotein for longitudinal baseline-subtracted M20 response to 6 months of drug treatment was calculated as described in the above section, calculation (2)(b), for the 34 patients who completed the trial. The response in one patient (ATX-001-046) was the only outlier more than 4.5 SD from the mean response of the M20 synthetic eigenprotein, and this individual’s samples were not considered in further analyses. The remaining 33 M20 response values constituted the target for prediction. To identify M20 response predictors, a matrix of candidate multiprotein predictors was assembled as follows: eighteen placebo samples without washout effect due to the crossover study design in the ATX cohort and the remainder of the 33 non-outlier naïve samples preceeding drug treatment—deriving from baseline samples—served as paired drug-naïve samples for the 33 baseline-subtracted samples. Their two-platform harmonized log_2_(abundance) matrix for all available MS plus SomaScan measures (N=6,008), and 32 synthetic eigenproteins successfully calculated in these naïve samples from the 34 eigenproteins as template, was subjected to samplewise subtraction of the median log_2_(abundance) of each sample’s 6,040 (N=6,008+32) values. Then, the matrix was subjected to a scaling normalization by multiplying all values in each column (sample) by a scale factor S, such that the sum of the squares of the values in each column becomes 1.0. This operation ensures that the data is scaled appropriately and is essential for subsequent analysis and follows the preparation of abundance matrix data for unweighted multifactor predictor signature calculations described previously(*53, 54*). Subsequently, scaled log_2_-transformed data was unlogged by exponentiating the values (raising 2 to the power of each value), resulting in high-precision, unlogged, scaled normalized values for all protein and synthetic eigenprotein measurements that could contribute to predictors calculated based on these data. Forty predictor candidate individual proteins were selected from the top 20 positive and top 20 negative correlates using bicor between the harmonized pre-scaled 6,008 proteins plus 32 synthetic eigenproteins and the M20 response. Then positive correlates were used as numerator factors and negative correlates as denominator factors in the assembly of ratios of up to 4 summed numerators divided by up to 4 summed denominators. Every 1:1, 2:2, and 3:3 combination ratio was calculated, and a random 10% of all possible 4:4 combination ratios were calculated for a total of 3.66 million ratios representing candidate predictors. The top multiprotein predictor ratio was identified by correlation using bicor, sorting all 3.66 million rho values.

### SomaScan SOMAmer Overlap with CSF Protein Quantitative Trait Loci (pQTLs)

Overlap of SomaScan-measured CSF pQTLs as previously described(*19*) with human SOMAmers after each QC step was counted and visualized using the venneuler R package (v1.1-0) venneuler function.

### Module Quantitative Trait Loci (mQTL) Analysis

Participants were genotyped using the Affymetrix Precision Medicine Array. Quality control of genotypes was performed using Plink v1.90(*55*). We excluded samples with genotype missing rate >10% and variants meeting any of the following criteria: genotype missing rate >10%, minor allele frequency <5%, Hardy-Weinberg equilibrium *p* value < 1 x 10^−7^. Genotypes were imputed to the 1000 Genome Project Phase 3 (*56*) using the Michigan Imputation Server (*57*) with mixed population parameter. SNPs with imputation *R*^#^ > 0.3 were retained for analysis. To identify genetic variants associated with a protein co-expression module, we modeled the first eigenprotein of the protein module as a function of genotype, adjusting for sex, age, and cognitive diagnosis using a univariate linear mixed model (GEMMA (*58*)). To control for false positives due to population stratification or other factors, we performed genomic control adjustment of *p* values(*23*). Variants associated with a module at an adjusted genome-wide significance (*p* <5 × 10^!$^) were categorized as either *cis-* or *trans-* protein module quantitative trait locus (modQTL). *cis*-mod-QTL was defined as a SNP within 1 megabase of any of the genes in the corresponding module; otherwise, they were categorized as *trans*-mod-QTLs.

### CSF pQTL Analysis by Platform

To ensure the homogeneity of the study population, only individuals of European ancestry were included in this analysis. Cryptic relatedness was checked and none was identified. To infer potentially hidden confounding variables, we performed surrogate variable analysis using the sva package in R (*59*). We regressed out the effects of age, sex, and cognitive diagnosis from the proteomic profile before using it for surrogate variable analysis. For each dataset, we determined the number of surrogate variables (SVs) using the sva() function. To identify cis-pQTLs in each dataset (TMT, SOMA_Retained and SOMA_Discarded) for 242 European individuals, we used linear regression implemented by PLINK2 (*55*). Sex, age, cognitive diagnosis, first 10 genetic principal components and SVs (specifically 7 SVs for TMT profiles; 19 SVs for SOMA_Retained, and 6 SVs for SOMA_Discarded) as covariates for each dataset separately. We used a 500kb window upstream and downstream of the gene for QTL analysis. Multiple testing was adjusted with false discovery rate (FDR). Sites with FDR < 0.05 were considered significant.

### Other Statistics

All statistical analyses were performed in R (v4.0.2). Boxplots represent the median, 25th, and 75th percentile extremes; thus, hinges of a box represent the interquartile range of the two middle quartiles of data within a group. The farthest data points up to 1.5 times the interquartile range away from box hinges define the extent of whiskers (error bars). Correlations were performed using the biweight midcorrelation function as implemented in the WGCNA R package or Pearson correlation. Comparisons between two groups were performed by two-tailed *t* test. Comparisons among three or more groups were performed with Kruskal-Wallis nonparametric ANOVA or standard ANOVA with Tukey post hoc pairwise comparison of significance. *P* values were adjusted for multiple comparisons by false discovery rate (FDR) correction according to the Benjamini-Hochberg method where indicated. *Z* score conversion of normalized protein data and normalized protein eigenproteins or synthetic eigenproteins were calculated as fold of standard deviation from the mean.

## Data and Code Availability

Raw data, case traits, and analyses related to this manuscript are available at https://www.synapse.org/Emory300ATX. The atomoxetine trial information and data were described in Levey *et al.(27)*. The TAMPOR algorithm used for median normalization has been described(*48*) and is fully documented and available as an R function, which can be downloaded from https://github.com/edammer/TAMPOR. The results published here are in whole or in part based on data obtained from the AMP-AD Knowledge Portal (https://adknowledgeportal.synapse.org). The AMP-AD Knowledge Portal is a platform for accessing data, analyses and tools generated by the AMP-AD Target Discovery Program and other programs supported by the National Institute on Aging to enable open-science practices and accelerate translational learning. The data, analyses and tools are shared early in the research cycle without a publication embargo on secondary use. Data are available for general research use according to the following requirements for data access and data attribution (https://adknowledgeportal.synapse.org/#/DataAccess/Instructions).

## Author Contributions

E.C.B.J., E.B.D, J.J.L., L.P., and D.M.D. designed the experiments. L.P. and D.M.D. carried out experiments. E.B.D., E.C.B.J., A.S., E.G., S.P.R., V.G., E.A.F., and G.T.G. analyzed data. K.A.W., V.E., L.L.J., V.G., D.W., C.C., J.J.L., T.S.W., A.P.W., N.T.S., and A.I.L. provided advice on the interpretation of data and manuscript review. E.C.B.J. wrote the manuscript with input from co-authors. All authors approved the final manuscript.

## Supporting information

Dammer et al Supplementary Tables

Dammer et al Extended Data

## Data Availability

Raw data, case traits, and analyses related to this manuscript are available at https://www.synapse.org/Emory300ATX.

---

**Supplementary Figure 1.**
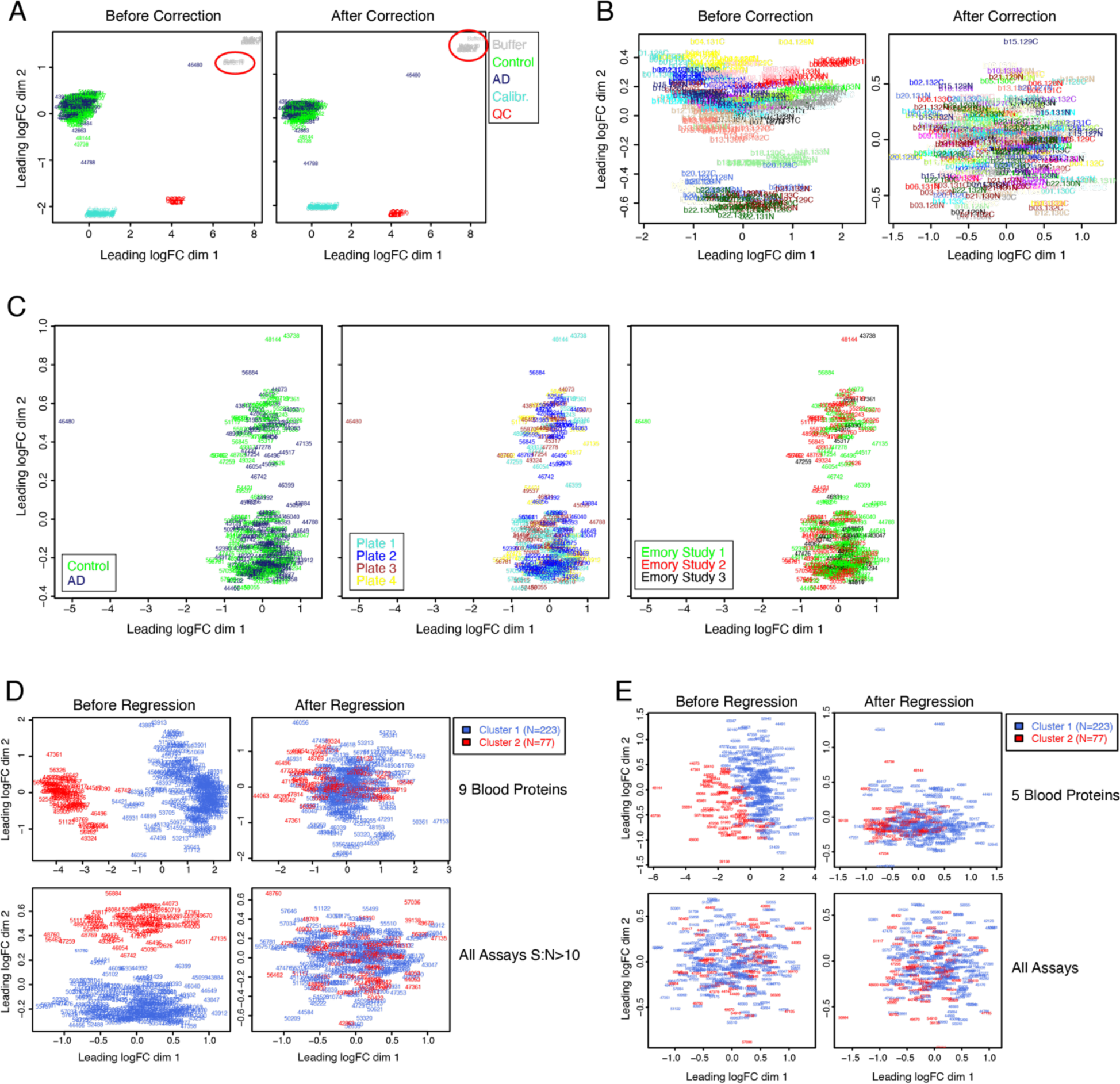
Correction of Batch Effects and Nuisance Variance. (A) Multi-dimensional scaling (MDS) analysis of the SomaScan data including experimental samples, buffer samples, calibrator samples, and QC samples (*N*=344 samples, 7596 assays). Plates with significantly different background buffer signal are circled in red (left). MDS analysis of SomaScan data after median buffer background signal difference correction in the outlier plates (right). (B) MDS analysis of TMT-MS data before batch correction (left) and after batch correction using the global internal standard (right). (C) MDS analysis of SomaScan data filtering on only high signal-to-noise (S:N greater than 10 after background subtraction, *n*=1119) proteins and coloring samples by diagnostic status (left), the plate on which they were analyzed (middle), or the Emory study from which they were obtained (right). (D) MDS analysis of SomaScan data for a 9-blood protein signature (top) or all proteins with S:N greater than 10 (bottom) before (left) and after (right) regression of the data on the 9-blood protein eigenprotein. (E) MDS analysis of TMT-MS data for a 5-blood protein signature (top) or all proteins (bottom) before (left) and after (right) regression of the data on the 5-blood protein eigenprotein.

**Supplementary Figure 2.**
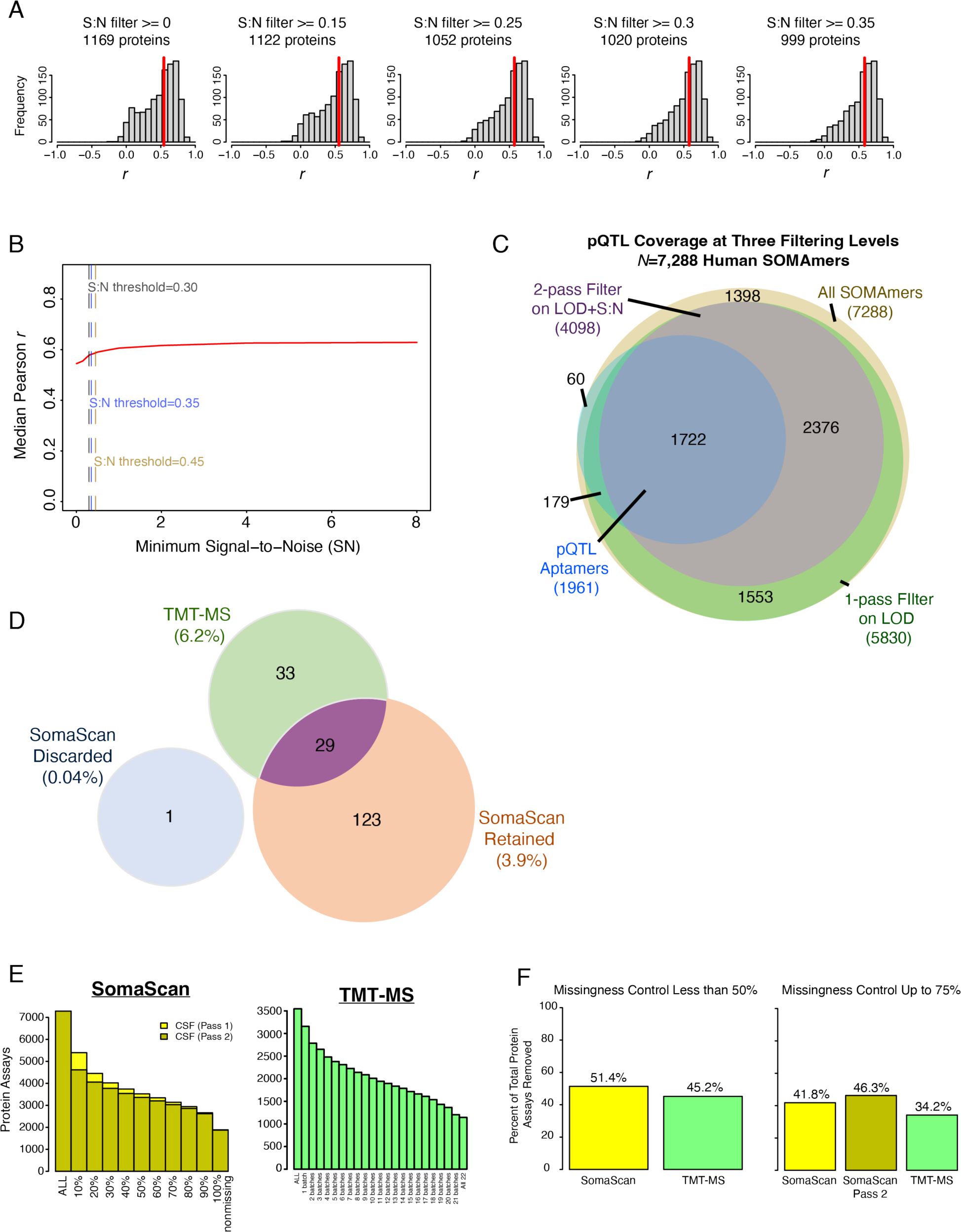
Signal-to-Noise and Missingness Control. (A-C) Signal-to-noise (S:N) filtering. (A) Correlation distributions between SomaScan and TMT-MS at different S:N thresholds applied to the SomaScan data. The number of proteins used for correlation are provided in each plot, with a threshold of at least 75 observations required for correlation. The red vertical line indicates the median correlation. (B) Median correlation across different S:N thresholds applied to the SomaScan data. The vertical dashed lines indicate S:N thresholds of 0.3, 0.35, and 0.45. (C) Overlap of CSF protein quantitative trait loci (pQTLs) with SOMAmers at different levels of filtering. 1722 out of 1961 SOMAmers with pQTLs (88%) were retained after filtering the data on limit of detection (LOD) and S:N greater than 0.3, which resulted in the removal of 44% of the SomaScan assays from further analyses. If multiple SOMAmers to the same gene symbol had pQTLs, only one SOMAmer was counted in the analysis. (D) Number of CSF proteins with pQTLs by platform. For SomaScan, counts are separated by SOMAmers that were retained versus discarded after filtering on LOD and S:N thresholds. The percentage of pQTLs out of the total number of proteins tested in each group is provided. For TMT-MS, only proteins with no missing values were considered in the analysis. Further information is provided in **Supplementary Table 3**. (E, F) Missingness control. (E) The number of protein assays at different levels of missingness in SomaScan (left) and TMT-MS (right) across 300 CSF samples. The number of SomaScan assays at different levels of missingness is given for filtering only on LOD (Pass 1) and filtering on both LOD and S:N thresholds (Pass 2). (F) The percent of assays removed in SomaScan and TMT-MS data when controlling for missingness at 50% (left) and 75% (right).

**Supplementary Figure 3.**
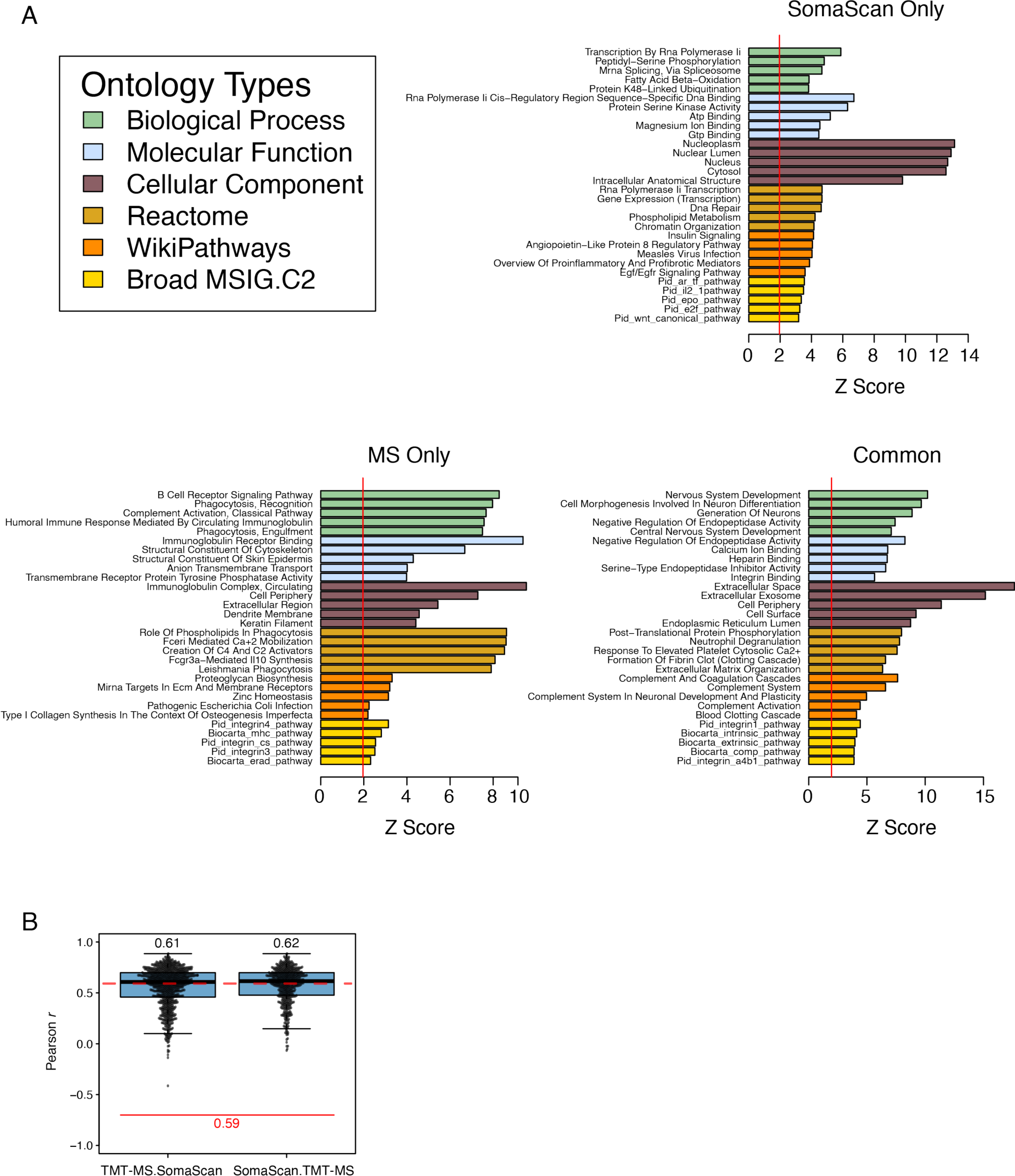
Ontology Analysis and Cross-Platform Correlation. (A) Ontology analysis corresponding to the Venn diagram in Figure 2A. The vertical red line indicates significant enrichment at a *z* score of 1.96. Broad MSIG.C2, Broad Institute Molecular Signatures C2 database. (B) Cross-platform correlation after filtering for significantly differentially abundant proteins in either TMT-MS or SomaScan platforms. The horizontal dashed red line indicated the median correlation without filtering on differentially abundant proteins in either platform. The median correlation did not significantly increase when filtering on differentially abundant proteins in TMT-MS prior to correlation (TMT-MS.SomaScan) or in SomaScan prior to correlation (SomaScan.TMT-MS).

**Supplementary Figure 4.**
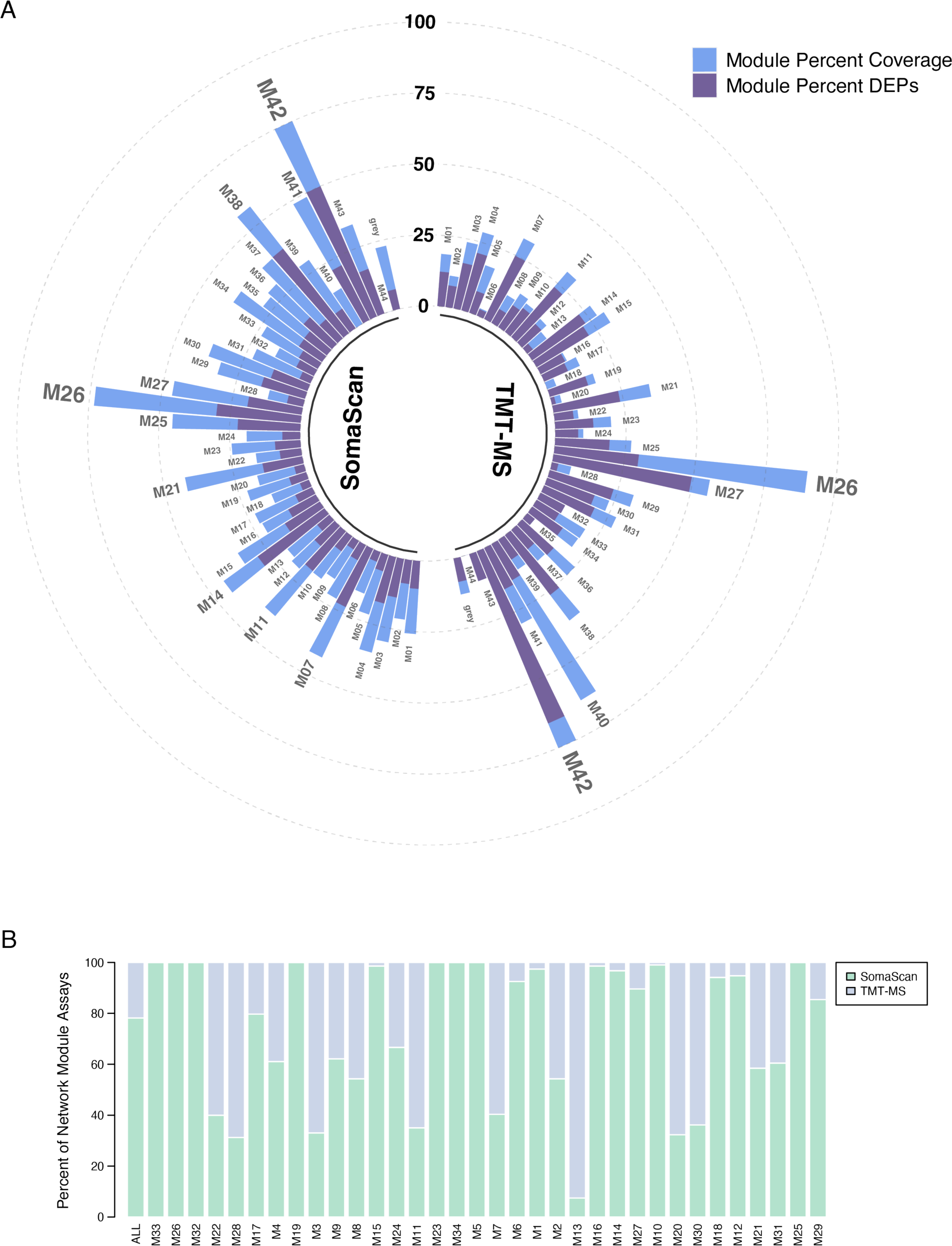
AD Brain Network Module Coverage and Network Module Platform Composition. (A) Module percent coverage of a previously described AD brain protein network(*2*) by SomaScan and TMT-MS platforms, including coverage of proteins that show significant changes in abundance in AD within each module. (B) Percent of assays in each CSF network module from SomaScan or TMT-MS measurements.

**Supplementary Figure 5.**
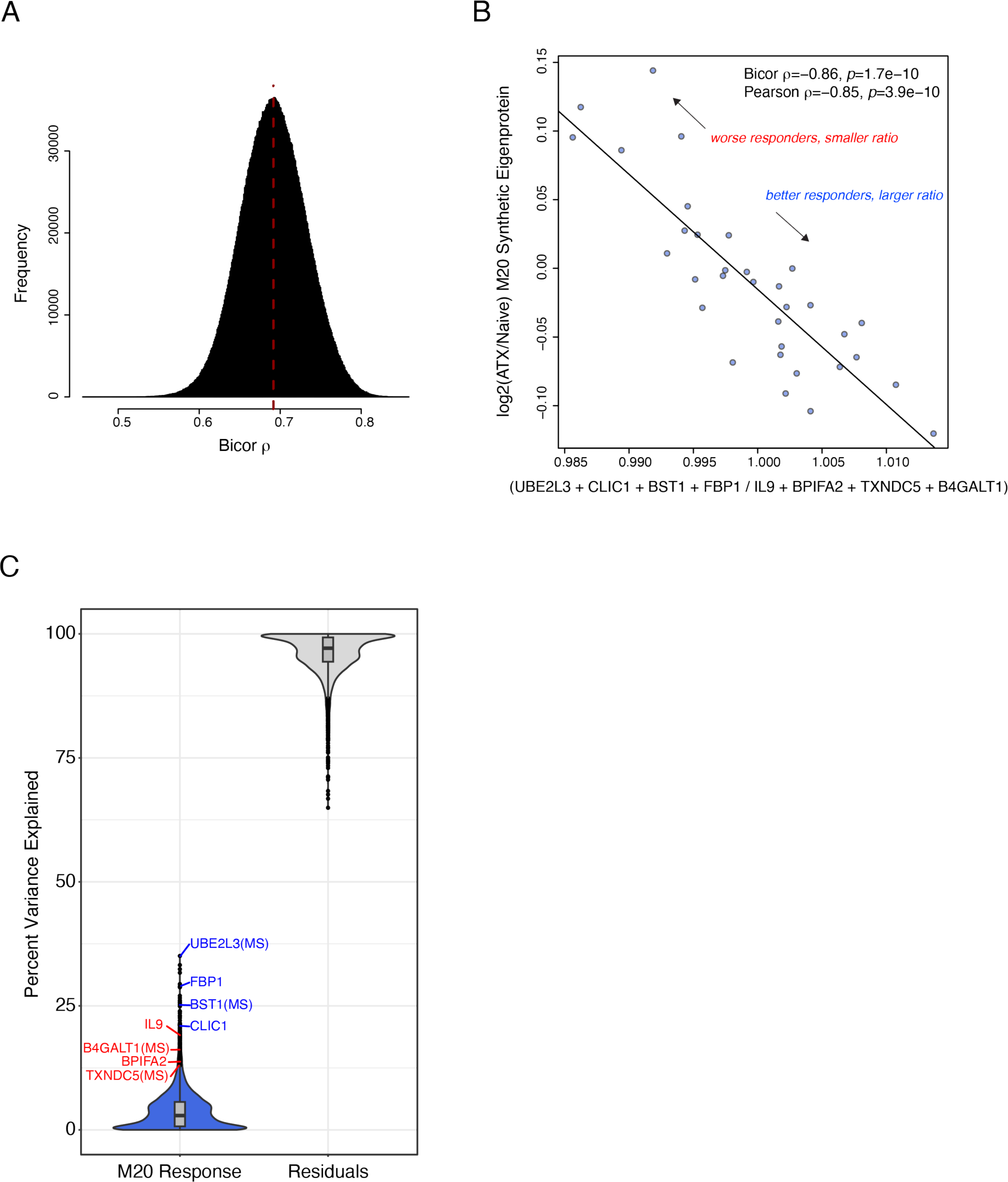
Prediction of ATX Response Using a Protein Ratio. (A) Possible combinations of a four-protein ratio in ATX naïve individuals in which the numerator contained four proteins positively correlated with M20 response and the denominator contained four proteins negatively correlated with M20 response were tested for correlation with the log2(ATX/naïve) M20 synthetic eigenprotein. Distribution of correlations with M20 response is shown. Red vertical line indicates the median correlation. (B) Correlation for the ratio able to best predict M20 response (*n*=34). One outlier is excluded from visualization. UBE2L3, ubiquitin-conjugating enzyme E2 L3; CLIC1, chloride intracellular channel protein 1; BST1, ADP-ribosyl cyclase/cyclic ADP-ribose hydrolase 2; FBP1, fructose-1,6-bisphosphatase 1; IL9, interleukin-9; BPIFA2, BPI fold-containing family A member 2; TXNDC5, thioredoxin domain-containing protein 5; B4GALT1, beta-1,4-galactosyltransferase 1. (C) Variance partition on the M20 response illustrating that all the proteins in the ratio had strong covariance with the log2(ATX/naïve) M20 synthetic eigenprotein (M20 response; blue, positive correlation; red, negative correlation; MS, mass spectrometry measurement).

**Supplementary Figure 6.**
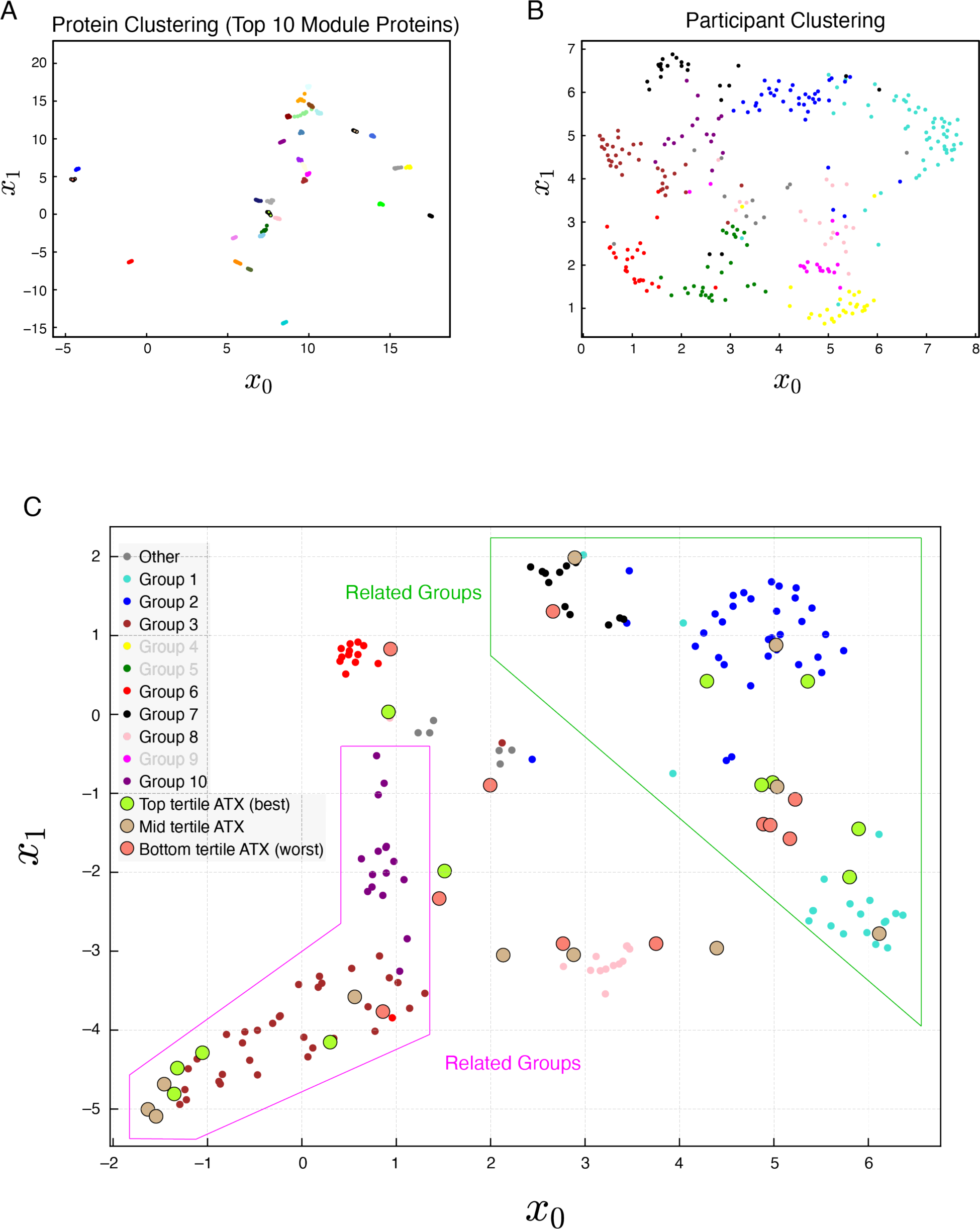
Multi-dimensional Data Reduction of Proteins and People. (A) The top ten hub proteins from each network module were visualized in two-dimensional space across participants by uniform manifold approximation and projection (UMAP). Points are colored by network module as defined by the weighted co-expression network algorithm (WGCNA) used to construct the co-expression network. (B) Participants were visualized in two-dimensional space across the top ten hub proteins from each network module by UMAP. Participants are colored based on their group as defined by WGCNA. (C) UMAP visualization of the ten participant groups as defined by WGCNA. Groups 4, 5, and 9 are excluded for clarity. The baseline CSF proteomes of individuals treated with ATX were divided into tertiles based on degree of ATX response and projected onto the groups. Green and magenta polygons highlight groups related in the hierarchical clustering network.

