## Supplementary material for "Proteomic Network Analysis of Alzheimer’s Disease Cerebrospinal Fluid Reveals Alterations Associated with *APOE* ε4 Genotype and Atomoxetine Treatment": Dammer et al Extended Data

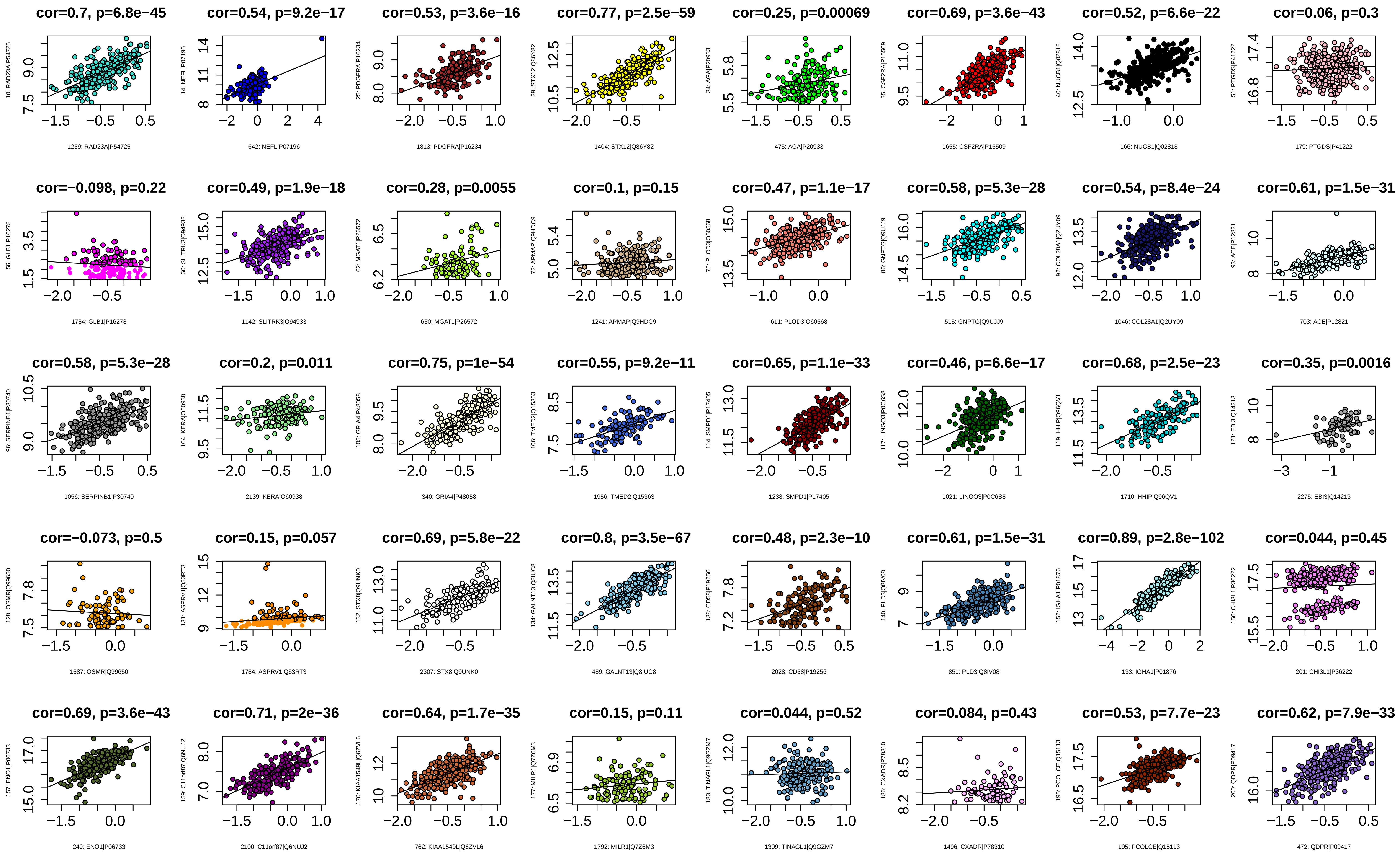
cor=0.7, p=6.8e−45
10: RAD23A|P54725
1259: RAD23A|P54725
cor=0.54, p=9.2e−17
14: NEFL|P07196
642: NEFL|P07196
cor=0.53, p=3.6e−16
25: PDGFRA|P16234
1813: PDGFRA|P16234
cor=0.77, p=2.5e−59
29: STX12|Q86Y82
1404: STX12|Q86Y82
cor=0.25, p=0.00069
34: AGA|P20933
475: AGA|P20933
cor=0.69, p=3.6e−43
35: CSF2RA|P15509
1655: CSF2RA|P15509
cor=0.52, p=6.6e−22
40: NUCB1|Q02818
166: NUCB1|Q02818
cor=0.06, p=0.3
51: PTGDS|P41222
179: PTGDS|P41222
cor=−0.098, p=0.22
56: GLB1|P16278
1754: GLB1|P16278
cor=0.49, p=1.9e−18
60: SLITRK3|O94933
1142: SLITRK3|O94933
cor=0.28, p=0.0055
62: MGAT1|P26572
650: MGAT1|P26572
cor=0.1, p=0.15
72: APMAP|Q9HDC9
1241: APMAP|Q9HDC9
cor=0.47, p=1.1e−17
75: PLOD3|O60568
611: PLOD3|O60568
cor=0.58, p=5.3e−28
86: GNPTG|Q9UJJ9
515: GNPTG|Q9UJJ9
cor=0.54, p=8.4e−24
92: COL28A1|Q2UY09
1046: COL28A1|Q2UY09
cor=0.61, p=1.5e−31
93: ACE|P12821
703: ACE|P12821
cor=0.58, p=5.3e−28
96: SERPINB1|P30740
1056: SERPINB1|P30740
cor=0.2, p=0.011
104: KERA|O60938
2139: KERA|O60938
cor=0.75, p=1e−54
105: GRIA4|P48058
340: GRIA4|P48058
cor=0.55, p=9.2e−11
106: TMED2|Q15363
1956: TMED2|Q15363
cor=0.65, p=1.1e−33
114: SMPD1|P17405
1238: SMPD1|P17405
cor=0.46, p=6.6e−17
117: LINGO3|P0C6S8
1021: LINGO3|P0C6S8
cor=0.68, p=2.5e−23
119: HHIP|Q96QV1
1710: HHIP|Q96QV1
cor=0.35, p=0.0016
121: EBI3|Q14213
2275: EBI3|Q14213
cor=−0.073, p=0.5
128: OSMR|Q99650
1587: OSMR|Q99650
cor=0.15, p=0.057
131: ASPRV1|Q53RT3
1784: ASPRV1|Q53RT3
cor=0.69, p=5.8e−22
132: STX8|Q9UNK0
2307: STX8|Q9UNK0
cor=0.8, p=3.5e−67
134: GALNT13|Q8IUC8
489: GALNT13|Q8IUC8
cor=0.48, p=2.3e−10
138: CD58|P19256
2028: CD58|P19256
cor=0.61, p=1.5e−31
140: PLD3|Q8IV08
851: PLD3|Q8IV08
cor=0.89, p=2.8e−102
152: IGHA1|P01876
133: IGHA1|P01876
cor=0.044, p=0.45
156: CHI3L1|P36222
201: CHI3L1|P36222
cor=0.69, p=3.6e−43
157: ENO1|P06733
249: ENO1|P06733
cor=0.71, p=2e−36
159: C11orf87|Q6NUJ2
2100: C11orf87|Q6NUJ2
cor=0.64, p=1.7e−35
170: KIAA1549L|Q6ZVL6
762: KIAA1549L|Q6ZVL6
cor=0.15, p=0.11
177: MILR1|Q7Z6M3
1792: MILR1|Q7Z6M3
cor=0.044, p=0.52
183: TINAGL1|Q9GZM7
1309: TINAGL1|Q9GZM7
cor=0.084, p=0.43
186: CXADR|P78310
1496: CXADR|P78310
cor=0.53, p=7.7e−23
195: PCOLCE|Q15113
195: PCOLCE|Q15113
cor=0.62, p=7.9e−33
200: QDPR|P09417
472: QDPR|P09417

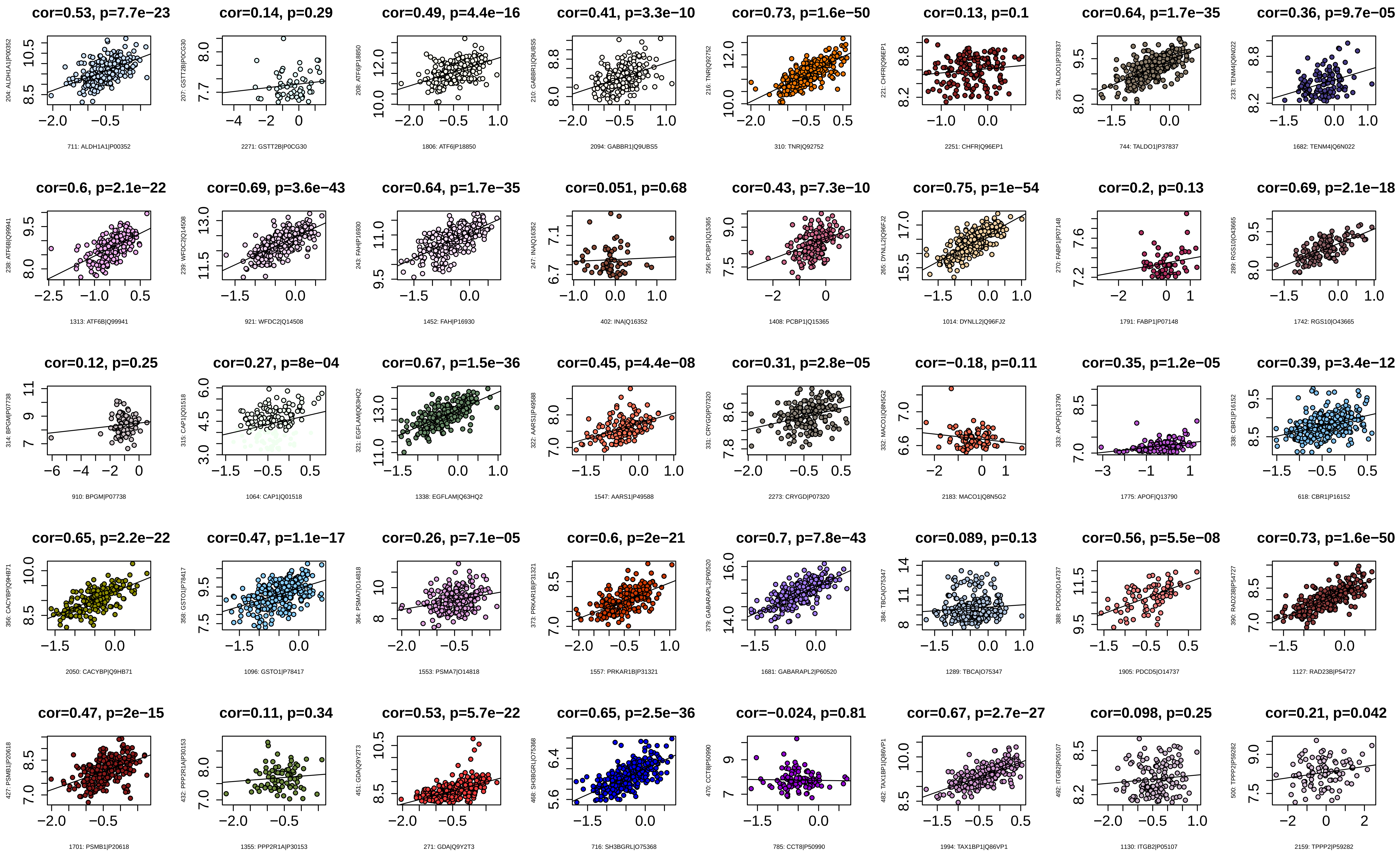
cor=0.53, p=7.7e−23
204: ALDH1A1|P00352
711: ALDH1A1|P00352
cor=0.14, p=0.29
207: GSTT2B|P0CG30
2271: GSTT2B|P0CG30
cor=0.49, p=4.4e−16
208: ATF6|P18850
1806: ATF6|P18850
cor=0.41, p=3.3e−10
210: GABBR1|Q9UBS5
2094: GABBR1|Q9UBS5
cor=0.73, p=1.6e−50
216: TNR|Q92752
310: TNR|Q92752
cor=0.13, p=0.1
221: CHFR|Q96EP1
2251: CHFR|Q96EP1
cor=0.64, p=1.7e−35
225: TALDO1|P37837
744: TALDO1|P37837
cor=0.36, p=9.7e−05
233: TENM4|Q6N022
1682: TENM4|Q6N022
cor=0.6, p=2.1e−22
238: ATF6B|Q99941
1313: ATF6B|Q99941
cor=0.69, p=3.6e−43
239: WFDC2|Q14508
921: WFDC2|Q14508
cor=0.64, p=1.7e−35
243: FAH|P16930
1452: FAH|P16930
cor=0.051, p=0.68
247: INA|Q16352
402: INA|Q16352
cor=0.43, p=7.3e−10
256: PCBP1|Q15365
1408: PCBP1|Q15365
cor=0.75, p=1e−54
265: DYNLL2|Q96FJ2
1014: DYNLL2|Q96FJ2
cor=0.2, p=0.13
270: FABP1|P07148
1791: FABP1|P07148
cor=0.69, p=2.1e−18
289: RGS10|O43665
1742: RGS10|O43665
cor=0.12, p=0.25
314: BPGM|P07738
910: BPGM|P07738
cor=0.27, p=8e−04
315: CAP1|Q01518
1064: CAP1|Q01518
cor=0.67, p=1.5e−36
321: EGFLAM|Q63HQ2
1338: EGFLAM|Q63HQ2
cor=0.45, p=4.4e−08
322: AARS1|P49588
1547: AARS1|P49588
cor=0.31, p=2.8e−05
331: CRYGD|P07320
2273: CRYGD|P07320
cor=−0.18, p=0.11
332: MACO1|Q8N5G2
2183: MACO1|Q8N5G2
cor=0.35, p=1.2e−05
333: APOF|Q13790
1775: APOF|Q13790
cor=0.39, p=3.4e−12
338: CBR1|P16152
618: CBR1|P16152
cor=0.65, p=2.2e−22
356: CACYBP|Q9HB71
2050: CACYBP|Q9HB71
cor=0.47, p=1.1e−17
358: GSTO1|P78417
1096: GSTO1|P78417
cor=0.26, p=7.1e−05
364: PSMA7|O14818
1553: PSMA7|O14818
cor=0.6, p=2e−21
373: PRKAR1B|P31321
1557: PRKAR1B|P31321
cor=0.7, p=7.8e−43
379: GABARAPL2|P60520
1681: GABARAPL2|P60520
cor=0.089, p=0.13
384: TBCA|O75347
1289: TBCA|O75347
cor=0.56, p=5.5e−08
388: PDCD5|O14737
1905: PDCD5|O14737
cor=0.73, p=1.6e−50
390: RAD23B|P54727
1127: RAD23B|P54727
cor=0.47, p=2e−15
427: PSMB1|P20618
1701: PSMB1|P20618
cor=0.11, p=0.34
432: PPP2R1A|P30153
1355: PPP2R1A|P30153
cor=0.53, p=5.7e−22
451: GDA|Q9Y2T3
271: GDA|Q9Y2T3
cor=0.65, p=2.5e−36
468: SH3BGRL|O75368
716: SH3BGRL|O75368
cor=−0.024, p=0.81
470: CCT8|P50990
785: CCT8|P50990
cor=0.67, p=2.7e−27
482: TAX1BP1|Q86VP1
1994: TAX1BP1|Q86VP1
cor=0.098, p=0.25
492: ITGB2|P05107
1130: ITGB2|P05107
cor=0.21, p=0.042
500: TPPP2|P59282
2159: TPPP2|P59282

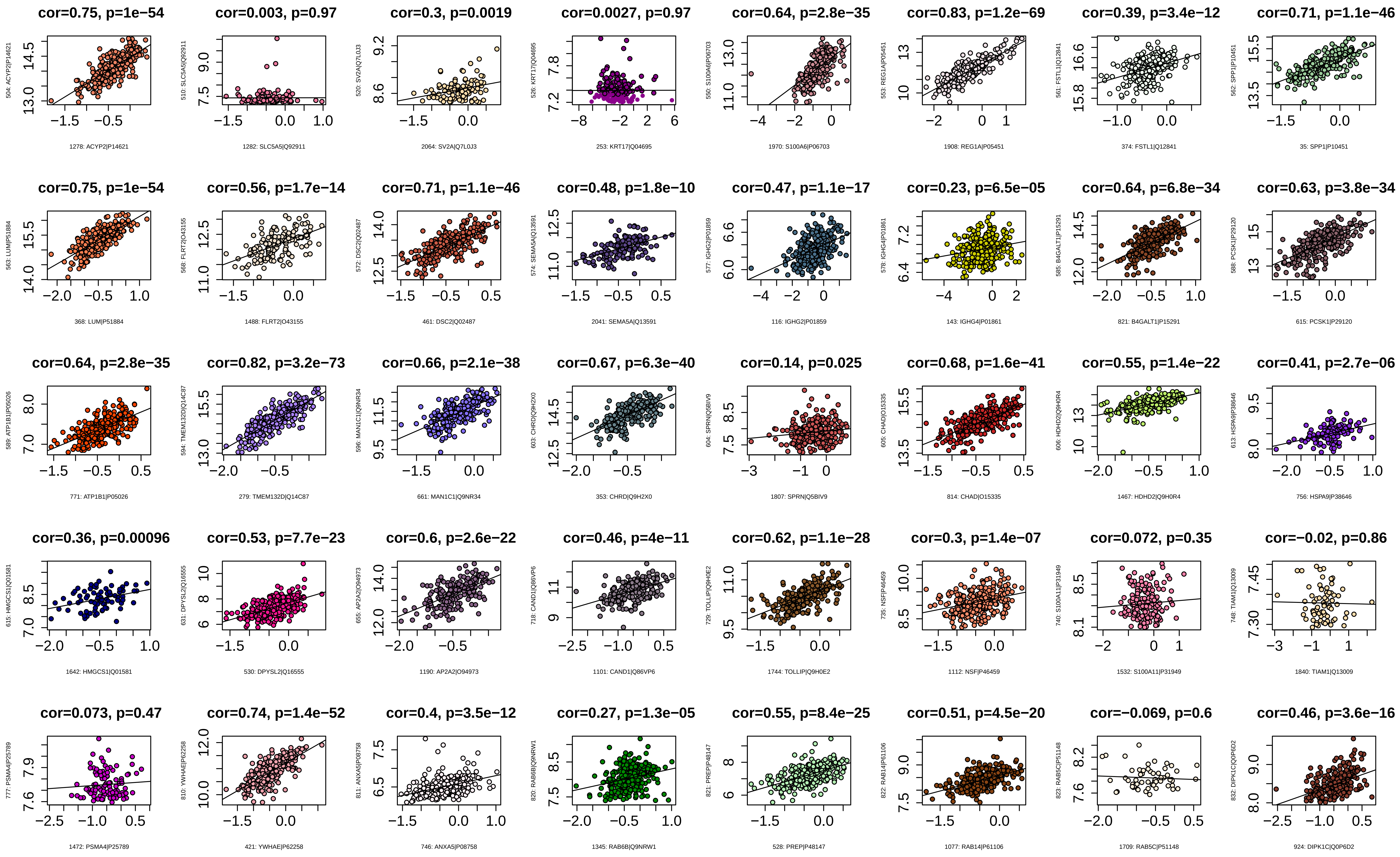
cor=0.75, p=1e−54
cor=0.003, p=0.97
cor=0.3, p=0.0019
cor=0.0027, p=0.97
cor=0.64, p=2.8e−35
cor=0.83, p=1.2e−69
cor=0.39, p=3.4e−12
cor=0.71, p=1.1e−46
cor=0.75, p=1e−54
cor=0.56, p=1.7e−14
cor=0.71, p=1.1e−46
cor=0.48, p=1.8e−10
cor=0.47, p=1.1e−17
cor=0.23, p=6.5e−05
cor=0.64, p=6.8e−34
cor=0.63, p=3.8e−34
cor=0.64, p=2.8e−35
cor=0.82, p=3.2e−73
cor=0.66, p=2.1e−38
cor=0.67, p=6.3e−40
cor=0.14, p=0.025
cor=0.68, p=1.6e−41
cor=0.55, p=1.4e−22
cor=0.41, p=2.7e−06
cor=0.36, p=0.00096
cor=0.53, p=7.7e−23
cor=0.6, p=2.6e−22
cor=0.46, p=4e−11
cor=0.62, p=1.1e−28
cor=0.3, p=1.4e−07
cor=0.072, p=0.35
cor=−0.02, p=0.86
cor=0.073, p=0.47
cor=0.74, p=1.4e−52
cor=0.4, p=3.5e−12
cor=0.27, p=1.3e−05
cor=0.55, p=8.4e−25
cor=0.51, p=4.5e−20
cor=−0.069, p=0.6
cor=0.46, p=3.6e−16

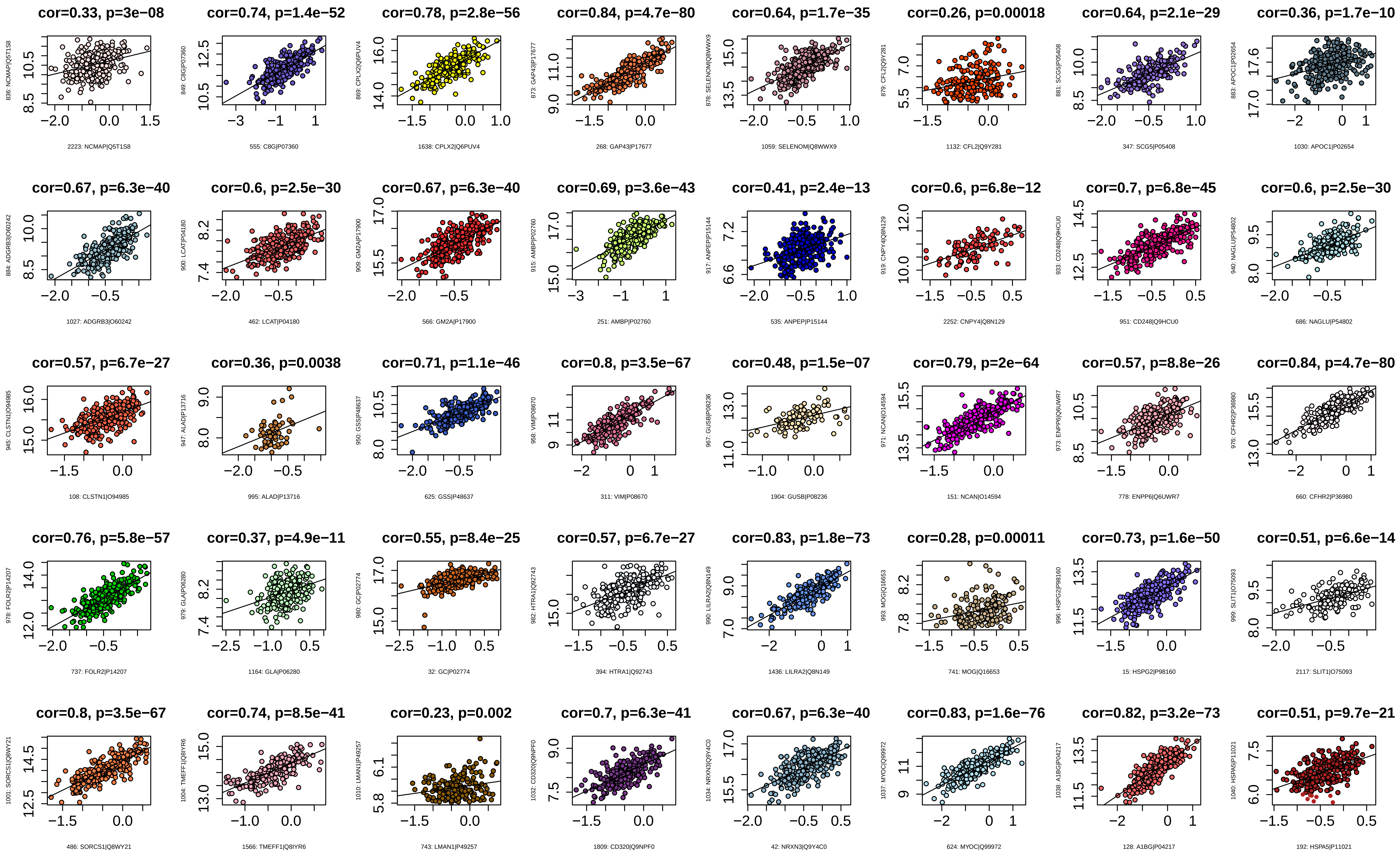

cor=0.33, p=3e−08
836: NCMAP|Q5T1S8
2223: NCMAP|Q5T1S8
cor=0.74, p=1.4e−52
849: C8G|P07360
555: C8G|P07360
cor=0.78, p=2.8e−56
869: CPLX2|Q6PUV4
1638: CPLX2|Q6PUV4
cor=0.84, p=4.7e−80
873: GAP43|P17677
268: GAP43|P17677
cor=0.64, p=1.7e−35
878: SELENOM|Q8WWX9
1059: SELENOM|Q8WWX9
cor=0.26, p=0.00018
879: CFL2|Q9Y281
1132: CFL2|Q9Y281
cor=0.64, p=2.1e−29
881: SCG5|P05408
347: SCG5|P05408
cor=0.36, p=1.7e−10
883: APOC1|P02654
1030: APOC1|P02654
cor=0.67, p=6.3e−40
884: ADGRB3|O60242
1027: ADGRB3|O60242
cor=0.6, p=2.5e−30
900: LCAT|P04180
462: LCAT|P04180
cor=0.67, p=6.3e−40
909: GM2A|P17900
566: GM2A|P17900
cor=0.69, p=3.6e−43
915: AMBP|P02760
251: AMBP|P02760
cor=0.41, p=2.4e−13
917: ANPEP|P15144
535: ANPEP|P15144
cor=0.6, p=6.8e−12
919: CNPY4|Q8N129
2252: CNPY4|Q8N129
cor=0.7, p=6.8e−45
933: CD248|Q9HCU0
951: CD248|Q9HCU0
cor=0.6, p=2.5e−30
940: NAGLU|P54802
686: NAGLU|P54802
cor=0.57, p=6.7e−27
945: CLSTN1|O94985
108: CLSTN1|O94985
cor=0.36, p=0.0038
947: ALAD|P13716
995: ALAD|P13716
cor=0.71, p=1.1e−46
950: GSS|P48637
625: GSS|P48637
cor=0.8, p=3.5e−67
958: VIM|P08670
311: VIM|P08670
cor=0.48, p=1.5e−07
967: GUSB|P08236
1904: GUSB|P08236
cor=0.79, p=2e−64
971: NCAN|O14594
151: NCAN|O14594
cor=0.57, p=8.8e−26
973: ENPP6|Q6UWR7
778: ENPP6|Q6UWR7
cor=0.84, p=4.7e−80
976: CFHR2|P36980
660: CFHR2|P36980
cor=0.76, p=5.8e−57
978: FOLR2|P14207
737: FOLR2|P14207
cor=0.37, p=4.9e−11
979: GLA|P06280
1164: GLA|P06280
cor=0.55, p=8.4e−25
980: GC|P02774
32: GC|P02774
cor=0.57, p=6.7e−27
982: HTRA1|Q92743
394: HTRA1|Q92743
cor=0.83, p=1.8e−73
990: LILRA2|Q8N149
1436: LILRA2|Q8N149
cor=0.28, p=0.00011
993: MOG|Q16653
741: MOG|Q16653
cor=0.73, p=1.6e−50
996: HSPG2|P98160
15: HSPG2|P98160
cor=0.51, p=6.6e−14
999: SLIT1|O75093
2117: SLIT1|O75093
cor=0.8, p=3.5e−67
1001: SORCS1|Q8WY21
486: SORCS1|Q8WY21
cor=0.74, p=8.5e−41
1004: TMEFF1|Q8IYR6
1566: TMEFF1|Q8IYR6
cor=0.23, p=0.002
1010: LMAN1|P49257
743: LMAN1|P49257
cor=0.7, p=6.3e−41
1032: CD320|Q9NPF0
1809: CD320|Q9NPF0
cor=0.67, p=6.3e−40
1034: NRXN3|Q9Y4C0
42: NRXN3|Q9Y4C0
cor=0.83, p=1.6e−76
1037: MYOC|Q99972
624: MYOC|Q99972
cor=0.82, p=3.2e−73
1038: A1BG|P04217
128: A1BG|P04217
cor=0.51, p=9.7e−21
1040: HSPA5|P11021
192: HSPA5|P11021

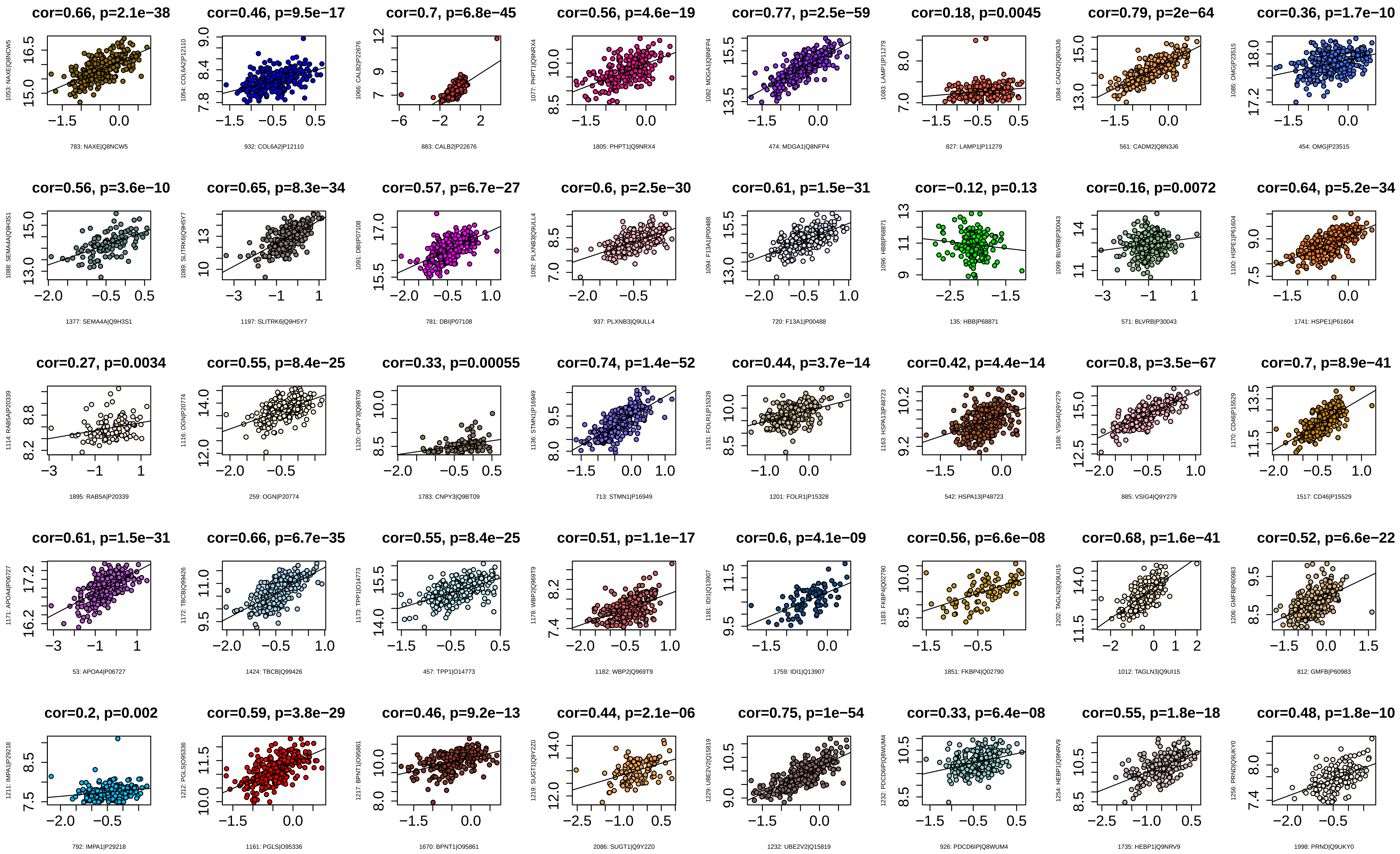

cor=0.66, p=2.1e−38
1053: NAXE|Q8NCW5
783: NAXE|Q8NCW5
cor=0.46, p=9.5e−17
1054: COL6A2|P12110
932: COL6A2|P12110
cor=0.7, p=6.8e−45
1066: CALB2|P22676
883: CALB2|P22676
cor=0.56, p=4.6e−19
1077: PHPT1|Q9NRX4
1805: PHPT1|Q9NRX4
cor=0.77, p=2.5e−59
1082: MDGA1|Q8NFP4
474: MDGA1|Q8NFP4
cor=0.18, p=0.0045
1083: LAMP1|P11279
827: LAMP1|P11279
cor=0.79, p=2e−64
1084: CADM2|Q8N3J6
561: CADM2|Q8N3J6
cor=0.36, p=1.7e−10
1085: OMG|P23515
454: OMG|P23515
cor=0.56, p=3.6e−10
1088: SEMA4A|Q9H3S1
1377: SEMA4A|Q9H3S1
cor=0.65, p=8.3e−34
1089: SLITRK6|Q9H5Y7
1197: SLITRK6|Q9H5Y7
cor=0.57, p=6.7e−27
1091: DBI|P07108
781: DBI|P07108
cor=0.6, p=2.5e−30
1092: PLXNB3|Q9ULL4
937: PLXNB3|Q9ULL4
cor=0.61, p=1.5e−31
1094: F13A1|P00488
720: F13A1|P00488
cor=−0.12, p=0.13
1096: HBB|P68871
135: HBB|P68871
cor=0.16, p=0.0072
1099: BLVRB|P30043
571: BLVRB|P30043
cor=0.64, p=5.2e−34
1100: HSPE1|P61604
1741: HSPE1|P61604
cor=0.27, p=0.0034
1114: RAB5A|P20339
1895: RAB5A|P20339
cor=0.55, p=8.4e−25
1116: OGN|P20774
259: OGN|P20774
cor=0.33, p=0.00055
1120: CNPY3|Q9BT09
1783: CNPY3|Q9BT09
cor=0.74, p=1.4e−52
1136: STMN1|P16949
713: STMN1|P16949
cor=0.44, p=3.7e−14
1151: FOLR1|P15328
1201: FOLR1|P15328
cor=0.42, p=4.4e−14
1163: HSPA13|P48723
542: HSPA13|P48723
cor=0.8, p=3.5e−67
1168: VSIG4|Q9Y279
885: VSIG4|Q9Y279
cor=0.7, p=8.9e−41
1170: CD46|P15529
1517: CD46|P15529
cor=0.61, p=1.5e−31
1171: APOA4|P06727
53: APOA4|P06727
cor=0.66, p=6.7e−35
1172: TBCB|Q99426
1424: TBCB|Q99426
cor=0.55, p=8.4e−25
1173: TPP1|O14773
457: TPP1|O14773
cor=0.51, p=1.1e−17
1178: WBP2|Q969T9
1182: WBP2|Q969T9
cor=0.6, p=4.1e−09
1181: IDI1|Q13907
1759: IDI1|Q13907
cor=0.56, p=6.6e−08
1183: FKBP4|Q02790
1851: FKBP4|Q02790
cor=0.68, p=1.6e−41
1202: TAGLN3|Q9UI15
1012: TAGLN3|Q9UI15
cor=0.52, p=6.6e−22
1206: GMFB|P60983
812: GMFB|P60983
cor=0.2, p=0.002
1211: IMPA1|P29218
792: IMPA1|P29218
cor=0.59, p=3.8e−29
1212: PGLS|O95336
1161: PGLS|O95336
cor=0.46, p=9.2e−13
1217: BPNT1|O95861
1670: BPNT1|O95861
cor=0.44, p=2.1e−06
1219: SUGT1|Q9Y2Z0
2086: SUGT1|Q9Y2Z0
cor=0.75, p=1e−54
1229: UBE2V2|Q15819
1232: UBE2V2|Q15819
cor=0.33, p=6.4e−08
1232: PDCD6IP|Q8WUM4
926: PDCD6IP|Q8WUM4
cor=0.55, p=1.8e−18
1254: HEBP1|Q9NRV9
1735: HEBP1|Q9NRV9
cor=0.48, p=1.8e−10
1256: PRND|Q9UKY0
1998: PRND|Q9UKY0

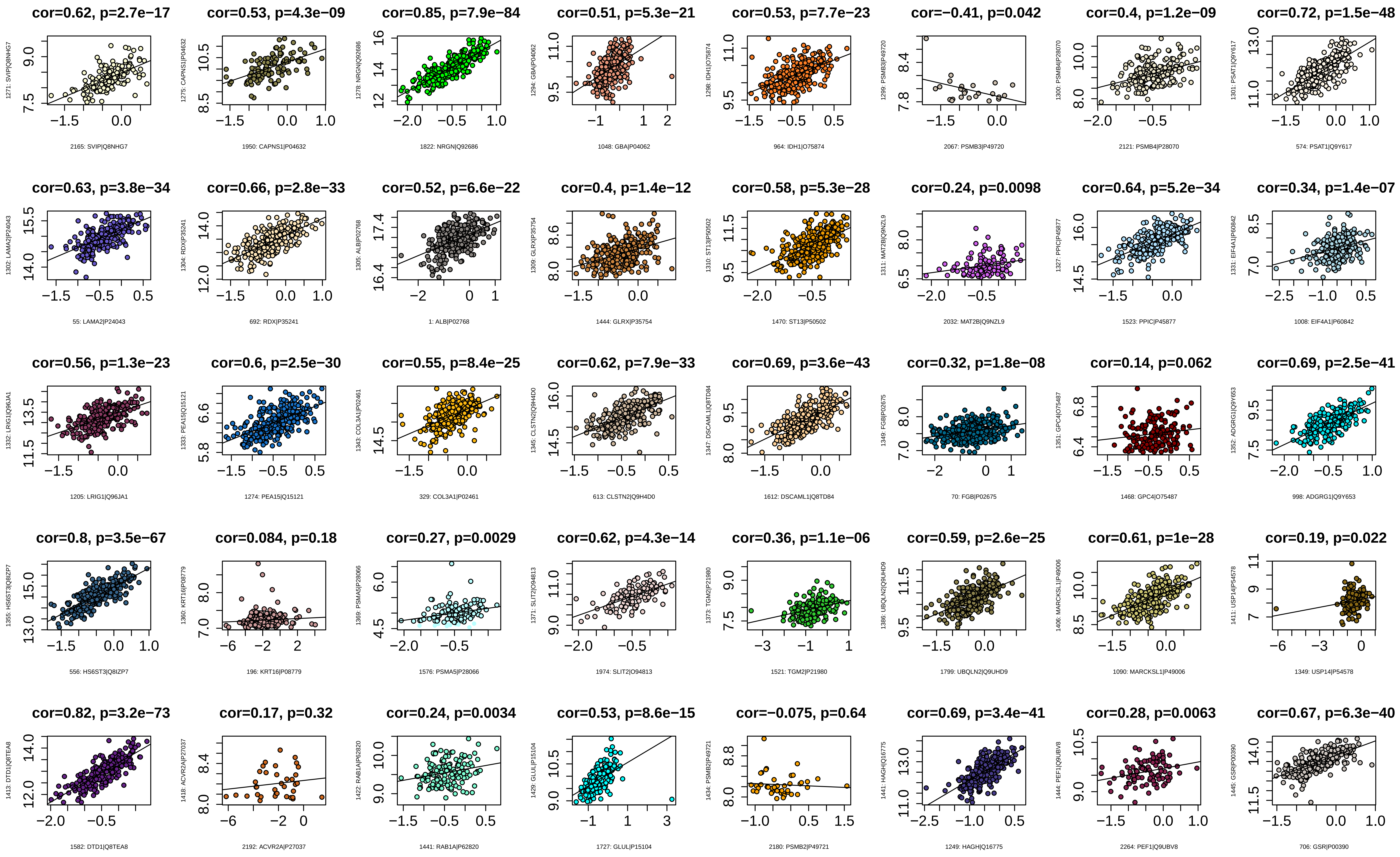

cor=0.62, p=2.7e−17
1271: SVIP|Q8NHG7
2165: SVIP|Q8NHG7
cor=0.53, p=4.3e−09
1275: CAPNS1|P04632
1950: CAPNS1|P04632
cor=0.85, p=7.9e−84
1278: NRGN|Q92686
1822: NRGN|Q92686
cor=0.51, p=5.3e−21
1294: GBA|P04062
1048: GBA|P04062
cor=0.53, p=7.7e−23
1298: IDH1|O75874
964: IDH1|O75874
cor=−0.41, p=0.042
1299: PSMB3|P49720
2067: PSMB3|P49720
cor=0.4, p=1.2e−09
1300: PSMB4|P28070
2121: PSMB4|P28070
cor=0.72, p=1.5e−48
1301: PSAT1|Q9Y617
574: PSAT1|Q9Y617
cor=0.63, p=3.8e−34
1302: LAMA2|P24043
55: LAMA2|P24043
cor=0.66, p=2.8e−33
1304: RDX|P35241
692: RDX|P35241
cor=0.52, p=6.6e−22
1305: ALB|P02768
1: ALB|P02768
cor=0.4, p=1.4e−12
1309: GLRX|P35754
1444: GLRX|P35754
cor=0.58, p=5.3e−28
1310: ST13|P50502
1470: ST13|P50502
cor=0.24, p=0.0098
1311: MAT2B|Q9NZL9
2032: MAT2B|Q9NZL9
cor=0.64, p=5.2e−34
1327: PPIC|P45877
1523: PPIC|P45877
cor=0.34, p=1.4e−07
1331: EIF4A1|P60842
1008: EIF4A1|P60842
cor=0.56, p=1.3e−23
1332: LRIG1|Q96JA1
1205: LRIG1|Q96JA1
cor=0.6, p=2.5e−30
1333: PEA15|Q15121
1274: PEA15|Q15121
cor=0.55, p=8.4e−25
1343: COL3A1|P02461
329: COL3A1|P02461
cor=0.62, p=7.9e−33
1345: CLSTN2|Q9H4D0
613: CLSTN2|Q9H4D0
cor=0.69, p=3.6e−43
1347: DSCAML1|Q8TD84
1612: DSCAML1|Q8TD84
cor=0.32, p=1.8e−08
1349: FGB|P02675
70: FGB|P02675
cor=0.14, p=0.062
1351: GPC4|O75487
1468: GPC4|O75487
cor=0.69, p=2.5e−41
1352: ADGRG1|Q9Y653
998: ADGRG1|Q9Y653
cor=0.8, p=3.5e−67
1355: HS6ST3|Q8IZP7
556: HS6ST3|Q8IZP7
cor=0.084, p=0.18
1360: KRT16|P08779
196: KRT16|P08779
cor=0.27, p=0.0029
1369: PSMA5|P28066
1576: PSMA5|P28066
cor=0.62, p=4.3e−14
1371: SLIT2|O94813
1974: SLIT2|O94813
cor=0.36, p=1.1e−06
1373: TGM2|P21980
1521: TGM2|P21980
cor=0.59, p=2.6e−25
1386: UBQLN2|Q9UHD9
1799: UBQLN2|Q9UHD9
cor=0.61, p=1e−28
1406: MARCKSL1|P49006
1090: MARCKSL1|P49006
cor=0.19, p=0.022
1411: USP14|P54578
1349: USP14|P54578
cor=0.82, p=3.2e−73
1413: DTD1|Q8TEA8
1582: DTD1|Q8TEA8
cor=0.17, p=0.32
1418: ACVR2A|P27037
2192: ACVR2A|P27037
cor=0.24, p=0.0034
1422: RAB1A|P62820
1441: RAB1A|P62820
cor=0.53, p=8.6e−15
1429: GLUL|P15104
1727: GLUL|P15104
cor=−0.075, p=0.64
1434: PSMB2|P49721
2180: PSMB2|P49721
cor=0.69, p=3.4e−41
1441: HAGH|Q16775
1249: HAGH|Q16775
cor=0.28, p=0.0063
1444: PEF1|Q9UBV8
2264: PEF1|Q9UBV8
cor=0.67, p=6.3e−40
1445: GSR|P00390
706: GSR|P00390

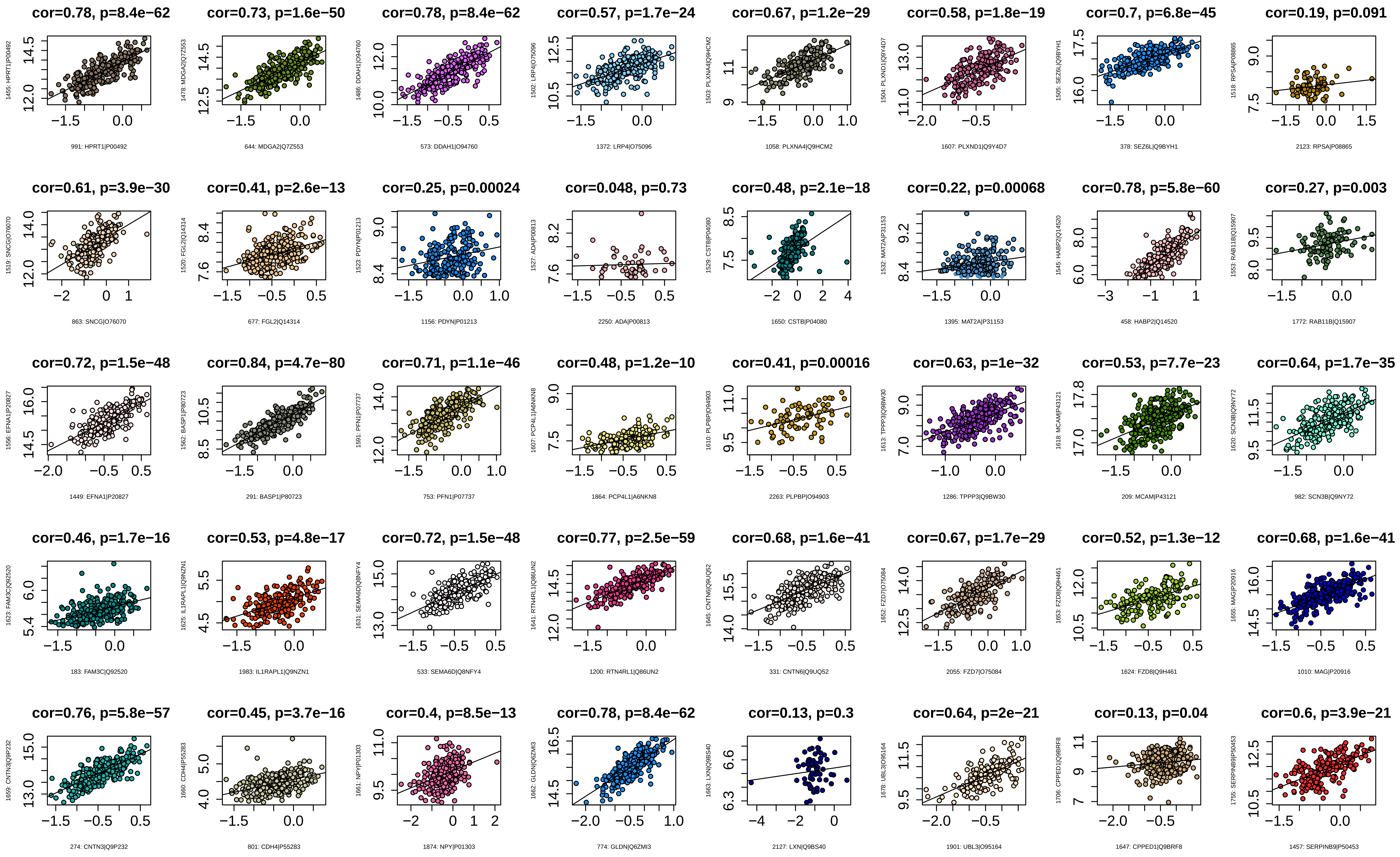
cor=0.78, p=8.4e−62
991: HPRT1|P00492
cor=0.73, p=1.6e−50
644: MDGA2|Q7Z553
cor=0.78, p=8.4e−62
573: DDAH1|O94760
cor=0.57, p=1.7e−24
1372: LRP4|O75096
cor=0.67, p=1.2e−29
1058: PLXNA4|Q9HCM2
cor=0.58, p=1.8e−19
1607: PLXND1|Q9Y4D7
cor=0.7, p=6.8e−45
378: SEZ6L|Q9BYH1
cor=0.19, p=0.091
2123: RPSA|P08865
cor=0.61, p=3.9e−30
863: SNCG|O76070
cor=0.41, p=2.6e−13
677: FGL2|Q14314
cor=0.25, p=0.00024
1156: PDYN|P01213
cor=0.048, p=0.73
2250: ADA|P00813
cor=0.48, p=2.1e−18
1650: CSTB|P04080
cor=0.22, p=0.00068
1395: MAT2A|P31153
cor=0.78, p=5.8e−60
458: HABP2|Q14520
cor=0.27, p=0.003
1772: RAB11B|Q15907
cor=0.72, p=1.5e−48
1449: EFNA1|P20827
cor=0.84, p=4.7e−80
291: BASP1|P80723
cor=0.71, p=1.1e−46
753: PFN1|P07737
cor=0.48, p=1.2e−10
1864: PCP4L1|A6NKN8
cor=0.41, p=0.00016
2263: PLPBP|O94903
cor=0.63, p=1e−32
1286: TPPP3|Q9BW30
cor=0.53, p=7.7e−23
209: MCAM|P43121
cor=0.64, p=1.7e−35
982: SCN3B|Q9NY72
cor=0.46, p=1.7e−16
183: FAM3C|Q92520
cor=0.53, p=4.8e−17
1983: IL1RAPL1|Q9NZN1
cor=0.72, p=1.5e−48
533: SEMA6D|Q8NFY4
cor=0.77, p=2.5e−59
1200: RTN4RL1|Q86UN2
cor=0.68, p=1.6e−41
331: CNTN6|Q9UQ52
cor=0.67, p=1.7e−29
2055: FZD7|O75084
cor=0.52, p=1.3e−12
1624: FZD8|Q9H461
cor=0.68, p=1.6e−41
1010: MAG|P20916
cor=0.76, p=5.8e−57
274: CNTN3|Q9P232
cor=0.45, p=3.7e−16
801: CDH4|P55283
cor=0.4, p=8.5e−13
1874: NPY|P01303
cor=0.78, p=8.4e−62
774: GLDN|Q6ZMI3
cor=0.13, p=0.3
2127: LXN|Q9BS40
cor=0.64, p=2e−21
1901: UBL3|O95164
cor=0.13, p=0.04
1647: CPPED1|Q9BRF8
cor=0.6, p=3.9e−21
1457: SERPINB9|P50453

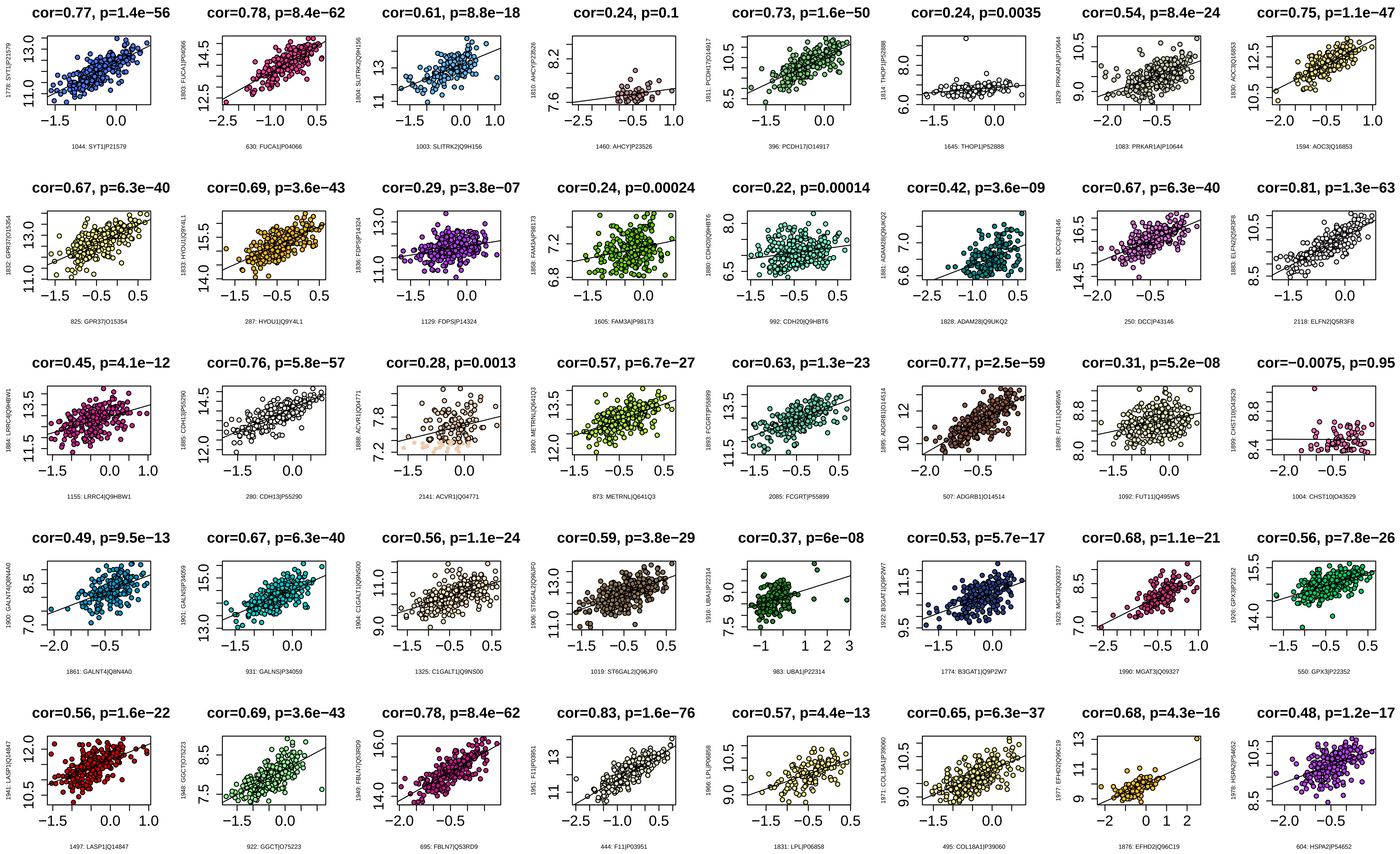
cor=0.77, p=1.4e−56
1044: SYT1|P21579
cor=0.78, p=8.4e−62
630: FUCA1|P04066
cor=0.61, p=8.8e−18
1003: SLITRK2|Q9H156
cor=0.24, p=0.1
1460: AHCY|P23526
cor=0.73, p=1.6e−50
396: PCDH17|O14917
cor=0.24, p=0.0035
1645: THOP1|P52888
cor=0.54, p=8.4e−24
1083: PRKAR1A|P10644
cor=0.75, p=1.1e−47
1594: AOC3|Q16853
cor=0.67, p=6.3e−40
825: GPR37|O15354
cor=0.69, p=3.6e−43
287: HYOU1|Q9Y4L1
cor=0.29, p=3.8e−07
1129: FDPS|P14324
cor=0.24, p=0.00024
1605: FAM3A|P98173
cor=0.22, p=0.00014
992: CDH20|Q9HBT6
cor=0.42, p=3.6e−09
1828: ADAM28|Q9UKQ2
cor=0.67, p=6.3e−40
250: DCC|P43146
cor=0.81, p=1.3e−63
2118: ELFN2|Q5R3F8
cor=0.45, p=4.1e−12
1155: LRRC4|Q9HBW1
cor=0.76, p=5.8e−57
280: CDH13|P55290
cor=0.28, p=0.0013
2141: ACVR1|Q04771
cor=0.57, p=6.7e−27
873: METRNL|Q641Q3
cor=0.63, p=1.3e−23
2085: FCGRT|P55899
cor=0.77, p=2.5e−59
507: ADGRB1|O14514
cor=0.31, p=5.2e−08
1092: FUT11|Q495W5
cor=−0.0075, p=0.95
1004: CHST10|O43529
cor=0.49, p=9.5e−13
1861: GALNT4|Q8N4A0
cor=0.67, p=6.3e−40
931: GALNS|P34059
cor=0.56, p=1.1e−24
1325: C1GALT1|Q9NS00
cor=0.59, p=3.8e−29
1019: ST6GAL2|Q96JF0
cor=0.37, p=6e−08
983: UBA1|P22314
cor=0.53, p=5.7e−17
1774: B3GAT1|Q9P2W7
cor=0.68, p=1.1e−21
1990: MGAT3|Q09327
cor=0.56, p=7.8e−26
550: GPX3|P22352
cor=0.56, p=1.6e−22
1497: LASP1|Q14847
cor=0.69, p=3.6e−43
922: GGCT|O75223
cor=0.78, p=8.4e−62
695: FBLN7|Q53RD9
cor=0.83, p=1.6e−76
444: F11|P03951
cor=0.57, p=4.4e−13
1831: LPL|P06858
cor=0.65, p=6.3e−37
495: COL18A1|P39060
cor=0.68, p=4.3e−16
1876: EFHD2|Q96C19
cor=0.48, p=1.2e−17
604: HSPA2|P54652

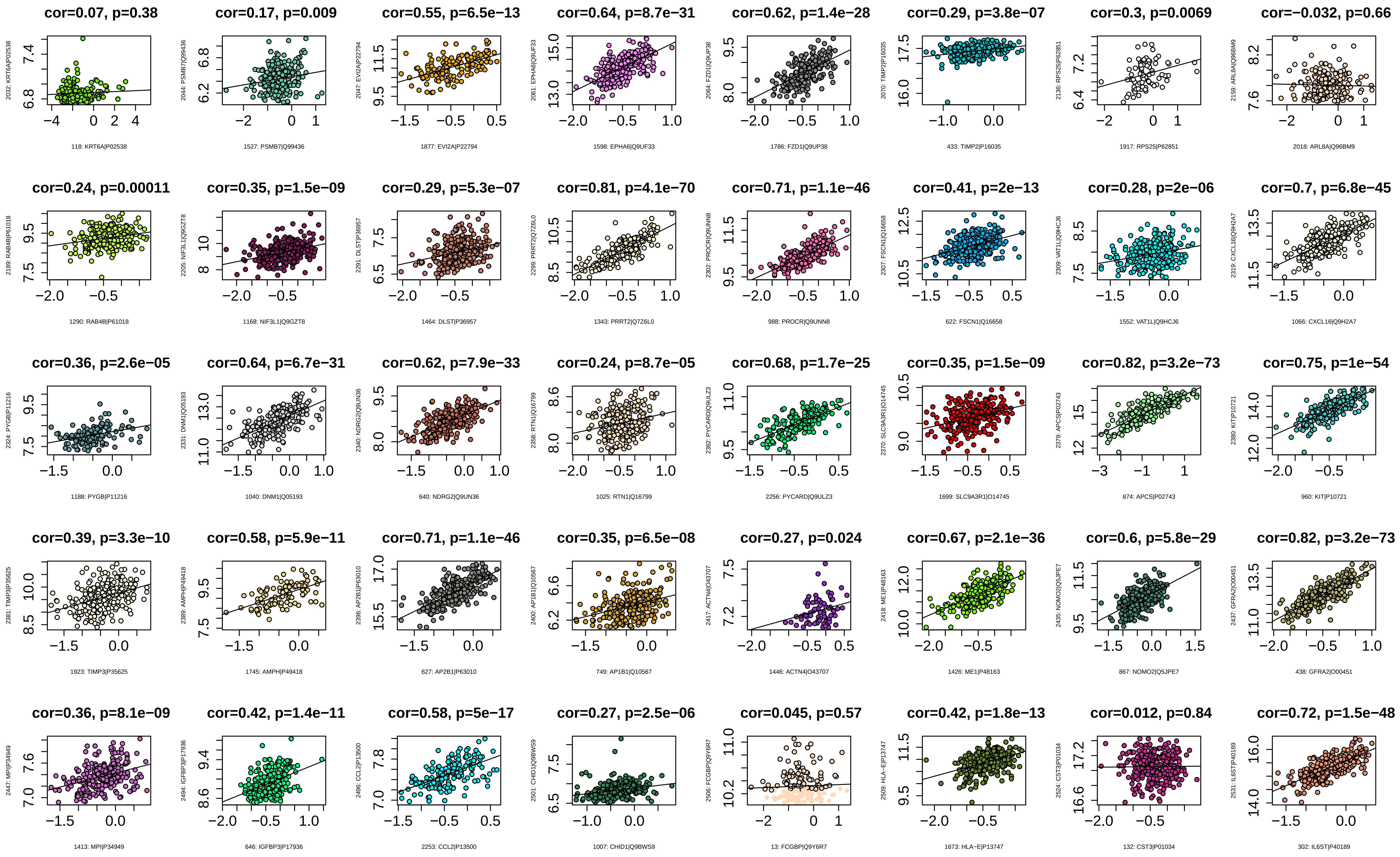
cor=0.07, p=0.38
118: KRT6A|P02538
cor=0.17, p=0.009
1527: PSMB7|Q99436
cor=0.55, p=6.5e−13
1877: EVI2A|P22794
cor=0.64, p=8.7e−31
1598: EPHA6|Q9UF33
cor=0.62, p=1.4e−28
1786: FZD1|Q9UP38
cor=0.29, p=3.8e−07
433: TIMP2|P16035
cor=0.3, p=0.0069
1917: RPS25|P62851
cor=−0.032, p=0.66
2018: ARL8A|Q96BM9
cor=0.24, p=0.00011
1290: RAB4B|P61018
cor=0.35, p=1.5e−09
1168: NIF3L1|Q9GZT8
cor=0.29, p=5.3e−07
1464: DLST|P36957
cor=0.81, p=4.1e−70
1343: PRRT2|Q7Z6L0
cor=0.71, p=1.1e−46
988: PROCR|Q9UNN8
cor=0.41, p=2e−13
622: FSCN1|Q16658
cor=0.28, p=2e−06
1552: VAT1L|Q9HCJ6
cor=0.7, p=6.8e−45
1066: CXCL16|Q9H2A7
cor=0.36, p=2.6e−05
1188: PYGB|P11216
cor=0.64, p=6.7e−31
1040: DNM1|Q05193
cor=0.62, p=7.9e−33
640: NDRG2|Q9UN36
cor=0.24, p=8.7e−05
1025: RTN1|Q16799
cor=0.68, p=1.7e−25
2256: PYCARD|Q9ULZ3
cor=0.35, p=1.5e−09
1699: SLC9A3R1|O14745
cor=0.82, p=3.2e−73
874: APCS|P02743
cor=0.75, p=1e−54
960: KIT|P10721
cor=0.39, p=3.3e−10
1923: TIMP3|P35625
cor=0.58, p=5.9e−11
1745: AMPH|P49418
cor=0.71, p=1.1e−46
627: AP2B1|P63010
cor=0.35, p=6.5e−08
749: AP1B1|Q10567
cor=0.27, p=0.024
1446: ACTN4|O43707
cor=0.67, p=2.1e−36
1426: ME1|P48163
cor=0.6, p=5.8e−29
867: NOMO2|Q5JPE7
cor=0.82, p=3.2e−73
438: GFRA2|O00451
cor=0.36, p=8.1e−09
1413: MPI|P34949
cor=0.42, p=1.4e−11
646: IGFBP3|P17936
cor=0.58, p=5e−17
2253: CCL2|P13500
cor=0.27, p=2.5e−06
1007: CHID1|Q9BWS9
cor=0.045, p=0.57
13: FCGBP|Q9Y6R7
cor=0.42, p=1.8e−13
1673: HLA−E|P13747
cor=0.012, p=0.84
132: CST3|P01034
cor=0.72, p=1.5e−48
302: IL6ST|P40189

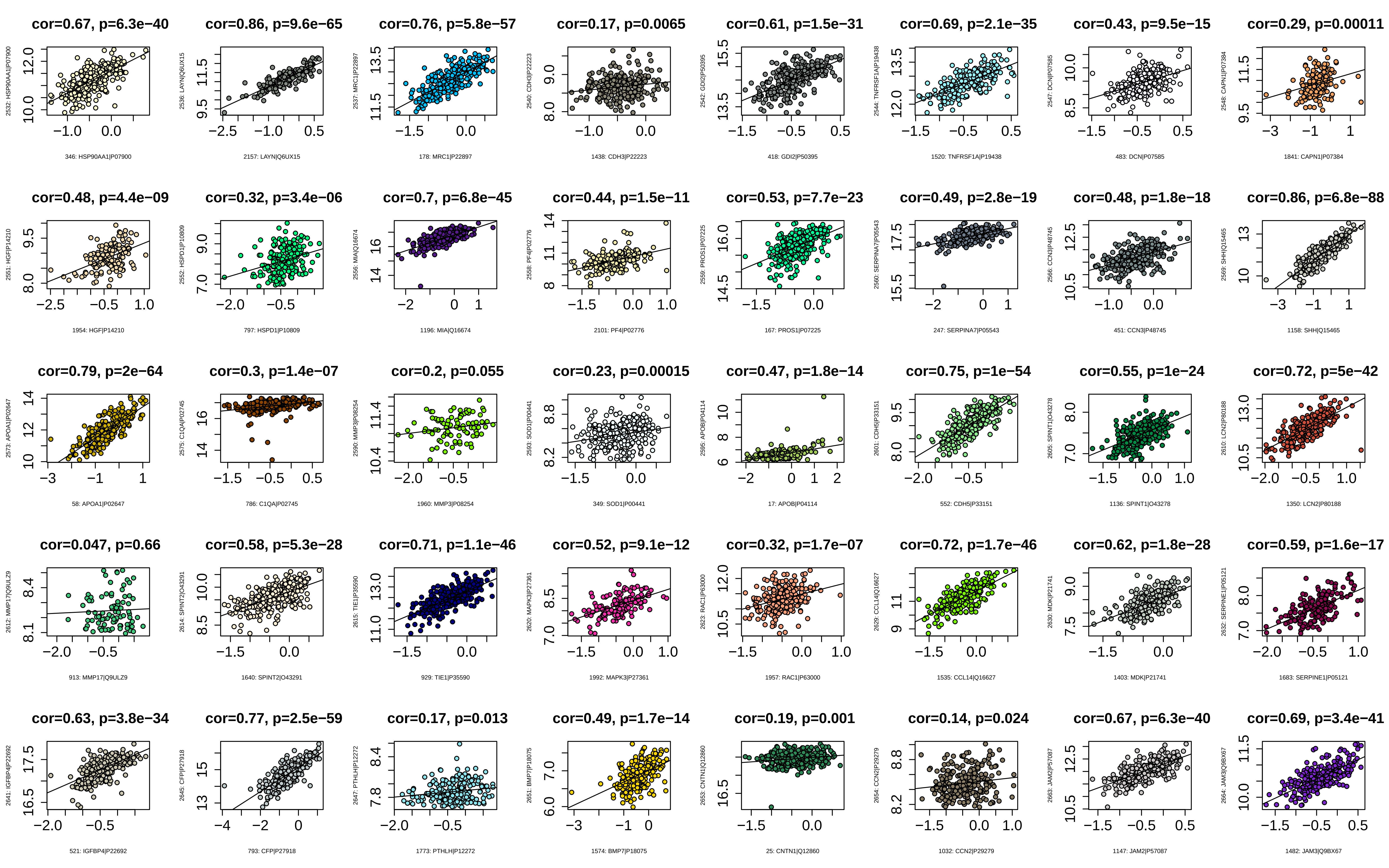

cor=0.67, p=6.3e−40
2532: HSP90AA1|P07900
346: HSP90AA1|P07900
cor=0.86, p=9.6e−65
2536: LAYN|Q6UX15
2157: LAYN|Q6UX15
cor=0.76, p=5.8e−57
2537: MRC1|P22897
178: MRC1|P22897
cor=0.17, p=0.0065
2540: CDH3|P22223
1438: CDH3|P22223
cor=0.61, p=1.5e−31
2542: GDI2|P50395
418: GDI2|P50395
cor=0.69, p=2.1e−35
2544: TNFRSF1A|P19438
1520: TNFRSF1A|P19438
cor=0.43, p=9.5e−15
2547: DCN|P07585
483: DCN|P07585
cor=0.29, p=0.00011
2548: CAPN1|P07384
1841: CAPN1|P07384
cor=0.48, p=4.4e−09
2551: HGF|P14210
1954: HGF|P14210
cor=0.32, p=3.4e−06
2552: HSPD1|P10809
797: HSPD1|P10809
cor=0.7, p=6.8e−45
2556: MIA|Q16674
1196: MIA|Q16674
cor=0.44, p=1.5e−11
2558: PF4|P02776
2101: PF4|P02776
cor=0.53, p=7.7e−23
2559: PROS1|P07225
167: PROS1|P07225
cor=0.49, p=2.8e−19
2560: SERPINA7|P05543
247: SERPINA7|P05543
cor=0.48, p=1.8e−18
2566: CCN3|P48745
451: CCN3|P48745
cor=0.86, p=6.8e−88
2569: SHH|Q15465
1158: SHH|Q15465
cor=0.79, p=2e−64
2573: APOA1|P02647
58: APOA1|P02647
cor=0.3, p=1.4e−07
2575: C1QA|P02745
786: C1QA|P02745
cor=0.2, p=0.055
2590: MMP3|P08254
1960: MMP3|P08254
cor=0.23, p=0.00015
2593: SOD1|P00441
349: SOD1|P00441
cor=0.47, p=1.8e−14
2595: APOB|P04114
17: APOB|P04114
cor=0.75, p=1e−54
2601: CDH5|P33151
552: CDH5|P33151
cor=0.55, p=1e−24
2605: SPINT1|O43278
1136: SPINT1|O43278
cor=0.72, p=5e−42
2610: LCN2|P80188
1350: LCN2|P80188
cor=0.047, p=0.66
2612: MMP17|Q9ULZ9
913: MMP17|Q9ULZ9
cor=0.58, p=5.3e−28
2614: SPINT2|O43291
1640: SPINT2|O43291
cor=0.71, p=1.1e−46
2615: TIE1|P35590
929: TIE1|P35590
cor=0.52, p=9.1e−12
2620: MAPK3|P27361
1992: MAPK3|P27361
cor=0.32, p=1.7e−07
2623: RAC1|P63000
1957: RAC1|P63000
cor=0.72, p=1.7e−46
2629: CCL14|Q16627
1535: CCL14|Q16627
cor=0.62, p=1.8e−28
2630: MDK|P21741
1403: MDK|P21741
cor=0.59, p=1.6e−17
2632: SERPINE1|P05121
1683: SERPINE1|P05121
cor=0.63, p=3.8e−34
2641: IGFBP4|P22692
521: IGFBP4|P22692
cor=0.77, p=2.5e−59
2645: CFP|P27918
793: CFP|P27918
cor=0.17, p=0.013
2647: PTHLH|P12272
1773: PTHLH|P12272
cor=0.49, p=1.7e−14
2651: BMP7|P18075
1574: BMP7|P18075
cor=0.19, p=0.001
2653: CNTN1|Q12860
25: CNTN1|Q12860
cor=0.14, p=0.024
2654: CCN2|P29279
1032: CCN2|P29279
cor=0.67, p=6.3e−40
2663: JAM2|P57087
1147: JAM2|P57087
cor=0.69, p=3.4e−41
2664: JAM3|Q9BX67
1482: JAM3|Q9BX67

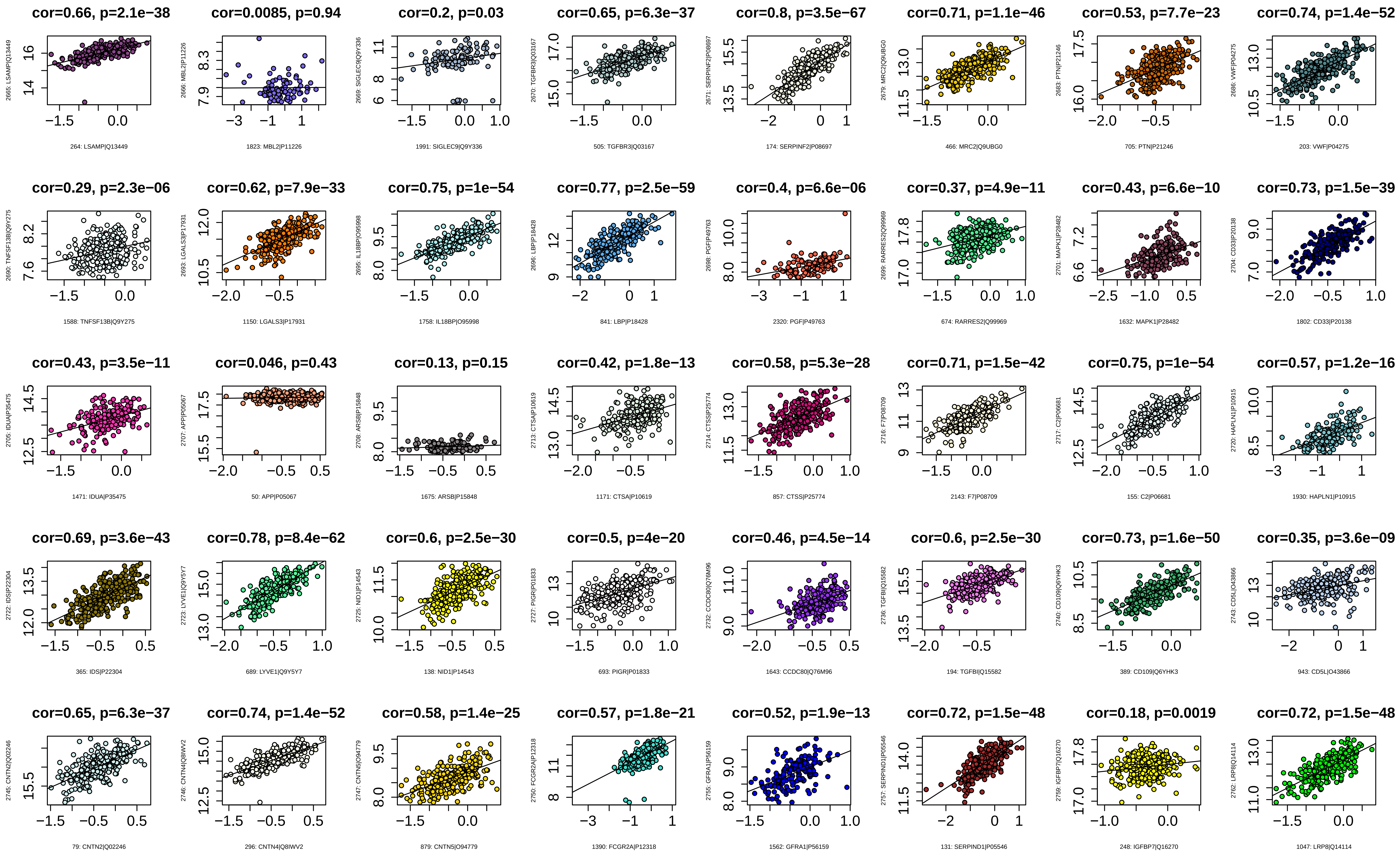
cor=0.66, p=2.1e−38
2665: LSAMP|Q13449
264: LSAMP|Q13449
cor=0.0085, p=0.94
2666: MBL2|P11226
1823: MBL2|P11226
cor=0.2, p=0.03
2669: SIGLEC9|Q9Y336
1991: SIGLEC9|Q9Y336
cor=0.65, p=6.3e−37
2670: TGFBR3|Q03167
505: TGFBR3|Q03167
cor=0.8, p=3.5e−67
2671: SERPINF2|P08697
174: SERPINF2|P08697
cor=0.71, p=1.1e−46
2679: MRC2|Q9UBG0
466: MRC2|Q9UBG0
cor=0.53, p=7.7e−23
2683: PTN|P21246
705: PTN|P21246
cor=0.74, p=1.4e−52
2686: VWF|P04275
203: VWF|P04275
cor=0.29, p=2.3e−06
2690: TNFSF13B|Q9Y275
1588: TNFSF13B|Q9Y275
cor=0.62, p=7.9e−33
2693: LGALS3|P17931
1150: LGALS3|P17931
cor=0.75, p=1e−54
2695: IL18BP|O95998
1758: IL18BP|O95998
cor=0.77, p=2.5e−59
2696: LBP|P18428
841: LBP|P18428
cor=0.4, p=6.6e−06
2698: PGF|P49763
2320: PGF|P49763
cor=0.37, p=4.9e−11
2699: RARRES2|Q99969
674: RARRES2|Q99969
cor=0.43, p=6.6e−10
2701: MAPK1|P28482
1632: MAPK1|P28482
cor=0.73, p=1.5e−39
2704: CD33|P20138
1802: CD33|P20138
cor=0.43, p=3.5e−11
2705: IDUA|P35475
1471: IDUA|P35475
cor=0.046, p=0.43
2707: APP|P05067
50: APP|P05067
cor=0.13, p=0.15
2708: ARSB|P15848
1675: ARSB|P15848
cor=0.42, p=1.8e−13
2713: CTSA|P10619
1171: CTSA|P10619
cor=0.58, p=5.3e−28
2714: CTSS|P25774
857: CTSS|P25774
cor=0.71, p=1.5e−42
2716: F7|P08709
2143: F7|P08709
cor=0.75, p=1e−54
2717: C2|P06681
155: C2|P06681
cor=0.57, p=1.2e−16
2720: HAPLN1|P10915
1930: HAPLN1|P10915
cor=0.69, p=3.6e−43
2722: IDS|P22304
365: IDS|P22304
cor=0.78, p=8.4e−62
2723: LYVE1|Q9Y5Y7
689: LYVE1|Q9Y5Y7
cor=0.6, p=2.5e−30
2725: NID1|P14543
138: NID1|P14543
cor=0.5, p=4e−20
2727: PIGR|P01833
693: PIGR|P01833
cor=0.46, p=4.5e−14
2732: CCDC80|Q76M96
1643: CCDC80|Q76M96
cor=0.6, p=2.5e−30
2736: TGFBI|Q15582
194: TGFBI|Q15582
cor=0.73, p=1.6e−50
2740: CD109|Q6YHK3
389: CD109|Q6YHK3
cor=0.35, p=3.6e−09
2743: CD5L|O43866
943: CD5L|O43866
cor=0.65, p=6.3e−37
2745: CNTN2|Q02246
79: CNTN2|Q02246
cor=0.74, p=1.4e−52
2746: CNTN4|Q8IWV2
296: CNTN4|Q8IWV2
cor=0.58, p=1.4e−25
2747: CNTN5|O94779
879: CNTN5|O94779
cor=0.57, p=1.8e−21
2750: FCGR2A|P12318
1390: FCGR2A|P12318
cor=0.52, p=1.9e−13
2755: GFRA1|P56159
1562: GFRA1|P56159
cor=0.72, p=1.5e−48
2757: SERPIND1|P05546
131: SERPIND1|P05546
cor=0.18, p=0.0019
2759: IGFBP7|Q16270
248: IGFBP7|Q16270
cor=0.72, p=1.5e−48
2762: LRP8|Q14114
1047: LRP8|Q14114

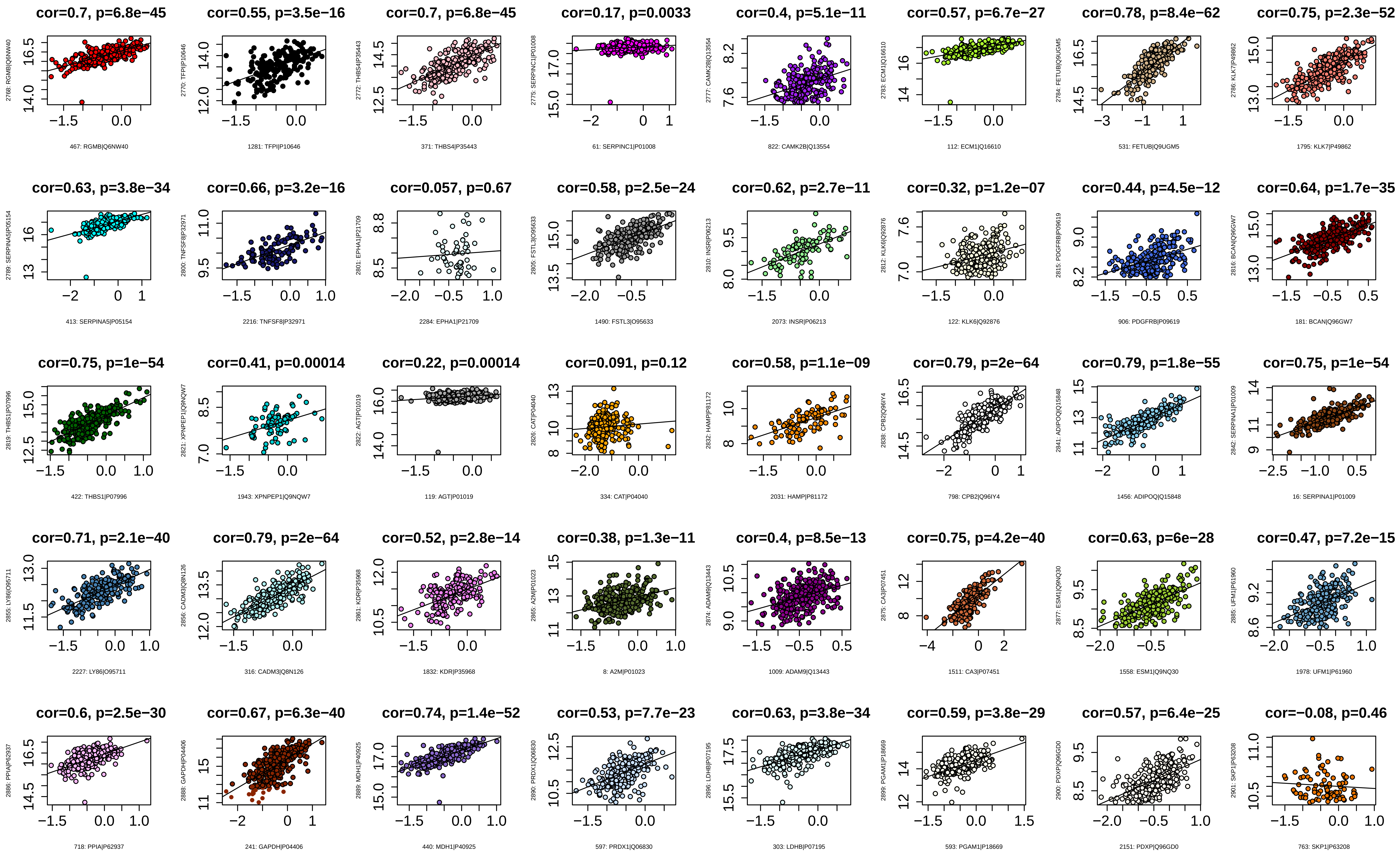
cor=0.7, p=6.8e−45
2768: RGMB|Q6NW40
467: RGMB|Q6NW40
cor=0.55, p=3.5e−16
2770: TFPI|P10646
1281: TFPI|P10646
cor=0.7, p=6.8e−45
2772: THBS4|P35443
371: THBS4|P35443
cor=0.17, p=0.0033
2775: SERPINC1|P01008
61: SERPINC1|P01008
cor=0.4, p=5.1e−11
2777: CAMK2B|Q13554
822: CAMK2B|Q13554
cor=0.57, p=6.7e−27
2783: ECM1|Q16610
112: ECM1|Q16610
cor=0.78, p=8.4e−62
2784: FETUB|Q9UGM5
531: FETUB|Q9UGM5
cor=0.75, p=2.3e−52
2786: KLK7|P49862
1795: KLK7|P49862
cor=0.63, p=3.8e−34
2789: SERPINA5|P05154
413: SERPINA5|P05154
cor=0.66, p=3.2e−16
2800: TNFSF8|P32971
2216: TNFSF8|P32971
cor=0.057, p=0.67
2801: EPHA1|P21709
2284: EPHA1|P21709
cor=0.58, p=2.5e−24
2805: FSTL3|O95633
1490: FSTL3|O95633
cor=0.62, p=2.7e−11
2810: INSR|P06213
2073: INSR|P06213
cor=0.32, p=1.2e−07
2812: KLK6|Q92876
122: KLK6|Q92876
cor=0.44, p=4.5e−12
2815: PDGFRB|P09619
906: PDGFRB|P09619
cor=0.64, p=1.7e−35
2816: BCAN|Q96GW7
181: BCAN|Q96GW7
cor=0.75, p=1e−54
2819: THBS1|P07996
422: THBS1|P07996
cor=0.41, p=0.00014
2821: XPNPEP1|Q9NQW7
1943: XPNPEP1|Q9NQW7
cor=0.22, p=0.00014
2822: AGT|P01019
119: AGT|P01019
cor=0.091, p=0.12
2826: CAT|P04040
334: CAT|P04040
cor=0.58, p=1.1e−09
2832: HAMP|P81172
2031: HAMP|P81172
cor=0.79, p=2e−64
2838: CPB2|Q96IY4
798: CPB2|Q96IY4
cor=0.79, p=1.8e−55
2841: ADIPOQ|Q15848
1456: ADIPOQ|Q15848
cor=0.75, p=1e−54
2842: SERPINA1|P01009
16: SERPINA1|P01009
cor=0.71, p=2.1e−40
2855: LY86|O95711
2227: LY86|O95711
cor=0.79, p=2e−64
2856: CADM3|Q8N126
316: CADM3|Q8N126
cor=0.52, p=2.8e−14
2861: KDR|P35968
1832: KDR|P35968
cor=0.38, p=1.3e−11
2865: A2M|P01023
8: A2M|P01023
cor=0.4, p=8.5e−13
2874: ADAM9|Q13443
1009: ADAM9|Q13443
cor=0.75, p=4.2e−40
2875: CA3|P07451
1511: CA3|P07451
cor=0.63, p=6e−28
2877: ESM1|Q9NQ30
1558: ESM1|Q9NQ30
cor=0.47, p=7.2e−15
2885: UFM1|P61960
1978: UFM1|P61960
cor=0.6, p=2.5e−30
2886: PPIA|P62937
718: PPIA|P62937
cor=0.67, p=6.3e−40
2888: GAPDH|P04406
241: GAPDH|P04406
cor=0.74, p=1.4e−52
2889: MDH1|P40925
440: MDH1|P40925
cor=0.53, p=7.7e−23
2890: PRDX1|Q06830
597: PRDX1|Q06830
cor=0.63, p=3.8e−34
2896: LDHB|P07195
303: LDHB|P07195
cor=0.59, p=3.8e−29
2899: PGAM1|P18669
593: PGAM1|P18669
cor=0.57, p=6.4e−25
2900: PDXP|Q96GD0
2151: PDXP|Q96GD0
cor=−0.08, p=0.46
2901: SKP1|P63208
763: SKP1|P63208

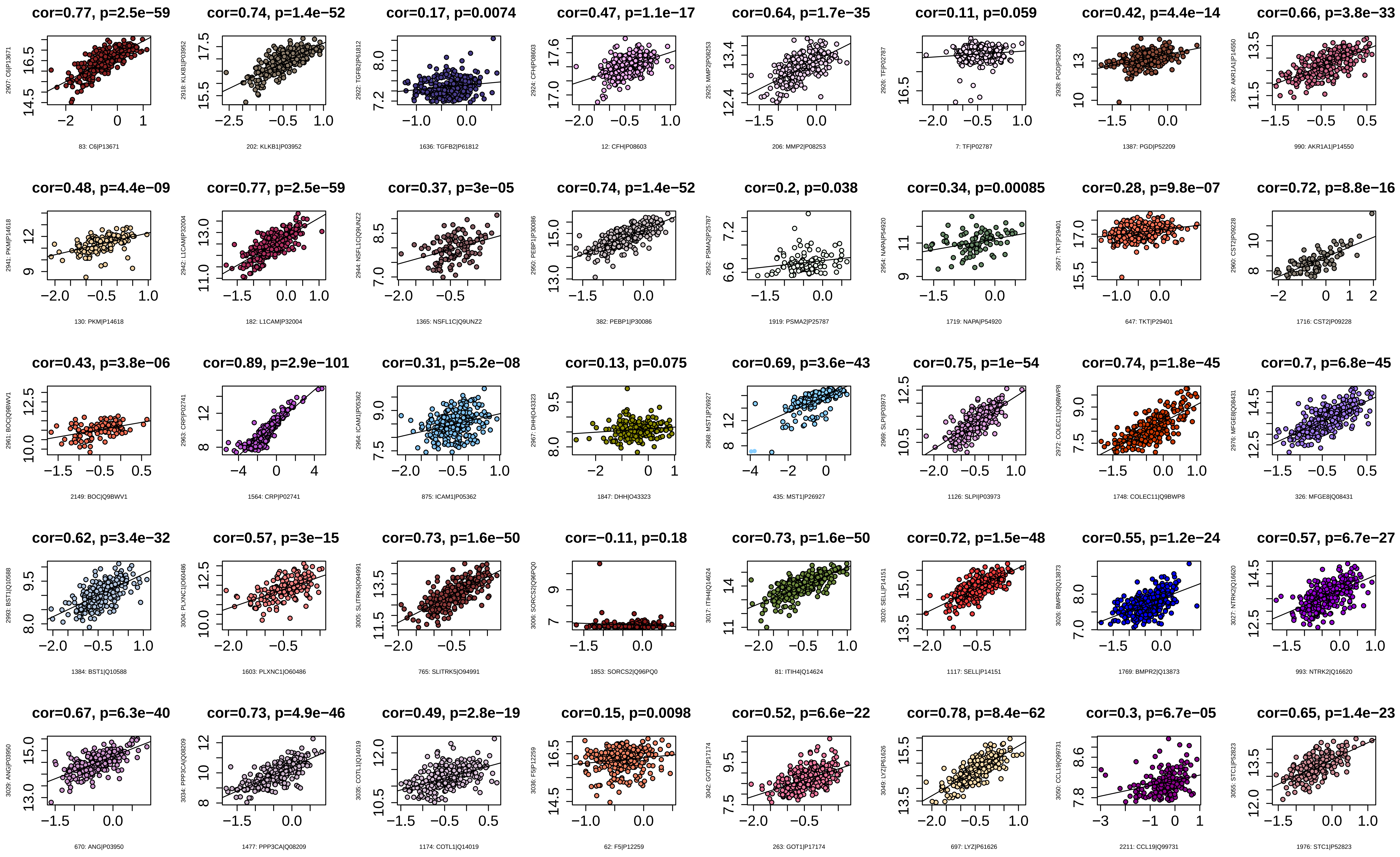

cor=0.77, p=2.5e−59
2907: C6|P13671
83: C6|P13671
cor=0.74, p=1.4e−52
2918: KLKB1|P03952
202: KLKB1|P03952
cor=0.17, p=0.0074
2922: TGFB2|P61812
1636: TGFB2|P61812
cor=0.47, p=1.1e−17
2924: CFH|P08603
12: CFH|P08603
cor=0.64, p=1.7e−35
2925: MMP2|P08253
206: MMP2|P08253
cor=0.11, p=0.059
2926: TF|P02787
7: TF|P02787
cor=0.42, p=4.4e−14
2928: PGD|P52209
1387: PGD|P52209
cor=0.66, p=3.8e−33
2930: AKR1A1|P14550
990: AKR1A1|P14550
cor=0.48, p=4.4e−09
2941: PKM|P14618
130: PKM|P14618
cor=0.77, p=2.5e−59
2942: L1CAM|P32004
182: L1CAM|P32004
cor=0.37, p=3e−05
2944: NSFL1C|Q9UNZ2
1365: NSFL1C|Q9UNZ2
cor=0.74, p=1.4e−52
2950: PEBP1|P30086
382: PEBP1|P30086
cor=0.2, p=0.038
2952: PSMA2|P25787
1919: PSMA2|P25787
cor=0.34, p=0.00085
2954: NAPA|P54920
1719: NAPA|P54920
cor=0.28, p=9.8e−07
2957: TKT|P29401
647: TKT|P29401
cor=0.72, p=8.8e−16
2960: CST2|P09228
1716: CST2|P09228
cor=0.43, p=3.8e−06
2961: BOC|Q9BWV1
2149: BOC|Q9BWV1
cor=0.89, p=2.9e−101
2963: CRP|P02741
1564: CRP|P02741
cor=0.31, p=5.2e−08
2964: ICAM1|P05362
875: ICAM1|P05362
cor=0.13, p=0.075
2967: DHH|O43323
1847: DHH|O43323
cor=0.69, p=3.6e−43
2968: MST1|P26927
435: MST1|P26927
cor=0.75, p=1e−54
2969: SLPI|P03973
1126: SLPI|P03973
cor=0.74, p=1.8e−45
2972: COLEC11|Q9BWP8
1748: COLEC11|Q9BWP8
cor=0.7, p=6.8e−45
2976: MFGE8|Q08431
326: MFGE8|Q08431
cor=0.62, p=3.4e−32
2993: BST1|Q10588
1384: BST1|Q10588
cor=0.57, p=3e−15
3004: PLXNC1|O60486
1603: PLXNC1|O60486
cor=0.73, p=1.6e−50
3005: SLITRK5|O94991
765: SLITRK5|O94991
cor=−0.11, p=0.18
3006: SORCS2|Q96PQ0
1853: SORCS2|Q96PQ0
cor=0.73, p=1.6e−50
3017: ITIH4|Q14624
81: ITIH4|Q14624
cor=0.72, p=1.5e−48
3020: SELL|P14151
1117: SELL|P14151
cor=0.55, p=1.2e−24
3026: BMPR2|Q13873
1769: BMPR2|Q13873
cor=0.57, p=6.7e−27
3027: NTRK2|Q16620
993: NTRK2|Q16620
cor=0.67, p=6.3e−40
3029: ANG|P03950
670: ANG|P03950
cor=0.73, p=4.9e−46
3034: PPP3CA|Q08209
1477: PPP3CA|Q08209
cor=0.49, p=2.8e−19
3035: COTL1|Q14019
1174: COTL1|Q14019
cor=0.15, p=0.0098
3036: F5|P12259
62: F5|P12259
cor=0.52, p=6.6e−22
3042: GOT1|P17174
263: GOT1|P17174
cor=0.78, p=8.4e−62
3049: LYZ|P61626
697: LYZ|P61626
cor=0.3, p=6.7e−05
3050: CCL19|Q99731
2211: CCL19|Q99731
cor=0.65, p=1.4e−23
3055: STC1|P52823
1976: STC1|P52823

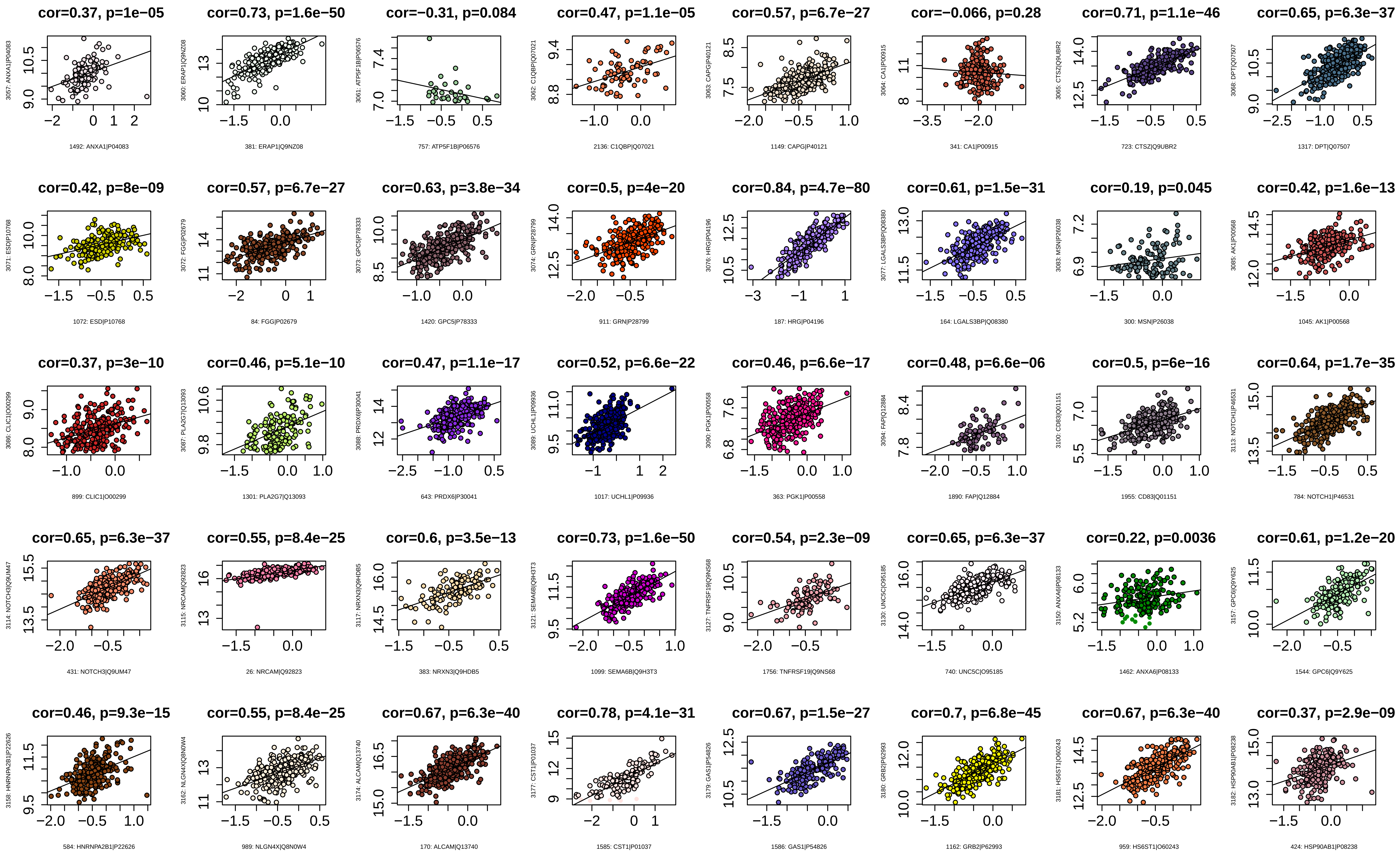
cor=0.37, p=1e−05
1492: ANXA1|P04083
cor=0.73, p=1.6e−50
381: ERAP1|Q9NZ08
cor=−0.31, p=0.084
757: ATP5F1B|P06576
cor=0.47, p=1.1e−05
2136: C1QBP|Q07021
cor=0.57, p=6.7e−27
1149: CAPG|P40121
cor=−0.066, p=0.28
341: CA1|P00915
cor=0.71, p=1.1e−46
723: CTSZ|Q9UBR2
cor=0.65, p=6.3e−37
1317: DPT|Q07507
cor=0.42, p=8e−09
1072: ESD|P10768
cor=0.57, p=6.7e−27
84: FGG|P02679
cor=0.63, p=3.8e−34
1420: GPC5|P78333
cor=0.5, p=4e−20
911: GRN|P28799
cor=0.84, p=4.7e−80
187: HRG|P04196
cor=0.61, p=1.5e−31
164: LGALS3BP|Q08380
cor=0.19, p=0.045
300: MSN|P26038
cor=0.42, p=1.6e−13
1045: AK1|P00568
cor=0.37, p=3e−10
899: CLIC1|O00299
cor=0.46, p=5.1e−10
1301: PLA2G7|Q13093
cor=0.47, p=1.1e−17
643: PRDX6|P30041
cor=0.52, p=6.6e−22
1017: UCHL1|P09936
cor=0.46, p=6.6e−17
363: PGK1|P00558
cor=0.48, p=6.6e−06
1890: FAP|Q12884
cor=0.5, p=6e−16
1955: CD83|Q01151
cor=0.64, p=1.7e−35
784: NOTCH1|P46531
cor=0.65, p=6.3e−37
431: NOTCH3|Q9UM47
cor=0.55, p=8.4e−25
26: NRCAM|Q92823
cor=0.6, p=3.5e−13
383: NRXN3|Q9HDB5
cor=0.73, p=1.6e−50
1099: SEMA6B|Q9H3T3
cor=0.54, p=2.3e−09
1756: TNFRSF19|Q9NS68
cor=0.65, p=6.3e−37
740: UNC5C|O95185
cor=0.22, p=0.0036
1462: ANXA6|P08133
cor=0.61, p=1.2e−20
1544: GPC6|Q9Y625
cor=0.46, p=9.3e−15
584: HNRNPA2B1|P22626
cor=0.55, p=8.4e−25
989: NLGN4X|Q8N0W4
cor=0.67, p=6.3e−40
170: ALCAM|Q13740
cor=0.78, p=4.1e−31
1585: CST1|P01037
cor=0.67, p=1.5e−27
1586: GAS1|P54826
cor=0.7, p=6.8e−45
1162: GRB2|P62993
cor=0.67, p=6.3e−40
959: HS6ST1|O60243
cor=0.37, p=2.9e−09
424: HSP90AB1|P08238

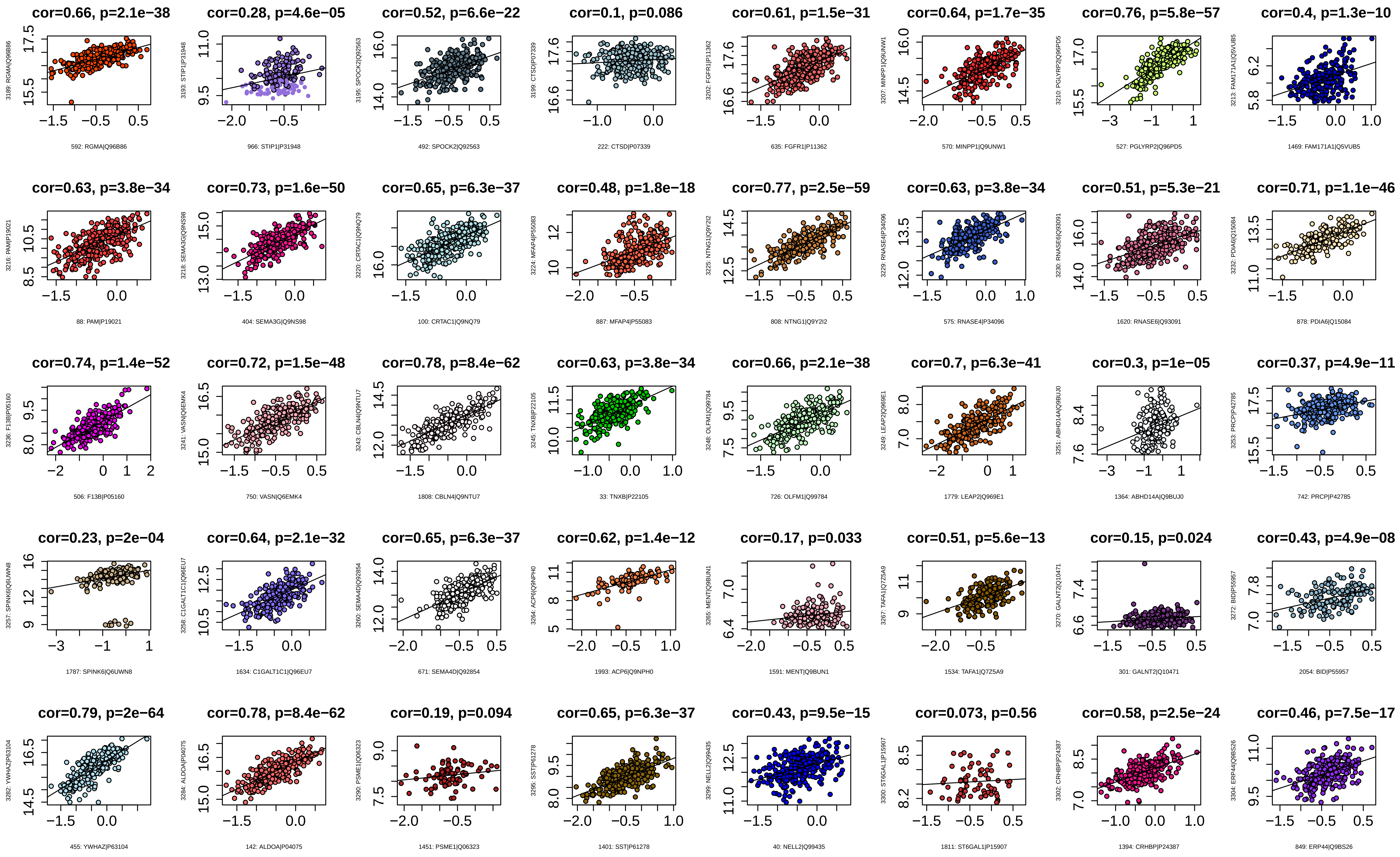
cor=0.66, p=2.1e−38
592: RGMA|Q96B86
cor=0.28, p=4.6e−05
966: STIP1|P31948
cor=0.52, p=6.6e−22
492: SPOCK2|Q92563
cor=0.1, p=0.086
222: CTSD|P07339
cor=0.61, p=1.5e−31
635: FGFR1|P11362
cor=0.64, p=1.7e−35
570: MINPP1|Q9UNW1
cor=0.76, p=5.8e−57
527: PGLYRP2|Q96PD5
cor=0.4, p=1.3e−10
1469: FAM171A1|Q5VUB5
cor=0.63, p=3.8e−34
88: PAM|P19021
cor=0.73, p=1.6e−50
404: SEMA3G|Q9NS98
cor=0.65, p=6.3e−37
100: CRTAC1|Q9NQ79
cor=0.48, p=1.8e−18
887: MFAP4|P55083
cor=0.77, p=2.5e−59
808: NTNG1|Q9Y2I2
cor=0.63, p=3.8e−34
575: RNASE4|P34096
cor=0.51, p=5.3e−21
1620: RNASE6|Q93091
cor=0.71, p=1.1e−46
878: PDIA6|Q15084
cor=0.74, p=1.4e−52
506: F13B|P05160
cor=0.72, p=1.5e−48
750: VASN|Q6EMK4
cor=0.78, p=8.4e−62
1808: CBLN4|Q9NTU7
cor=0.63, p=3.8e−34
33: TNXB|P22105
cor=0.66, p=2.1e−38
726: OLFM1|Q99784
cor=0.7, p=6.3e−41
1779: LEAP2|Q969E1
cor=0.3, p=1e−05
1364: ABHD14A|Q9BUJ0
cor=0.37, p=4.9e−11
742: PRCP|P42785
cor=0.23, p=2e−04
1787: SPINK6|Q6UWN8
cor=0.64, p=2.1e−32
1634: C1GALT1C1|Q96EU7
cor=0.65, p=6.3e−37
671: SEMA4D|Q92854
cor=0.62, p=1.4e−12
1993: ACP6|Q9NPH0
cor=0.17, p=0.033
1591: MENT|Q9BUN1
cor=0.51, p=5.6e−13
1534: TAFA1|Q7Z5A9
cor=0.15, p=0.024
301: GALNT2|Q10471
cor=0.43, p=4.9e−08
2054: BID|P55957
cor=0.79, p=2e−64
455: YWHAZ|P63104
cor=0.78, p=8.4e−62
142: ALDOA|P04075
cor=0.19, p=0.094
1451: PSME1|Q06323
cor=0.65, p=6.3e−37
1401: SST|P61278
cor=0.43, p=9.5e−15
40: NELL2|Q99435
cor=0.073, p=0.56
1811: ST6GAL1|P15907
cor=0.58, p=2.5e−24
1394: CRHBP|P24387
cor=0.46, p=7.5e−17
849: ERP44|Q9BS26

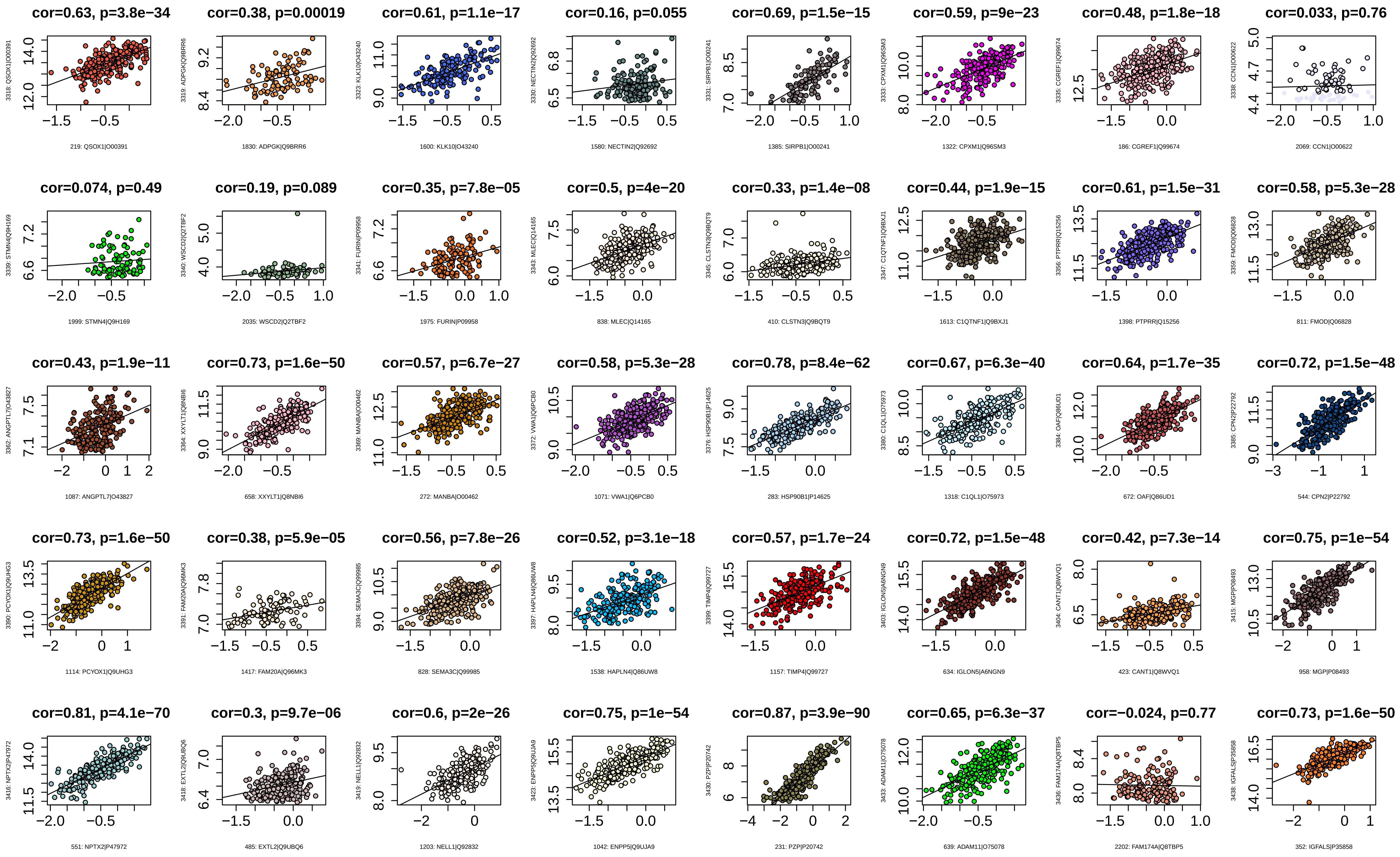
cor=0.63, p=3.8e−34
3318: QSOX1|O00391
219: QSOX1|O00391
cor=0.38, p=0.00019
3319: ADPGK|Q9BRR6
1830: ADPGK|Q9BRR6
cor=0.61, p=1.1e−17
3323: KLK10|O43240
1600: KLK10|O43240
cor=0.16, p=0.055
3330: NECTIN2|Q92692
1580: NECTIN2|Q92692
cor=0.69, p=1.5e−15
3331: SIRPB1|O00241
1385: SIRPB1|O00241
cor=0.59, p=9e−23
3333: CPXM1|Q96SM3
1322: CPXM1|Q96SM3
cor=0.48, p=1.8e−18
3335: CGREF1|Q99674
186: CGREF1|Q99674
cor=0.033, p=0.76
3338: CCN1|O00622
2069: CCN1|O00622
cor=0.074, p=0.49
3339: STMN4|Q9H169
1999: STMN4|Q9H169
cor=0.19, p=0.089
3340: WSCD2|Q2TBF2
2035: WSCD2|Q2TBF2
cor=0.35, p=7.8e−05
3341: FURIN|P09958
1975: FURIN|P09958
cor=0.5, p=4e−20
3343: MLEC|Q14165
838: MLEC|Q14165
cor=0.33, p=1.4e−08
3345: CLSTN3|Q9BQT9
410: CLSTN3|Q9BQT9
cor=0.44, p=1.9e−15
3347: C1QTNF1|Q9BXJ1
1613: C1QTNF1|Q9BXJ1
cor=0.61, p=1.5e−31
3356: PTPRR|Q15256
1398: PTPRR|Q15256
cor=0.58, p=5.3e−28
3359: FMOD|Q06828
811: FMOD|Q06828
cor=0.43, p=1.9e−11
3362: ANGPTL7|O43827
1087: ANGPTL7|O43827
cor=0.73, p=1.6e−50
3364: XXYLT1|Q8NBI6
658: XXYLT1|Q8NBI6
cor=0.57, p=6.7e−27
3369: MANBA|O00462
272: MANBA|O00462
cor=0.58, p=5.3e−28
3372: VWA1|Q6PCB0
1071: VWA1|Q6PCB0
cor=0.78, p=8.4e−62
3376: HSP90B1|P14625
283: HSP90B1|P14625
cor=0.67, p=6.3e−40
3380: C1QL1|O75973
1318: C1QL1|O75973
cor=0.64, p=1.7e−35
3384: OAF|Q86UD1
672: OAF|Q86UD1
cor=0.72, p=1.5e−48
3385: CPN2|P22792
544: CPN2|P22792
cor=0.73, p=1.6e−50
3390: PCYOX1|Q9UHG3
1114: PCYOX1|Q9UHG3
cor=0.38, p=5.9e−05
3391: FAM20A|Q96MK3
1417: FAM20A|Q96MK3
cor=0.56, p=7.8e−26
3394: SEMA3C|Q99985
828: SEMA3C|Q99985
cor=0.52, p=3.1e−18
3397: HAPLN4|Q86UW8
1538: HAPLN4|Q86UW8
cor=0.57, p=1.7e−24
3399: TIMP4|Q99727
1157: TIMP4|Q99727
cor=0.72, p=1.5e−48
3403: IGLON5|A6NGN9
634: IGLON5|A6NGN9
cor=0.42, p=7.3e−14
3404: CANT1|Q8WVQ1
423: CANT1|Q8WVQ1
cor=0.75, p=1e−54
3415: MGP|P08493
958: MGP|P08493
cor=0.81, p=4.1e−70
3416: NPTX2|P47972
551: NPTX2|P47972
cor=0.3, p=9.7e−06
3418: EXTL2|Q9UBQ6
485: EXTL2|Q9UBQ6
cor=0.6, p=2e−26
3419: NELL1|Q92832
1203: NELL1|Q92832
cor=0.75, p=1e−54
3423: ENPP5|Q9UJA9
1042: ENPP5|Q9UJA9
cor=0.87, p=3.9e−90
3430: PZP|P20742
231: PZP|P20742
cor=0.65, p=6.3e−37
3433: ADAM11|O75078
639: ADAM11|O75078
cor=−0.024, p=0.77
3436: FAM174A|Q8TBP5
2202: FAM174A|Q8TBP5
cor=0.73, p=1.6e−50
3438: IGFALS|P35858
352: IGFALS|P35858

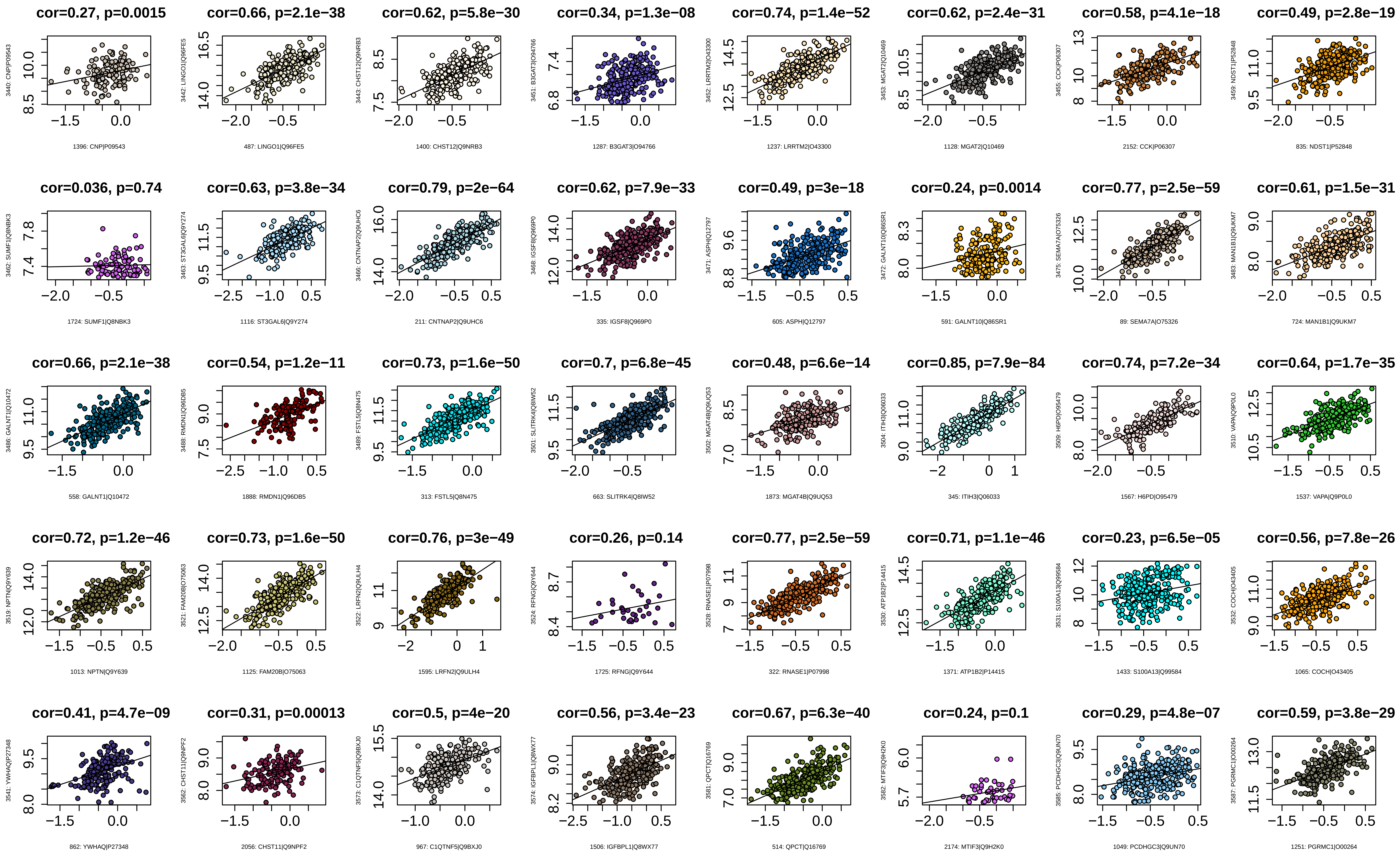
cor=0.27, p=0.0015
1396: CNP|P09543
cor=0.66, p=2.1e−38
487: LINGO1|Q96FE5
cor=0.62, p=5.8e−30
1400: CHST12|Q9NRB3
cor=0.34, p=1.3e−08
1287: B3GAT3|O94766
cor=0.74, p=1.4e−52
1237: LRRTM2|O43300
cor=0.62, p=2.4e−31
1128: MGAT2|Q10469
cor=0.58, p=4.1e−18
2152: CCK|P06307
cor=0.49, p=2.8e−19
835: NDST1|P52848
cor=0.036, p=0.74
1724: SUMF1|Q8NBK3
cor=0.63, p=3.8e−34
1116: ST3GAL6|Q9Y274
cor=0.79, p=2e−64
211: CNTNAP2|Q9UHC6
cor=0.62, p=7.9e−33
335: IGSF8|Q969P0
cor=0.49, p=3e−18
605: ASPH|Q12797
cor=0.24, p=0.0014
591: GALNT10|Q86SR1
cor=0.77, p=2.5e−59
89: SEMA7A|O75326
cor=0.61, p=1.5e−31
724: MAN1B1|Q9UKM7
cor=0.66, p=2.1e−38
558: GALNT1|Q10472
cor=0.54, p=1.2e−11
1888: RMDN1|Q96DB5
cor=0.73, p=1.6e−50
313: FSTL5|Q8N475
cor=0.7, p=6.8e−45
663: SLITRK4|Q8IW52
cor=0.48, p=6.6e−14
1873: MGAT4B|Q9UQ53
cor=0.85, p=7.9e−84
345: ITIH3|Q06033
cor=0.74, p=7.2e−34
1567: H6PD|O95479
cor=0.64, p=1.7e−35
1537: VAPA|Q9P0L0
cor=0.72, p=1.2e−46
1013: NPTN|Q9Y639
cor=0.73, p=1.6e−50
1125: FAM20B|O75063
cor=0.76, p=3e−49
1595: LRFN2|Q9ULH4
cor=0.26, p=0.14
1725: RFNG|Q9Y644
cor=0.77, p=2.5e−59
322: RNASE1|P07998
cor=0.71, p=1.1e−46
1371: ATP1B2|P14415
cor=0.23, p=6.5e−05
1433: S100A13|Q99584
cor=0.56, p=7.8e−26
1065: COCH|O43405
cor=0.41, p=4.7e−09
862: YWHAQ|P27348
cor=0.31, p=0.00013
2056: CHST11|Q9NPF2
cor=0.5, p=4e−20
967: C1QTNF5|Q9BXJ0
cor=0.56, p=3.4e−23
1506: IGFBPL1|Q8WX77
cor=0.67, p=6.3e−40
514: QPCT|Q16769
cor=0.24, p=0.1
2174: MTIF3|Q9H2K0
cor=0.29, p=4.8e−07
1049: PCDHGC3|Q9UN70
cor=0.59, p=3.8e−29
1251: PGRMC1|O00264

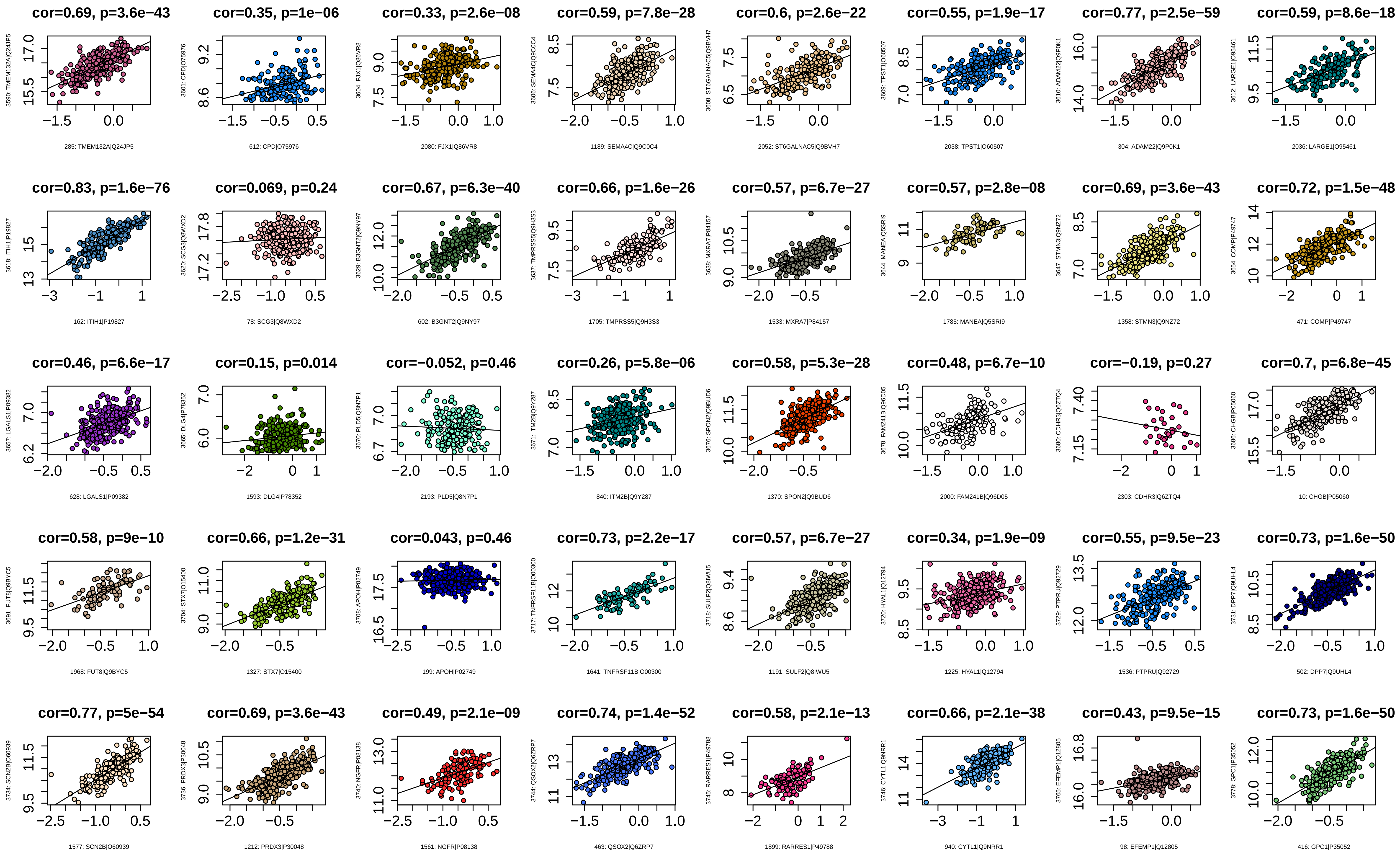
cor=0.69, p=3.6e−43
285: TMEM132A|Q24JP5
cor=0.35, p=1e−06
612: CPD|O75976
cor=0.33, p=2.6e−08
2080: FJX1|Q86VR8
cor=0.59, p=7.8e−28
1189: SEMA4C|Q9C0C4
cor=0.6, p=2.6e−22
2052: ST6GALNAC5|Q9BVH7
cor=0.55, p=1.9e−17
2038: TPST1|O60507
cor=0.77, p=2.5e−59
304: ADAM22|Q9P0K1
cor=0.59, p=8.6e−18
2036: LARGE1|O95461
cor=0.83, p=1.6e−76
162: ITIH1|P19827
cor=0.069, p=0.24
78: SCG3|Q8WXD2
cor=0.67, p=6.3e−40
602: B3GNT2|Q9NY97
cor=0.66, p=1.6e−26
1705: TMPRSS5|Q9H3S3
cor=0.57, p=6.7e−27
1533: MXRA7|P84157
cor=0.57, p=2.8e−08
1785: MANEA|Q5SRI9
cor=0.69, p=3.6e−43
1358: STMN3|Q9NZ72
cor=0.72, p=1.5e−48
471: COMP|P49747
cor=0.46, p=6.6e−17
628: LGALS1|P09382
cor=0.15, p=0.014
1593: DLG4|P78352
cor=−0.052, p=0.46
2193: PLD5|Q8N7P1
cor=0.26, p=5.8e−06
840: ITM2B|Q9Y287
cor=0.58, p=5.3e−28
1370: SPON2|Q9BUD6
cor=0.48, p=6.7e−10
2000: FAM241B|Q96D05
cor=−0.19, p=0.27
2303: CDHR3|Q6ZTQ4
cor=0.7, p=6.8e−45
10: CHGB|P05060
cor=0.58, p=9e−10
1968: FUT8|Q9BYC5
cor=0.66, p=1.2e−31
1327: STX7|O15400
cor=0.043, p=0.46
199: APOH|P02749
cor=0.73, p=2.2e−17
1641: TNFRSF11B|O00300
cor=0.57, p=6.7e−27
1191: SULF2|Q8IWU5
cor=0.34, p=1.9e−09
1225: HYAL1|Q12794
cor=0.55, p=9.5e−23
1536: PTPRU|Q92729
cor=0.73, p=1.6e−50
502: DPP7|Q9UHL4
cor=0.77, p=5e−54
1577: SCN2B|O60939
cor=0.69, p=3.6e−43
1212: PRDX3|P30048
cor=0.49, p=2.1e−09
1561: NGFR|P08138
cor=0.74, p=1.4e−52
463: QSOX2|Q6ZRP7
cor=0.58, p=2.1e−13
1899: RARRES1|P49788
cor=0.66, p=2.1e−38
940: CYTL1|Q9NRR1
cor=0.43, p=9.5e−15
98: EFEMP1|Q12805
cor=0.73, p=1.6e−50
416: GPC1|P35052

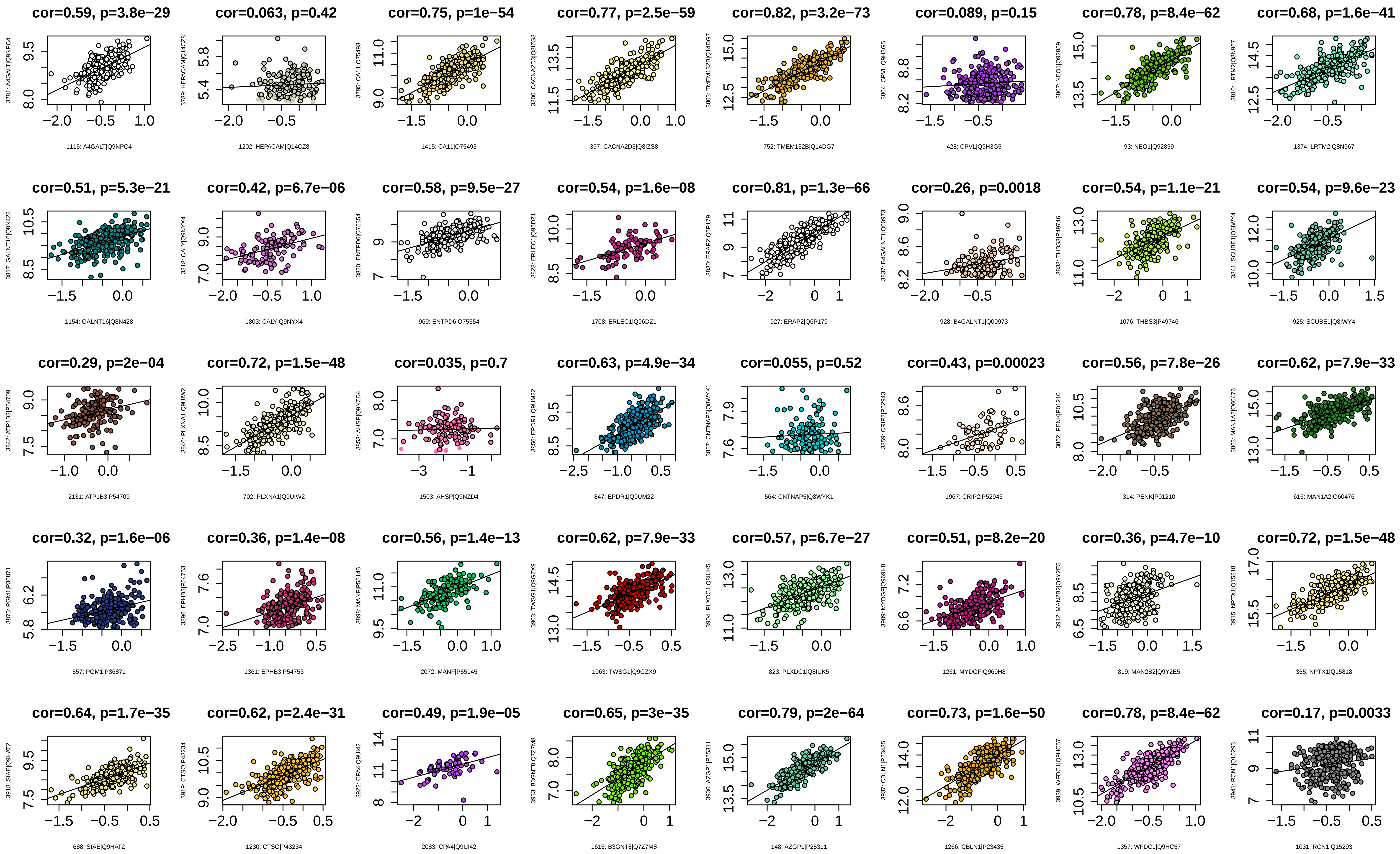
cor=0.59, p=3.8e−29
3781: A4GALT|Q9NPC4
1115: A4GALT|Q9NPC4
cor=0.063, p=0.42
3789: HEPACAM|Q14CZ8
1202: HEPACAM|Q14CZ8
cor=0.75, p=1e−54
3795: CA11|O75493
1415: CA11|O75493
cor=0.77, p=2.5e−59
3800: CACNA2D3|Q8IZS8
397: CACNA2D3|Q8IZS8
cor=0.82, p=3.2e−73
3803: TMEM132B|Q14DG7
752: TMEM132B|Q14DG7
cor=0.089, p=0.15
3804: CPVL|Q9H3G5
428: CPVL|Q9H3G5
cor=0.78, p=8.4e−62
3807: NEO1|Q92859
93: NEO1|Q92859
cor=0.68, p=1.6e−41
3810: LRTM2|Q8N967
1374: LRTM2|Q8N967
cor=0.51, p=5.3e−21
3817: GALNT16|Q8N428
1154: GALNT16|Q8N428
cor=0.42, p=6.7e−06
3818: CALY|Q9NYX4
1803: CALY|Q9NYX4
cor=0.58, p=9.5e−27
3820: ENTPD6|O75354
969: ENTPD6|O75354
cor=0.54, p=1.6e−08
3828: ERLEC1|Q96DZ1
1708: ERLEC1|Q96DZ1
cor=0.81, p=1.3e−66
3830: ERAP2|Q6P179
927: ERAP2|Q6P179
cor=0.26, p=0.0018
3837: B4GALNT1|Q00973
928: B4GALNT1|Q00973
cor=0.54, p=1.1e−21
3838: THBS3|P49746
1076: THBS3|P49746
cor=0.54, p=9.6e−23
3841: SCUBE1|Q8IWY4
925: SCUBE1|Q8IWY4
cor=0.29, p=2e−04
3842: ATP1B3|P54709
2131: ATP1B3|P54709
cor=0.72, p=1.5e−48
3846: PLXNA1|Q9UIW2
702: PLXNA1|Q9UIW2
cor=0.035, p=0.7
3853: AHSP|Q9NZD4
1503: AHSP|Q9NZD4
cor=0.63, p=4.9e−34
3856: EPDR1|Q9UM22
847: EPDR1|Q9UM22
cor=0.055, p=0.52
3857: CNTNAP5|Q8WYK1
564: CNTNAP5|Q8WYK1
cor=0.43, p=0.00023
3859: CRIP2|P52943
1967: CRIP2|P52943
cor=0.56, p=7.8e−26
3862: PENK|P01210
314: PENK|P01210
cor=0.62, p=7.9e−33
3863: MAN1A2|O60476
616: MAN1A2|O60476
cor=0.32, p=1.6e−06
3875: PGM1|P36871
557: PGM1|P36871
cor=0.36, p=1.4e−08
3896: EPHB3|P54753
1361: EPHB3|P54753
cor=0.56, p=1.4e−13
3898: MANF|P55145
2072: MANF|P55145
cor=0.62, p=7.9e−33
3903: TWSG1|Q9GZX9
1063: TWSG1|Q9GZX9
cor=0.57, p=6.7e−27
3904: PLXDC1|Q8IUK5
823: PLXDC1|Q8IUK5
cor=0.51, p=8.2e−20
3909: MYDGF|Q969H8
1261: MYDGF|Q969H8
cor=0.36, p=4.7e−10
3912: MAN2B2|Q9Y2E5
819: MAN2B2|Q9Y2E5
cor=0.72, p=1.5e−48
3915: NPTX1|Q15818
355: NPTX1|Q15818
cor=0.64, p=1.7e−35
3918: SIAE|Q9HAT2
688: SIAE|Q9HAT2
cor=0.62, p=2.4e−31
3919: CTSO|P43234
1230: CTSO|P43234
cor=0.49, p=1.9e−05
3922: CPA4|Q9UI42
2083: CPA4|Q9UI42
cor=0.65, p=3e−35
3933: B3GNT8|Q7Z7M8
1616: B3GNT8|Q7Z7M8
cor=0.79, p=2e−64
3936: AZGP1|P25311
148: AZGP1|P25311
cor=0.73, p=1.6e−50
3937: CBLN1|P23435
1266: CBLN1|P23435
cor=0.78, p=8.4e−62
3939: WFDC1|Q9HC57
1357: WFDC1|Q9HC57
cor=0.17, p=0.0033
3941: RCN1|Q15293
1031: RCN1|Q15293

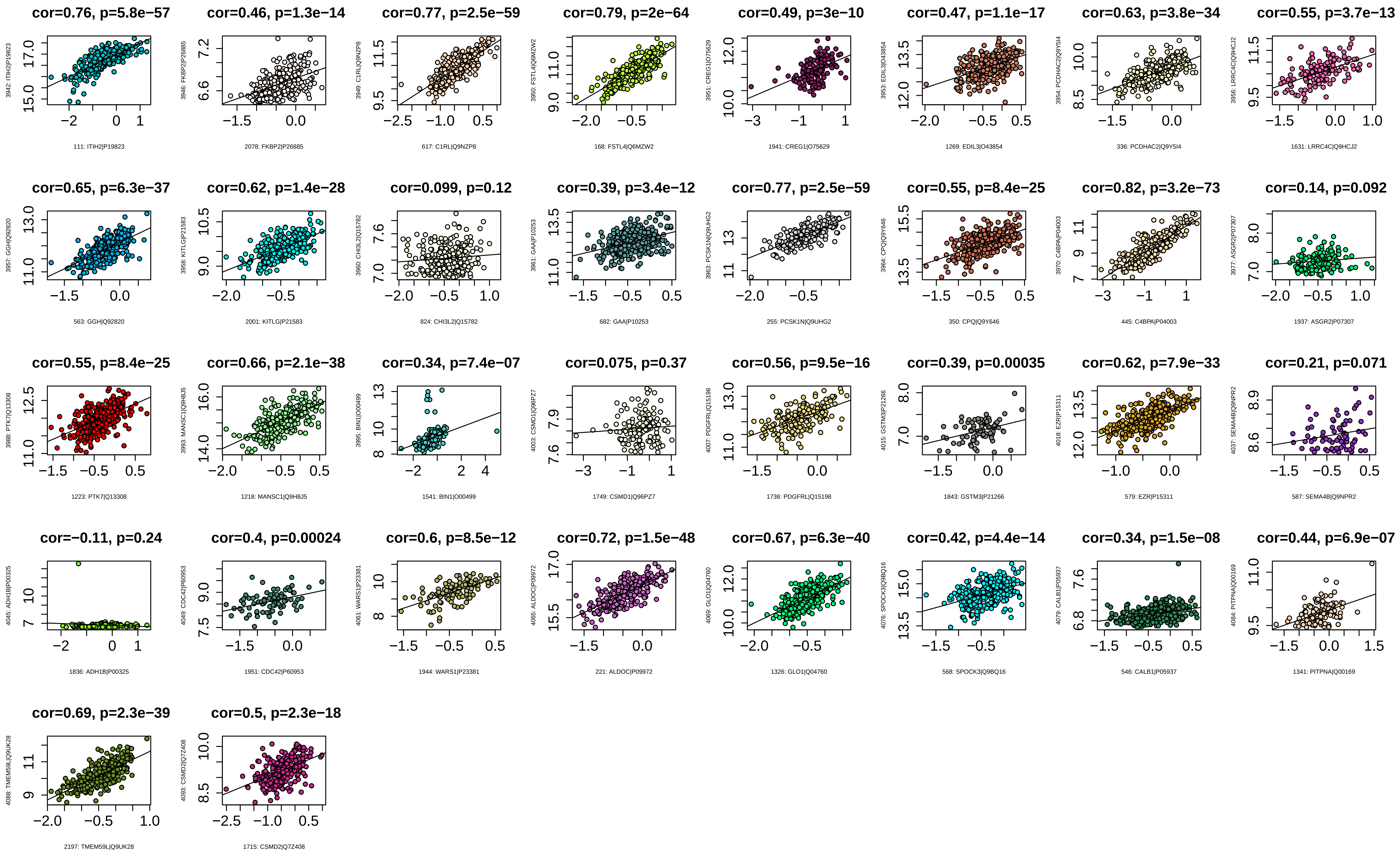
cor=0.76, p=5.8e−57
3942: ITIH2|P19823
111: ITIH2|P19823
cor=0.46, p=1.3e−14
3946: FKBP2|P26885
2078: FKBP2|P26885
cor=0.77, p=2.5e−59
3949: C1RL|Q9NZP8
617: C1RL|Q9NZP8
cor=0.79, p=2e−64
3950: FSTL4|Q6MZW2
168: FSTL4|Q6MZW2
cor=0.49, p=3e−10
3951: CREG1|O75629
1941: CREG1|O75629
cor=0.47, p=1.1e−17
3953: EDIL3|O43854
1269: EDIL3|O43854
cor=0.63, p=3.8e−34
3954: PCDHAC2|Q9Y5I4
336: PCDHAC2|Q9Y5I4
cor=0.55, p=3.7e−13
3956: LRRC4C|Q9HCJ2
1631: LRRC4C|Q9HCJ2
cor=0.65, p=6.3e−37
3957: GGH|Q92820
563: GGH|Q92820
cor=0.62, p=1.4e−28
3958: KITLG|P21583
2001: KITLG|P21583
cor=0.099, p=0.12
3960: CHI3L2|Q15782
824: CHI3L2|Q15782
cor=0.39, p=3.4e−12
3961: GAA|P10253
682: GAA|P10253
cor=0.77, p=2.5e−59
3963: PCSK1N|Q9UHG2
255: PCSK1N|Q9UHG2
cor=0.55, p=8.4e−25
3964: CPQ|Q9Y646
350: CPQ|Q9Y646
cor=0.82, p=3.2e−73
3970: C4BPA|P04003
445: C4BPA|P04003
cor=0.14, p=0.092
3977: ASGR2|P07307
1937: ASGR2|P07307
cor=0.55, p=8.4e−25
3988: PTK7|Q13308
1223: PTK7|Q13308
cor=0.66, p=2.1e−38
3993: MANSC1|Q9H8J5
1218: MANSC1|Q9H8J5
cor=0.34, p=7.4e−07
3995: BIN1|O00499
1541: BIN1|O00499
cor=0.075, p=0.37
4003: CSMD1|Q96PZ7
1749: CSMD1|Q96PZ7
cor=0.56, p=9.5e−16
4007: PDGFRL|Q15198
1736: PDGFRL|Q15198
cor=0.39, p=0.00035
4015: GSTM3|P21266
1843: GSTM3|P21266
cor=0.62, p=7.9e−33
4018: EZR|P15311
579: EZR|P15311
cor=0.21, p=0.071
4037: SEMA4B|Q9NPR2
587: SEMA4B|Q9NPR2
cor=−0.11, p=0.24
4045: ADH1B|P00325
1836: ADH1B|P00325
cor=0.4, p=0.00024
4049: CDC42|P60953
1951: CDC42|P60953
cor=0.6, p=8.5e−12
4061: WARS1|P23381
1944: WARS1|P23381
cor=0.72, p=1.5e−48
4065: ALDOC|P09972
221: ALDOC|P09972
cor=0.67, p=6.3e−40
4069: GLO1|Q04760
1326: GLO1|Q04760
cor=0.42, p=4.4e−14
4076: SPOCK3|Q9BQ16
568: SPOCK3|Q9BQ16
cor=0.34, p=1.5e−08
4079: CALB1|P05937
546: CALB1|P05937
cor=0.44, p=6.9e−07
4084: PITPNA|Q00169
1341: PITPNA|Q00169
cor=0.69, p=2.3e−39
4088: TMEM59L|Q9UK28
2197: TMEM59L|Q9UK28
cor=0.5, p=2.3e−18
4093: CSMD2|Q7Z408
1715: CSMD2|Q7Z408

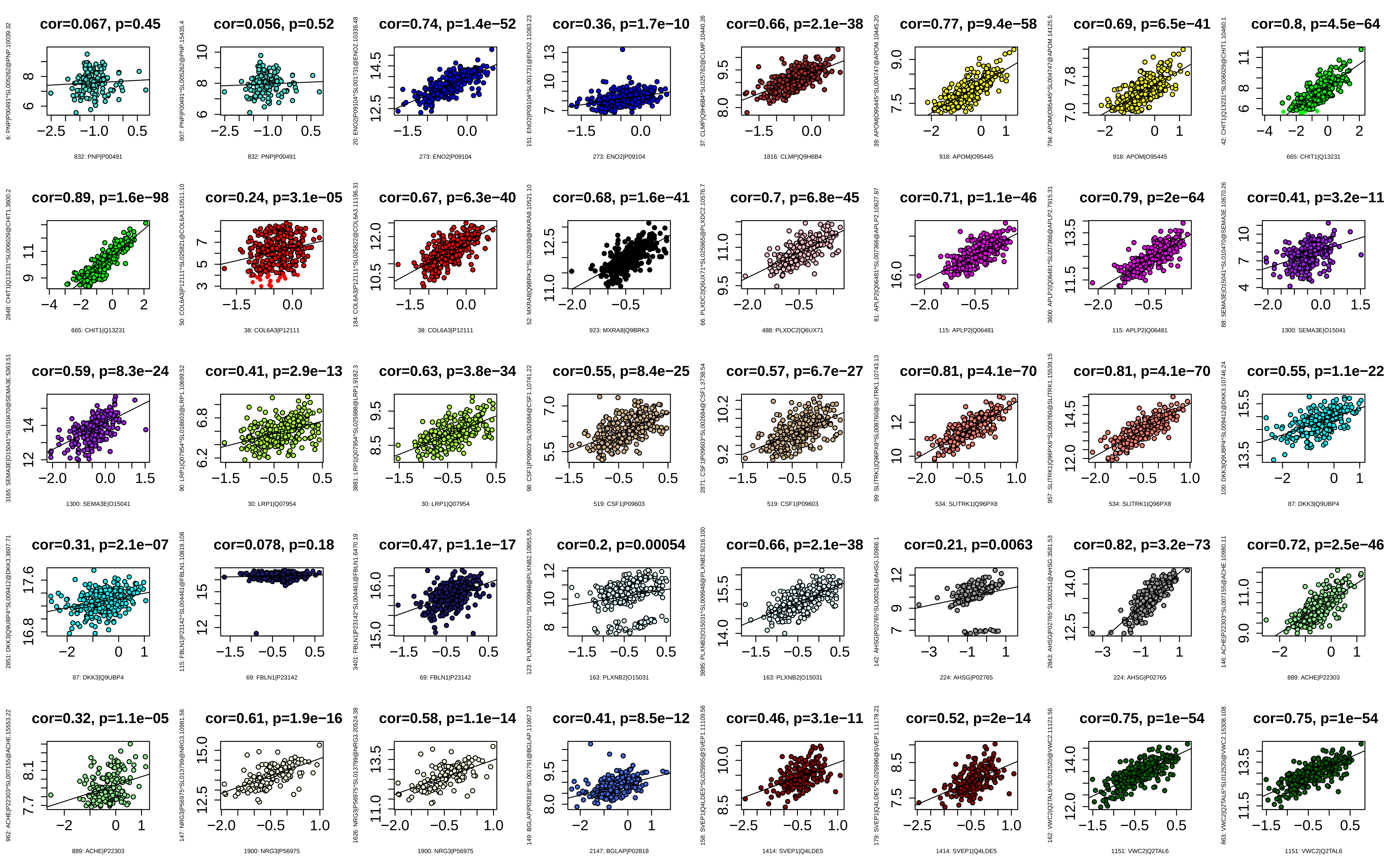
cor=0.067, p=0.45
832: PNP|P00491
cor=0.056, p=0.52
832: PNP|P00491
cor=0.74, p=1.4e−52
273: ENO2|P09104
cor=0.36, p=1.7e−10
273: ENO2|P09104
cor=0.66, p=2.1e−38
1816: CLMP|Q9H6B4
cor=0.77, p=9.4e−58
918: APOM|O95445
cor=0.69, p=6.5e−41
918: APOM|O95445
cor=0.8, p=4.5e−64
665: CHIT1|Q13231
cor=0.89, p=1.6e−98
665: CHIT1|Q13231
cor=0.24, p=3.1e−05
38: COL6A3|P12111
cor=0.67, p=6.3e−40
38: COL6A3|P12111
cor=0.68, p=1.6e−41
923: MXRA8|Q9BRK3
cor=0.7, p=6.8e−45
488: PLXDC2|Q6UX71
cor=0.71, p=1.1e−46
115: APLP2|Q06481
cor=0.79, p=2e−64
115: APLP2|Q06481
cor=0.41, p=3.2e−11
1300: SEMA3E|O15041
cor=0.59, p=8.3e−24
1300: SEMA3E|O15041
cor=0.41, p=2.9e−13
30: LRP1|Q07954
cor=0.63, p=3.8e−34
30: LRP1|Q07954
cor=0.55, p=8.4e−25
519: CSF1|P09603
cor=0.57, p=6.7e−27
519: CSF1|P09603
cor=0.81, p=4.1e−70
534: SLITRK1|Q96PX8
cor=0.81, p=4.1e−70
534: SLITRK1|Q96PX8
cor=0.55, p=1.1e−22
87: DKK3|Q9UBP4
cor=0.31, p=2.1e−07
87: DKK3|Q9UBP4
cor=0.078, p=0.18
69: FBLN1|P23142
cor=0.47, p=1.1e−17
69: FBLN1|P23142
cor=0.2, p=0.00054
163: PLXNB2|O15031
cor=0.66, p=2.1e−38
163: PLXNB2|O15031
cor=0.21, p=0.0063
224: AHSG|P02765
cor=0.82, p=3.2e−73
224: AHSG|P02765
cor=0.72, p=2.5e−46
889: ACHE|P22303
cor=0.32, p=1.1e−05
889: ACHE|P22303
cor=0.61, p=1.9e−16
1900: NRG3|P56975
cor=0.58, p=1.1e−14
1900: NRG3|P56975
cor=0.41, p=8.5e−12
2147: BGLAP|P02818
cor=0.46, p=3.1e−11
1414: SVEP1|Q4LDE5
cor=0.52, p=2e−14
1414: SVEP1|Q4LDE5
cor=0.75, p=1e−54
1151: VWC2|Q2TAL6
cor=0.75, p=1e−54
1151: VWC2|Q2TAL6

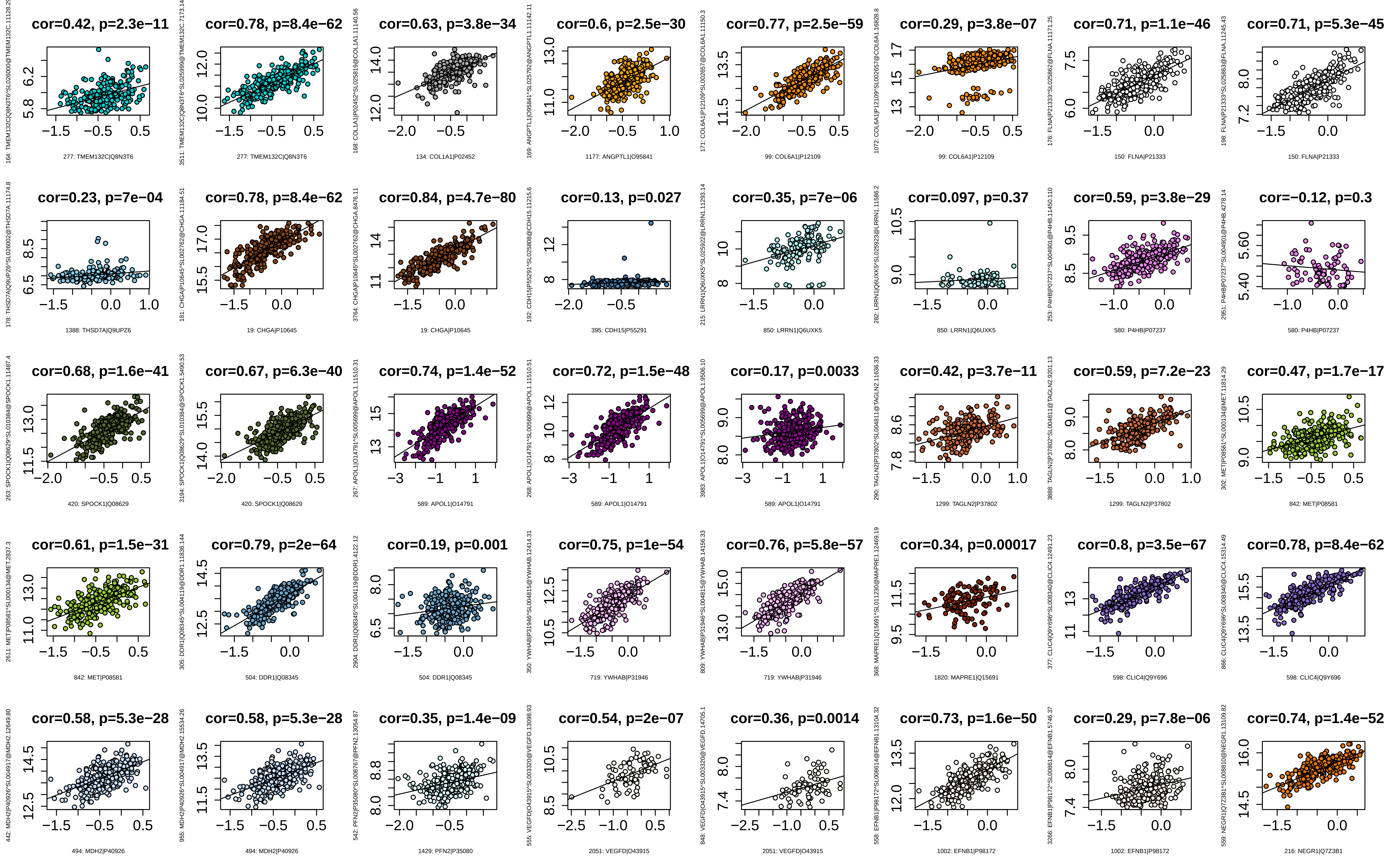
cor=0.42, p=2.3e−11
277: TMEM132C|Q8N3T6
cor=0.78, p=8.4e−62
277: TMEM132C|Q8N3T6
cor=0.63, p=3.8e−34
134: COL1A1|P02452
cor=0.6, p=2.5e−30
1177: ANGPTL1|O95841
cor=0.77, p=2.5e−59
99: COL6A1|P12109
cor=0.29, p=3.8e−07
99: COL6A1|P12109
cor=0.71, p=1.1e−46
150: FLNA|P21333
cor=0.71, p=5.3e−45
150: FLNA|P21333
cor=0.23, p=7e−04
1388: THSD7A|Q9UPZ6
cor=0.78, p=8.4e−62
19: CHGA|P10645
cor=0.84, p=4.7e−80
19: CHGA|P10645
cor=0.13, p=0.027
395: CDH15|P55291
cor=0.35, p=7e−06
850: LRRN1|Q6UXK5
cor=0.097, p=0.37
850: LRRN1|Q6UXK5
cor=0.59, p=3.8e−29
580: P4HB|P07237
cor=−0.12, p=0.3
580: P4HB|P07237
cor=0.68, p=1.6e−41
420: SPOCK1|Q08629
cor=0.67, p=6.3e−40
420: SPOCK1|Q08629
cor=0.74, p=1.4e−52
589: APOL1|O14791
cor=0.72, p=1.5e−48
589: APOL1|O14791
cor=0.17, p=0.0033
589: APOL1|O14791
cor=0.42, p=3.7e−11
1299: TAGLN2|P37802
cor=0.59, p=7.2e−23
1299: TAGLN2|P37802
cor=0.47, p=1.7e−17
842: MET|P08581
cor=0.61, p=1.5e−31
842: MET|P08581
cor=0.79, p=2e−64
504: DDR1|Q08345
cor=0.19, p=0.001
504: DDR1|Q08345
cor=0.75, p=1e−54
719: YWHAB|P31946
cor=0.76, p=5.8e−57
719: YWHAB|P31946
cor=0.34, p=0.00017
1820: MAPRE1|Q15691
cor=0.8, p=3.5e−67
598: CLIC4|Q9Y696
cor=0.78, p=8.4e−62
598: CLIC4|Q9Y696
cor=0.58, p=5.3e−28
494: MDH2|P40926
cor=0.58, p=5.3e−28
494: MDH2|P40926
cor=0.35, p=1.4e−09
1429: PFN2|P35080
cor=0.54, p=2e−07
2051: VEGFD|O43915
cor=0.36, p=0.0014
2051: VEGFD|O43915
cor=0.73, p=1.6e−50
1002: EFNB1|P98172
cor=0.29, p=7.8e−06
1002: EFNB1|P98172
cor=0.74, p=1.4e−52
216: NEGR1|Q7Z3B1

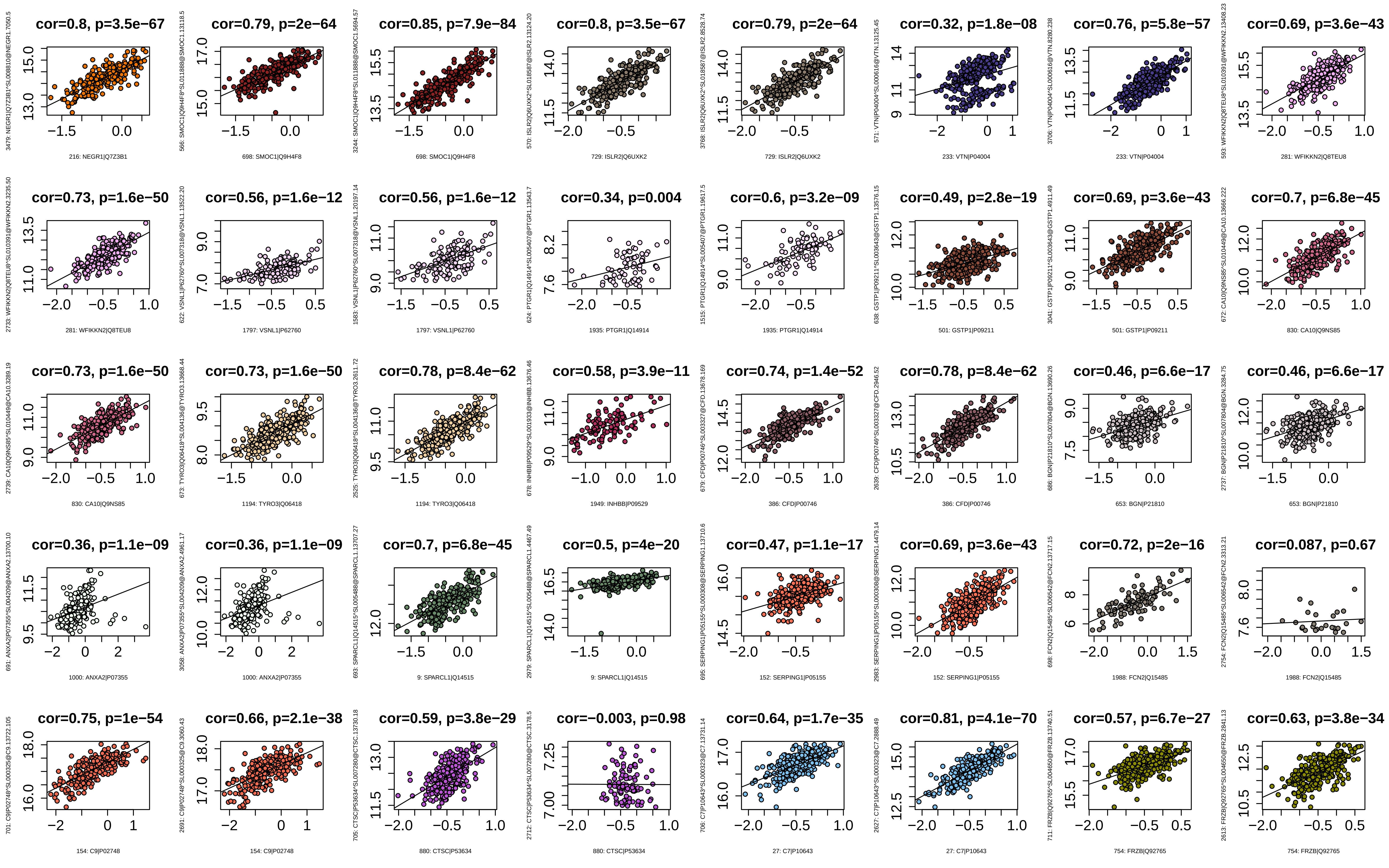
cor=0.8, p=3.5e−67
216: NEGR1|Q7Z3B1
cor=0.79, p=2e−64
698: SMOC1|Q9H4F8
cor=0.85, p=7.9e−84
698: SMOC1|Q9H4F8
cor=0.8, p=3.5e−67
729: ISLR2|Q6UXK2
cor=0.79, p=2e−64
729: ISLR2|Q6UXK2
cor=0.32, p=1.8e−08
233: VTN|P04004
cor=0.76, p=5.8e−57
233: VTN|P04004
cor=0.69, p=3.6e−43
281: WFIKKN2|Q8TEU8
cor=0.73, p=1.6e−50
281: WFIKKN2|Q8TEU8
cor=0.56, p=1.6e−12
1797: VSNL1|P62760
cor=0.56, p=1.6e−12
1797: VSNL1|P62760
cor=0.34, p=0.004
1935: PTGR1|Q14914
cor=0.6, p=3.2e−09
1935: PTGR1|Q14914
cor=0.49, p=2.8e−19
501: GSTP1|P09211
cor=0.69, p=3.6e−43
501: GSTP1|P09211
cor=0.7, p=6.8e−45
830: CA10|Q9NS85
cor=0.73, p=1.6e−50
830: CA10|Q9NS85
cor=0.73, p=1.6e−50
1194: TYRO3|Q06418
cor=0.78, p=8.4e−62
1194: TYRO3|Q06418
cor=0.58, p=3.9e−11
1949: INHBB|P09529
cor=0.74, p=1.4e−52
386: CFD|P00746
cor=0.78, p=8.4e−62
386: CFD|P00746
cor=0.46, p=6.6e−17
653: BGN|P21810
cor=0.46, p=6.6e−17
653: BGN|P21810
cor=0.36, p=1.1e−09
1000: ANXA2|P07355
cor=0.36, p=1.1e−09
1000: ANXA2|P07355
cor=0.7, p=6.8e−45
9: SPARCL1|Q14515
cor=0.5, p=4e−20
9: SPARCL1|Q14515
cor=0.47, p=1.1e−17
152: SERPING1|P05155
cor=0.69, p=3.6e−43
152: SERPING1|P05155
cor=0.72, p=2e−16
1988: FCN2|Q15485
cor=0.087, p=0.67
1988: FCN2|Q15485
cor=0.75, p=1e−54
154: C9|P02748
cor=0.66, p=2.1e−38
154: C9|P02748
cor=0.59, p=3.8e−29
880: CTSC|P53634
cor=−0.003, p=0.98
880: CTSC|P53634
cor=0.64, p=1.7e−35
27: C7|P10643
cor=0.81, p=4.1e−70
27: C7|P10643
cor=0.57, p=6.7e−27
754: FRZB|Q92765
cor=0.63, p=3.8e−34
754: FRZB|Q92765

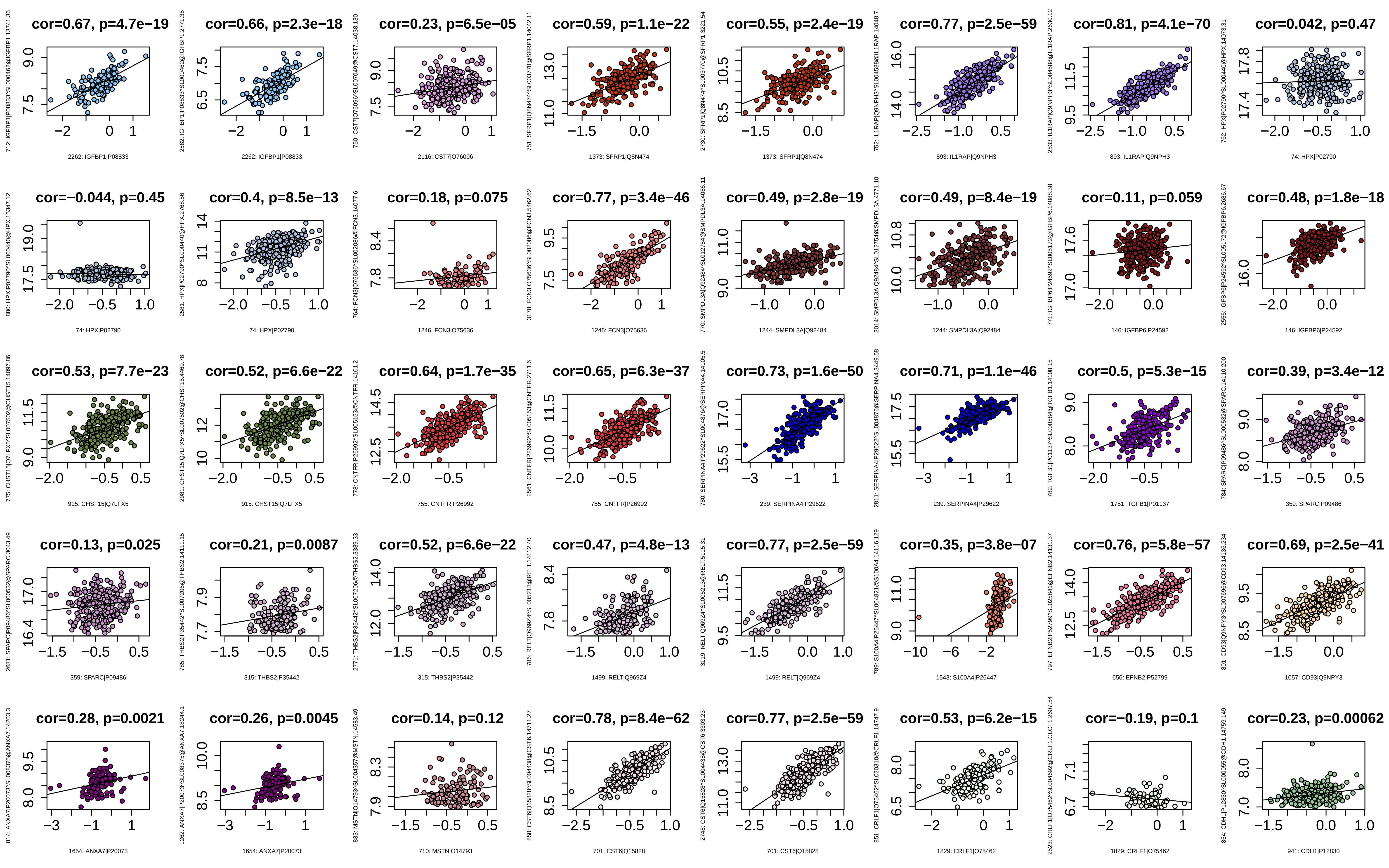
cor=0.67, p=4.7e−19
2262: IGFBP1|P08833
cor=0.66, p=2.3e−18
2262: IGFBP1|P08833
cor=0.23, p=6.5e−05
2116: CST7|O76096
cor=0.59, p=1.1e−22
1373: SFRP1|Q8N474
cor=0.55, p=2.4e−19
1373: SFRP1|Q8N474
cor=0.77, p=2.5e−59
893: IL1RAP|Q9NPH3
cor=0.81, p=4.1e−70
893: IL1RAP|Q9NPH3
cor=0.042, p=0.47
74: HPX|P02790
cor=−0.044, p=0.45
74: HPX|P02790
cor=0.4, p=8.5e−13
74: HPX|P02790
cor=0.18, p=0.075
1246: FCN3|O75636
cor=0.77, p=3.4e−46
1246: FCN3|O75636
cor=0.49, p=2.8e−19
1244: SMPDL3A|Q92484
cor=0.49, p=8.4e−19
1244: SMPDL3A|Q92484
cor=0.11, p=0.059
146: IGFBP6|P24592
cor=0.48, p=1.8e−18
146: IGFBP6|P24592
cor=0.53, p=7.7e−23
915: CHST15|Q7LFX5
cor=0.52, p=6.6e−22
915: CHST15|Q7LFX5
cor=0.64, p=1.7e−35
755: CNTFR|P26992
cor=0.65, p=6.3e−37
755: CNTFR|P26992
cor=0.73, p=1.6e−50
239: SERPINA4|P29622
cor=0.71, p=1.1e−46
239: SERPINA4|P29622
cor=0.5, p=5.3e−15
1751: TGFB1|P01137
cor=0.39, p=3.4e−12
359: SPARC|P09486
cor=0.13, p=0.025
359: SPARC|P09486
cor=0.21, p=0.0087
315: THBS2|P35442
cor=0.52, p=6.6e−22
315: THBS2|P35442
cor=0.47, p=4.8e−13
1499: RELT|Q969Z4
cor=0.77, p=2.5e−59
1499: RELT|Q969Z4
cor=0.35, p=3.8e−07
1543: S100A4|P26447
cor=0.76, p=5.8e−57
656: EFNB2|P52799
cor=0.69, p=2.5e−41
1057: CD93|Q9NPY3
cor=0.28, p=0.0021
1654: ANXA7|P20073
cor=0.26, p=0.0045
1654: ANXA7|P20073
cor=0.14, p=0.12
710: MSTN|O14793
cor=0.78, p=8.4e−62
701: CST6|Q15828
cor=0.77, p=2.5e−59
701: CST6|Q15828
cor=0.53, p=6.2e−15
1829: CRLF1|O75462
cor=−0.19, p=0.1
1829: CRLF1|O75462
cor=0.23, p=0.00062
941: CDH1|P12830

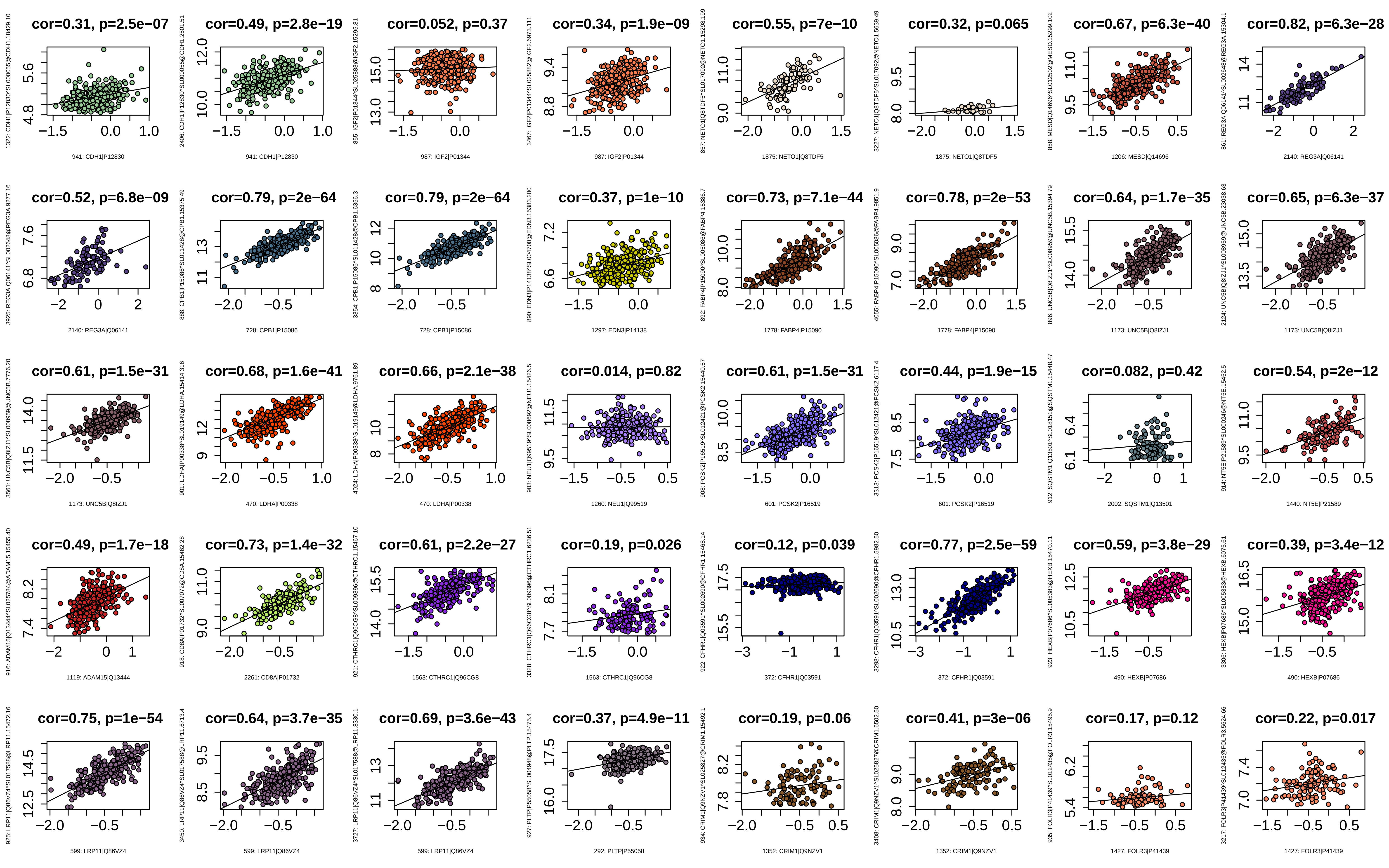

cor=0.31, p=2.5e−07
941: CDH1|P12830
cor=0.49, p=2.8e−19
941: CDH1|P12830
cor=0.052, p=0.37
987: IGF2|P01344
cor=0.34, p=1.9e−09
987: IGF2|P01344
cor=0.55, p=7e−10
1875: NETO1|Q8TDF5
cor=0.32, p=0.065
1875: NETO1|Q8TDF5
cor=0.67, p=6.3e−40
1206: MESD|Q14696
cor=0.82, p=6.3e−28
2140: REG3A|Q06141
cor=0.52, p=6.8e−09
2140: REG3A|Q06141
cor=0.79, p=2e−64
728: CPB1|P15086
cor=0.79, p=2e−64
728: CPB1|P15086
cor=0.37, p=1e−10
1297: EDN3|P14138
cor=0.73, p=7.1e−44
1778: FABP4|P15090
cor=0.78, p=2e−53
1778: FABP4|P15090
cor=0.64, p=1.7e−35
1173: UNC5B|Q8IZJ1
cor=0.65, p=6.3e−37
1173: UNC5B|Q8IZJ1
cor=0.61, p=1.5e−31
1173: UNC5B|Q8IZJ1
cor=0.68, p=1.6e−41
470: LDHA|P00338
cor=0.66, p=2.1e−38
470: LDHA|P00338
cor=0.014, p=0.82
1260: NEU1|Q99519
cor=0.61, p=1.5e−31
601: PCSK2|P16519
cor=0.44, p=1.9e−15
601: PCSK2|P16519
cor=0.082, p=0.42
2002: SQSTM1|Q13501
cor=0.54, p=2e−12
1440: NT5E|P21589
cor=0.49, p=1.7e−18
1119: ADAM15|Q13444
cor=0.73, p=1.4e−32
2261: CD8A|P01732
cor=0.61, p=2.2e−27
1563: CTHRC1|Q96CG8
cor=0.19, p=0.026
1563: CTHRC1|Q96CG8
cor=0.12, p=0.039
372: CFHR1|Q03591
cor=0.77, p=2.5e−59
372: CFHR1|Q03591
cor=0.59, p=3.8e−29
490: HEXB|P07686
cor=0.39, p=3.4e−12
490: HEXB|P07686
cor=0.75, p=1e−54
599: LRP11|Q86VZ4
cor=0.64, p=3.7e−35
599: LRP11|Q86VZ4
cor=0.69, p=3.6e−43
599: LRP11|Q86VZ4
cor=0.37, p=4.9e−11
292: PLTP|P55058
cor=0.19, p=0.06
1352: CRIM1|Q9NZV1
cor=0.41, p=3e−06
1352: CRIM1|Q9NZV1
cor=0.17, p=0.12
1427: FOLR3|P41439
cor=0.22, p=0.017
1427: FOLR3|P41439

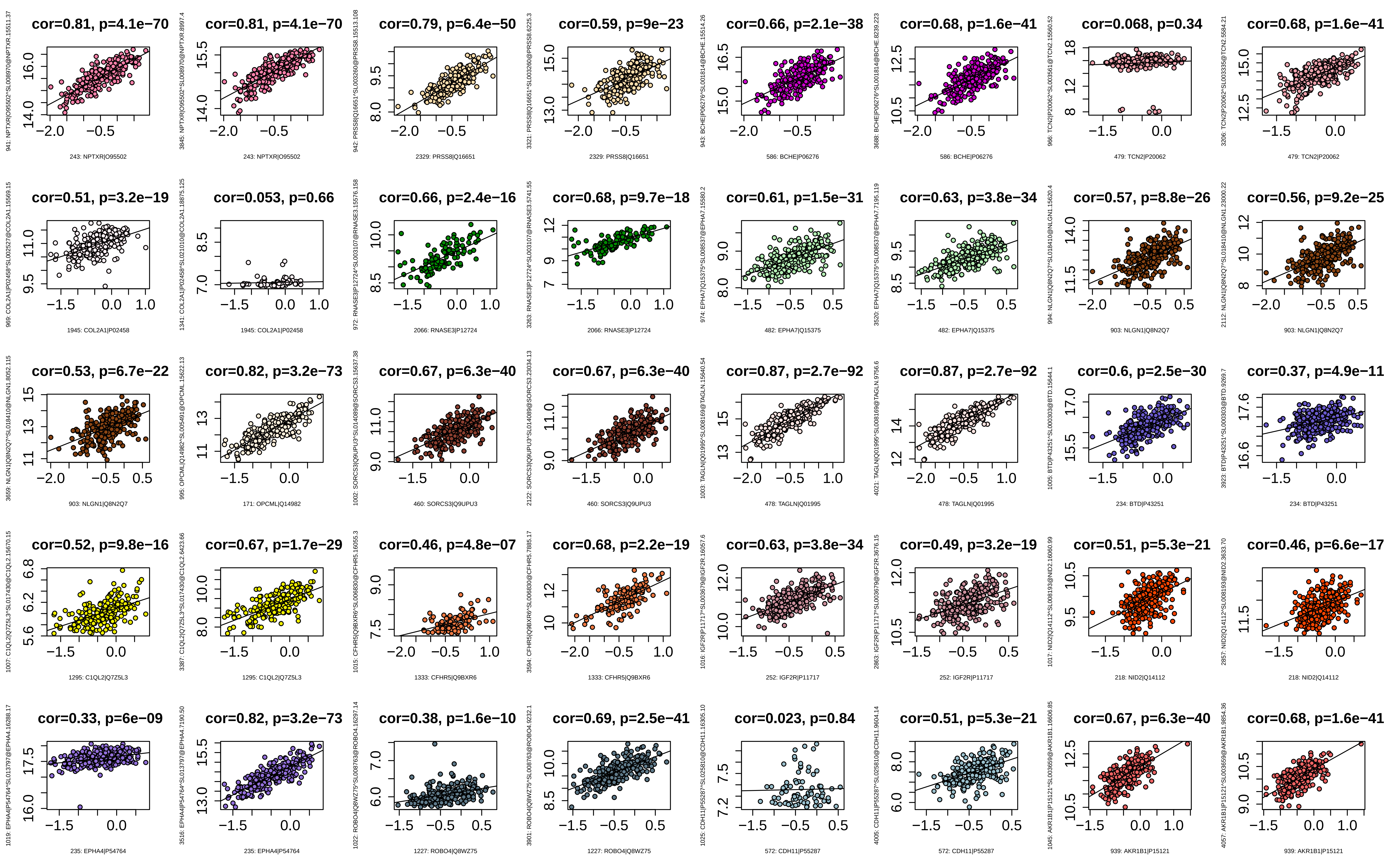

cor=0.81, p=4.1e−70
243: NPTXR|O95502
cor=0.81, p=4.1e−70
243: NPTXR|O95502
cor=0.79, p=6.4e−50
2329: PRSS8|Q16651
cor=0.59, p=9e−23
2329: PRSS8|Q16651
cor=0.66, p=2.1e−38
586: BCHE|P06276
cor=0.68, p=1.6e−41
586: BCHE|P06276
cor=0.068, p=0.34
479: TCN2|P20062
cor=0.68, p=1.6e−41
479: TCN2|P20062
cor=0.51, p=3.2e−19
1945: COL2A1|P02458
cor=0.053, p=0.66
1945: COL2A1|P02458
cor=0.66, p=2.4e−16
2066: RNASE3|P12724
cor=0.68, p=9.7e−18
2066: RNASE3|P12724
cor=0.61, p=1.5e−31
482: EPHA7|Q15375
cor=0.63, p=3.8e−34
482: EPHA7|Q15375
cor=0.57, p=8.8e−26
903: NLGN1|Q8N2Q7
cor=0.56, p=9.2e−25
903: NLGN1|Q8N2Q7
cor=0.53, p=6.7e−22
903: NLGN1|Q8N2Q7
cor=0.82, p=3.2e−73
171: OPCML|Q14982
cor=0.67, p=6.3e−40
460: SORCS3|Q9UPU3
cor=0.67, p=6.3e−40
460: SORCS3|Q9UPU3
cor=0.87, p=2.7e−92
478: TAGLN|Q01995
cor=0.87, p=2.7e−92
478: TAGLN|Q01995
cor=0.6, p=2.5e−30
234: BTD|P43251
cor=0.37, p=4.9e−11
234: BTD|P43251
cor=0.52, p=9.8e−16
1295: C1QL2|Q7Z5L3
cor=0.67, p=1.7e−29
1295: C1QL2|Q7Z5L3
cor=0.46, p=4.8e−07
1333: CFHR5|Q9BXR6
cor=0.68, p=2.2e−19
1333: CFHR5|Q9BXR6
cor=0.63, p=3.8e−34
252: IGF2R|P11717
cor=0.49, p=3.2e−19
252: IGF2R|P11717
cor=0.51, p=5.3e−21
218: NID2|Q14112
cor=0.46, p=6.6e−17
218: NID2|Q14112
cor=0.33, p=6e−09
235: EPHA4|P54764
cor=0.82, p=3.2e−73
235: EPHA4|P54764
cor=0.38, p=1.6e−10
1227: ROBO4|Q8WZ75
cor=0.69, p=2.5e−41
1227: ROBO4|Q8WZ75
cor=0.023, p=0.84
572: CDH11|P55287
cor=0.51, p=5.3e−21
572: CDH11|P55287
cor=0.67, p=6.3e−40
939: AKR1B1|P15121
cor=0.68, p=1.6e−41
939: AKR1B1|P15121

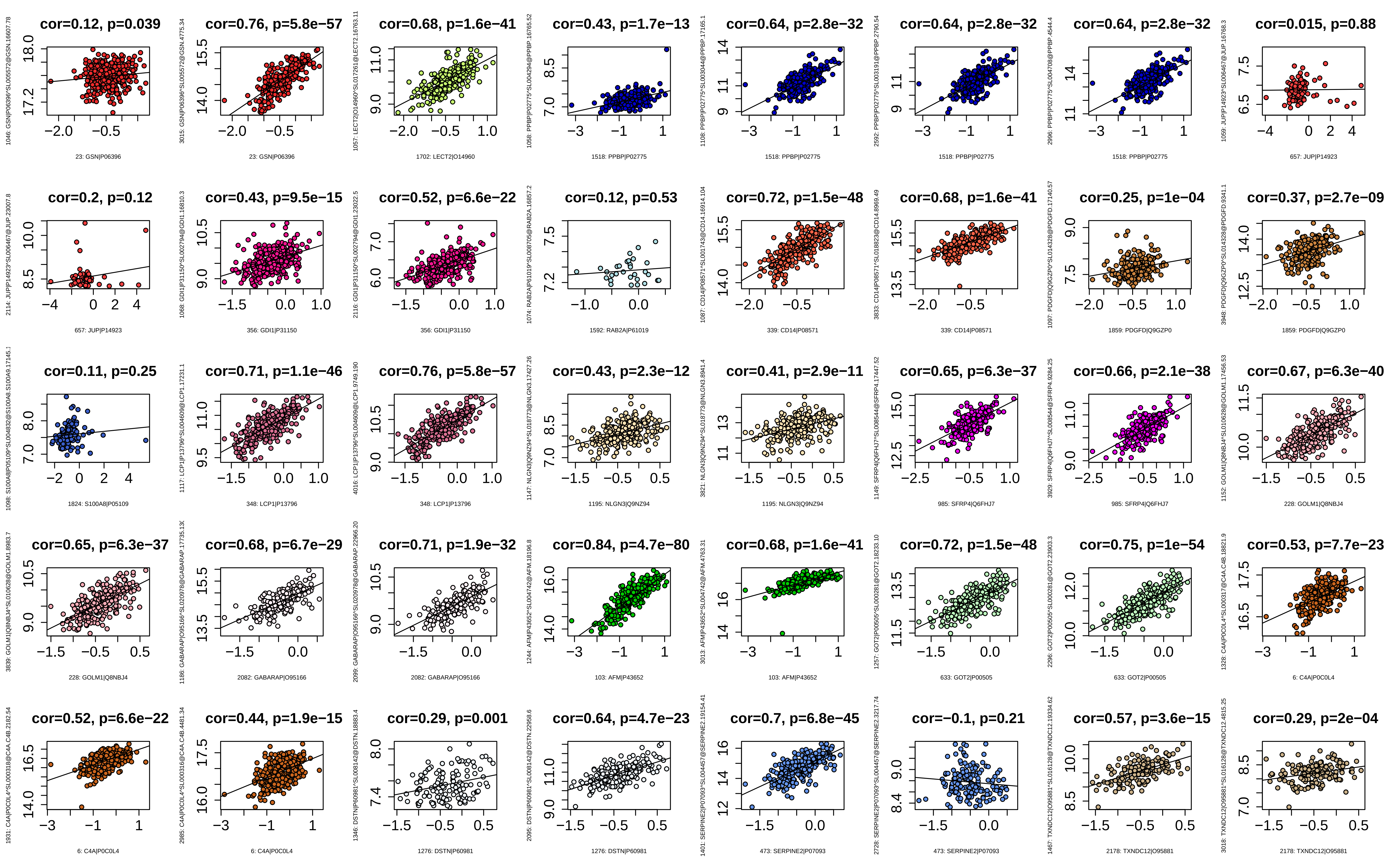
cor=0.12, p=0.039
23: GSN|P06396
cor=0.76, p=5.8e−57
23: GSN|P06396
cor=0.68, p=1.6e−41
1702: LECT2|O14960
cor=0.43, p=1.7e−13
1518: PPBP|P02775
cor=0.64, p=2.8e−32
1518: PPBP|P02775
cor=0.64, p=2.8e−32
1518: PPBP|P02775
cor=0.64, p=2.8e−32
1518: PPBP|P02775
cor=0.015, p=0.88
657: JUP|P14923
cor=0.2, p=0.12
657: JUP|P14923
cor=0.43, p=9.5e−15
356: GDI1|P31150
cor=0.52, p=6.6e−22
356: GDI1|P31150
cor=0.12, p=0.53
1592: RAB2A|P61019
cor=0.72, p=1.5e−48
339: CD14|P08571
cor=0.68, p=1.6e−41
339: CD14|P08571
cor=0.25, p=1e−04
1859: PDGFD|Q9GZP0
cor=0.37, p=2.7e−09
1859: PDGFD|Q9GZP0
cor=0.11, p=0.25
1824: S100A8|P05109
cor=0.71, p=1.1e−46
348: LCP1|P13796
cor=0.76, p=5.8e−57
348: LCP1|P13796
cor=0.43, p=2.3e−12
1195: NLGN3|Q9NZ94
cor=0.41, p=2.9e−11
1195: NLGN3|Q9NZ94
cor=0.65, p=6.3e−37
985: SFRP4|Q6FHJ7
cor=0.66, p=2.1e−38
985: SFRP4|Q6FHJ7
cor=0.67, p=6.3e−40
228: GOLM1|Q8NBJ4
cor=0.65, p=6.3e−37
228: GOLM1|Q8NBJ4
cor=0.68, p=6.7e−29
2082: GABARAP|O95166
cor=0.71, p=1.9e−32
2082: GABARAP|O95166
cor=0.84, p=4.7e−80
103: AFM|P43652
cor=0.68, p=1.6e−41
103: AFM|P43652
cor=0.72, p=1.5e−48
633: GOT2|P00505
cor=0.75, p=1e−54
633: GOT2|P00505
cor=0.53, p=7.7e−23
6: C4A|P0C0L4
cor=0.52, p=6.6e−22
6: C4A|P0C0L4
cor=0.44, p=1.9e−15
6: C4A|P0C0L4
cor=0.29, p=0.001
1276: DSTN|P60981
cor=0.64, p=4.7e−23
1276: DSTN|P60981
cor=0.7, p=6.8e−45
473: SERPINE2|P07093
cor=−0.1, p=0.21
473: SERPINE2|P07093
cor=0.57, p=3.6e−15
2178: TXNDC12|O95881
cor=0.29, p=2e−04
2178: TXNDC12|O95881

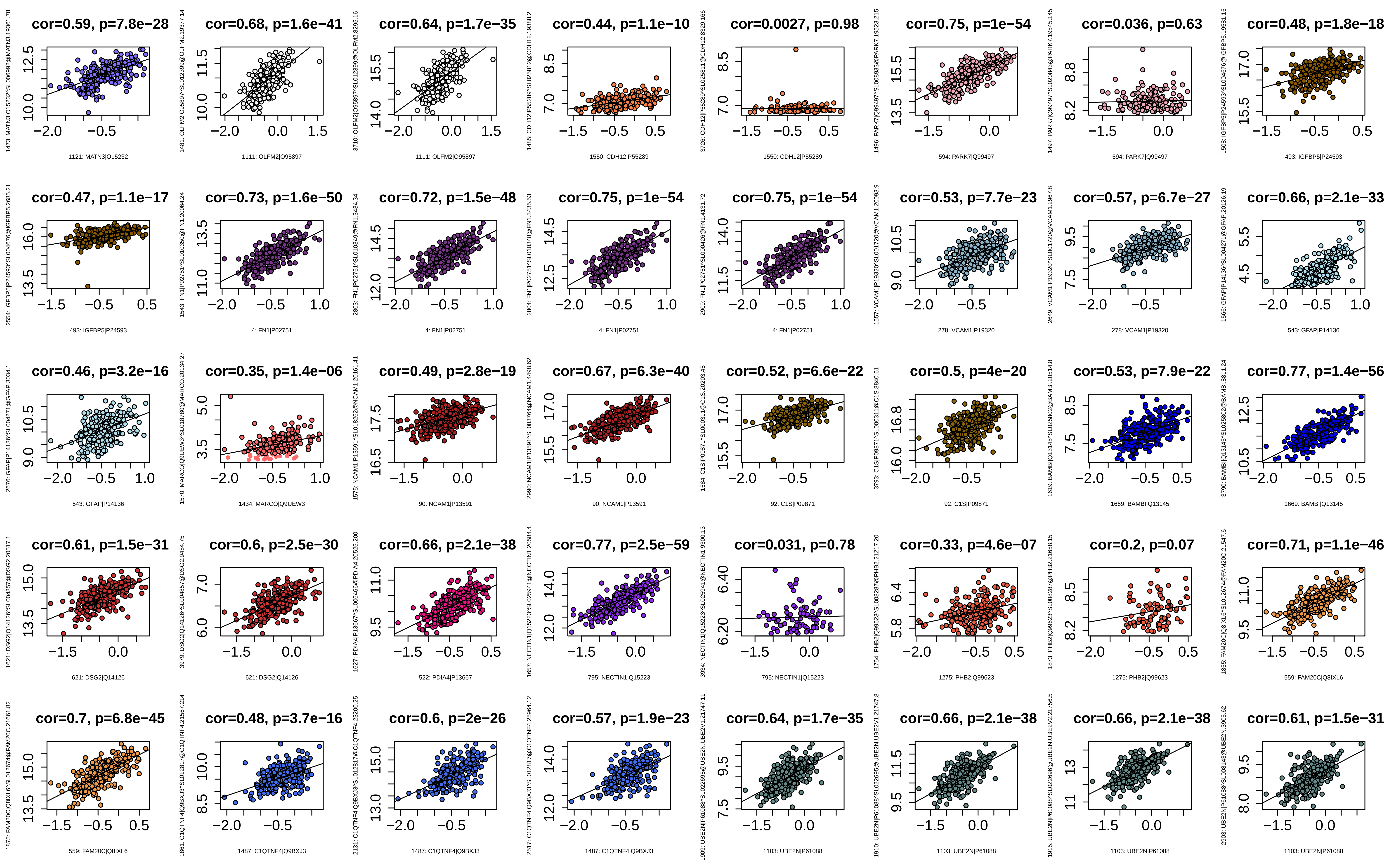

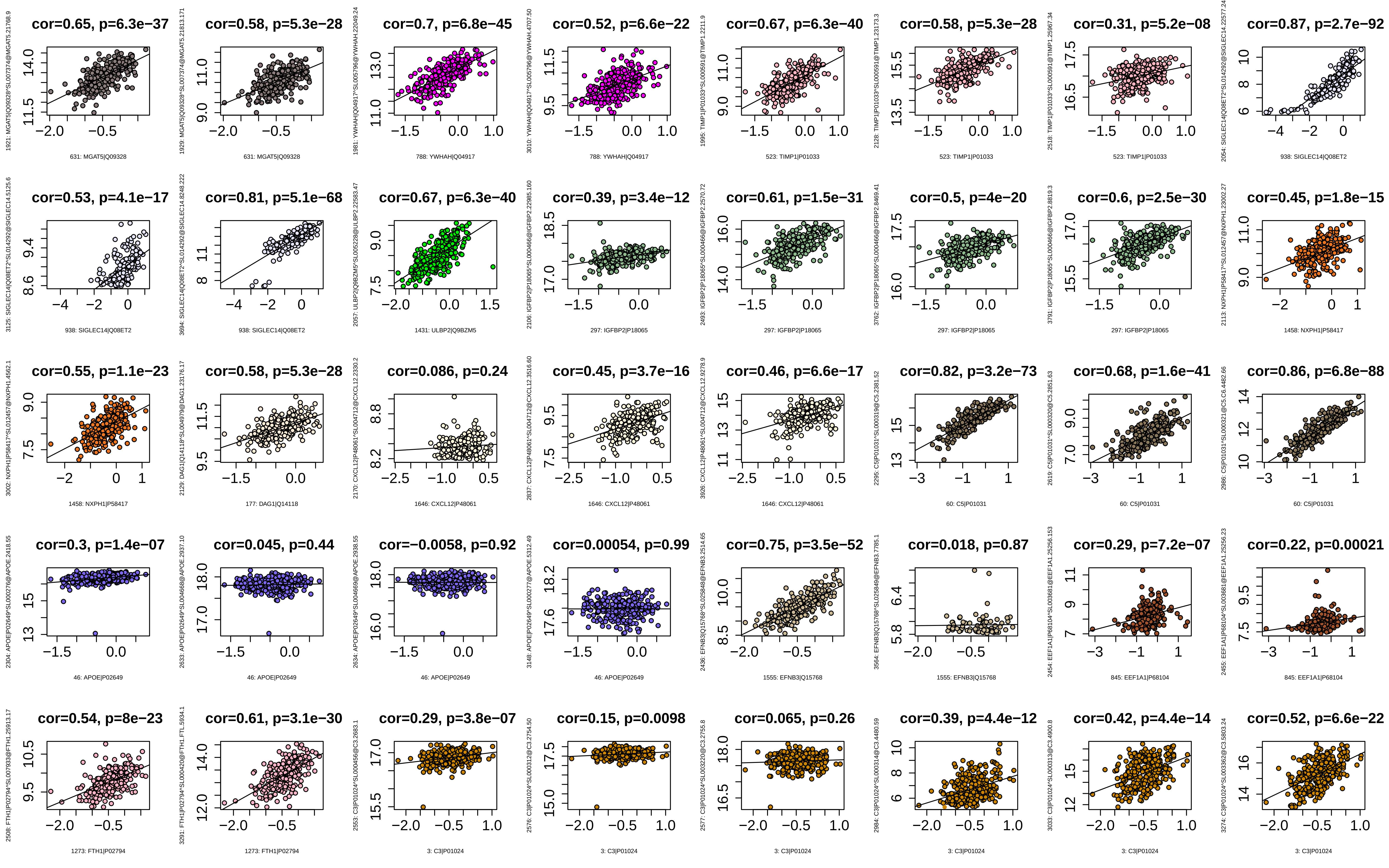
cor=0.65, p=6.3e−37
631: MGAT5|Q09328
cor=0.58, p=5.3e−28
631: MGAT5|Q09328
cor=0.7, p=6.8e−45
788: YWHAH|Q04917
cor=0.52, p=6.6e−22
788: YWHAH|Q04917
cor=0.67, p=6.3e−40
523: TIMP1|P01033
cor=0.58, p=5.3e−28
523: TIMP1|P01033
cor=0.31, p=5.2e−08
523: TIMP1|P01033
cor=0.87, p=2.7e−92
938: SIGLEC14|Q08ET2
cor=0.53, p=4.1e−17
938: SIGLEC14|Q08ET2
cor=0.81, p=5.1e−68
938: SIGLEC14|Q08ET2
cor=0.67, p=6.3e−40
1431: ULBP2|Q9BZM5
cor=0.39, p=3.4e−12
297: IGFBP2|P18065
cor=0.61, p=1.5e−31
297: IGFBP2|P18065
cor=0.5, p=4e−20
297: IGFBP2|P18065
cor=0.6, p=2.5e−30
297: IGFBP2|P18065
cor=0.45, p=1.8e−15
1458: NXPH1|P58417
cor=0.55, p=1.1e−23
1458: NXPH1|P58417
cor=0.58, p=5.3e−28
177: DAG1|Q14118
cor=0.086, p=0.24
1646: CXCL12|P48061
cor=0.45, p=3.7e−16
1646: CXCL12|P48061
cor=0.46, p=6.6e−17
1646: CXCL12|P48061
cor=0.82, p=3.2e−73
60: C5|P01031
cor=0.68, p=1.6e−41
60: C5|P01031
cor=0.86, p=6.8e−88
60: C5|P01031
cor=0.3, p=1.4e−07
46: APOE|P02649
cor=0.045, p=0.44
46: APOE|P02649
cor=−0.0058, p=0.92
46: APOE|P02649
cor=0.00054, p=0.99
46: APOE|P02649
cor=0.75, p=3.5e−52
1555: EFNB3|Q15768
cor=0.018, p=0.87
1555: EFNB3|Q15768
cor=0.29, p=7.2e−07
845: EEF1A1|P68104
cor=0.22, p=0.00021
845: EEF1A1|P68104
cor=0.54, p=8e−23
1273: FTH1|P02794
cor=0.61, p=3.1e−30
1273: FTH1|P02794
cor=0.29, p=3.8e−07
3: C3|P01024
cor=0.15, p=0.0098
3: C3|P01024
cor=0.065, p=0.26
3: C3|P01024
cor=0.39, p=4.4e−12
3: C3|P01024
cor=0.42, p=4.4e−14
3: C3|P01024
cor=0.52, p=6.6e−22
3: C3|P01024

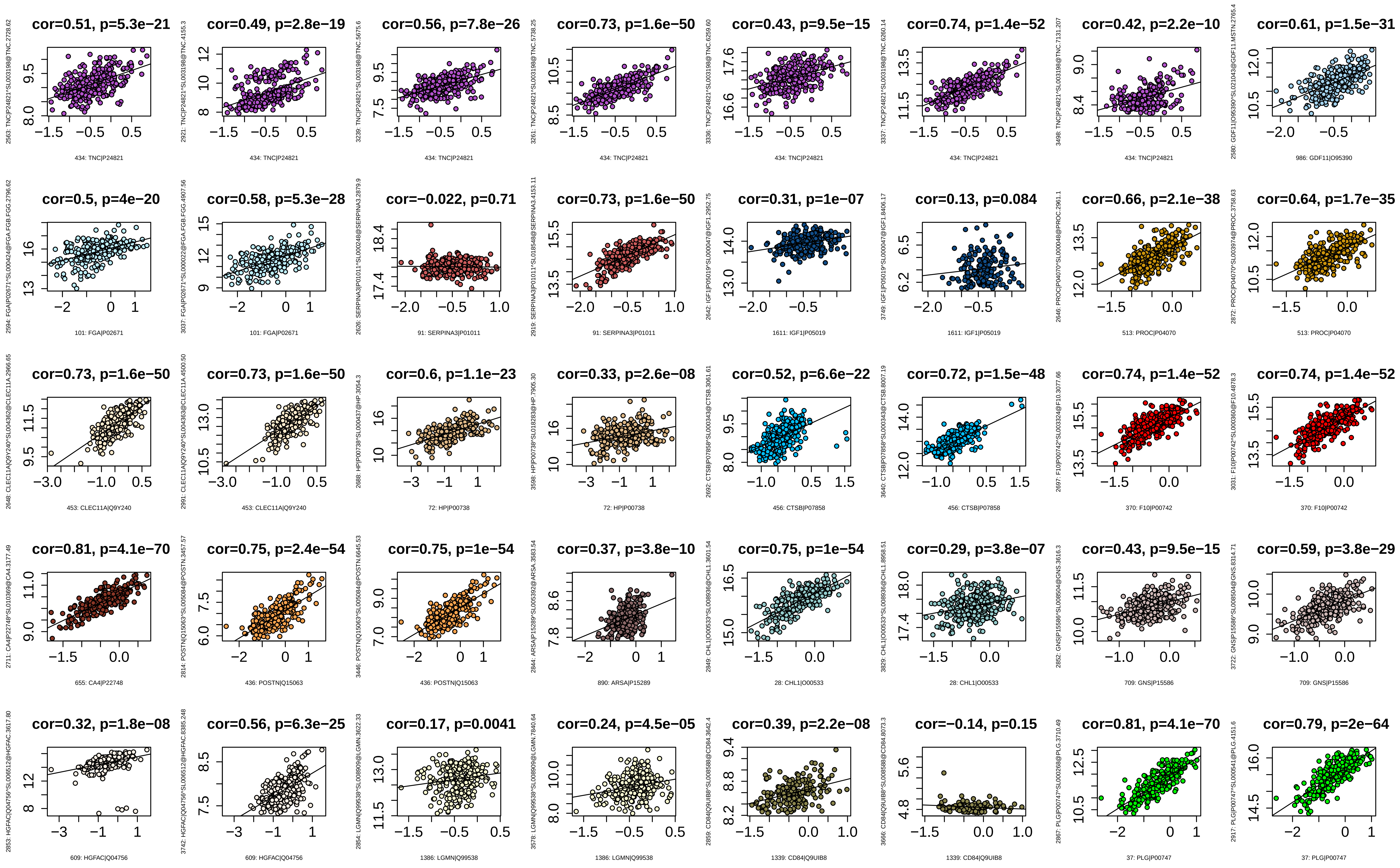
cor=0.51, p=5.3e−21
434: TNC|P24821
cor=0.49, p=2.8e−19
434: TNC|P24821
cor=0.56, p=7.8e−26
434: TNC|P24821
cor=0.73, p=1.6e−50
434: TNC|P24821
cor=0.43, p=9.5e−15
434: TNC|P24821
cor=0.74, p=1.4e−52
434: TNC|P24821
cor=0.42, p=2.2e−10
434: TNC|P24821
cor=0.61, p=1.5e−31
986: GDF11|O95390
cor=0.5, p=4e−20
101: FGA|P02671
cor=0.58, p=5.3e−28
101: FGA|P02671
cor=−0.022, p=0.71
91: SERPINA3|P01011
cor=0.73, p=1.6e−50
91: SERPINA3|P01011
cor=0.31, p=1e−07
1611: IGF1|P05019
cor=0.13, p=0.084
1611: IGF1|P05019
cor=0.66, p=2.1e−38
513: PROC|P04070
cor=0.64, p=1.7e−35
513: PROC|P04070
cor=0.73, p=1.6e−50
453: CLEC11A|Q9Y240
cor=0.73, p=1.6e−50
453: CLEC11A|Q9Y240
cor=0.6, p=1.1e−23
72: HP|P00738
cor=0.33, p=2.6e−08
72: HP|P00738
cor=0.52, p=6.6e−22
456: CTSB|P07858
cor=0.72, p=1.5e−48
456: CTSB|P07858
cor=0.74, p=1.4e−52
370: F10|P00742
cor=0.74, p=1.4e−52
370: F10|P00742
cor=0.81, p=4.1e−70
655: CA4|P22748
cor=0.75, p=2.4e−54
436: POSTN|Q15063
cor=0.75, p=1e−54
436: POSTN|Q15063
cor=0.37, p=3.8e−10
890: ARSA|P15289
cor=0.75, p=1e−54
28: CHL1|O00533
cor=0.29, p=3.8e−07
28: CHL1|O00533
cor=0.43, p=9.5e−15
709: GNS|P15586
cor=0.59, p=3.8e−29
709: GNS|P15586
cor=0.32, p=1.8e−08
609: HGFAC|Q04756
cor=0.56, p=6.3e−25
609: HGFAC|Q04756
cor=0.17, p=0.0041
1386: LGMN|Q99538
cor=0.24, p=4.5e−05
1386: LGMN|Q99538
cor=0.39, p=2.2e−08
1339: CD84|Q9UIB8
cor=−0.14, p=0.15
1339: CD84|Q9UIB8
cor=0.81, p=4.1e−70
37: PLG|P00747
cor=0.79, p=2e−64
37: PLG|P00747

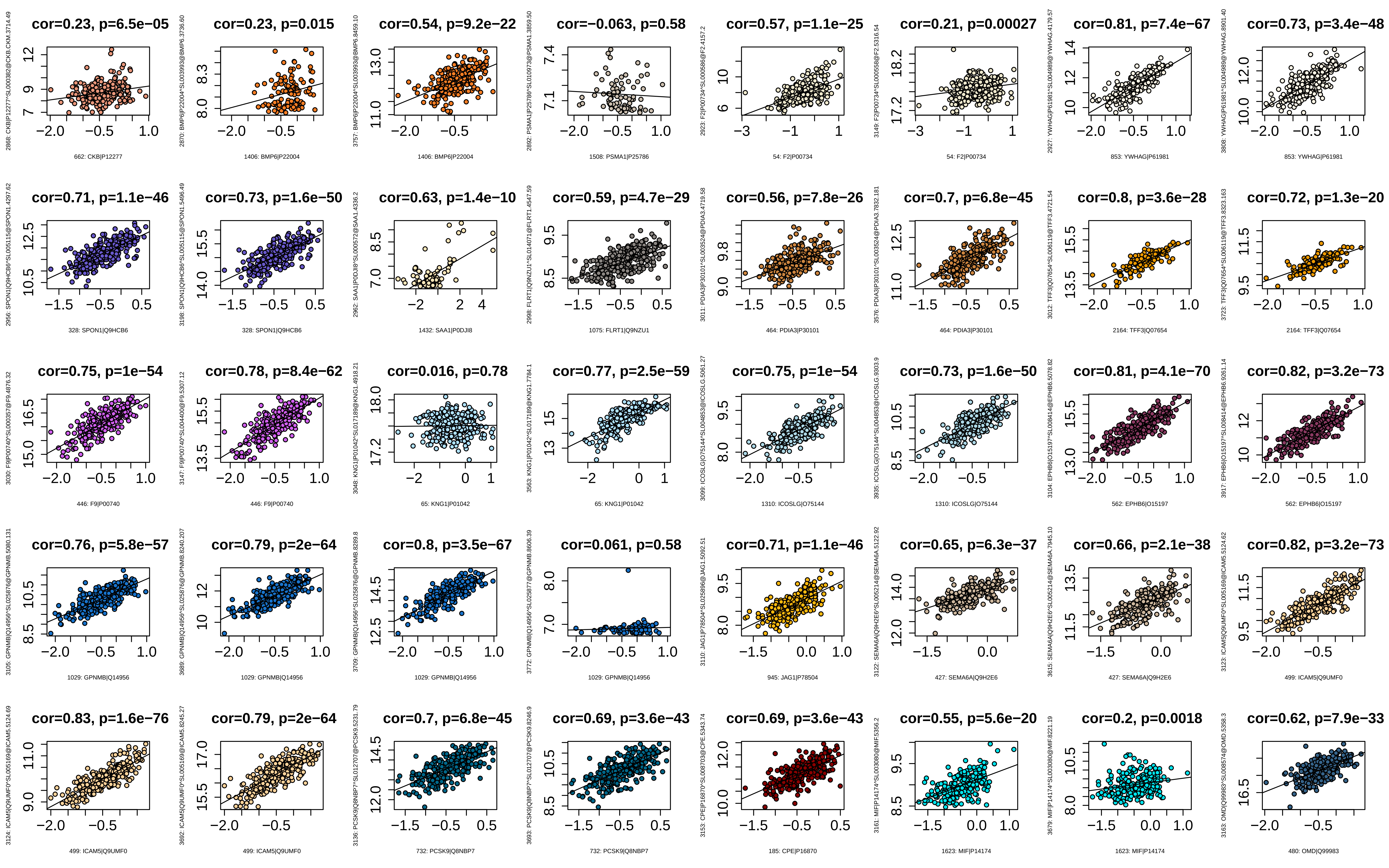
cor=0.23, p=6.5e−05
662: CKB|P12277
cor=0.23, p=0.015
1406: BMP6|P22004
cor=0.54, p=9.2e−22
1406: BMP6|P22004
cor=−0.063, p=0.58
1508: PSMA1|P25786
cor=0.57, p=1.1e−25
54: F2|P00734
cor=0.21, p=0.00027
54: F2|P00734
cor=0.81, p=7.4e−67
853: YWHAG|P61981
cor=0.73, p=3.4e−48
853: YWHAG|P61981
cor=0.71, p=1.1e−46
328: SPON1|Q9HCB6
cor=0.73, p=1.6e−50
328: SPON1|Q9HCB6
cor=0.63, p=1.4e−10
1432: SAA1|P0DJI8
cor=0.59, p=4.7e−29
1075: FLRT1|Q9NZU1
cor=0.56, p=7.8e−26
464: PDIA3|P30101
cor=0.7, p=6.8e−45
464: PDIA3|P30101
cor=0.8, p=3.6e−28
2164: TFF3|Q07654
cor=0.72, p=1.3e−20
2164: TFF3|Q07654
cor=0.75, p=1e−54
446: F9|P00740
cor=0.78, p=8.4e−62
446: F9|P00740
cor=0.016, p=0.78
65: KNG1|P01042
cor=0.77, p=2.5e−59
65: KNG1|P01042
cor=0.75, p=1e−54
1310: ICOSLG|O75144
cor=0.73, p=1.6e−50
1310: ICOSLG|O75144
cor=0.81, p=4.1e−70
562: EPHB6|O15197
cor=0.82, p=3.2e−73
562: EPHB6|O15197
cor=0.76, p=5.8e−57
1029: GPNMB|Q14956
cor=0.79, p=2e−64
1029: GPNMB|Q14956
cor=0.8, p=3.5e−67
1029: GPNMB|Q14956
cor=0.061, p=0.58
1029: GPNMB|Q14956
cor=0.71, p=1.1e−46
945: JAG1|P78504
cor=0.65, p=6.3e−37
427: SEMA6A|Q9H2E6
cor=0.66, p=2.1e−38
427: SEMA6A|Q9H2E6
cor=0.82, p=3.2e−73
499: ICAM5|Q9UMF0
cor=0.83, p=1.6e−76
499: ICAM5|Q9UMF0
cor=0.79, p=2e−64
499: ICAM5|Q9UMF0
cor=0.7, p=6.8e−45
732: PCSK9|Q8NBP7
cor=0.69, p=3.6e−43
732: PCSK9|Q8NBP7
cor=0.69, p=3.6e−43
185: CPE|P16870
cor=0.55, p=5.6e−20
1623: MIF|P14174
cor=0.2, p=0.0018
1623: MIF|P14174
cor=0.62, p=7.9e−33
480: OMD|Q99983

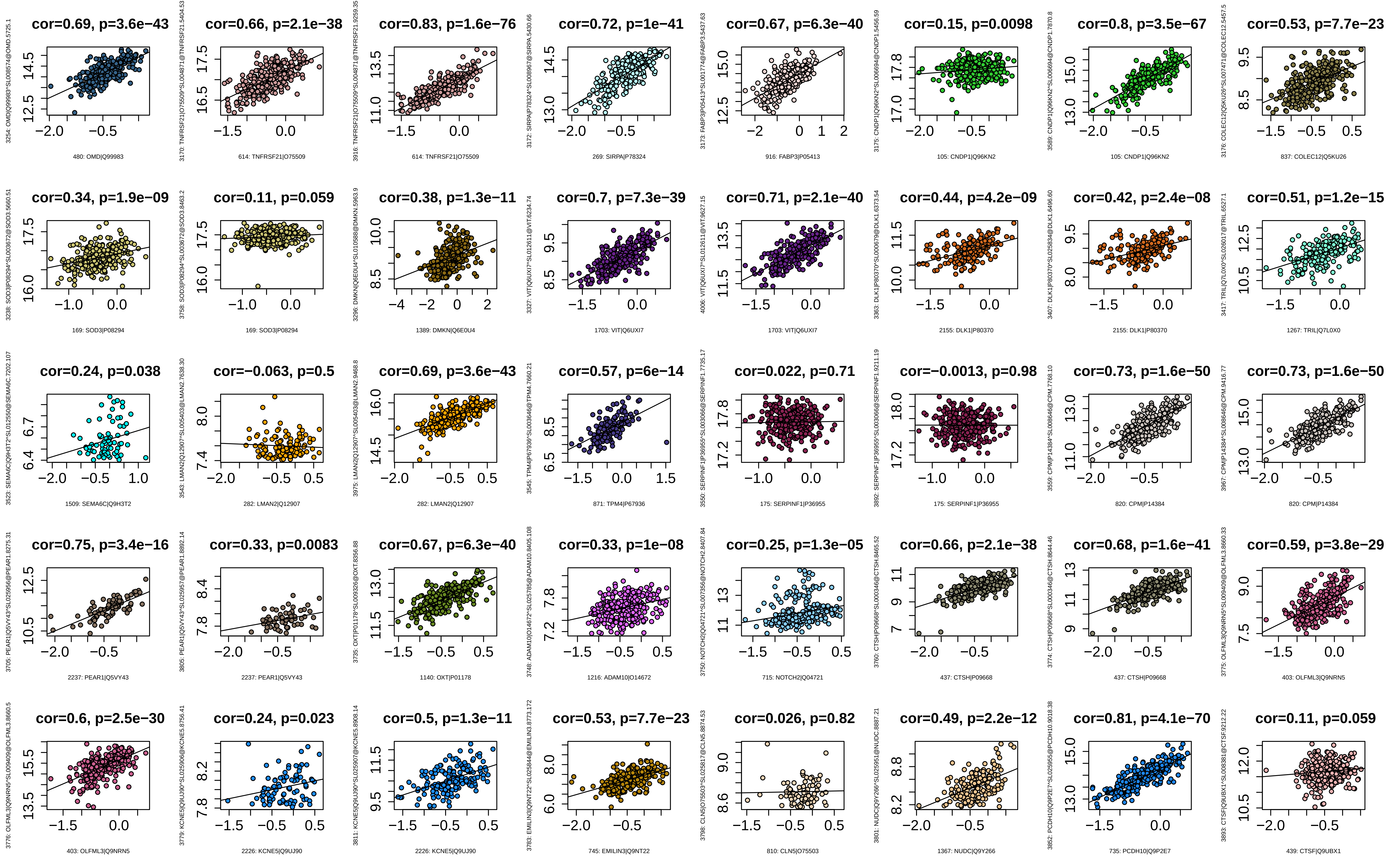
cor=0.69, p=3.6e−43
3254: OMD|Q99983^SL008574@OMD.5725.1
480: OMD|Q99983
cor=0.66, p=2.1e−38
3170: TNFRSF21|O75509^SL004871@TNFRSF21.5404.53
614: TNFRSF21|O75509
cor=0.83, p=1.6e−76
3916: TNFRSF21|O75509^SL004871@TNFRSF21.9259.35
614: TNFRSF21|O75509
cor=0.72, p=1e−41
3172: SIRPA|P78324^SL008967@SIRPA.5430.66
269: SIRPA|P78324
cor=0.67, p=6.3e−40
3173: FABP3|P05413^SL001774@FABP3.5437.63
916: FABP3|P05413
cor=0.15, p=0.0098
3175: CNDP1|Q96KN2^SL006694@CNDP1.5456.59
105: CNDP1|Q96KN2
cor=0.8, p=3.5e−67
3589: CNDP1|Q96KN2^SL006694@CNDP1.7870.8
105: CNDP1|Q96KN2
cor=0.53, p=7.7e−23
3176: COLEC12|Q5KU26^SL007471@COLEC12.5457.5
837: COLEC12|Q5KU26
cor=0.34, p=1.9e−09
3238: SOD3|P08294^SL003672@SOD3.5660.51
169: SOD3|P08294
cor=0.11, p=0.059
3758: SOD3|P08294^SL003672@SOD3.8463.2
169: SOD3|P08294
cor=0.38, p=1.3e−11
3296: DMKN|Q6E0U4^SL010588@DMKN.5963.9
1389: DMKN|Q6E0U4
cor=0.7, p=7.3e−39
3327: VIT|Q6UXI7^SL012611@VIT.6234.74
1703: VIT|Q6UXI7
cor=0.71, p=2.1e−40
4006: VIT|Q6UXI7^SL012611@VIT.9627.15
1703: VIT|Q6UXI7
cor=0.44, p=4.2e−09
3363: DLK1|P80370^SL000679@DLK1.6373.54
2155: DLK1|P80370
cor=0.42, p=2.4e−08
3407: DLK1|P80370^SL025834@DLK1.6496.60
2155: DLK1|P80370
cor=0.51, p=1.2e−15
3417: TRIL|Q7L0X0^SL026017@TRIL.6527.1
1267: TRIL|Q7L0X0
cor=0.24, p=0.038
3523: SEMA6C|Q9H3T2^SL012550@SEMA6C.7202.107
1509: SEMA6C|Q9H3T2
cor=−0.063, p=0.5
3543: LMAN2|Q12907^SL005403@LMAN2.7638.30
282: LMAN2|Q12907
cor=0.69, p=3.6e−43
3975: LMAN2|Q12907^SL005403@LMAN2.9468.8
282: LMAN2|Q12907
cor=0.57, p=6e−14
3545: TPM4|P67936^SL003646@TPM4.7660.21
871: TPM4|P67936
cor=0.022, p=0.71
3550: SERPINF1|P36955^SL003066@SERPINF1.7735.17
175: SERPINF1|P36955
cor=−0.0013, p=0.98
3892: SERPINF1|P36955^SL003066@SERPINF1.9211.19
175: SERPINF1|P36955
cor=0.73, p=1.6e−50
3559: CPM|P14384^SL008646@CPM.7768.10
820: CPM|P14384
cor=0.73, p=1.6e−50
3967: CPM|P14384^SL008646@CPM.9416.77
820: CPM|P14384
cor=0.75, p=3.4e−16
3705: PEAR1|Q5VY43^SL025956@PEAR1.8275.31
2237: PEAR1|Q5VY43
cor=0.33, p=0.0083
3805: PEAR1|Q5VY43^SL025957@PEAR1.8892.14
2237: PEAR1|Q5VY43
cor=0.67, p=6.3e−40
3735: OXT|P01178^SL009326@OXT.8356.88
1140: OXT|P01178
cor=0.33, p=1e−08
3748: ADAM10|O14672^SL025785@ADAM10.8405.108
1216: ADAM10|O14672
cor=0.25, p=1.3e−05
3750: NOTCH2|Q04721^SL007356@NOTCH2.8407.84
715: NOTCH2|Q04721
cor=0.66, p=2.1e−38
3760: CTSH|P09668^SL000346@CTSH.8465.52
437: CTSH|P09668
cor=0.68, p=1.6e−41
3774: CTSH|P09668^SL000346@CTSH.8644.46
437: CTSH|P09668
cor=0.59, p=3.8e−29
3775: OLFML3|Q9NRN5^SL009409@OLFML3.8660.33
403: OLFML3|Q9NRN5
cor=0.6, p=2.5e−30
3776: OLFML3|Q9NRN5^SL009409@OLFML3.8660.5
403: OLFML3|Q9NRN5
cor=0.24, p=0.023
3779: KCNE5|Q9UJ90^SL025906@KCNE5.8756.41
2226: KCNE5|Q9UJ90
cor=0.5, p=1.3e−11
3811: KCNE5|Q9UJ90^SL025907@KCNE5.8908.14
2226: KCNE5|Q9UJ90
cor=0.53, p=7.7e−23
3783: EMILIN3|Q9NT22^SL025844@EMILIN3.8773.172
745: EMILIN3|Q9NT22
cor=0.026, p=0.82
3798: CLN5|O75503^SL025817@CLN5.8874.53
810: CLN5|O75503
cor=0.49, p=2.2e−12
3801: NUDC|Q9Y266^SL025951@NUDC.8887.21
1367: NUDC|Q9Y266
cor=0.81, p=4.1e−70
3852: PCDH10|Q9P2E7^SL025955@PCDH10.9018.38
735: PCDH10|Q9P2E7
cor=0.11, p=0.059
3893: CTSF|Q9UBX1^SL008381@CTSF.9212.22
439: CTSF|Q9UBX1

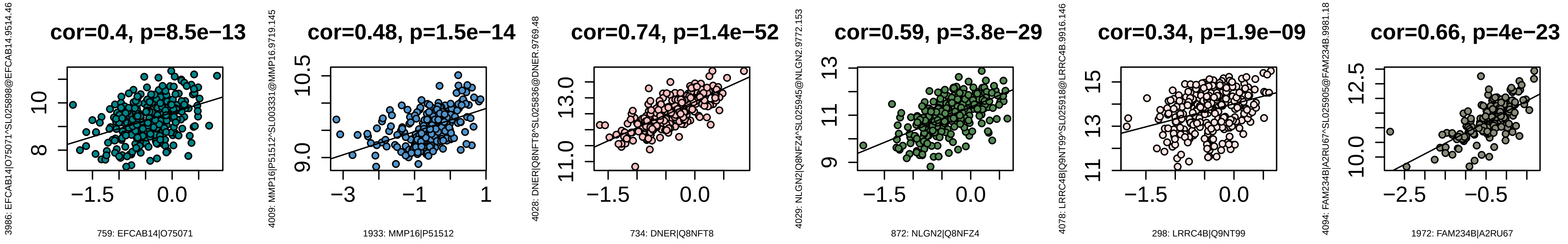
cor=0.4, p=8.5e−13
3986: EFCAB14|O75071^SL025898@EFCAB14.9514.46
759: EFCAB14|O75071
cor=0.48, p=1.5e−14
4009: MMP16|P51512^SL003331@MMP16.9719.145
1933: MMP16|P51512
cor=0.74, p=1.4e−52
4028: DNER|Q8NFT8^SL025836@DNER.9769.48
734: DNER|Q8NFT8
cor=0.59, p=3.8e−29
4029: NLGN2|Q8NFZ4^SL025945@NLGN2.9772.153
872: NLGN2|Q8NFZ4
cor=0.34, p=1.9e−09
4078: LRRC4B|Q9NT99^SL025918@LRRC4B.9916.146
298: LRRC4B|Q9NT99
cor=0.66, p=4e−23
4094: FAM234B|A2RU67^SL025905@FAM234B.9981.18
1972: FAM234B|A2RU67

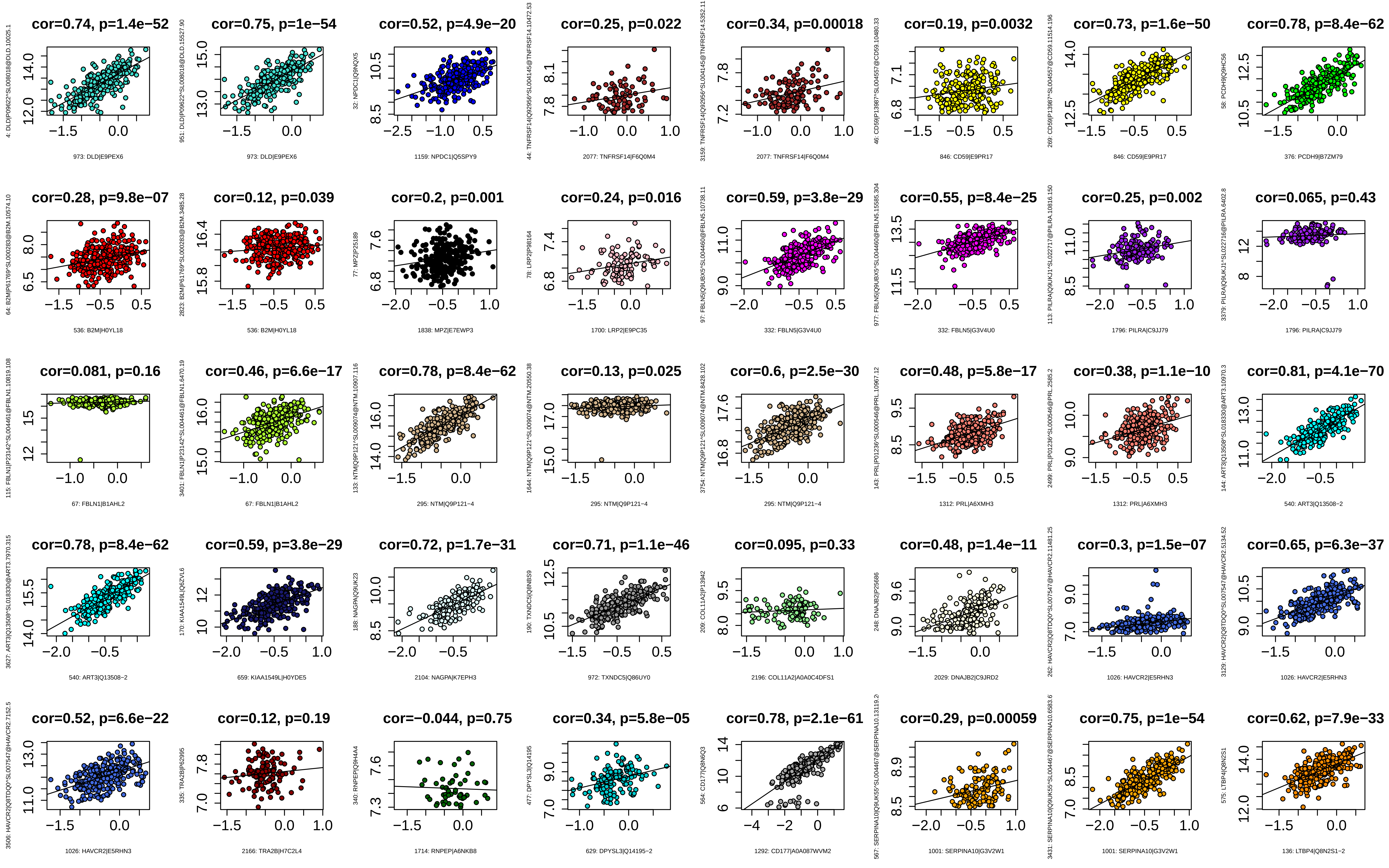
cor=0.74, p=1.4e−52
973: DLD|E9PEX6
cor=0.75, p=1e−54
973: DLD|E9PEX6
cor=0.52, p=4.9e−20
1159: NPDC1|Q5SPY9
cor=0.25, p=0.022
2077: TNFRSF14|F6Q0M4
cor=0.34, p=0.00018
2077: TNFRSF14|F6Q0M4
cor=0.19, p=0.0032
846: CD59|E9PR17
cor=0.73, p=1.6e−50
846: CD59|E9PR17
cor=0.78, p=8.4e−62
376: PCDH9|B7ZM79
cor=0.28, p=9.8e−07
536: B2M|H0YL18
cor=0.12, p=0.039
536: B2M|H0YL18
cor=0.2, p=0.001
1838: MPZ|E7EWP3
cor=0.24, p=0.016
1700: LRP2|E9PC35
cor=0.59, p=3.8e−29
332: FBLN5|G3V4U0
cor=0.55, p=8.4e−25
332: FBLN5|G3V4U0
cor=0.25, p=0.002
1796: PILRA|C9JJ79
cor=0.065, p=0.43
1796: PILRA|C9JJ79
cor=0.081, p=0.16
67: FBLN1|B1AHL2
cor=0.46, p=6.6e−17
67: FBLN1|B1AHL2
cor=0.78, p=8.4e−62
295: NTM|Q9P121−4
cor=0.13, p=0.025
295: NTM|Q9P121−4
cor=0.6, p=2.5e−30
295: NTM|Q9P121−4
cor=0.48, p=5.8e−17
1312: PRL|A6XMH3
cor=0.38, p=1.1e−10
1312: PRL|A6XMH3
cor=0.81, p=4.1e−70
540: ART3|Q13508−2
cor=0.78, p=8.4e−62
540: ART3|Q13508−2
cor=0.59, p=3.8e−29
659: KIAA1549L|H0YDE5
cor=0.72, p=1.7e−31
2104: NAGPA|K7EPH3
cor=0.71, p=1.1e−46
972: TXNDC5|Q86UY0
cor=0.095, p=0.33
2196: COL11A2|A0A0C4DFS1
cor=0.48, p=1.4e−11
2029: DNAJB2|C9JRD2
cor=0.3, p=1.5e−07
1026: HAVCR2|E5RHN3
cor=0.65, p=6.3e−37
1026: HAVCR2|E5RHN3
cor=0.52, p=6.6e−22
1026: HAVCR2|E5RHN3
cor=0.12, p=0.19
2166: TRA2B|H7C2L4
cor=−0.044, p=0.75
1714: RNPEP|A6NKB8
cor=0.34, p=5.8e−05
629: DPYSL3|Q14195−2
cor=0.78, p=2.1e−61
1292: CD177|A0A087WVM2
cor=0.29, p=0.00059
1001: SERPINA10|G3V2W1
cor=0.75, p=1e−54
1001: SERPINA10|G3V2W1
cor=0.62, p=7.9e−33
136: LTBP4|Q8N2S1−2

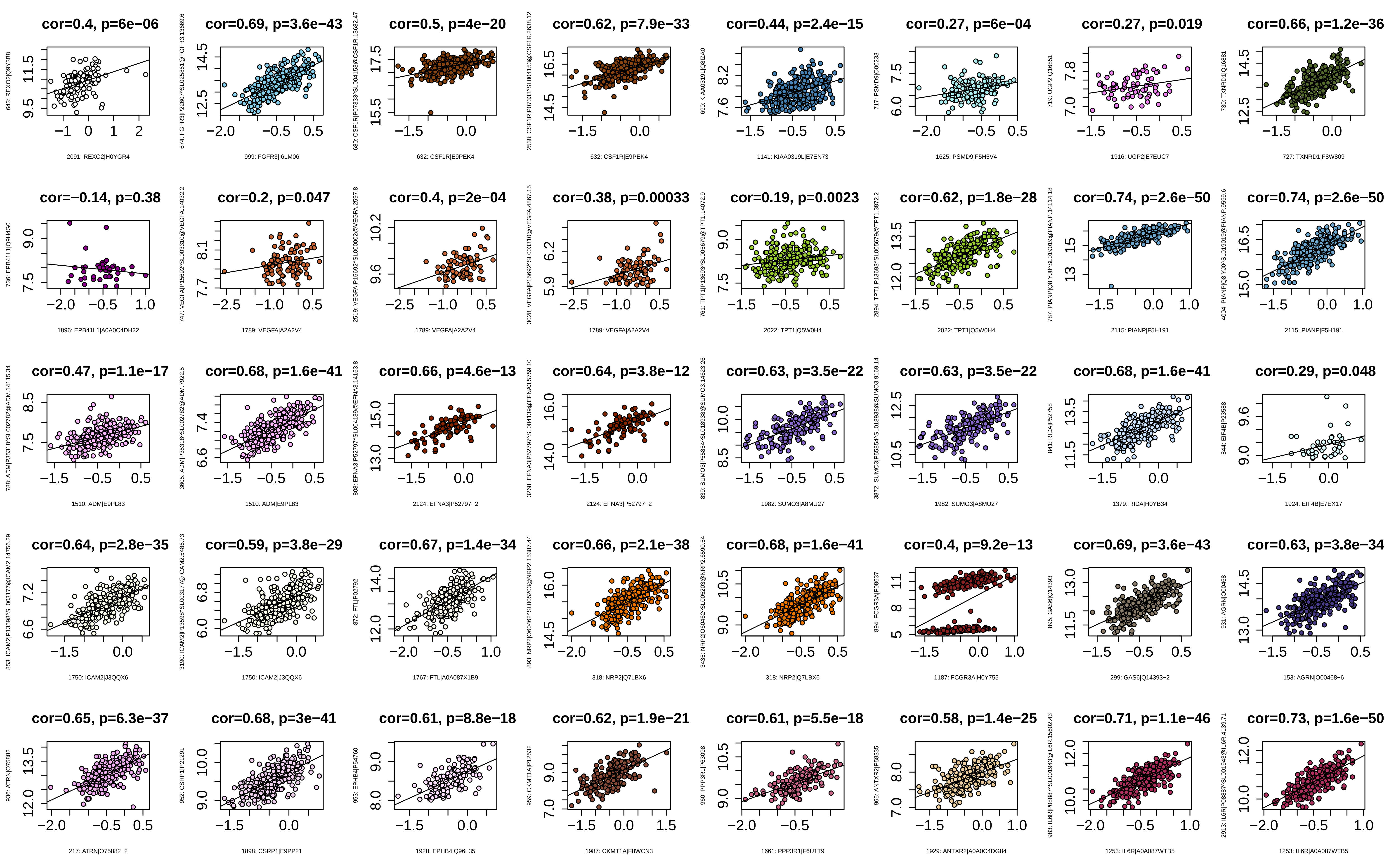
cor=0.4, p=6e−06
2091: REXO2|H0YGR4
cor=0.69, p=3.6e−43
999: FGFR3|I6LM06
cor=0.5, p=4e−20
632: CSF1R|E9PEK4
cor=0.62, p=7.9e−33
632: CSF1R|E9PEK4
cor=0.44, p=2.4e−15
1141: KIAA0319L|E7EN73
cor=0.27, p=6e−04
1625: PSMD9|F5H5V4
cor=0.27, p=0.019
1916: UGP2|E7EUC7
cor=0.66, p=1.2e−36
727: TXNRD1|F8W809
cor=−0.14, p=0.38
1896: EPB41L1|A0A0C4DH22
cor=0.2, p=0.047
1789: VEGFA|A2A2V4
cor=0.4, p=2e−04
1789: VEGFA|A2A2V4
cor=0.38, p=0.00033
1789: VEGFA|A2A2V4
cor=0.19, p=0.0023
2022: TPT1|Q5W0H4
cor=0.62, p=1.8e−28
2022: TPT1|Q5W0H4
cor=0.74, p=2.6e−50
2115: PIANP|F5H191
cor=0.74, p=2.6e−50
2115: PIANP|F5H191
cor=0.47, p=1.1e−17
1510: ADM|E9PL83
cor=0.68, p=1.6e−41
1510: ADM|E9PL83
cor=0.66, p=4.6e−13
2124: EFNA3|P52797−2
cor=0.64, p=3.8e−12
2124: EFNA3|P52797−2
cor=0.63, p=3.5e−22
1982: SUMO3|A8MU27
cor=0.63, p=3.5e−22
1982: SUMO3|A8MU27
cor=0.68, p=1.6e−41
1379: RIDA|H0YB34
cor=0.29, p=0.048
1924: EIF4B|E7EX17
cor=0.64, p=2.8e−35
1750: ICAM2|J3QQX6
cor=0.59, p=3.8e−29
1750: ICAM2|J3QQX6
cor=0.67, p=1.4e−34
1767: FTL|A0A087X1B9
cor=0.66, p=2.1e−38
318: NRP2|Q7LBX6
cor=0.68, p=1.6e−41
318: NRP2|Q7LBX6
cor=0.4, p=9.2e−13
1187: FCGR3A|H0Y755
cor=0.69, p=3.6e−43
299: GAS6|Q14393−2
cor=0.63, p=3.8e−34
153: AGRN|O00468−6
cor=0.65, p=6.3e−37
217: ATRN|O75882−2
cor=0.68, p=3e−41
1898: CSRP1|E9PP21
cor=0.61, p=8.8e−18
1928: EPHB4|Q96L35
cor=0.62, p=1.9e−21
1987: CKMT1A|F8WCN3
cor=0.61, p=5.5e−18
1661: PPP3R1|F6U1T9
cor=0.58, p=1.4e−25
1929: ANTXR2|A0A0C4DG84
cor=0.71, p=1.1e−46
1253: IL6R|A0A087WTB5
cor=0.73, p=1.6e−50
1253: IL6R|A0A087WTB5

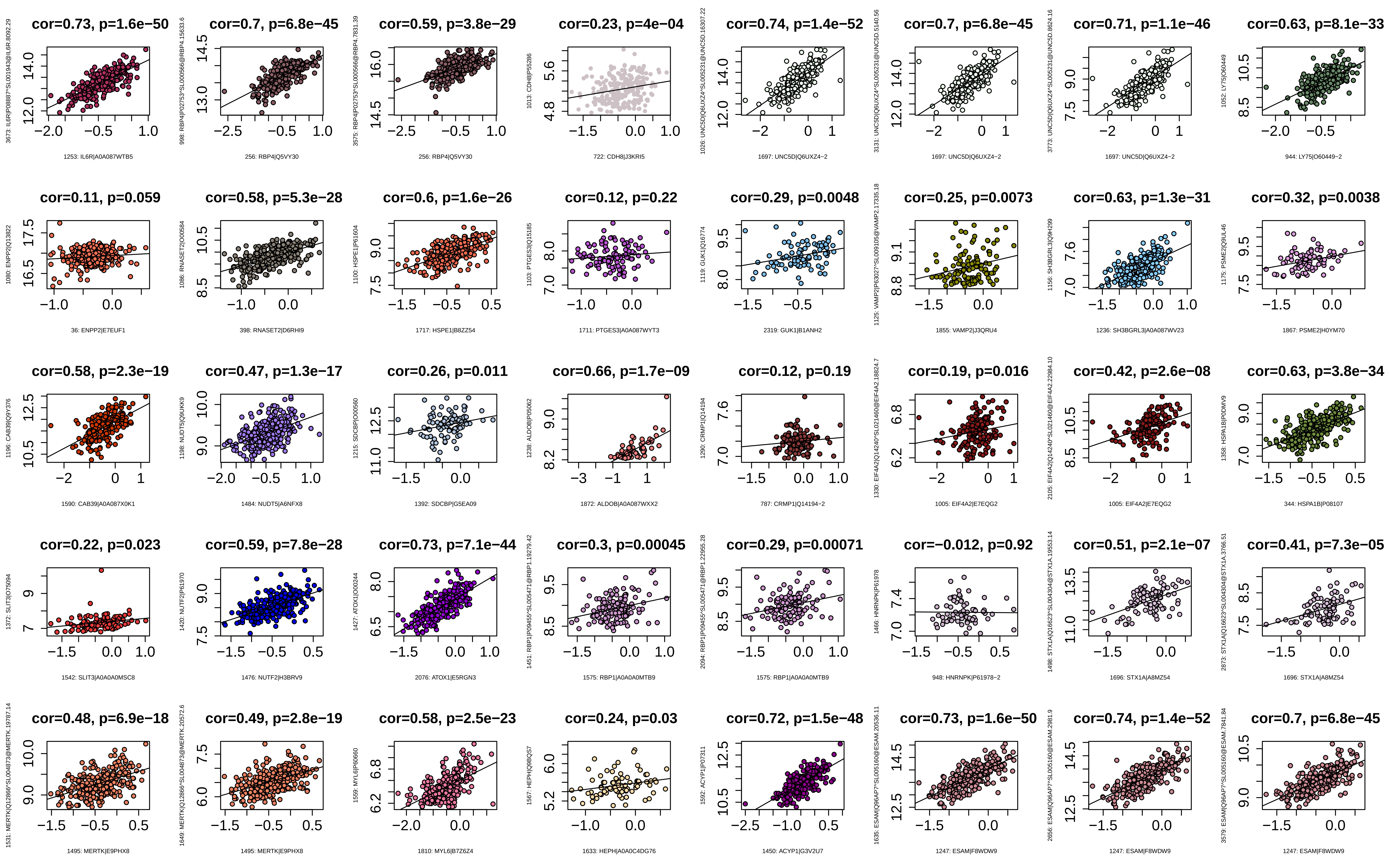
cor=0.73, p=1.6e−50
1253: IL6R|A0A087WTB5
cor=0.7, p=6.8e−45
256: RBP4|Q5VY30
cor=0.59, p=3.8e−29
256: RBP4|Q5VY30
cor=0.23, p=4e−04
722: CDH8|J3KRI5
cor=0.74, p=1.4e−52
1697: UNC5D|Q6UXZ4−2
cor=0.7, p=6.8e−45
1697: UNC5D|Q6UXZ4−2
cor=0.71, p=1.1e−46
1697: UNC5D|Q6UXZ4−2
cor=0.63, p=8.1e−33
944: LY75|O60449−2
cor=0.11, p=0.059
36: ENPP2|E7EUF1
cor=0.58, p=5.3e−28
398: RNASET2|D6RHI9
cor=0.6, p=1.6e−26
1717: HSPE1|B8ZZ54
cor=0.12, p=0.22
1711: PTGES3|A0A087WYT3
cor=0.29, p=0.0048
2319: GUK1|B1ANH2
cor=0.25, p=0.0073
1855: VAMP2|J3QRU4
cor=0.63, p=1.3e−31
1236: SH3BGRL3|A0A087WV23
cor=0.32, p=0.0038
1867: PSME2|H0YM70
cor=0.58, p=2.3e−19
1590: CAB39|A0A087X0K1
cor=0.47, p=1.3e−17
1484: NUDT5|A6NFX8
cor=0.26, p=0.011
1392: SDCBP|G5EA09
cor=0.66, p=1.7e−09
1872: ALDOB|A0A087WXX2
cor=0.12, p=0.19
787: CRMP1|Q14194−2
cor=0.19, p=0.016
1005: EIF4A2|E7EQG2
cor=0.42, p=2.6e−08
1005: EIF4A2|E7EQG2
cor=0.63, p=3.8e−34
344: HSPA1B|P08107
cor=0.22, p=0.023
1542: SLIT3|A0A0A0MSC8
cor=0.59, p=7.8e−28
1476: NUTF2|H3BRV9
cor=0.73, p=7.1e−44
2076: ATOX1|E5RGN3
cor=0.3, p=0.00045
1575: RBP1|A0A0A0MTB9
cor=0.29, p=0.00071
1575: RBP1|A0A0A0MTB9
cor=−0.012, p=0.92
948: HNRNPK|P61978−2
cor=0.51, p=2.1e−07
1696: STX1A|A8MZ54
cor=0.41, p=7.3e−05
1696: STX1A|A8MZ54
cor=0.48, p=6.9e−18
1495: MERTK|E9PHX8
cor=0.49, p=2.8e−19
1495: MERTK|E9PHX8
cor=0.58, p=2.5e−23
1810: MYL6|B7Z6Z4
cor=0.24, p=0.03
1633: HEPH|A0A0C4DG76
cor=0.72, p=1.5e−48
1450: ACYP1|G3V2U7
cor=0.73, p=1.6e−50
1247: ESAM|F8WDW9
cor=0.74, p=1.4e−52
1247: ESAM|F8WDW9
cor=0.7, p=6.8e−45
1247: ESAM|F8WDW9

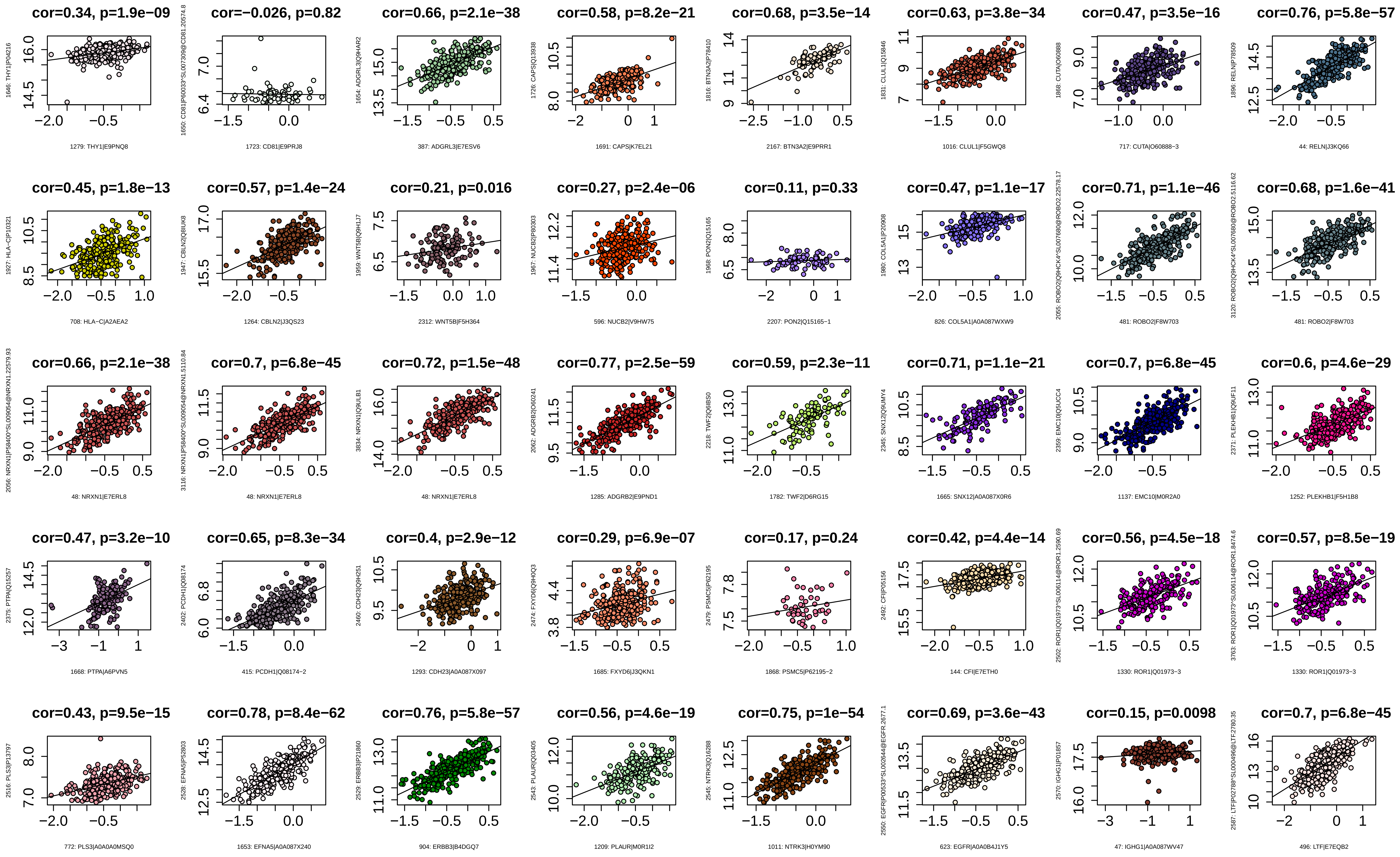

cor=0.34, p=1.9e−09
1646: THY1|P04216
1279: THY1|E9PNQ8
cor=−0.026, p=0.82
1723: CD81|E9PRJ8
cor=0.66, p=2.1e−38
1654: ADGRL3|Q9HAR2
387: ADGRL3|E7ESV6
cor=0.58, p=8.2e−21
1726: CAPS|Q13938
1691: CAPS|K7EL21
cor=0.68, p=3.5e−14
1816: BTN3A2|P78410
2167: BTN3A2|E9PRR1
cor=0.63, p=3.8e−34
1831: CLUL1|Q15846
1016: CLUL1|F5GWQ8
cor=0.47, p=3.5e−16
1868: CUTA|O60888
717: CUTA|O60888−3
cor=0.76, p=5.8e−57
1896: RELN|P78509
44: RELN|J3KQ66
cor=0.45, p=1.8e−13
1927: HLA−C|P10321
708: HLA−C|A2AEA2
cor=0.57, p=1.4e−24
1947: CBLN2|Q8IUK8
1264: CBLN2|J3QS23
cor=0.21, p=0.016
1959: WNT5B|Q9H1J7
2312: WNT5B|F5H364
cor=0.27, p=2.4e−06
1967: NUCB2|P80303
596: NUCB2|V9HW75
cor=0.11, p=0.33
1968: PON2|Q15165
2207: PON2|Q15165−1
cor=0.47, p=1.1e−17
1980: COL5A1|P20908
826: COL5A1|A0A087WXW9
cor=0.71, p=1.1e−46
481: ROBO2|F8W703
cor=0.68, p=1.6e−41
481: ROBO2|F8W703
cor=0.66, p=2.1e−38
48: NRXN1|E7ERL8
cor=0.7, p=6.8e−45
48: NRXN1|E7ERL8
cor=0.72, p=1.5e−48
3834: NRXN1|Q9ULB1
48: NRXN1|E7ERL8
cor=0.77, p=2.5e−59
2062: ADGRB2|O60241
1285: ADGRB2|E9PND1
cor=0.59, p=2.3e−11
2218: TWF2|Q6IBS0
1782: TWF2|D6RG15
cor=0.71, p=1.1e−21
2345: SNX12|Q9UMY4
1665: SNX12|A0A087X0R6
cor=0.7, p=6.8e−45
2359: EMC10|Q5UCC4
1137: EMC10|M0R2A0
cor=0.6, p=4.6e−29
2371: PLEKHB1|Q9UF11
1252: PLEKHB1|F5H1B8
cor=0.47, p=3.2e−10
2375: PTPA|Q15257
1668: PTPA|A6PVN5
cor=0.65, p=8.3e−34
2402: PCDH1|Q08174
415: PCDH1|Q08174−2
cor=0.4, p=2.9e−12
2460: CDH23|Q9H251
1293: CDH23|A0A087X097
cor=0.29, p=6.9e−07
2474: FXYD6|Q9H0Q3
1685: FXYD6|J3QKN1
cor=0.17, p=0.24
2479: PSMC5|P62195
1868: PSMC5|P62195−2
cor=0.42, p=4.4e−14
2492: CFI|P05156
144: CFI|E7ETH0
cor=0.56, p=4.5e−18
1330: ROR1|Q01973−3
cor=0.57, p=8.5e−19
1330: ROR1|Q01973−3
cor=0.43, p=9.5e−15
2516: PLS3|P13797
772: PLS3|A0A0A0MSQ0
cor=0.78, p=8.4e−62
2528: EFNA5|P52803
1653: EFNA5|A0A087X240
cor=0.76, p=5.8e−57
2529: ERBB3|P21860
904: ERBB3|B4DGQ7
cor=0.56, p=4.6e−19
2543: PLAUR|Q03405
1209: PLAUR|M0R1I2
cor=0.75, p=1e−54
2545: NTRK3|Q16288
1011: NTRK3|H0YM90
cor=0.69, p=3.6e−43
623: EGFR|A0A0B4J1Y5
cor=0.15, p=0.0098
2570: IGHG1|P01857
47: IGHG1|A0A087WV47
cor=0.7, p=6.8e−45
496: LTF|E7EQB2

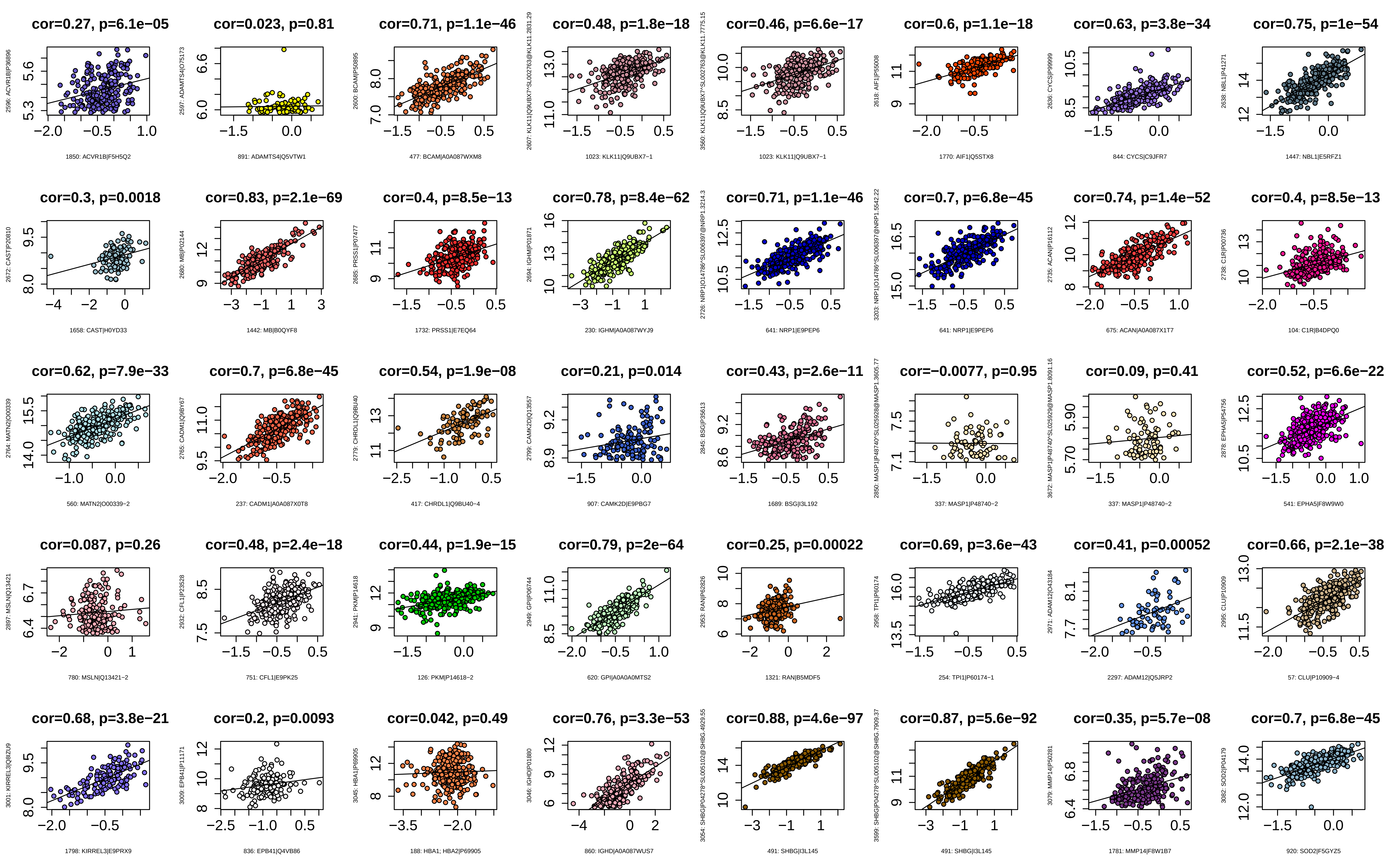

cor=0.27, p=6.1e−05
1850: ACVR1B|F5H5Q2
cor=0.023, p=0.81
891: ADAMTS4|Q5VTW1
cor=0.71, p=1.1e−46
477: BCAM|A0A087WXM8
cor=0.48, p=1.8e−18
1023: KLK11|Q9UBX7−1
cor=0.46, p=6.6e−17
1023: KLK11|Q9UBX7−1
cor=0.6, p=1.1e−18
1770: AIF1|Q5STX8
cor=0.63, p=3.8e−34
844: CYCS|C9JFR7
cor=0.75, p=1e−54
1447: NBL1|E5RFZ1
cor=0.3, p=0.0018
1658: CAST|H0YD33
cor=0.83, p=2.1e−69
1442: MB|B0QYF8
cor=0.4, p=8.5e−13
1732: PRSS1|E7EQ64
cor=0.78, p=8.4e−62
230: IGHM|A0A087WYJ9
cor=0.71, p=1.1e−46
641: NRP1|E9PEP6
cor=0.7, p=6.8e−45
641: NRP1|E9PEP6
cor=0.74, p=1.4e−52
675: ACAN|A0A087X1T7
cor=0.4, p=8.5e−13
104: C1R|B4DPQ0
cor=0.62, p=7.9e−33
560: MATN2|O00339−2
cor=0.7, p=6.8e−45
237: CADM1|A0A087X0T8
cor=0.54, p=1.9e−08
417: CHRDL1|Q9BU40−4
cor=0.21, p=0.014
907: CAMK2D|E9PBG7
cor=0.43, p=2.6e−11
1689: BSG|I3L192
cor=−0.0077, p=0.95
337: MASP1|P48740−2
cor=0.09, p=0.41
337: MASP1|P48740−2
cor=0.52, p=6.6e−22
541: EPHA5|F8W9W0
cor=0.087, p=0.26
780: MSLN|Q13421−2
cor=0.48, p=2.4e−18
751: CFL1|E9PK25
cor=0.44, p=1.9e−15
126: PKM|P14618−2
cor=0.79, p=2e−64
620: GPI|A0A0A0MTS2
cor=0.25, p=0.00022
1321: RAN|B5MDF5
cor=0.69, p=3.6e−43
254: TPI1|P60174−1
cor=0.41, p=0.00052
2297: ADAM12|Q5JRP2
cor=0.66, p=2.1e−38
57: CLU|P10909−4
cor=0.68, p=3.8e−21
1798: KIRREL3|E9PRX9
cor=0.2, p=0.0093
836: EPB41|Q4VB86
cor=0.042, p=0.49
188: HBA1; HBA2|P69905
cor=0.76, p=3.3e−53
860: IGHD|A0A087WUS7
cor=0.88, p=4.6e−97
491: SHBG|I3L145
cor=0.87, p=5.6e−92
491: SHBG|I3L145
cor=0.35, p=5.7e−08
1781: MMP14|F8W1B7
cor=0.7, p=6.8e−45
920: SOD2|F5GYZ5

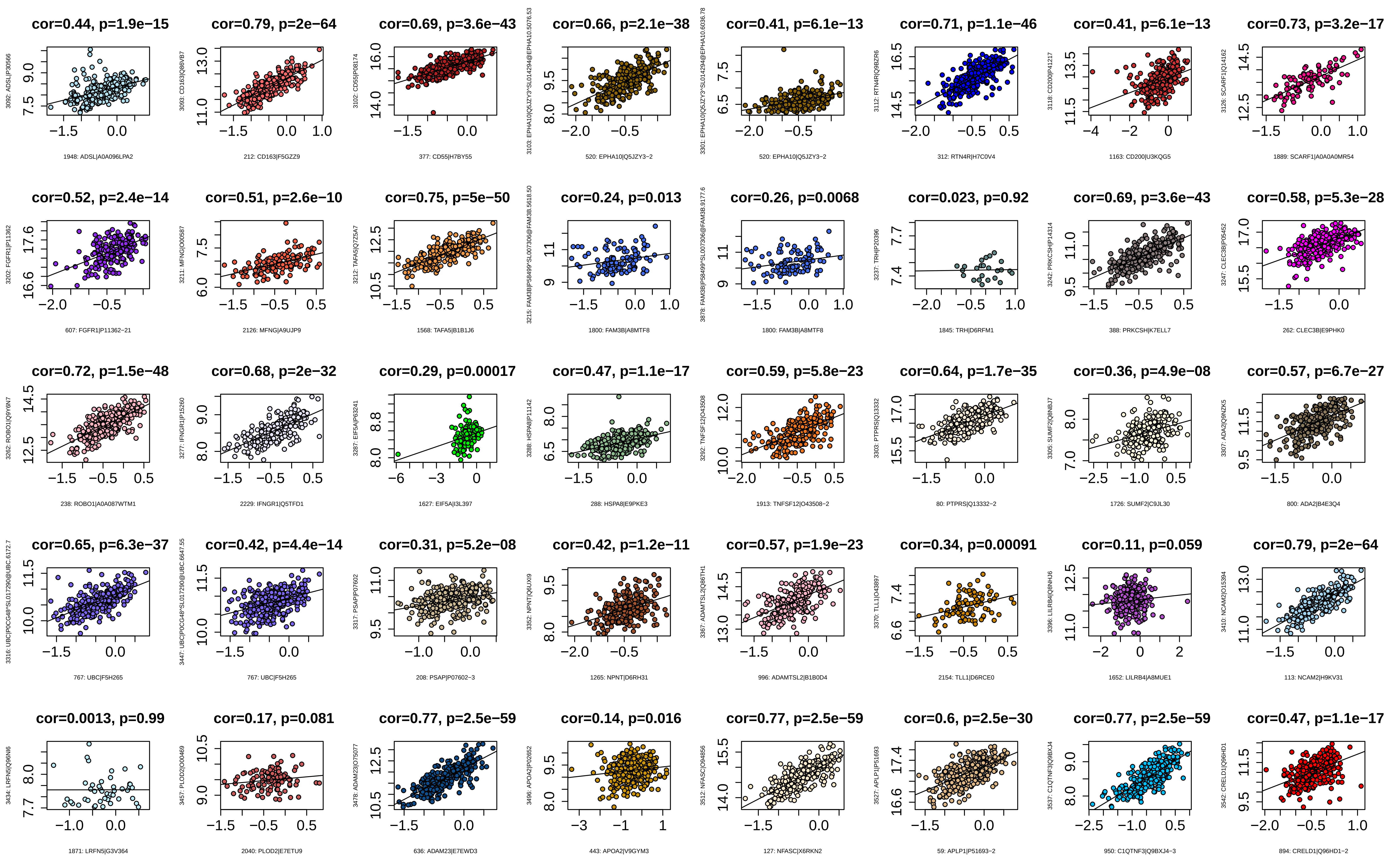

cor=0.44, p=1.9e−15
cor=0.79, p=2e−64
cor=0.69, p=3.6e−43
cor=0.66, p=2.1e−38
cor=0.41, p=6.1e−13
cor=0.71, p=1.1e−46
cor=0.41, p=6.1e−13
cor=0.73, p=3.2e−17
1948: ADSL|A0A096LPA2
212: CD163|F5GZZ9
377: CD55|H7BY55
520: EPHA10|Q5JZY3−2
520: EPHA10|Q5JZY3−2
312: RTN4R|H7C0V4
1163: CD200|U3KQG5
1889: SCARF1|A0A0A0MR54
cor=0.52, p=2.4e−14
cor=0.51, p=2.6e−10
cor=0.75, p=5e−50
cor=0.24, p=0.013
cor=0.26, p=0.0068
cor=0.023, p=0.92
cor=0.69, p=3.6e−43
cor=0.58, p=5.3e−28
607: FGFR1|P11362−21
2126: MFNG|A9UJP9
1568: TAFA5|B1B1J6
1800: FAM3B|A8MTF8
1800: FAM3B|A8MTF8
1845: TRH|D6RFM1
388: PRKCSH|K7ELL7
262: CLEC3B|E9PHK0
cor=0.72, p=1.5e−48
cor=0.68, p=2e−32
cor=0.29, p=0.00017
cor=0.47, p=1.1e−17
cor=0.59, p=5.8e−23
cor=0.64, p=1.7e−35
cor=0.36, p=4.9e−08
cor=0.57, p=6.7e−27
238: ROBO1|A0A087WTM1
2229: IFNGR1|Q5TFD1
1627: EIF5A|I3L397
288: HSPA8|E9PKE3
1913: TNFSF12|O43508−2
80: PTPRS|Q13332−2
1726: SUMF2|C9JL30
800: ADA2|B4E3Q4
cor=0.65, p=6.3e−37
cor=0.42, p=4.4e−14
cor=0.31, p=5.2e−08
cor=0.42, p=1.2e−11
cor=0.57, p=1.9e−23
cor=0.34, p=0.00091
cor=0.11, p=0.059
cor=0.79, p=2e−64
767: UBC|F5H265
767: UBC|F5H265
208: PSAP|P07602−3
1265: NPNT|D6RH31
996: ADAMTSL2|B1B0D4
2154: TLL1|D6RCE0
1652: LILRB4|A8MUE1
113: NCAM2|H9KV31
cor=0.0013, p=0.99
cor=0.17, p=0.081
cor=0.77, p=2.5e−59
cor=0.14, p=0.016
cor=0.77, p=2.5e−59
cor=0.6, p=2.5e−30
cor=0.77, p=2.5e−59
cor=0.47, p=1.1e−17
1871: LRFN5|G3V364
2040: PLOD2|E7ETU9
636: ADAM23|E7EWD3
443: APOA2|V9GYM3
127: NFASC|X6RKN2
59: APLP1|P51693−2
950: C1QTNF3|Q9BXJ4−3
894: CRELD1|Q96HD1−2

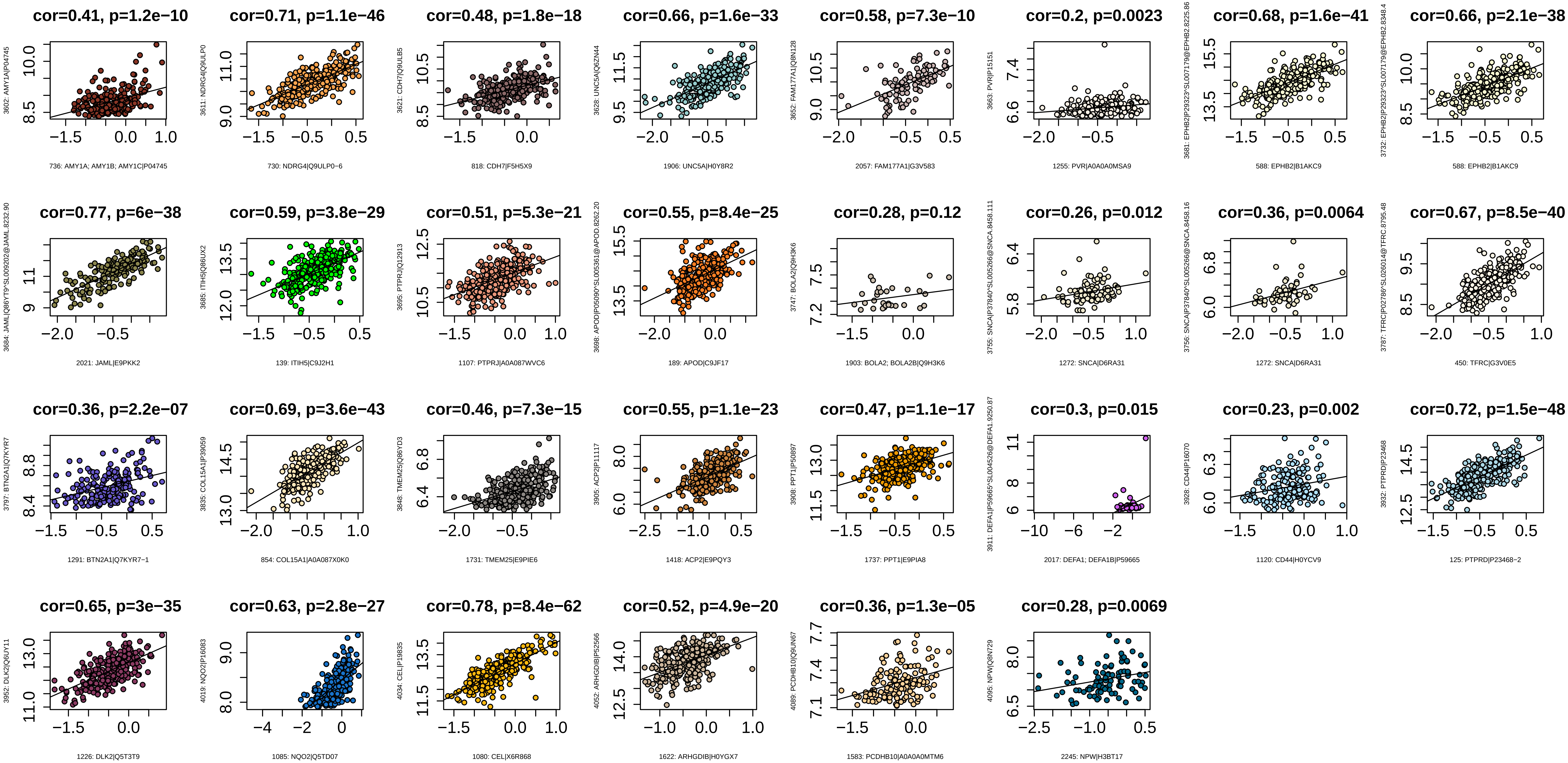
cor=0.41, p=1.2e−10
3602: AMY1A|P04745
736: AMY1A; AMY1B; AMY1C|P04745
cor=0.71, p=1.1e−46
3611: NDRG4|Q9ULP0
730: NDRG4|Q9ULP0−6
cor=0.48, p=1.8e−18
3621: CDH7|Q9ULB5
818: CDH7|F5H5X9
cor=0.66, p=1.6e−33
3628: UNC5A|Q6ZN44
1906: UNC5A|H0Y8R2
cor=0.58, p=7.3e−10
3652: FAM177A1|Q8N128
2057: FAM177A1|G3V583
cor=0.2, p=0.0023
3663: PVR|P15151
1255: PVR|A0A0A0MSA9
cor=0.68, p=1.6e−41
3681: EPHB2|P29323^SL007179@EPHB2.8225.86
588: EPHB2|B1AKC9
cor=0.66, p=2.1e−38
3732: EPHB2|P29323^SL007179@EPHB2.8348.4
588: EPHB2|B1AKC9
cor=0.77, p=6e−38
3684: JAML|Q86YT9^SL009202@JAML.8232.90
2021: JAML|E9PKK2
cor=0.59, p=3.8e−29
3685: ITIH5|Q86UX2
139: ITIH5|C9J2H1
cor=0.51, p=5.3e−21
3695: PTPRJ|Q12913
1107: PTPRJ|A0A087WVC6
cor=0.55, p=8.4e−25
3698: APOD|P05090^SL005361@APOD.8262.20
189: APOD|C9JF17
cor=0.28, p=0.12
3747: BOLA2|Q9H3K6
1903: BOLA2; BOLA2B|Q9H3K6
cor=0.26, p=0.012
3755: SNCA|P37840^SL005266@SNCA.8458.111
1272: SNCA|D6RA31
cor=0.36, p=0.0064
3756: SNCA|P37840^SL005266@SNCA.8458.16
1272: SNCA|D6RA31
cor=0.67, p=8.5e−40
3787: TFRC|P02786^SL026014@TFRC.8795.48
450: TFRC|G3V0E5
cor=0.36, p=2.2e−07
3797: BTN2A1|Q7KYR7
1291: BTN2A1|Q7KYR7−1
cor=0.69, p=3.6e−43
3835: COL15A1|P39059
854: COL15A1|A0A087X0K0
cor=0.46, p=7.3e−15
3848: TMEM25|Q86YD3
1731: TMEM25|E9PIE6
cor=0.55, p=1.1e−23
3905: ACP2|P11117
1418: ACP2|E9PQY3
cor=0.47, p=1.1e−17
3908: PPT1|P50897
1737: PPT1|E9PIA8
cor=0.3, p=0.015
3911: DEFA1|P59665^SL004526@DEFA1.9250.87
2017: DEFA1; DEFA1B|P59665
cor=0.23, p=0.002
3928: CD44|P16070
1120: CD44|H0YCV9
cor=0.72, p=1.5e−48
3932: PTPRD|P23468
125: PTPRD|P23468−2
cor=0.65, p=3e−35
3952: DLK2|Q6UY11
1226: DLK2|Q5T3T9
cor=0.63, p=2.8e−27
4019: NQO2|P16083
1085: NQO2|Q5TD07
cor=0.78, p=8.4e−62
4034: CEL|P19835
1080: CEL|X6R868
cor=0.52, p=4.9e−20
4052: ARHGDIB|P52566
1622: ARHGDIB|H0YGX7
cor=0.36, p=1.3e−05
4089: PCDHB10|Q9UN67
1583: PCDHB10|A0A0A0MTM6
cor=0.28, p=0.0069
4095: NPW|Q8N729
2245: NPW|H3BT17

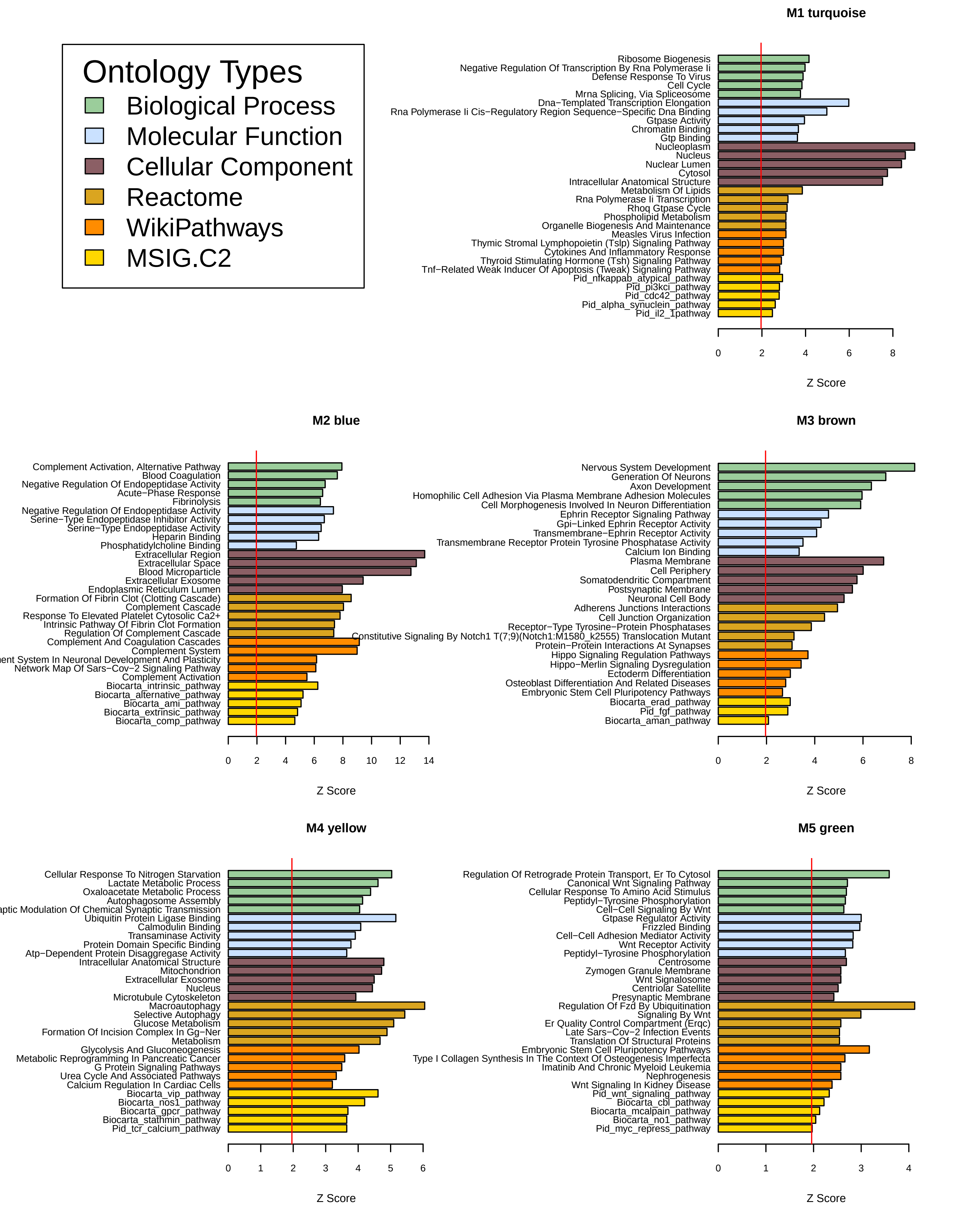
Ontology Types
Biological Process
Molecular Function
Cellular Component
Reactome
WikiPathways
MSIG.C2
M1 turquoise
Ribosome Biogenesis
Negative Regulation Of Transcription By Rna Polymerase Ii
Defense Response To Virus
Cell Cycle
Mrna Splicing, Via Spliceosome
Dna−Templated Transcription Elongation
Rna Polymerase Ii Cis−Regulatory Region Sequence−Specific Dna Binding
Gtpase Activity
Chromatin Binding
Gtp Binding
Nucleoplasm
Nucleus
Nuclear Lumen
Cytosol
Intracellular Anatomical Structure
Metabolism Of Lipids
Rna Polymerase Ii Transcription
Rhoq Gtpase Cycle
Phospholipid Metabolism
Organelle Biogenesis And Maintenance
Measles Virus Infection
Thymic Stromal Lymphopoietin (Tslp) Signaling Pathway
Cytokines And Inflammatory Response
Thyroid Stimulating Hormone (Tsh) Signaling Pathway
Tnf−Related Weak Inducer Of Apoptosis (Tweak) Signaling Pathway
Pid_nfkappab_atypical_pathway
Pid_pi3kci_pathway
Pid_cdc42_pathway
Pid_alpha_synuclein_pathway
Pid_il2_1pathway
0 2 4 6 8
Z Score
M2 blue
Complement Activation, Alternative Pathway
Blood Coagulation
Negative Regulation Of Endopeptidase Activity
Acute−Phase Response
Fibrinolysis
Negative Regulation Of Endopeptidase Activity
Serine−Type Endopeptidase Inhibitor Activity
Serine−Type Endopeptidase Activity
Heparin Binding
Phosphatidylcholine Binding
Extracellular Region
Extracellular Space
Blood Microparticle
Extracellular Exosome
Endoplasmic Reticulum Lumen
Formation Of Fibrin Clot (Clotting Cascade)
Complement Cascade
Response To Elevated Platelet Cytosolic Ca2+
Intrinsic Pathway Of Fibrin Clot Formation
Regulation Of Complement Cascade
Complement And Coagulation Cascades
Complement System
ent System In Neuronal Development And Plasticity
Network Map Of Sars−Cov−2 Signaling Pathway
Complement Activation
Biocarta_intrinsic_pathway
Biocarta_alternative_pathway
Biocarta_ami_pathway
Biocarta_extrinsic_pathway
Biocarta_comp_pathway
0 2 4 6 8 10 12 14
Z Score
M3 brown
Nervous System Development
Generation Of Neurons
Axon Development
Homophilic Cell Adhesion Via Plasma Membrane Adhesion Molecules
Cell Morphogenesis Involved In Neuron Differentiation
Ephrin Receptor Signaling Pathway
Gpi−Linked Ephrin Receptor Activity
Transmembrane−Ephrin Receptor Activity
Transmembrane Receptor Protein Tyrosine Phosphatase Activity
Calcium Ion Binding
Plasma Membrane
Cell Periphery
Somatodendritic Compartment
Postsynaptic Membrane
Neuronal Cell Body
Adherens Junctions Interactions
Cell Junction Organization
Receptor−Type Tyrosine−Protein Phosphatases
Constitutive Signaling By Notch1 T(7;9)(Notch1:M1580_k2555) Translocation Mutant
Protein−Protein Interactions At Synapses
Hippo Signaling Regulation Pathways
Hippo−Merlin Signaling Dysregulation
Ectoderm Differentiation
Osteoblast Differentiation And Related Diseases
Embryonic Stem Cell Pluripotency Pathways
Biocarta_erad_pathway
Pid_fgf_pathway
Biocarta_aman_pathway
0 2 4 6 8
Z Score
M4 yellow
Cellular Response To Nitrogen Starvation
Lactate Metabolic Process
Oxaloacetate Metabolic Process
Autophagosome Assembly
aptic Modulation Of Chemical Synaptic Transmission
Ubiquitin Protein Ligase Binding
Calmodulin Binding
Transaminase Activity
Protein Domain Specific Binding
Atp−Dependent Protein Disaggregase Activity
Intracellular Anatomical Structure
Mitochondrion
Extracellular Exosome
Nucleus
Microtubule Cytoskeleton
Macroautophagy
Selective Autophagy
Glucose Metabolism
Formation Of Incision Complex In Gg−Ner
Metabolism
Glycolysis And Gluconeogenesis
Metabolic Reprogramming In Pancreatic Cancer
G Protein Signaling Pathways
Urea Cycle And Associated Pathways
Calcium Regulation In Cardiac Cells
Biocarta_vip_pathway
Biocarta_nos1_pathway
Biocarta_gpcr_pathway
Biocarta_stathmin_pathway
Pid_tcr_calcium_pathway
0 1 2 3 4 5 6
Z Score
M5 green
Regulation Of Retrograde Protein Transport, Er To Cytosol
Canonical Wnt Signaling Pathway
Cellular Response To Amino Acid Stimulus
Peptidyl−Tyrosine Phosphorylation
Cell−Cell Signaling By Wnt
Gtpase Regulator Activity
Frizzled Binding
Cell−Cell Adhesion Mediator Activity
Wnt Receptor Activity
Peptidyl−Tyrosine Phosphorylation
Centrosome
Zymogen Granule Membrane
Wnt Signalosome
Centriolar Satellite
Presynaptic Membrane
Regulation Of Fzd By Ubiquitination
Signaling By Wnt
Er Quality Control Compartment (Erqc)
Late Sars−Cov−2 Infection Events
Translation Of Structural Proteins
Embryonic Stem Cell Pluripotency Pathways
Type I Collagen Synthesis In The Context Of Osteogenesis Imperfecta
Imatinib And Chronic Myeloid Leukemia
Nephrogenesis
Wnt Signaling In Kidney Disease
Pid_wnt_signaling_pathway
Biocarta_cbl_pathway
Biocarta_mcalpain_pathway
Biocarta_no1_pathway
Pid_myc_repress_pathway
0 1 2 3 4
Z Score

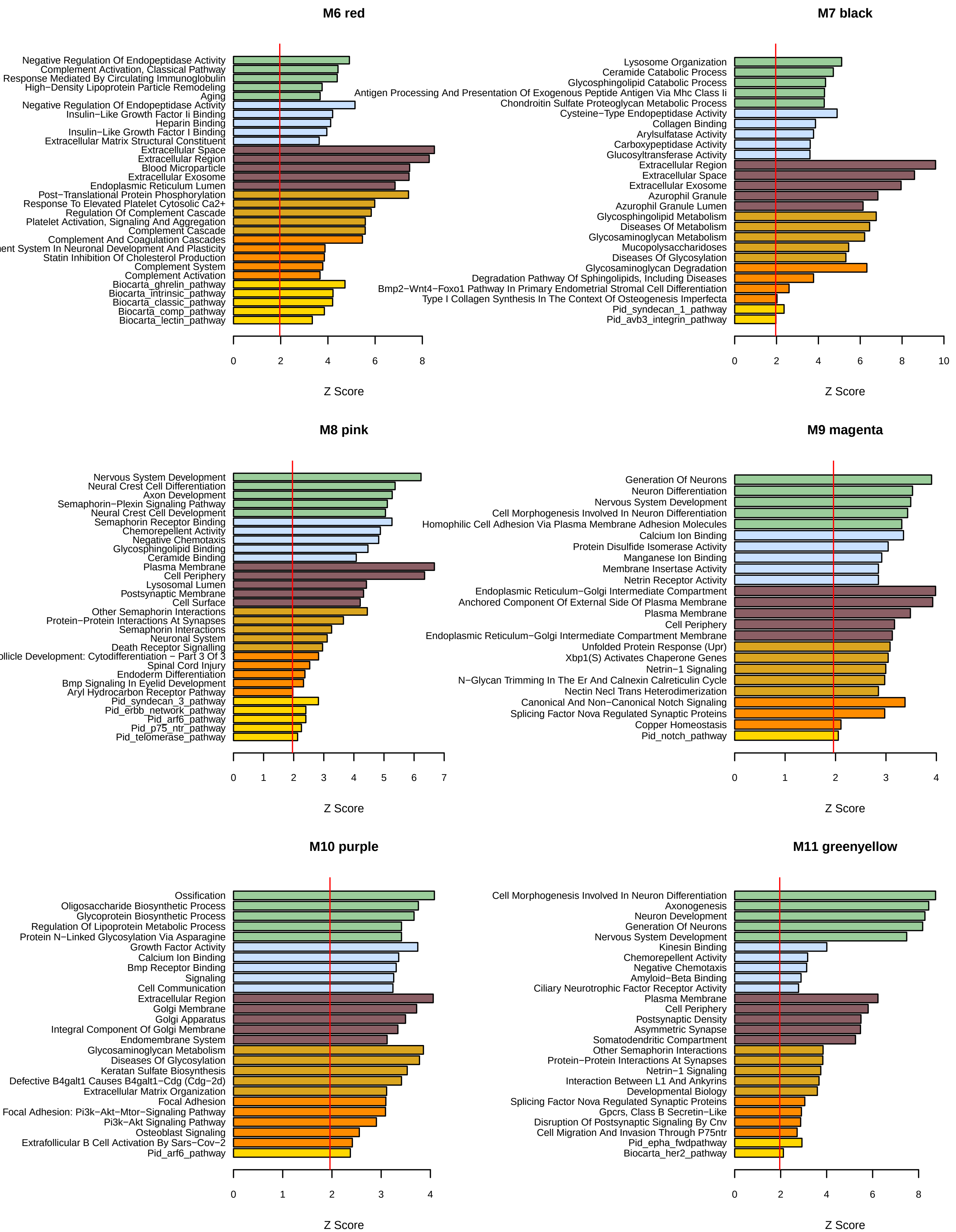
M6 red
Negative Regulation Of Endopeptidase Activity
Complement Activation, Classical Pathway
Response Mediated By Circulating Immunoglobulin
High−Density Lipoprotein Particle Remodeling
Aging
Negative Regulation Of Endopeptidase Activity
Insulin−Like Growth Factor Ii Binding
Heparin Binding
Insulin−Like Growth Factor I Binding
Extracellular Matrix Structural Constituent
Extracellular Space
Extracellular Region
Blood Microparticle
Extracellular Exosome
Endoplasmic Reticulum Lumen
Post−Translational Protein Phosphorylation
Response To Elevated Platelet Cytosolic Ca2+
Regulation Of Complement Cascade
Platelet Activation, Signaling And Aggregation
Complement Cascade
Complement And Coagulation Cascades
ent System In Neuronal Development And Plasticity
Statin Inhibition Of Cholesterol Production
Complement System
Complement Activation
Biocarta_ghrelin_pathway
Biocarta_intrinsic_pathway
Biocarta_classic_pathway
Biocarta_comp_pathway
Biocarta_lectin_pathway
0
2
4
6
8
Z Score
M7 black
Lysosome Organization
Ceramide Catabolic Process
Glycosphingolipid Catabolic Process
Antigen Processing And Presentation Of Exogenous Peptide Antigen Via Mhc Class Ii
Chondroitin Sulfate Proteoglycan Metabolic Process
Cysteine−Type Endopeptidase Activity
Collagen Binding
Arylsulfatase Activity
Carboxypeptidase Activity
Glucosyltransferase Activity
Extracellular Region
Extracellular Space
Extracellular Exosome
Azurophil Granule
Azurophil Granule Lumen
Glycosphingolipid Metabolism
Diseases Of Metabolism
Glycosaminoglycan Metabolism
Mucopolysaccharidoses
Diseases Of Glycosylation
Glycosaminoglycan Degradation
Degradation Pathway Of Sphingolipids, Including Diseases
Bmp2−Wnt4−Foxo1 Pathway In Primary Endometrial Stromal Cell Differentiation
Type I Collagen Synthesis In The Context Of Osteogenesis Imperfecta
Pid_syndecan_1_pathway
Pid_avb3_integrin_pathway
0
2
4
6
8
10
Z Score
M8 pink
Nervous System Development
Neural Crest Cell Differentiation
Axon Development
Semaphorin−Plexin Signaling Pathway
Neural Crest Cell Development
Semaphorin Receptor Binding
Chemorepellent Activity
Negative Chemotaxis
Glycosphingolipid Binding
Ceramide Binding
Plasma Membrane
Cell Periphery
Lysosomal Lumen
Postsynaptic Membrane
Cell Surface
Other Semaphorin Interactions
Protein−Protein Interactions At Synapses
Semaphorin Interactions
Neuronal System
Death Receptor Signalling
llicle Development: Cytodifferentiation − Part 3 Of 3
Spinal Cord Injury
Endoderm Differentiation
Bmp Signaling In Eyelid Development
Aryl Hydrocarbon Receptor Pathway
Pid_syndecan_3_pathway
Pid_erbb_network_pathway
Pid_arf6_pathway
Pid_p75_ntr_pathway
Pid_telomerase_pathway
0
1
2
3
4
5
6
7
Z Score
M9 magenta
Generation Of Neurons
Neuron Differentiation
Nervous System Development
Cell Morphogenesis Involved In Neuron Differentiation
Homophilic Cell Adhesion Via Plasma Membrane Adhesion Molecules
Calcium Ion Binding
Protein Disulfide Isomerase Activity
Manganese Ion Binding
Membrane Insertase Activity
Netrin Receptor Activity
Endoplasmic Reticulum−Golgi Intermediate Compartment
Anchored Component Of External Side Of Plasma Membrane
Plasma Membrane
Cell Periphery
Endoplasmic Reticulum−Golgi Intermediate Compartment Membrane
Unfolded Protein Response (Upr)
Xbp1(S) Activates Chaperone Genes
Netrin−1 Signaling
N−Glycan Trimming In The Er And Calnexin Calreticulin Cycle
Nectin Necl Trans Heterodimerization
Canonical And Non−Canonical Notch Signaling
Splicing Factor Nova Regulated Synaptic Proteins
Copper Homeostasis
Pid_notch_pathway
0
1
2
3
4
Z Score
M10 purple
Ossification
Oligosaccharide Biosynthetic Process
Glycoprotein Biosynthetic Process
Regulation Of Lipoprotein Metabolic Process
Protein N−Linked Glycosylation Via Asparagine
Growth Factor Activity
Calcium Ion Binding
Bmp Receptor Binding
Signaling
Cell Communication
Extracellular Region
Golgi Membrane
Golgi Apparatus
Integral Component Of Golgi Membrane
Endomembrane System
Glycosaminoglycan Metabolism
Diseases Of Glycosylation
Keratan Sulfate Biosynthesis
Defective B4galt1 Causes B4galt1−Cdg (Cdg−2d)
Extracellular Matrix Organization
Focal Adhesion
Focal Adhesion: Pi3k−Akt−Mtor−Signaling Pathway
Pi3k−Akt Signaling Pathway
Osteoblast Signaling
Extrafollicular B Cell Activation By Sars−Cov−2
Pid_arf6_pathway
0
1
2
3
4
Z Score
M11 greenyellow
Cell Morphogenesis Involved In Neuron Differentiation
Axonogenesis
Neuron Development
Generation Of Neurons
Nervous System Development
Kinesin Binding
Chemorepellent Activity
Negative Chemotaxis
Amyloid−Beta Binding
Ciliary Neurotrophic Factor Receptor Activity
Plasma Membrane
Cell Periphery
Postsynaptic Density
Asymmetric Synapse
Somatodendritic Compartment
Other Semaphorin Interactions
Protein−Protein Interactions At Synapses
Netrin−1 Signaling
Interaction Between L1 And Ankyrins
Developmental Biology
Splicing Factor Nova Regulated Synaptic Proteins
Gpcrs, Class B Secretin−Like
Disruption Of Postsynaptic Signaling By Cnv
Cell Migration And Invasion Through P75ntr
Pid_epha_fwdpathway
Biocarta_her2_pathway
0
2
4
6
8
Z Score

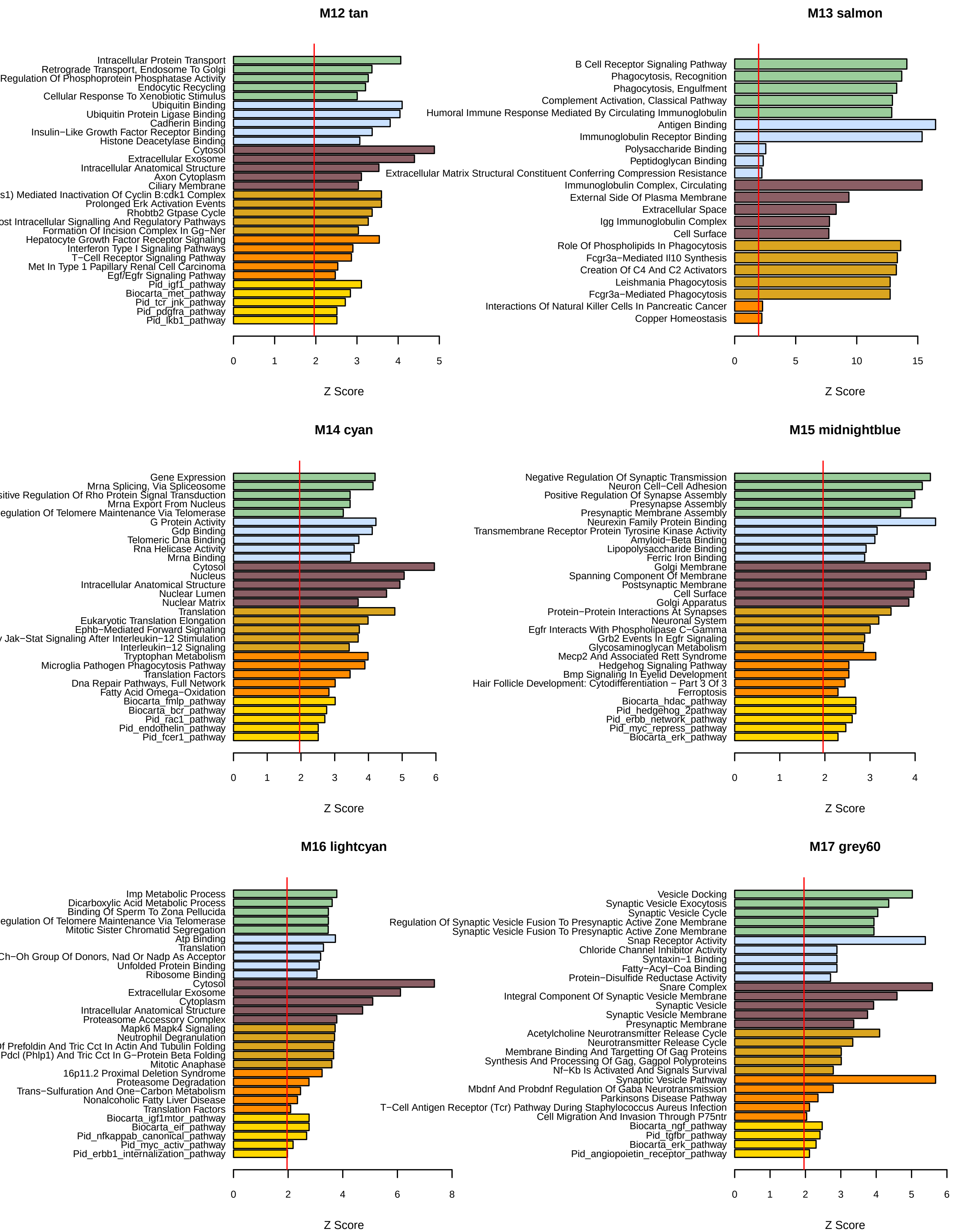
M12 tan
Intracellular Protein Transport
Retrograde Transport, Endosome To Golgi
Regulation Of Phosphoprotein Phosphatase Activity
Endocytic Recycling
Cellular Response To Xenobiotic Stimulus
Ubiquitin Binding
Ubiquitin Protein Ligase Binding
Cadherin Binding
Insulin−Like Growth Factor Receptor Binding
Histone Deacetylase Binding
Cytosol
Extracellular Exosome
Intracellular Anatomical Structure
Axon Cytoplasm
Ciliary Membrane
s1) Mediated Inactivation Of Cyclin B:cdk1 Complex
Prolonged Erk Activation Events
Rhobtb2 Gtpase Cycle
ost Intracellular Signalling And Regulatory Pathways
Formation Of Incision Complex In Gg−Ner
Hepatocyte Growth Factor Receptor Signaling
Interferon Type I Signaling Pathways
T−Cell Receptor Signaling Pathway
Met In Type 1 Papillary Renal Cell Carcinoma
Egf/Egfr Signaling Pathway
Pid_igf1_pathway
Biocarta_met_pathway
Pid_tcr_jnk_pathway
Pid_pdgfra_pathway
Pid_lkb1_pathway
0 1 2 3 4 5
Z Score
M13 salmon
B Cell Receptor Signaling Pathway
Phagocytosis, Recognition
Phagocytosis, Engulfment
Complement Activation, Classical Pathway
Humoral Immune Response Mediated By Circulating Immunoglobulin
Antigen Binding
Immunoglobulin Receptor Binding
Polysaccharide Binding
Peptidoglycan Binding
Extracellular Matrix Structural Constituent Conferring Compression Resistance
Immunoglobulin Complex, Circulating
External Side Of Plasma Membrane
Extracellular Space
Igg Immunoglobulin Complex
Cell Surface
Role Of Phospholipids In Phagocytosis
Fcgr3a−Mediated Il10 Synthesis
Creation Of C4 And C2 Activators
Leishmania Phagocytosis
Fcgr3a−Mediated Phagocytosis
Interactions Of Natural Killer Cells In Pancreatic Cancer
Copper Homeostasis
0 5 10 15
Z Score
M14 cyan
Gene Expression
Mrna Splicing, Via Spliceosome
sitive Regulation Of Rho Protein Signal Transduction
Mrna Export From Nucleus
egulation Of Telomere Maintenance Via Telomerase
G Protein Activity
Gdp Binding
Telomeric Dna Binding
Rna Helicase Activity
Mrna Binding
Cytosol
Nucleus
Intracellular Anatomical Structure
Nuclear Lumen
Nuclear Matrix
Translation
Eukaryotic Translation Elongation
Ephb−Mediated Forward Signaling
Jak−Stat Signaling After Interleukin−12 Stimulation
Interleukin−12 Signaling
Tryptophan Metabolism
Microglia Pathogen Phagocytosis Pathway
Translation Factors
Dna Repair Pathways, Full Network
Fatty Acid Omega−Oxidation
Biocarta_fmlp_pathway
Biocarta_bcr_pathway
Pid_rac1_pathway
Pid_endothelin_pathway
Pid_fcer1_pathway
0 1 2 3 4 5 6
Z Score
M15 midnightblue
Negative Regulation Of Synaptic Transmission
Neuron Cell−Cell Adhesion
Positive Regulation Of Synapse Assembly
Presynapse Assembly
Presynaptic Membrane Assembly
Neurexin Family Protein Binding
Transmembrane Receptor Protein Tyrosine Kinase Activity
Amyloid−Beta Binding
Lipopolysaccharide Binding
Ferric Iron Binding
Golgi Membrane
Spanning Component Of Membrane
Postsynaptic Membrane
Cell Surface
Golgi Apparatus
Protein−Protein Interactions At Synapses
Neuronal System
Egfr Interacts With Phospholipase C−Gamma
Grb2 Events In Egfr Signaling
Glycosaminoglycan Metabolism
Mecp2 And Associated Rett Syndrome
Hedgehog Signaling Pathway
Bmp Signaling In Eyelid Development
Hair Follicle Development: Cytodifferentiation − Part 3 Of 3
Ferroptosis
Biocarta_hdac_pathway
Pid_hedgehog_2pathway
Pid_erbb_network_pathway
Pid_myc_repress_pathway
Biocarta_erk_pathway
0 1 2 3 4
Z Score
M16 lightcyan
Imp Metabolic Process
Dicarboxylic Acid Metabolic Process
Binding Of Sperm To Zona Pellucida
egulation Of Telomere Maintenance Via Telomerase
Mitotic Sister Chromatid Segregation
Atp Binding
Translation
Ch−Oh Group Of Donors, Nad Or Nadp As Acceptor
Unfolded Protein Binding
Ribosome Binding
Cytosol
Extracellular Exosome
Cytoplasm
Intracellular Anatomical Structure
Proteasome Accessory Complex
Mapk6 Mapk4 Signaling
Neutrophil Degranulation
f Prefoldin And Tric Cct In Actin And Tubulin Folding
Pdcl (Phlp1) And Tric Cct In G−Protein Beta Folding
Mitotic Anaphase
16p11.2 Proximal Deletion Syndrome
Proteasome Degradation
Trans−Sulfuration And One−Carbon Metabolism
Nonalcoholic Fatty Liver Disease
Translation Factors
Biocarta_igf1mtor_pathway
Biocarta_eif_pathway
Pid_nfkappab_canonical_pathway
Pid_myc_activ_pathway
Pid_erbb1_internalization_pathway
0 2 4 6 8
Z Score
M17 grey60
Vesicle Docking
Synaptic Vesicle Exocytosis
Synaptic Vesicle Cycle
Regulation Of Synaptic Vesicle Fusion To Presynaptic Active Zone Membrane
Synaptic Vesicle Fusion To Presynaptic Active Zone Membrane
Snap Receptor Activity
Chloride Channel Inhibitor Activity
Syntaxin−1 Binding
Fatty−Acyl−Coa Binding
Protein−Disulfide Reductase Activity
Snare Complex
Integral Component Of Synaptic Vesicle Membrane
Synaptic Vesicle
Synaptic Vesicle Membrane
Presynaptic Membrane
Acetylcholine Neurotransmitter Release Cycle
Neurotransmitter Release Cycle
Membrane Binding And Targetting Of Gag Proteins
Synthesis And Processing Of Gag, Gagpol Polyproteins
Nf−Kb Is Activated And Signals Survival
Synaptic Vesicle Pathway
Mbdnf And Probdnf Regulation Of Gaba Neurotransmission
Parkinsons Disease Pathway
T−Cell Antigen Receptor (Tcr) Pathway During Staphylococcus Aureus Infection
Cell Migration And Invasion Through P75ntr
Biocarta_ngf_pathway
Pid_tgfbr_pathway
Biocarta_erk_pathway
Pid_angiopoietin_receptor_pathway
0 1 2 3 4 5 6
Z Score

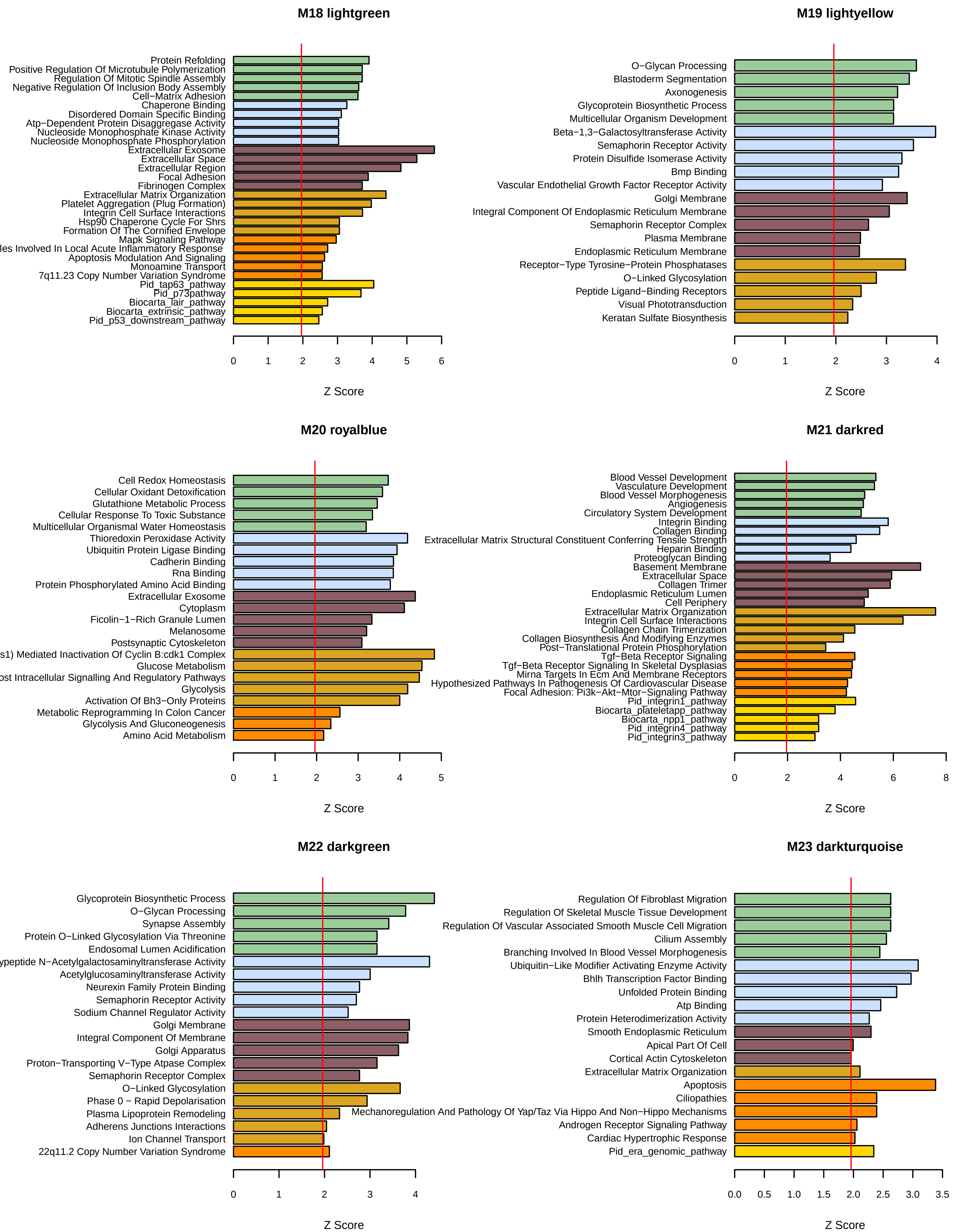

M18 lightgreen
Protein Refolding
Positive Regulation Of Microtubule Polymerization
Regulation Of Mitotic Spindle Assembly
Negative Regulation Of Inclusion Body Assembly
Cell−Matrix Adhesion
Chaperone Binding
Disordered Domain Specific Binding
Atp−Dependent Protein Disaggregase Activity
Nucleoside Monophosphate Kinase Activity
Nucleoside Monophosphate Phosphorylation
Extracellular Exosome
Extracellular Space
Extracellular Region
Focal Adhesion
Fibrinogen Complex
Extracellular Matrix Organization
Platelet Aggregation (Plug Formation)
Integrin Cell Surface Interactions
Hsp90 Chaperone Cycle For Shrs
Formation Of The Cornified Envelope
Mapk Signaling Pathway
les Involved In Local Acute Inflammatory Response
Apoptosis Modulation And Signaling
Monoamine Transport
7q11.23 Copy Number Variation Syndrome
Pid_tap63_pathway
Pid_p73pathway
Biocarta_lair_pathway
Biocarta_extrinsic_pathway
Pid_p53_downstream_pathway
0 1 2 3 4 5 6
Z Score
M19 lightyellow
O−Glycan Processing
Blastoderm Segmentation
Axonogenesis
Glycoprotein Biosynthetic Process
Multicellular Organism Development
Beta−1,3−Galactosyltransferase Activity
Semaphorin Receptor Activity
Protein Disulfide Isomerase Activity
Bmp Binding
Vascular Endothelial Growth Factor Receptor Activity
Golgi Membrane
Integral Component Of Endoplasmic Reticulum Membrane
Semaphorin Receptor Complex
Plasma Membrane
Endoplasmic Reticulum Membrane
Receptor−Type Tyrosine−Protein Phosphatases
O−Linked Glycosylation
Peptide Ligand−Binding Receptors
Visual Phototransduction
Keratan Sulfate Biosynthesis
0 1 2 3 4
Z Score
M20 royalblue
Cell Redox Homeostasis
Cellular Oxidant Detoxification
Glutathione Metabolic Process
Cellular Response To Toxic Substance
Multicellular Organismal Water Homeostasis
Thioredoxin Peroxidase Activity
Ubiquitin Protein Ligase Binding
Cadherin Binding
Rna Binding
Protein Phosphorylated Amino Acid Binding
Extracellular Exosome
Cytoplasm
Ficolin−1−Rich Granule Lumen
Melanosome
Postsynaptic Cytoskeleton
s1) Mediated Inactivation Of Cyclin B:cdk1 Complex
Glucose Metabolism
ost Intracellular Signalling And Regulatory Pathways
Glycolysis
Activation Of Bh3−Only Proteins
Metabolic Reprogramming In Colon Cancer
Glycolysis And Gluconeogenesis
Amino Acid Metabolism
0 1 2 3 4 5
Z Score
M21 darkred
Blood Vessel Development
Vasculature Development
Blood Vessel Morphogenesis
Angiogenesis
Circulatory System Development
Integrin Binding
Collagen Binding
Extracellular Matrix Structural Constituent Conferring Tensile Strength
Heparin Binding
Proteoglycan Binding
Basement Membrane
Extracellular Space
Collagen Trimer
Endoplasmic Reticulum Lumen
Cell Periphery
Extracellular Matrix Organization
Integrin Cell Surface Interactions
Collagen Chain Trimerization
Collagen Biosynthesis And Modifying Enzymes
Post−Translational Protein Phosphorylation
Tgf−Beta Receptor Signaling
Tgf−Beta Receptor Signaling In Skeletal Dysplasias
Mirna Targets In Ecm And Membrane Receptors
Hypothesized Pathways In Pathogenesis Of Cardiovascular Disease
Focal Adhesion: Pi3k−Akt−Mtor−Signaling Pathway
Pid_integrin1_pathway
Biocarta_plateletapp_pathway
Biocarta_npp1_pathway
Pid_integrin4_pathway
Pid_integrin3_pathway
0 2 4 6 8
Z Score
M22 darkgreen
Glycoprotein Biosynthetic Process
O−Glycan Processing
Synapse Assembly
Protein O−Linked Glycosylation Via Threonine
Endosomal Lumen Acidification
ypeptide N−Acetylgalactosaminyltransferase Activity
Acetylglucosaminyltransferase Activity
Neurexin Family Protein Binding
Semaphorin Receptor Activity
Sodium Channel Regulator Activity
Golgi Membrane
Integral Component Of Membrane
Golgi Apparatus
Proton−Transporting V−Type Atpase Complex
Semaphorin Receptor Complex
O−Linked Glycosylation
Phase 0 − Rapid Depolarisation
Plasma Lipoprotein Remodeling
Adherens Junctions Interactions
Ion Channel Transport
22q11.2 Copy Number Variation Syndrome
0 1 2 3 4
Z Score
M23 darkturquoise
Regulation Of Fibroblast Migration
Regulation Of Skeletal Muscle Tissue Development
Regulation Of Vascular Associated Smooth Muscle Cell Migration
Cilium Assembly
Branching Involved In Blood Vessel Morphogenesis
Ubiquitin−Like Modifier Activating Enzyme Activity
Bhlh Transcription Factor Binding
Unfolded Protein Binding
Atp Binding
Protein Heterodimerization Activity
Smooth Endoplasmic Reticulum
Apical Part Of Cell
Cortical Actin Cytoskeleton
Extracellular Matrix Organization
Apoptosis
Ciliopathies
Mechanoregulation And Pathology Of Yap/Taz Via Hippo And Non−Hippo Mechanisms
Androgen Receptor Signaling Pathway
Cardiac Hypertrophic Response
Pid_era_genomic_pathway
0.0 0.5 1.0 1.5 2.0 2.5 3.0 3.5
Z Score

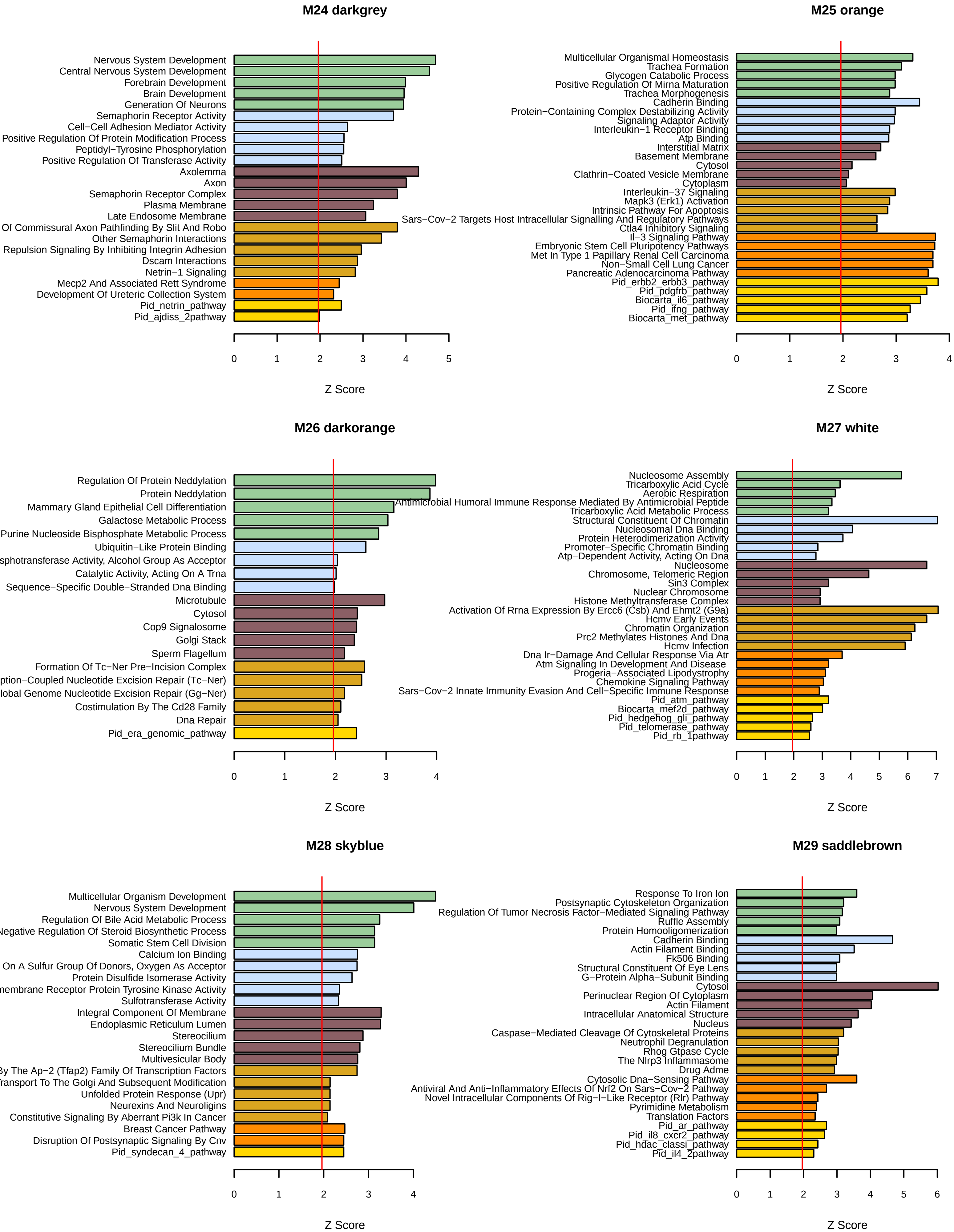
M24 darkgrey
Nervous System Development
Central Nervous System Development
Forebrain Development
Brain Development
Generation Of Neurons
Semaphorin Receptor Activity
Cell−Cell Adhesion Mediator Activity
Positive Regulation Of Protein Modification Process
Peptidyl−Tyrosine Phosphorylation
Positive Regulation Of Transferase Activity
Axolemma
Axon
Semaphorin Receptor Complex
Plasma Membrane
Late Endosome Membrane
Of Commissural Axon Pathfinding By Slit And Robo
Other Semaphorin Interactions
Repulsion Signaling By Inhibiting Integrin Adhesion
Dscam Interactions
Netrin−1 Signaling
Mecp2 And Associated Rett Syndrome
Development Of Ureteric Collection System
Pid_netrin_pathway
Pid_ajdiss_2pathway
0 1 2 3 4 5
Z Score
M25 orange
Multicellular Organismal Homeostasis
Trachea Formation
Glycogen Catabolic Process
Positive Regulation Of Mirna Maturation
Trachea Morphogenesis
Cadherin Binding
Protein−Containing Complex Destabilizing Activity
Signaling Adaptor Activity
Interleukin−1 Receptor Binding
Atp Binding
Interstitial Matrix
Basement Membrane
Cytosol
Clathrin−Coated Vesicle Membrane
Cytoplasm
Interleukin−37 Signaling
Mapk3 (Erk1) Activation
Intrinsic Pathway For Apoptosis
Sars−Cov−2 Targets Host Intracellular Signalling And Regulatory Pathways
Ctla4 Inhibitory Signaling
Il−3 Signaling Pathway
Embryonic Stem Cell Pluripotency Pathways
Met In Type 1 Papillary Renal Cell Carcinoma
Non−Small Cell Lung Cancer
Pancreatic Adenocarcinoma Pathway
Pid_erbb2_erbb3_pathway
Pid_pdgfrb_pathway
Biocarta_il6_pathway
Pid_ifng_pathway
Biocarta_met_pathway
0 1 2 3 4
Z Score
M26 darkorange
Regulation Of Protein Neddylation
Protein Neddylation
Mammary Gland Epithelial Cell Differentiation
Galactose Metabolic Process
Purine Nucleoside Bisphosphate Metabolic Process
Ubiquitin−Like Protein Binding
sphotransferase Activity, Alcohol Group As Acceptor
Catalytic Activity, Acting On A Trna
Sequence−Specific Double−Stranded Dna Binding
Microtubule
Cytosol
Cop9 Signalosome
Golgi Stack
Sperm Flagellum
Formation Of Tc−Ner Pre−Incision Complex
ption−Coupled Nucleotide Excision Repair (Tc−Ner)
lobal Genome Nucleotide Excision Repair (Gg−Ner)
Costimulation By The Cd28 Family
Dna Repair
Pid_era_genomic_pathway
0 1 2 3 4
Z Score
M27 white
Nucleosome Assembly
Tricarboxylic Acid Cycle
Aerobic Respiration
Antimicrobial Humoral Immune Response Mediated By Antimicrobial Peptide
Tricarboxylic Acid Metabolic Process
Structural Constituent Of Chromatin
Nucleosomal Dna Binding
Protein Heterodimerization Activity
Promoter−Specific Chromatin Binding
Atp−Dependent Activity, Acting On Dna
Nucleosome
Chromosome, Telomeric Region
Sin3 Complex
Nuclear Chromosome
Histone Methyltransferase Complex
Activation Of Rrna Expression By Ercc6 (Csb) And Ehmt2 (G9a)
Hcmv Early Events
Chromatin Organization
Prc2 Methylates Histones And Dna
Hcmv Infection
Dna Ir−Damage And Cellular Response Via Atr
Atm Signaling In Development And Disease
Progeria−Associated Lipodystrophy
Chemokine Signaling Pathway
Sars−Cov−2 Innate Immunity Evasion And Cell−Specific Immune Response
Pid_atm_pathway
Biocarta_mef2d_pathway
Pid_hedgehog_gli_pathway
Pid_telomerase_pathway
Pid_rb_1pathway
0 1 2 3 4 5 6 7
Z Score
M28 skyblue
Multicellular Organism Development
Nervous System Development
Regulation Of Bile Acid Metabolic Process
Negative Regulation Of Steroid Biosynthetic Process
Somatic Stem Cell Division
Calcium Ion Binding
On A Sulfur Group Of Donors, Oxygen As Acceptor
Protein Disulfide Isomerase Activity
nembrane Receptor Protein Tyrosine Kinase Activity
Sulfotransferase Activity
Integral Component Of Membrane
Endoplasmic Reticulum Lumen
Stereocilium
Stereocilium Bundle
Multivesicular Body
By The Ap−2 (Tfap2) Family Of Transcription Factors
ransport To The Golgi And Subsequent Modification
Unfolded Protein Response (Upr)
Neurexins And Neuroligins
Constitutive Signaling By Aberrant Pi3k In Cancer
Breast Cancer Pathway
Disruption Of Postsynaptic Signaling By Cnv
Pid_syndecan_4_pathway
0 1 2 3 4
Z Score
M29 saddlebrown
Response To Iron Ion
Postsynaptic Cytoskeleton Organization
Regulation Of Tumor Necrosis Factor−Mediated Signaling Pathway
Ruffle Assembly
Protein Homooligomerization
Cadherin Binding
Actin Filament Binding
Fk506 Binding
Structural Constituent Of Eye Lens
G−Protein Alpha−Subunit Binding
Cytosol
Perinuclear Region Of Cytoplasm
Actin Filament
Intracellular Anatomical Structure
Nucleus
Caspase−Mediated Cleavage Of Cytoskeletal Proteins
Neutrophil Degranulation
Rhog Gtpase Cycle
The Nlrp3 Inflammasome
Drug Adme
Cytosolic Dna−Sensing Pathway
Antiviral And Anti−Inflammatory Effects Of Nrf2 On Sars−Cov−2 Pathway
Novel Intracellular Components Of Rig−I−Like Receptor (Rlr) Pathway
Pyrimidine Metabolism
Translation Factors
Pid_ar_pathway
Pid_il8_cxcr2_pathway
Pid_hdac_classi_pathway
Pid_il4_2pathway
0 1 2 3 4 5 6
Z Score

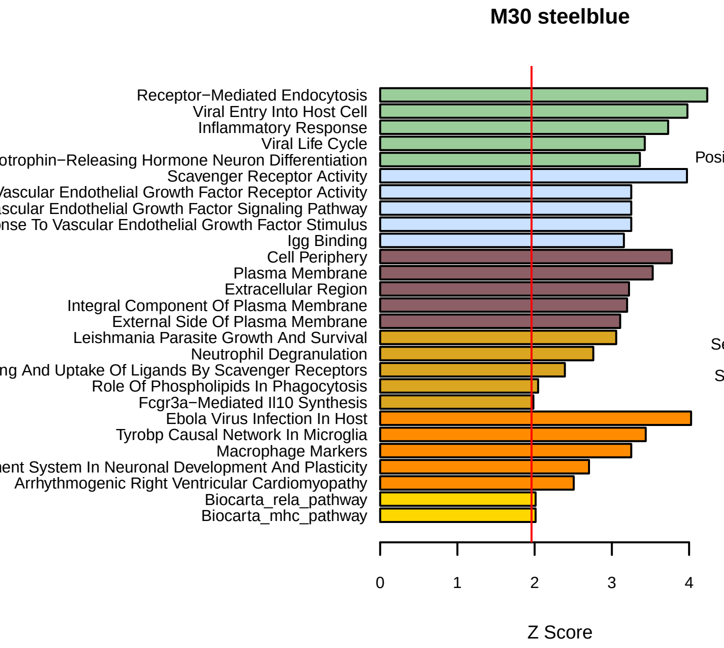
M30 steelblue
Receptor−Mediated Endocytosis
Viral Entry Into Host Cell
Inflammatory Response
Viral Life Cycle
otrophin−Releasing Hormone Neuron Differentiation
Scavenger Receptor Activity
ascular Endothelial Growth Factor Receptor Activity
scular Endothelial Growth Factor Signaling Pathway
nse To Vascular Endothelial Growth Factor Stimulus
Igg Binding
Cell Periphery
Plasma Membrane
Extracellular Region
Integral Component Of Plasma Membrane
External Side Of Plasma Membrane
Leishmania Parasite Growth And Survival
Neutrophil Degranulation
ng And Uptake Of Ligands By Scavenger Receptors
Role Of Phospholipids In Phagocytosis
Fcgr3a−Mediated Il10 Synthesis
Ebola Virus Infection In Host
Tyrobp Causal Network In Microglia
Macrophage Markers
ent System In Neuronal Development And Plasticity
Arrhythmogenic Right Ventricular Cardiomyopathy
Biocarta_rela_pathway
Biocarta_mhc_pathway
0
1
2
3
4
Z Score

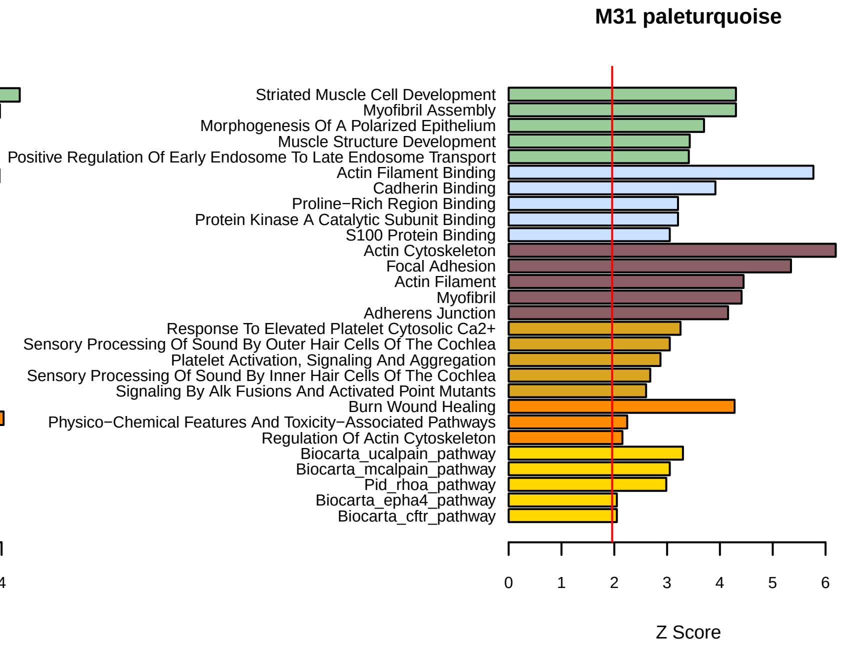
M31 paleturquoise
Striated Muscle Cell Development
Myofibril Assembly
Morphogenesis Of A Polarized Epithelium
Muscle Structure Development
Positive Regulation Of Early Endosome To Late Endosome Transport
Actin Filament Binding
Cadherin Binding
Proline−Rich Region Binding
Protein Kinase A Catalytic Subunit Binding
S100 Protein Binding
Actin Cytoskeleton
Focal Adhesion
Actin Filament
Myofibril
Adherens Junction
Response To Elevated Platelet Cytosolic Ca2+
Sensory Processing Of Sound By Outer Hair Cells Of The Cochlea
Platelet Activation, Signaling And Aggregation
Sensory Processing Of Sound By Inner Hair Cells Of The Cochlea
Signaling By Alk Fusions And Activated Point Mutants
Burn Wound Healing
Physico−Chemical Features And Toxicity−Associated Pathways
Regulation Of Actin Cytoskeleton
Biocarta_ucalpain_pathway
Biocarta_mcalpain_pathway
Pid_rhoa_pathway
Biocarta_epha4_pathway
Biocarta_cftr_pathway
0
1
2
3
4
5
6
Z Score

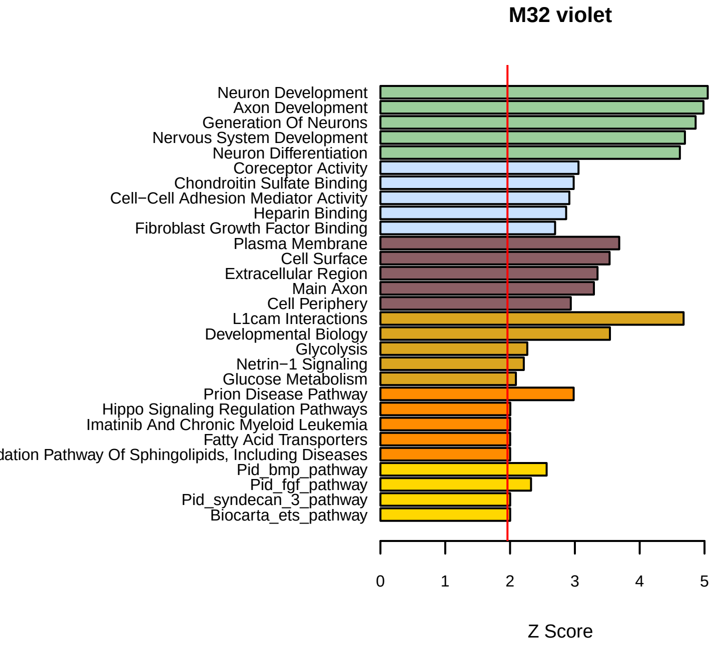
M32 violet
Neuron Development
Axon Development
Generation Of Neurons
Nervous System Development
Neuron Differentiation
Coreceptor Activity
Chondroitin Sulfate Binding
Cell−Cell Adhesion Mediator Activity
Heparin Binding
Fibroblast Growth Factor Binding
Plasma Membrane
Cell Surface
Extracellular Region
Main Axon
Cell Periphery
L1cam Interactions
Developmental Biology
Glycolysis
Netrin−1 Signaling
Glucose Metabolism
Prion Disease Pathway
Hippo Signaling Regulation Pathways
Imatinib And Chronic Myeloid Leukemia
Fatty Acid Transporters
dation Pathway Of Sphingolipids, Including Diseases
Pid_bmp_pathway
Pid_fgf_pathway
Pid_syndecan_3_pathway
Biocarta_ets_pathway
0
1
2
3
4
5
Z Score

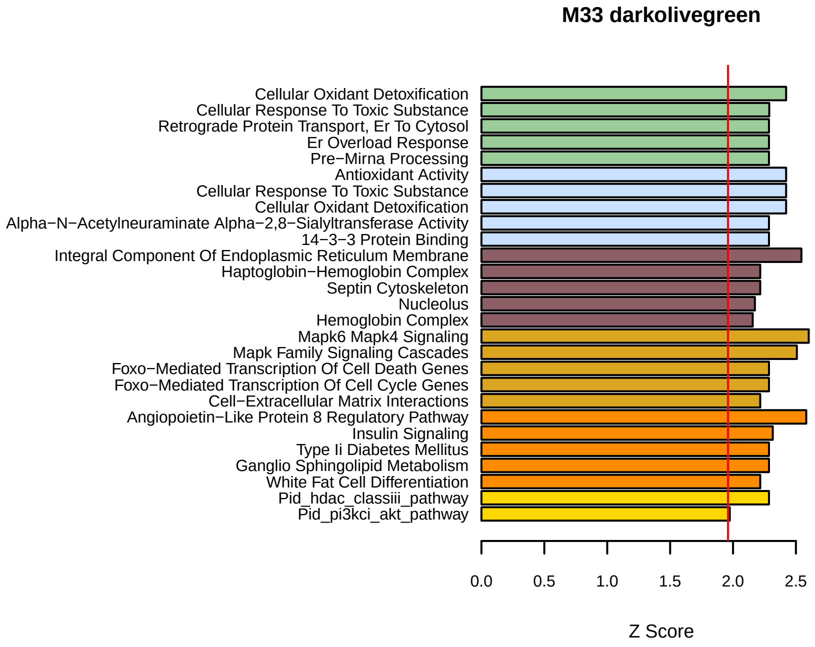
M33 darkolivegreen
Cellular Oxidant Detoxification
Cellular Response To Toxic Substance
Retrograde Protein Transport, Er To Cytosol
Er Overload Response
Pre−Mirna Processing
Antioxidant Activity
Cellular Response To Toxic Substance
Cellular Oxidant Detoxification
Alpha−N−Acetylneuraminate Alpha−2,8−Sialyltransferase Activity
14−3−3 Protein Binding
Integral Component Of Endoplasmic Reticulum Membrane
Haptoglobin−Hemoglobin Complex
Septin Cytoskeleton
Nucleolus
Hemoglobin Complex
Mapk6 Mapk4 Signaling
Mapk Family Signaling Cascades
Foxo−Mediated Transcription Of Cell Death Genes
Foxo−Mediated Transcription Of Cell Cycle Genes
Cell−Extracellular Matrix Interactions
Angiopoietin−Like Protein 8 Regulatory Pathway
Insulin Signaling
Type Ii Diabetes Mellitus
Ganglio Sphingolipid Metabolism
White Fat Cell Differentiation
Pid_hdac_classiii_pathway
Pid_pi3kci_akt_pathway
0.0
0.5
1.0
1.5
2.0
2.5
Z Score

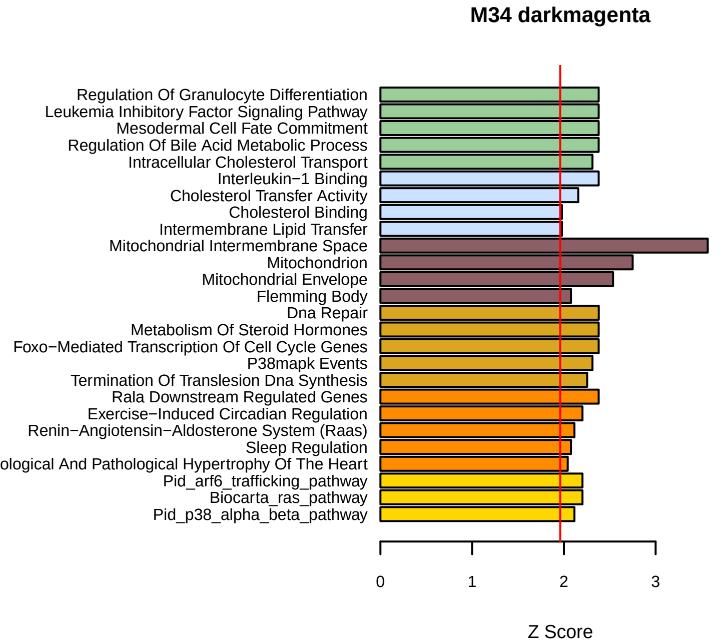
M34 darkmagenta
Regulation Of Granulocyte Differentiation
Leukemia Inhibitory Factor Signaling Pathway
Mesodermal Cell Fate Commitment
Regulation Of Bile Acid Metabolic Process
Intracellular Cholesterol Transport
Interleukin−1 Binding
Cholesterol Transfer Activity
Cholesterol Binding
Intermembrane Lipid Transfer
Mitochondrial Intermembrane Space
Mitochondrion
Mitochondrial Envelope
Flemming Body
Dna Repair
Metabolism Of Steroid Hormones
Foxo−Mediated Transcription Of Cell Cycle Genes
P38mapk Events
Termination Of Translesion Dna Synthesis
Rala Downstream Regulated Genes
Exercise−Induced Circadian Regulation
Renin−Angiotensin−Aldosterone System (Raas)
Sleep Regulation
ological And Pathological Hypertrophy Of The Heart
Pid_arf6_trafficking_pathway
Biocarta_ras_pathway
Pid_p38_alpha_beta_pathway
0
1
2
3
Z Score

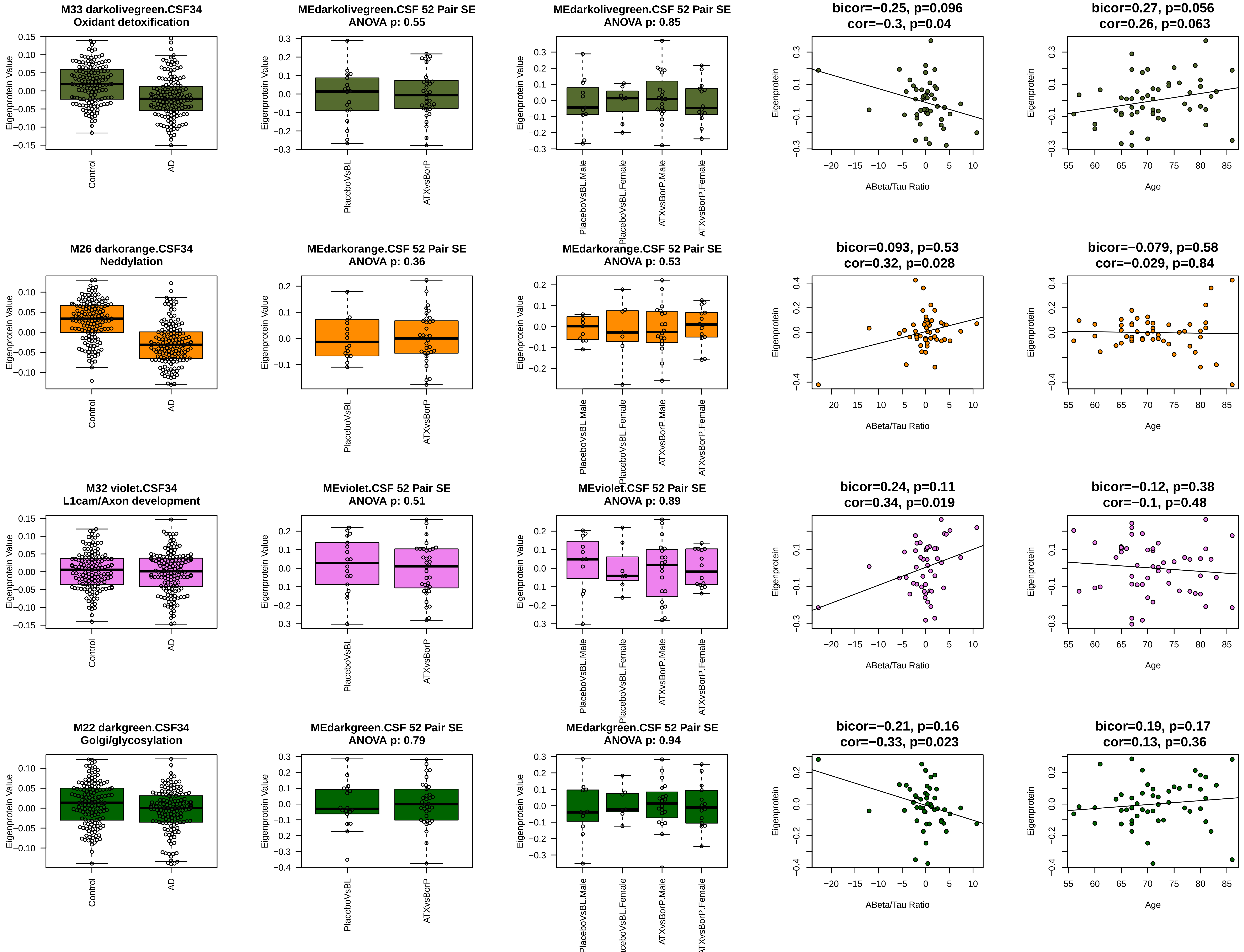
M33 darkolivegreen.CSF34
Oxidant detoxification
MEdarkolivegreen.CSF 52 Pair SE
ANOVA p: 0.55
MEdarkolivegreen.CSF 52 Pair SE
ANOVA p: 0.85
bicor=−0.25, p=0.096
cor=−0.3, p=0.04
bicor=0.27, p=0.056
cor=0.26, p=0.063
M26 darkorange.CSF34
Neddylation
MEdarkorange.CSF 52 Pair SE
ANOVA p: 0.36
MEdarkorange.CSF 52 Pair SE
ANOVA p: 0.53
bicor=0.093, p=0.53
cor=0.32, p=0.028
bicor=−0.079, p=0.58
cor=−0.029, p=0.84
M32 violet.CSF34
L1cam/Axon development
MEviolet.CSF 52 Pair SE
ANOVA p: 0.51
MEviolet.CSF 52 Pair SE
ANOVA p: 0.89
bicor=0.24, p=0.11
cor=0.34, p=0.019
bicor=−0.12, p=0.38
cor=−0.1, p=0.48
M22 darkgreen.CSF34
Golgi/glycosylation
MEdarkgreen.CSF 52 Pair SE
ANOVA p: 0.79
MEdarkgreen.CSF 52 Pair SE
ANOVA p: 0.94
bicor=−0.21, p=0.16
cor=−0.33, p=0.023
bicor=0.19, p=0.17
cor=0.13, p=0.36
Eigenprotein Value
Eigenprotein
Control
AD
PlaceboVsBL
ATXvsBorP
PlaceboVsBL.Male
PlaceboVsBL.Female
ATXvsBorP.Male
ATXvsBorP.Female
ABeta/Tau Ratio
Age

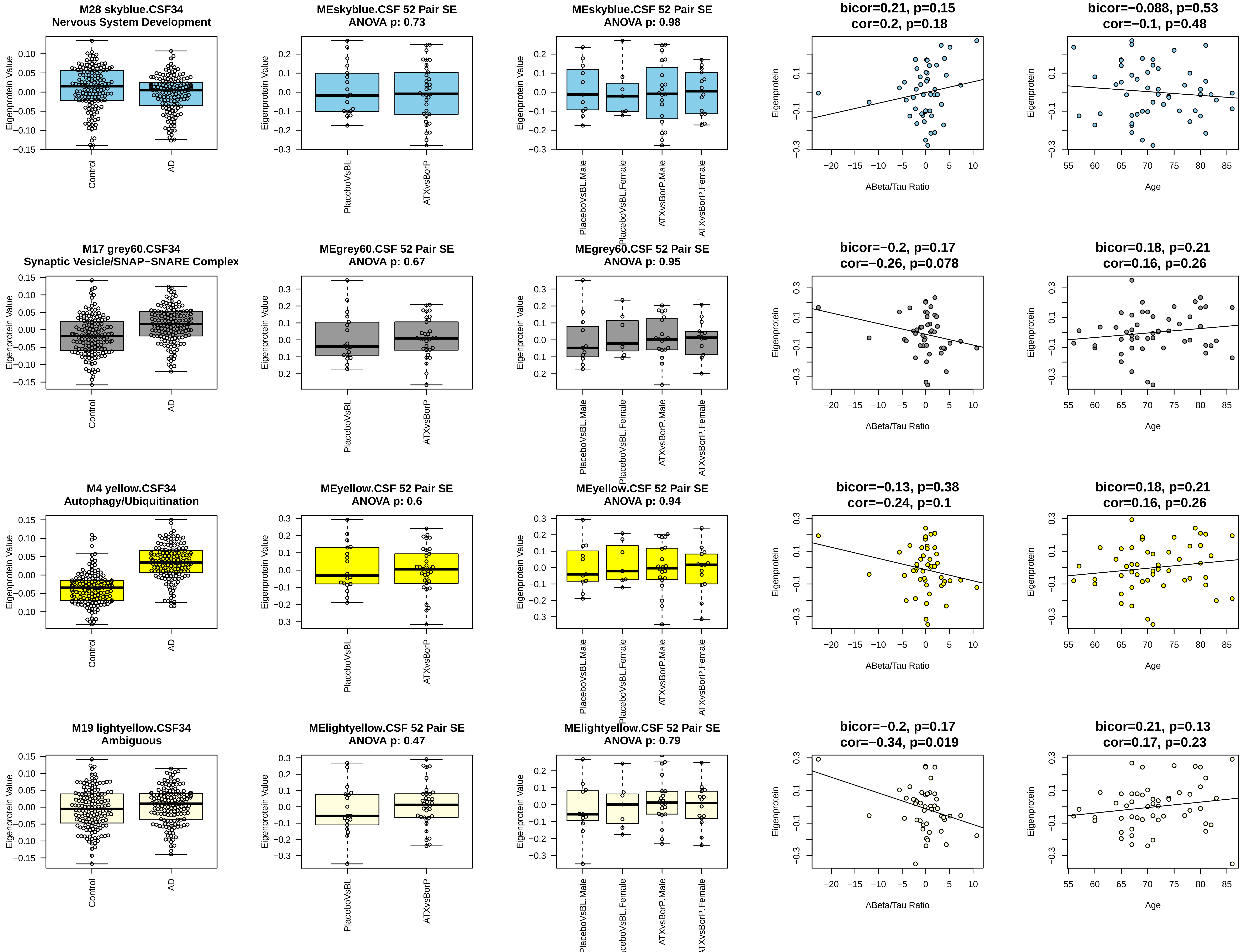
M28 skyblue.CSF34
Nervous System Development
MEskyblue.CSF 52 Pair SE
ANOVA p: 0.73
MEskyblue.CSF 52 Pair SE
ANOVA p: 0.98
bicor=0.21, p=0.15
cor=0.2, p=0.18
bicor=−0.088, p=0.53
cor=−0.1, p=0.48
M17 grey60.CSF34
Synaptic Vesicle/SNAP−SNARE Complex
MEgrey60.CSF 52 Pair SE
ANOVA p: 0.67
MEgrey60.CSF 52 Pair SE
ANOVA p: 0.95
bicor=−0.2, p=0.17
cor=−0.26, p=0.078
bicor=0.18, p=0.21
cor=0.16, p=0.26
M4 yellow.CSF34
Autophagy/Ubiquitination
MEyellow.CSF 52 Pair SE
ANOVA p: 0.6
MEyellow.CSF 52 Pair SE
ANOVA p: 0.94
bicor=−0.13, p=0.38
cor=−0.24, p=0.1
bicor=0.18, p=0.21
cor=0.16, p=0.26
M19 lightyellow.CSF34
Ambiguous
MElightyellow.CSF 52 Pair SE
ANOVA p: 0.47
MElightyellow.CSF 52 Pair SE
ANOVA p: 0.79
bicor=−0.2, p=0.17
cor=−0.34, p=0.019
bicor=0.21, p=0.13
cor=0.17, p=0.23
Eigenprotein Value
Eigenprotein
Control
AD
PlaceboVsBL
ATXvsBorP
PlaceboVsBL.Male
PlaceboVsBL.Female
ATXvsBorP.Male
ATXvsBorP.Female
ABeta/Tau Ratio
Age

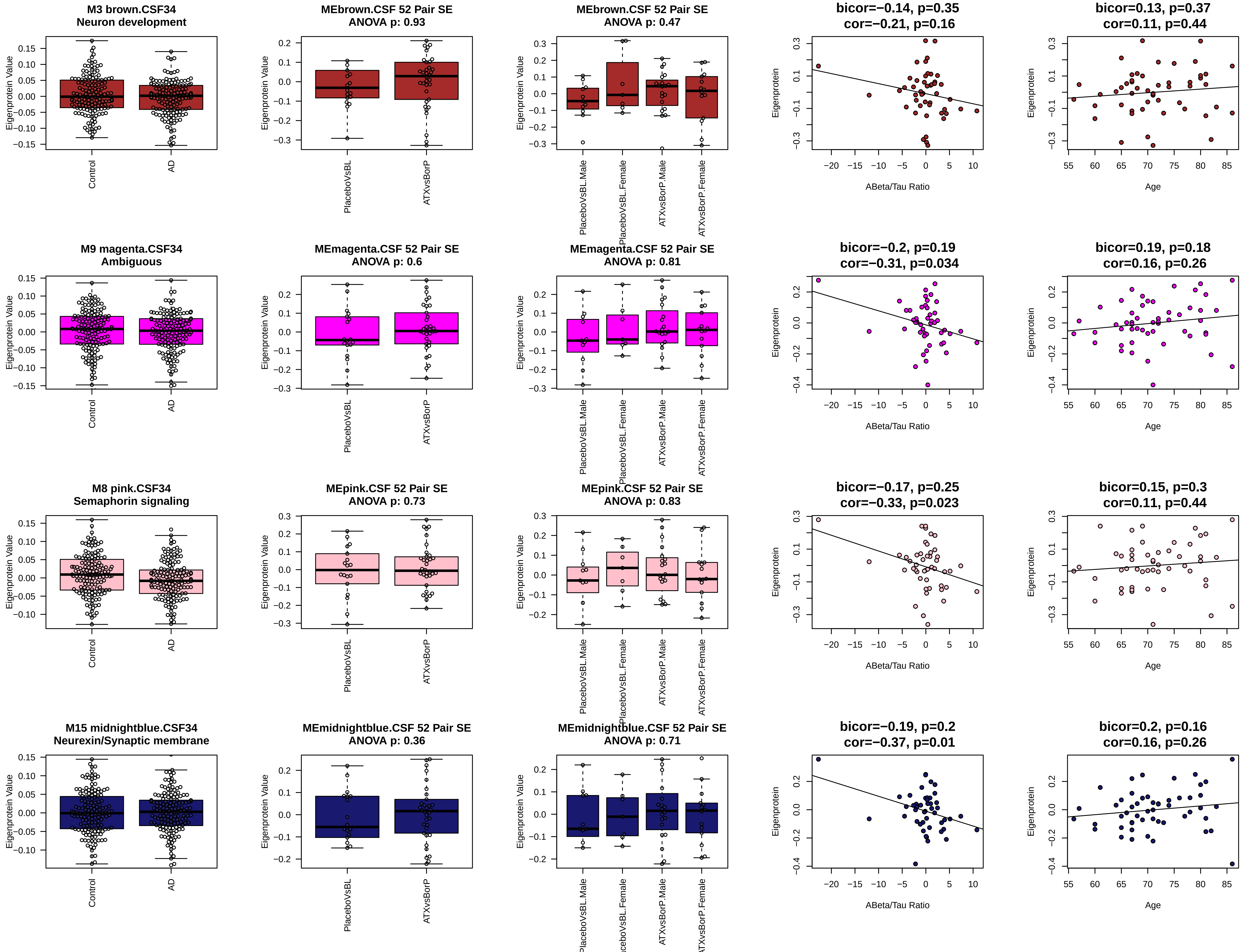

M3 brown.CSF34
Neuron development
MEbrown.CSF 52 Pair SE
ANOVA p: 0.93
MEbrown.CSF 52 Pair SE
ANOVA p: 0.47
bicor=−0.14, p=0.35
cor=−0.21, p=0.16
bicor=0.13, p=0.37
cor=0.11, p=0.44
M9 magenta.CSF34
Ambiguous
MEmagenta.CSF 52 Pair SE
ANOVA p: 0.6
MEmagenta.CSF 52 Pair SE
ANOVA p: 0.81
bicor=−0.2, p=0.19
cor=−0.31, p=0.034
bicor=0.19, p=0.18
cor=0.16, p=0.26
M8 pink.CSF34
Semaphorin signaling
MEpink.CSF 52 Pair SE
ANOVA p: 0.73
MEpink.CSF 52 Pair SE
ANOVA p: 0.83
bicor=−0.17, p=0.25
cor=−0.33, p=0.023
bicor=0.15, p=0.3
cor=0.11, p=0.44
M15 midnightblue.CSF34
Neurexin/Synaptic membrane
MEmidnightblue.CSF 52 Pair SE
ANOVA p: 0.36
MEmidnightblue.CSF 52 Pair SE
ANOVA p: 0.71
bicor=−0.19, p=0.2
cor=−0.37, p=0.01
bicor=0.2, p=0.16
cor=0.16, p=0.26
Eigenprotein Value
Eigenprotein
Control
AD
PlaceboVsBL
ATXvsBorP
PlaceboVsBL.Male
PlaceboVsBL.Female
ATXvsBorP.Male
ATXvsBorP.Female
ABeta/Tau Ratio
Age

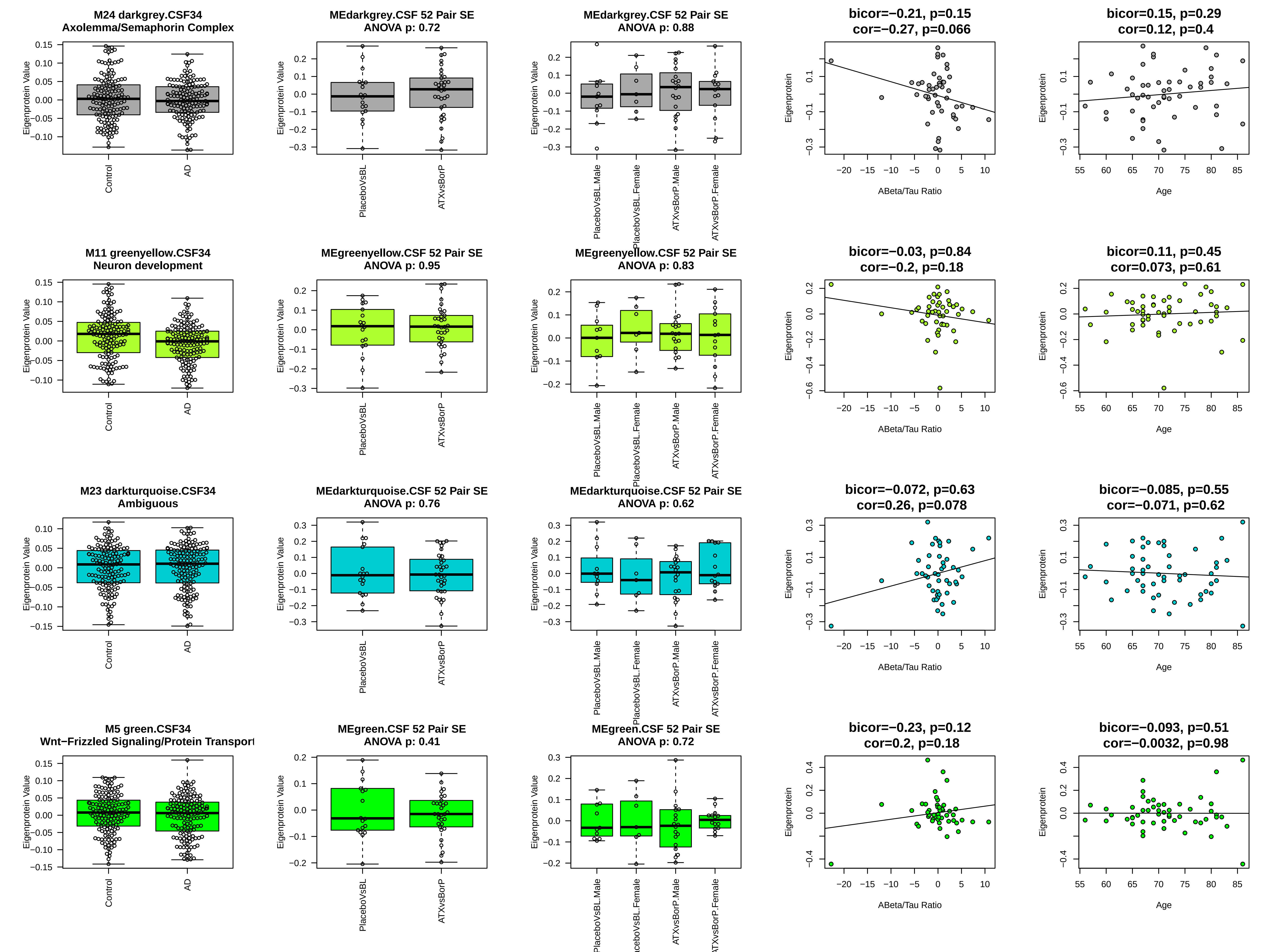
M24 darkgrey.CSF34
Axolemma/Semaphorin Complex
MEdarkgrey.CSF 52 Pair SE
ANOVA p: 0.72
MEdarkgrey.CSF 52 Pair SE
ANOVA p: 0.88
bicor=−0.21, p=0.15
cor=−0.27, p=0.066
bicor=0.15, p=0.29
cor=0.12, p=0.4
M11 greenyellow.CSF34
Neuron development
MEgreenyellow.CSF 52 Pair SE
ANOVA p: 0.95
MEgreenyellow.CSF 52 Pair SE
ANOVA p: 0.83
bicor=−0.03, p=0.84
cor=−0.2, p=0.18
bicor=0.11, p=0.45
cor=0.073, p=0.61
M23 darkturquoise.CSF34
Ambiguous
MEdarkturquoise.CSF 52 Pair SE
ANOVA p: 0.76
MEdarkturquoise.CSF 52 Pair SE
ANOVA p: 0.62
bicor=−0.072, p=0.63
cor=0.26, p=0.078
bicor=−0.085, p=0.55
cor=−0.071, p=0.62
M5 green.CSF34
Wnt−Frizzled Signaling/Protein Transport
MEgreen.CSF 52 Pair SE
ANOVA p: 0.41
MEgreen.CSF 52 Pair SE
ANOVA p: 0.72
bicor=−0.23, p=0.12
cor=0.2, p=0.18
bicor=−0.093, p=0.51
cor=−0.0032, p=0.98
Eigenprotein Value
Eigenprotein
Control
AD
PlaceboVsBL
ATXvsBorP
PlaceboVsBL.Male
PlaceboVsBL.Female
ATXvsBorP.Male
ATXvsBorP.Female
ABeta/Tau Ratio
Age

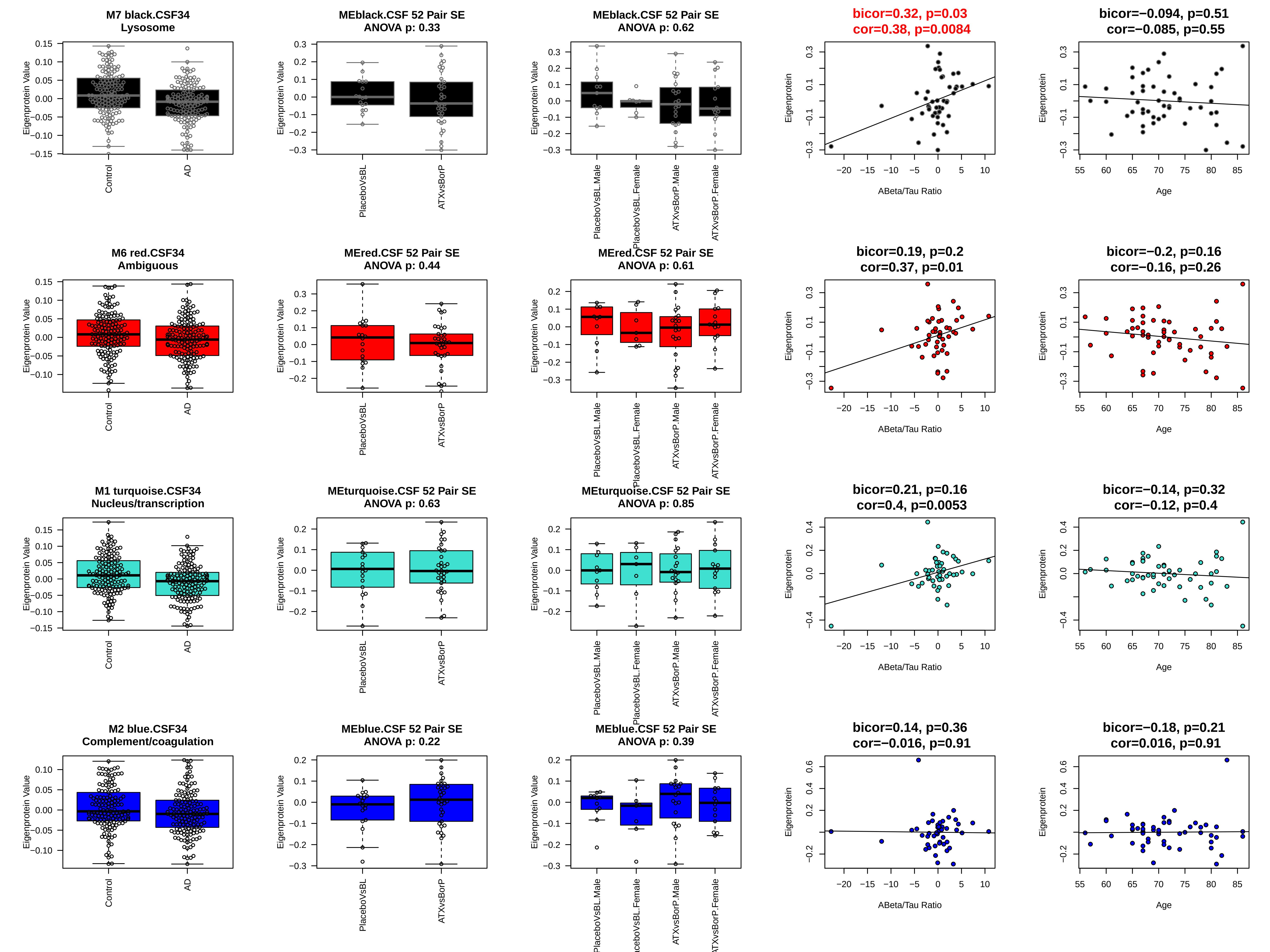
M7 black.CSF34
Lysosome
Eigenprotein Value
Control
AD
MEblack.CSF 52 Pair SE
ANOVA p: 0.33
PlaceboVsBL
ATXvsBorP
MEblack.CSF 52 Pair SE
ANOVA p: 0.62
PlaceboVsBL.Male
PlaceboVsBL.Female
ATXvsBorP.Male
ATXvsBorP.Female
bicor=0.32, p=0.03
cor=0.38, p=0.0084
Eigenprotein
ABeta/Tau Ratio
bicor=−0.094, p=0.51
cor=−0.085, p=0.55
Age
M6 red.CSF34
Ambiguous
MEred.CSF 52 Pair SE
ANOVA p: 0.44
MEred.CSF 52 Pair SE
ANOVA p: 0.61
bicor=0.19, p=0.2
cor=0.37, p=0.01
bicor=−0.2, p=0.16
cor=−0.16, p=0.26
M1 turquoise.CSF34
Nucleus/transcription
MEturquoise.CSF 52 Pair SE
ANOVA p: 0.63
MEturquoise.CSF 52 Pair SE
ANOVA p: 0.85
bicor=0.21, p=0.16
cor=0.4, p=0.0053
bicor=−0.14, p=0.32
cor=−0.12, p=0.4
M2 blue.CSF34
Complement/coagulation
MEblue.CSF 52 Pair SE
ANOVA p: 0.22
MEblue.CSF 52 Pair SE
ANOVA p: 0.39
bicor=0.14, p=0.36
cor=−0.016, p=0.91
bicor=−0.18, p=0.21
cor=0.016, p=0.91

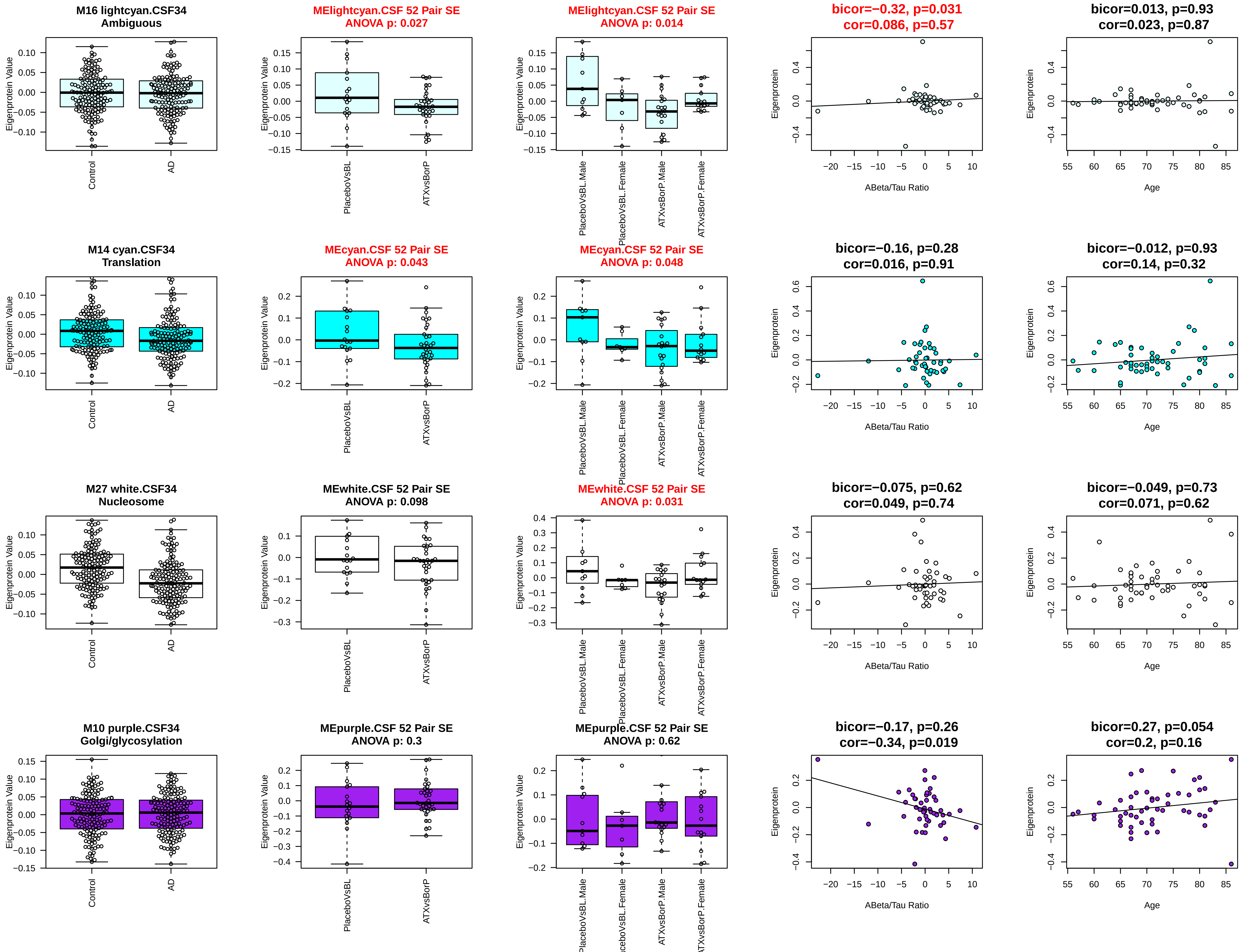
M16 lightcyan.CSF34
Ambiguous
MElightcyan.CSF 52 Pair SE
ANOVA p: 0.027
MElightcyan.CSF 52 Pair SE
ANOVA p: 0.014
bicor=−0.32, p=0.031
cor=0.086, p=0.57
bicor=0.013, p=0.93
cor=0.023, p=0.87
M14 cyan.CSF34
Translation
MEcyan.CSF 52 Pair SE
ANOVA p: 0.043
MEcyan.CSF 52 Pair SE
ANOVA p: 0.048
bicor=−0.16, p=0.28
cor=0.016, p=0.91
bicor=−0.012, p=0.93
cor=0.14, p=0.32
M27 white.CSF34
Nucleosome
MEwhite.CSF 52 Pair SE
ANOVA p: 0.098
MEwhite.CSF 52 Pair SE
ANOVA p: 0.031
bicor=−0.075, p=0.62
cor=0.049, p=0.74
bicor=−0.049, p=0.73
cor=0.071, p=0.62
M10 purple.CSF34
Golgi/glycosylation
MEpurple.CSF 52 Pair SE
ANOVA p: 0.3
MEpurple.CSF 52 Pair SE
ANOVA p: 0.62
bicor=−0.17, p=0.26
cor=−0.34, p=0.019
bicor=0.27, p=0.054
cor=0.2, p=0.16
Eigenprotein Value
Eigenprotein
Control
AD
PlaceboVsBL
ATXvsBorP
PlaceboVsBL.Male
PlaceboVsBL.Female
ATXvsBorP.Male
ATXvsBorP.Female
ABeta/Tau Ratio
Age

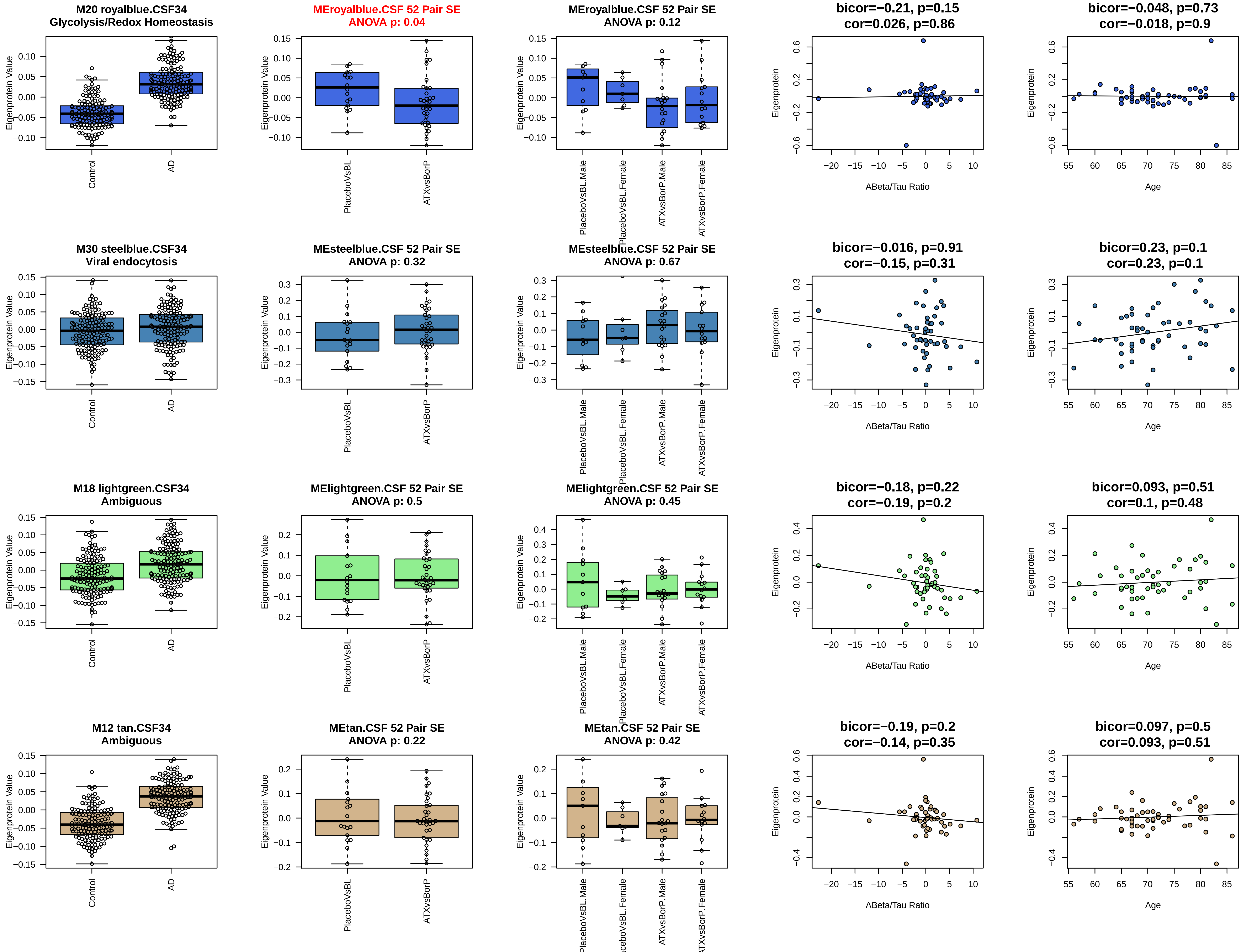
M20 royalblue.CSF34
Glycolysis/Redox Homeostasis
Eigenprotein Value
Control
AD
MEroyalblue.CSF 52 Pair SE
ANOVA p: 0.04
Eigenprotein Value
PlaceboVsBL
ATXvsBorP
MEroyalblue.CSF 52 Pair SE
ANOVA p: 0.12
Eigenprotein Value
PlaceboVsBL.Male
PlaceboVsBL.Female
ATXvsBorP.Male
ATXvsBorP.Female
bicor=−0.21, p=0.15
cor=0.026, p=0.86
Eigenprotein
ABeta/Tau Ratio
bicor=−0.048, p=0.73
cor=−0.018, p=0.9
Eigenprotein
Age
M30 steelblue.CSF34
Viral endocytosis
Eigenprotein Value
Control
AD
MEsteelblue.CSF 52 Pair SE
ANOVA p: 0.32
Eigenprotein Value
PlaceboVsBL
ATXvsBorP
MEsteelblue.CSF 52 Pair SE
ANOVA p: 0.67
Eigenprotein Value
PlaceboVsBL.Male
PlaceboVsBL.Female
ATXvsBorP.Male
ATXvsBorP.Female
bicor=−0.016, p=0.91
cor=−0.15, p=0.31
Eigenprotein
ABeta/Tau Ratio
bicor=0.23, p=0.1
cor=0.23, p=0.1
Eigenprotein
Age
M18 lightgreen.CSF34
Ambiguous
Eigenprotein Value
Control
AD
MElightgreen.CSF 52 Pair SE
ANOVA p: 0.5
Eigenprotein Value
PlaceboVsBL
ATXvsBorP
MElightgreen.CSF 52 Pair SE
ANOVA p: 0.45
Eigenprotein Value
PlaceboVsBL.Male
PlaceboVsBL.Female
ATXvsBorP.Male
ATXvsBorP.Female
bicor=−0.18, p=0.22
cor=−0.19, p=0.2
Eigenprotein
ABeta/Tau Ratio
bicor=0.093, p=0.51
cor=0.1, p=0.48
Eigenprotein
Age
M12 tan.CSF34
Ambiguous
Eigenprotein Value
Control
AD
MEtan.CSF 52 Pair SE
ANOVA p: 0.22
Eigenprotein Value
PlaceboVsBL
ATXvsBorP
MEtan.CSF 52 Pair SE
ANOVA p: 0.42
Eigenprotein Value
PlaceboVsBL.Male
PlaceboVsBL.Female
ATXvsBorP.Male
ATXvsBorP.Female
bicor=−0.19, p=0.2
cor=−0.14, p=0.35
Eigenprotein
ABeta/Tau Ratio
bicor=0.097, p=0.5
cor=0.093, p=0.51
Eigenprotein
Age

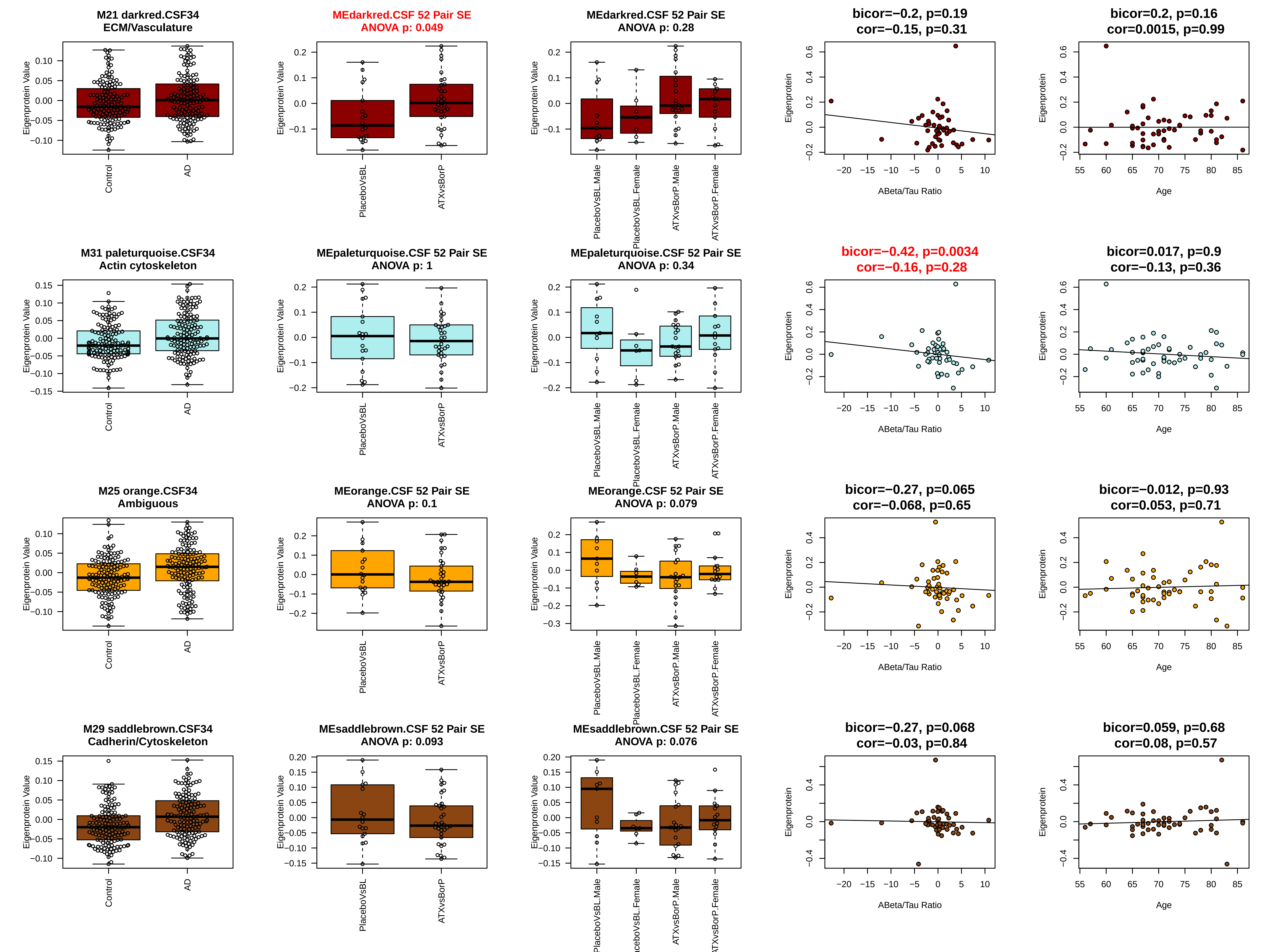
M21 darkred.CSF34
ECM/Vasculature
Eigenprotein Value
Control
AD
MEdarkred.CSF 52 Pair SE
ANOVA p: 0.049
PlaceboVsBL
ATXvsBorP
MEdarkred.CSF 52 Pair SE
ANOVA p: 0.28
PlaceboVsBL.Male
PlaceboVsBL.Female
ATXvsBorP.Male
ATXvsBorP.Female
bicor=−0.2, p=0.19
cor=−0.15, p=0.31
Eigenprotein
ABeta/Tau Ratio
bicor=0.2, p=0.16
cor=0.0015, p=0.99
Age
M31 paleturquoise.CSF34
Actin cytoskeleton
MEpaleturquoise.CSF 52 Pair SE
ANOVA p: 1
MEpaleturquoise.CSF 52 Pair SE
ANOVA p: 0.34
bicor=−0.42, p=0.0034
cor=−0.16, p=0.28
bicor=0.017, p=0.9
cor=−0.13, p=0.36
M25 orange.CSF34
Ambiguous
MEorange.CSF 52 Pair SE
ANOVA p: 0.1
MEorange.CSF 52 Pair SE
ANOVA p: 0.079
bicor=−0.27, p=0.065
cor=−0.068, p=0.65
bicor=−0.012, p=0.93
cor=0.053, p=0.71
M29 saddlebrown.CSF34
Cadherin/Cytoskeleton
MEsaddlebrown.CSF 52 Pair SE
ANOVA p: 0.093
MEsaddlebrown.CSF 52 Pair SE
ANOVA p: 0.076
bicor=−0.27, p=0.068
cor=−0.03, p=0.84
bicor=0.059, p=0.68
cor=0.08, p=0.57
